# SARS-COV-2 δ variant drives the pandemic in India and Europe via two subvariants

**DOI:** 10.1101/2021.10.16.21265096

**Authors:** Xiang-Jiao Yang

## Abstract

SARS-COV-2 evolution generates different variants and drives the pandemic. As the current main driver, δ variant bears little resemblance to the other three variants of concern, raising the question what features future variants of concern may possess. To address this important question, I compared different variant genomes and specifically analyzed δ genomes in the GISAID database for potential clues. The analysis revealed that δ genomes identified in India by April 2021 form four different groups (referred to as δ1, δ2, δ3 and δ4) with signature spike, nucleocapsid and NSP3 substitutions defining each group. Since May 2021, δ1 has gradually overtaken all other subvariants and become the dominant pandemic driver, whereas δ2 has played a less prominent role and the remaining two (δ3 and δ4) are insignificant. This group composition and variant transition are also apparent across Europe. In the United Kingdom, δ1 has quickly become predominant and is the sole pandemic driver underlying the current wave of COVID-19 cases. Alarmingly, δ1 subvariant has evolved further in the country and yielded a sublineage encoding spike V36F, A222V and V1264L. These substitutions may make the sublineage more virulent than δ1 itself. In the rest of Europe, δ1 is also the main pandemic driver, but δ2 still plays a role. In many European countries, there is a δ1 sublineage encoding spike T29A, T250I and Q613H. This sublineage originated from Morocco and has been a key pandemic driver there. Therefore, δ variant drives the pandemic in India and across Europe mainly through δ1 and δ2, with the former acquiring additional substitutions and yielding sublineages with the potential to drive the pandemic further. These results suggest a continuously branching model by which δ variant evolves and generates more virulent subvariants.

## INTRODUCTION

Since the initial cases were identified in December 2019, coronavirus disease 2019 (COVID-19) has caused the global pandemic in a way totally unexpected to almost everyone in the world. In a short 22 months, this pandemic has led to almost 238 million confirmed cases around the world, accounting to almost 3.0% of the entire population (according to the Our World in Data website, https://ourworldindata.org/; accessed on October 10, 2021). As there are many exposed but unreported cases, the real case count is much larger than this. As for mortality, almost 4.9 million people have died of this disease, accounting for over 1 out of 2,000 people. Moreover, many of the deceased were adult caregivers, thereby leaving behind thousands of orphans around the world [1,2]. For example, 1.5 million children across 21 countries lost a care-giving parent or grandparent due to COVID-19 from March 2020 to April 2021 [1]. Thus, this pandemic has caused lots of suffering and tragic loss of life. Moreover, it has crippled the economy and hindered its recovery around the world. To make the matter even much worse, this pandemic is still actively ongoing, with new waves of cases in many countries, including Israel, the United Kingdom, the United States of America, Canada, Singapore, Malaysia and Indonesia (Our World in Data). Notably, many of them, including Israel, the United Kingdom, U.S.A., Canada and Singapore are among the most vaccinated countries in the world. Thus, this pandemic remains a major and urgent public health crisis unprecedented in the recent human history.

The disease is caused by severe acute respiratory syndrome coronavirus 2 (SARS-CoV-2), with its first isolates sequenced and reported at the beginning of January 2020 [3]. The genomes sequences indicate that the virus is homologous to SARS-COV-1, an enveloped, positive-sense and single-stranded RNA virus that caused the severe acute respiratory syndrome (SARS) epidemic mainly in Asia from 2003 to 2004. By comparison, COVID-19 was declared a pandemic by the WHO at the beginning of March 2020 [3]. Since December 2019, there are multiple waves, with the newer ones much worse than the first one. From genomic surveillance, it has become gradually clear that virus evolution plays a key role in driving waves of cases in different countries and continents around the world. Thus, it is an important task to map out how the virus mutates and evolves. This also sheds light on the evolutionary trajectories of the virus, which will be helpful for preventing pandemics in the future.

For genomic surveillance, data sharing and annotation are critical. In this regard, the global initiative on sharing avian influenza data (GISAID) has played a critical role since the beginning of the pandemic and as of October 13, 2021, >4.25 million SARS-COV-2 genomes have been deposited into the database [4]. From genomic surveillance, it is clear that the virus has evolved dynamically and yielded many variants, with some being the main drivers of the pandemic. Among them are four dominant variants, α (B.1.1.7) [5], β (B.1.351) [6], γ (P.1) [7] and δ (B.1.617.2) [8], initially identified in the United Kingdom, South Africa, Brazil and India/Japan, respectively. The WHO has designated these strains as variants of concerns. One astonishing common theme is that they emerged from complete obscurity and then rapidly rose to become the pandemic drivers, raising the question about candidates as the potential future variants of concern. This question would greatly help us get well ahead of SARS-COV-2, (instead of ‘tail-gating’ it) and take ‘pre-emptive’ strikes against such variants. To address this important question, I have closely tracked how different variants evolve. Here, I describe that δ variant drives the pandemic in India and Europe mainly through δ1 and δ2, with the former acquiring additional virulent substitutions and yielding multiple sublineages with the potential to drive the pandemic further. This study complements two companion reports on how δ1 and δ2 subvariants drive the pandemic in the USA [9], Indonesia, Singapore and Malaysia [10].

## RESULTS AND DISCUSSION

### SARS-COV-2 evolution drives the pandemic around the world

As shown in Fig. 1A, there have been six waves of COVID-19 cases in the world since December 2019. The initial identification of α [5], β [6] and γ (P.1) [7] variants at the end of 2020 and the beginning of 2021 signaled that SARS-COV-2 evolution is the major driver of the pandemic. Accordingly, the pandemic can be divided into pre-VOC (variant of concern) and VOC phases. In the first few months of 2021, these three VOCs were the main pandemic driver but just as they caught the world by surprise, δ variant suddenly emerged from complete obscurity [8], rapidly overtook the three other VOCs and multiple variants of interests (such as μ, κ and Ida, Fig. 1A) [9], and gradually became the major pandemic driver. This raises an important question about what features such a pandemic driver possesses. Answers to this question will help identify potential new VOCs. The initial and obvious guess was that δ variant possess key spike L452R, T478K and P681R substitutions (Table 1). These substitutions are individually present in many other variants, so one possibility is that their combination is deadly. Even if so, one question is whether this combination is sufficient. Related to this, despite its possession of L452R and P682R, κ variant has not spread from India and colonized other countries as δ variant has done. One main difference is that T478K of δ variant is replaced with E484K in κ variant. Structural and molecular analysis [11-15] indicated that this difference is insufficient to explain the huge difference between these two variants in terms of their ability to become pandemic drivers. This raises the question what else makes δ variant become a dominant pandemic driver so rapidly. To answer this question, it is necessary to understand how this variant has evolved. As it originated in India, I first analyzed its evolution there.

**Figure 1.**
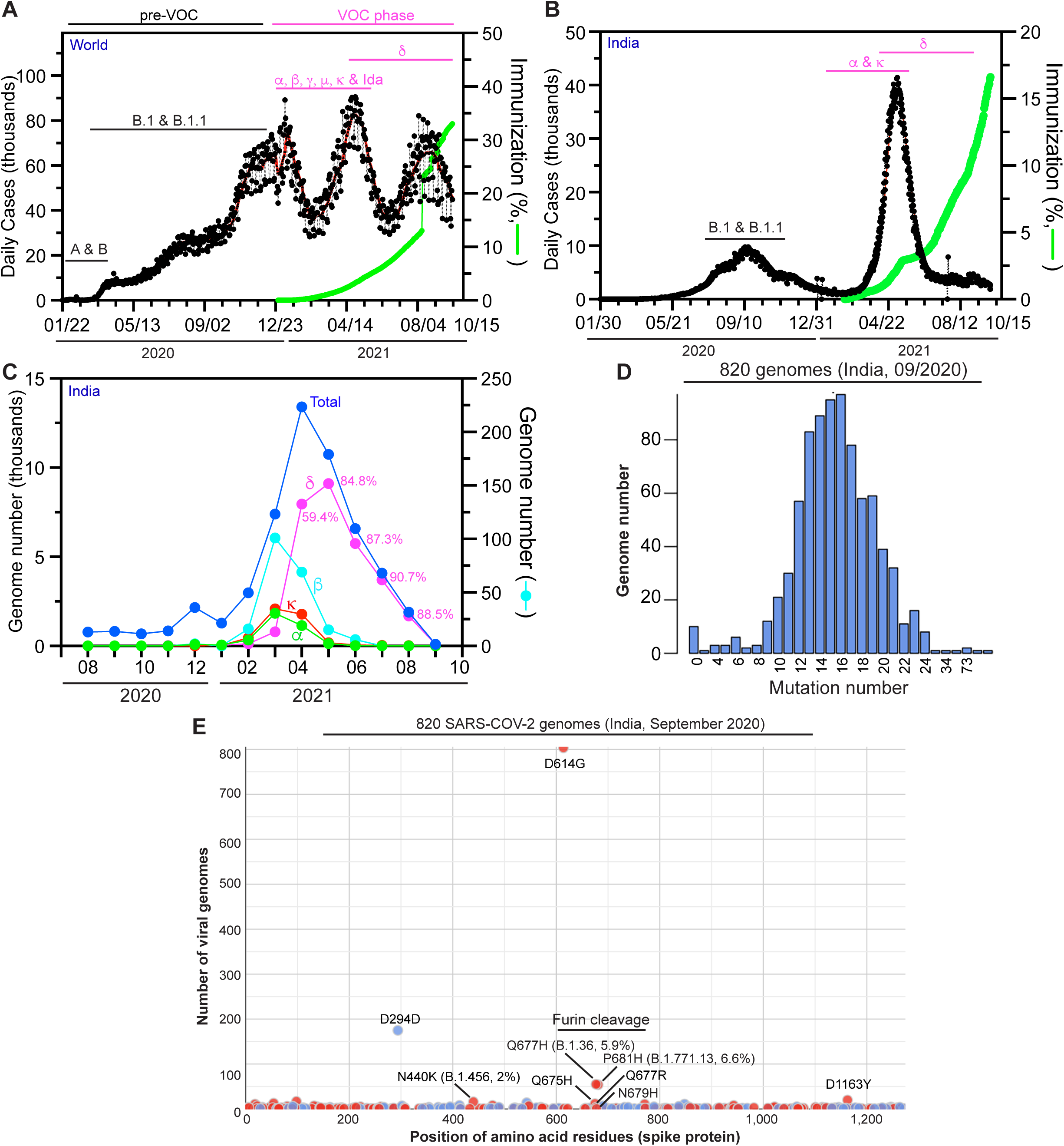
SARS-COV-2 evolution drives the pandemic in India. (**A-B**) Epidemiological curve and immunization progress in the world (A) and India (B). The pandemic can be divided into pre-VOC (variant of concern) and VOC phases. The initial reports of α, β and γ variants in December 2020 and at the beginning of 2021 are the defining moments that separate the two phases of the pandemic. For preparation of panels A-B, the Our World in Data website (https://ourworldindata.org/) was accessed on September 29, 2021. (**C**) Monthly distribution of variant genomes detected in India. (**D**-**E**) Mutation profile of SARS-COV-2 genomes from 820 COVID-19 genomes identified in India during September 2020. The genomes were downloaded from the GISAID SARS-COV-2 sequence database on September 29, 2021 for mutation profiling via Coronapp [16,17]. Shown here are the mutation load (D) and spike substitutions (E).

**Table 1.**
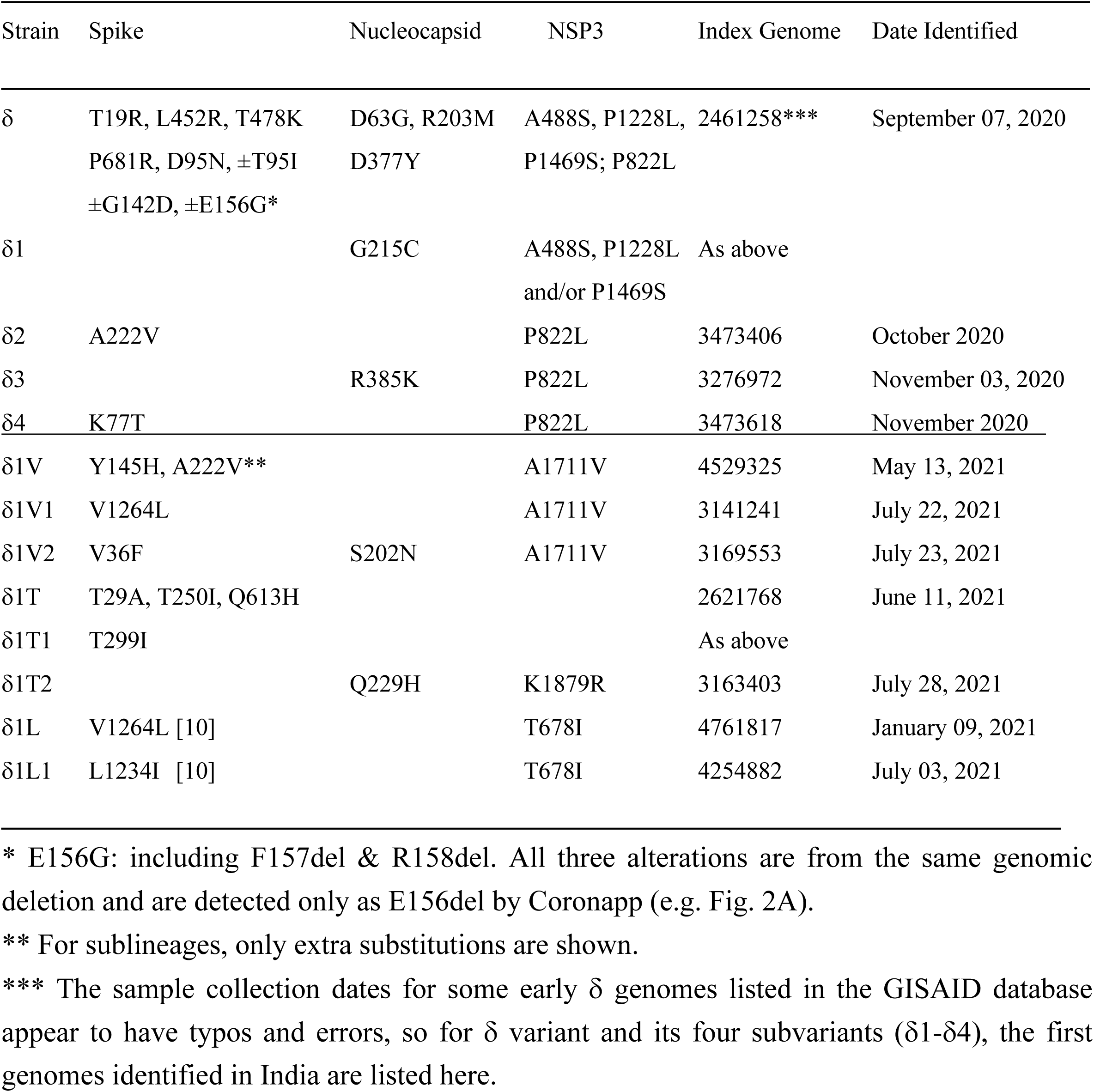
List of substitutions in δ variant and its subvariants.

### δ variant rapidly emerged as the major pandemic driver in India

As shown in Fig. 1B, there have been two waves of COVID-19 cases in India, with the first one starting in the summer of 2020 and ending a few months later. To understand the genomic basis of the epidemiological wave, I analyzed different variant genomes identified in India and deposited into the GISAID database [4]. As shown in Fig. 1C, α and κ variants were the major drivers in March 2021, but δ variant rapidly overtook all variants and became the major driver only one month later. Since May 2021, it has been present over 80% COVID-19 cases (Fig. 1C). Its rapid rise raises important questions about its evolution and on its molecular and genomic characteristics.

Related to its evolution, one would wonder whether it came from the first wave of cases in 2020. To understand the genomic basis of that wave, I performed mutation profiling of 820 SARS-COV-2 genomes in COVID-19 cases identified in India during September 2020. For this, I analyzed the genomes by utilizing Coronapp, an efficient web-based mutation annotation application [16,17]. The analysis indicated that on the average, there are 15-16 mutations per genome (Fig. 1D). In terms of spike substitutions, only D614G is present in all genomes, while P681H, Q677H and N440K are only encoded in 6.6%, 5.9% and 2% genomes, respectively (Fig. 1D & Table 1). δ variant displays signature spike substitutions such as L452R, T478K and P681R, as well as three signature nucleocapsid substitutions, including R203M (Table 1). These spike and nucleocapsid signature substitutions are absent in α [5], β [6] and γ [7] variants. Strikingly, among the 820 genomes, there is only one genome possessing these four signature substitutions (Table S1). If the sample collection date for this genome is correct, it may be a δ variant precursor. Together, these observations suggest that δ variant did not evolve from the majority of cases identified during the first wave, but rather originated from some rare evolutionary events. Conceptually, this is reminiscent of the way by which α variant emerged in the United Kingdom in September 2020 [5]. Emergence of α and δ variants as two major pandemic drivers from complete obscurity also raises an important issue about how to detect them in the early stages of their evolution. Due to the low abundance at such stages (often lower than 1%), this is challenging for genomic surveillance unless all COVID-19 cases are sequenced.

### δ variant has evolved and yielded different subvariants in India

Then I asked how δ variant had evolved by March 2020 when it was still obscure. To address this question, I utilized Coronapp [16,17] to analyze the mutation profile of 985 δ-genomes identified in India by March 2021. This analysis revealed different subgroups of δ-genomes (Fig. 2). In addition to what is presented in Fig. 2, Coronapp generated a list of mutations that each genome carries. I manually inspected this list (Table S2) to identify the subgroups. Initially, spike substitutions such as E156del, D950N, G142D and A222V (Fig. 2A & Table 1) were used to identify such subgroups, but the outcome was not clear cut, perhaps due to loss of G142D, E156del and/or D950N in some genomes during evolution. Then I noticed that nucleocapsid substitutions G215C and R385K (Fig. 2B) were mutually exclusive. Moreover, they were associated with two distinct sets of NSP3 substitutions: 1) G215C with A488S, P1228L and P1469S of NSP3; and 2) R385K with P822L of NSP3 (Fig. 2B-C). With this guide, I noticed that spike K77T and A222V are mutually exclusive, but both are associated with P822L. Thus, it appeared that the NSP3 substitutions are good markers to separate two large groups, with one group defined by nucleocapsid G215C along with three substitutions of NSP3 (A488S, P1228L and P1469S). The other group is defined by P822L of NSP3. This group can be further divided into three subgroups defined by spike K77T, A222V and nucleocapsid R385K (Table 1). This classification also appears to be true when I inspected mutation reports generated by CoVsurver, enabled by GISAID [4]. Thus, δ variant had yielded four distinct groups of subvariants in India by March 2021.

**Figure 2.**
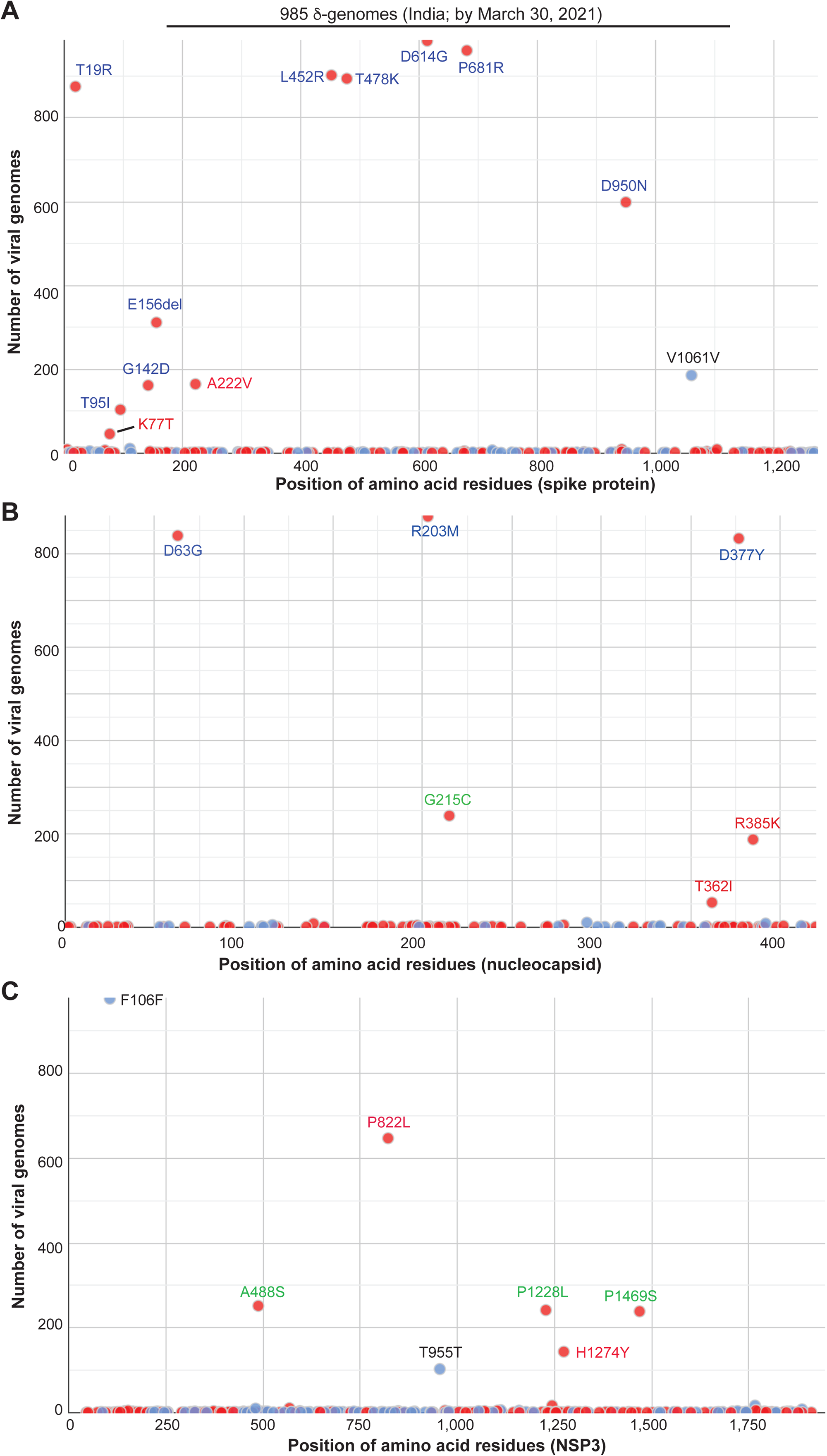
Mutation profile of 985 δ-genomes identified in India by the end of March 2021. The genomes were downloaded from the GISAID database on September 28, 2021 for mutation profiling via Coronapp. Shown are substitutions in spike (A), nucleocapsid (B) and NSP3 (C) proteins. Note that CoVsuver identifies the genomic deletion spanning spike codons for residues 156-158 as E156G, F157del & R158del, whereas this samll deletion is detected only as E156del by Coronapp (A).

To substantiate this, I carried out phylogenetic analysis of 291 δ-genomes identified in India by March 21, 2021. This analysis uncovered 5 major clusters (Fig. 3A). To assign the clusters, CoVsurver was used to generate mutation reports for some representative genomes of each cluster. Manual inspection of the reports indicated that four clusters correspond to the four groups mentioned above, so these four groups are referred to as δ1, δ2, δ3 and δ4, with nucleocapsid G215C, spike A222V, nucleocapsid R385K and spike K77T as signature substitutions, respectively. While δ1 is associated with A488S, P1228L and P1469S of NSP3, δ2, δ3 and δ4 encode P822L of NSP3. The fifth cluster from phylogenetic analysis is related to δ3 but does not carry nucleocapsid R385K, so this cluster is referred to as pre-δ3. Therefore, δ variant had generated 4-5 subvariants in India by March 2021.

**Figure 3.**
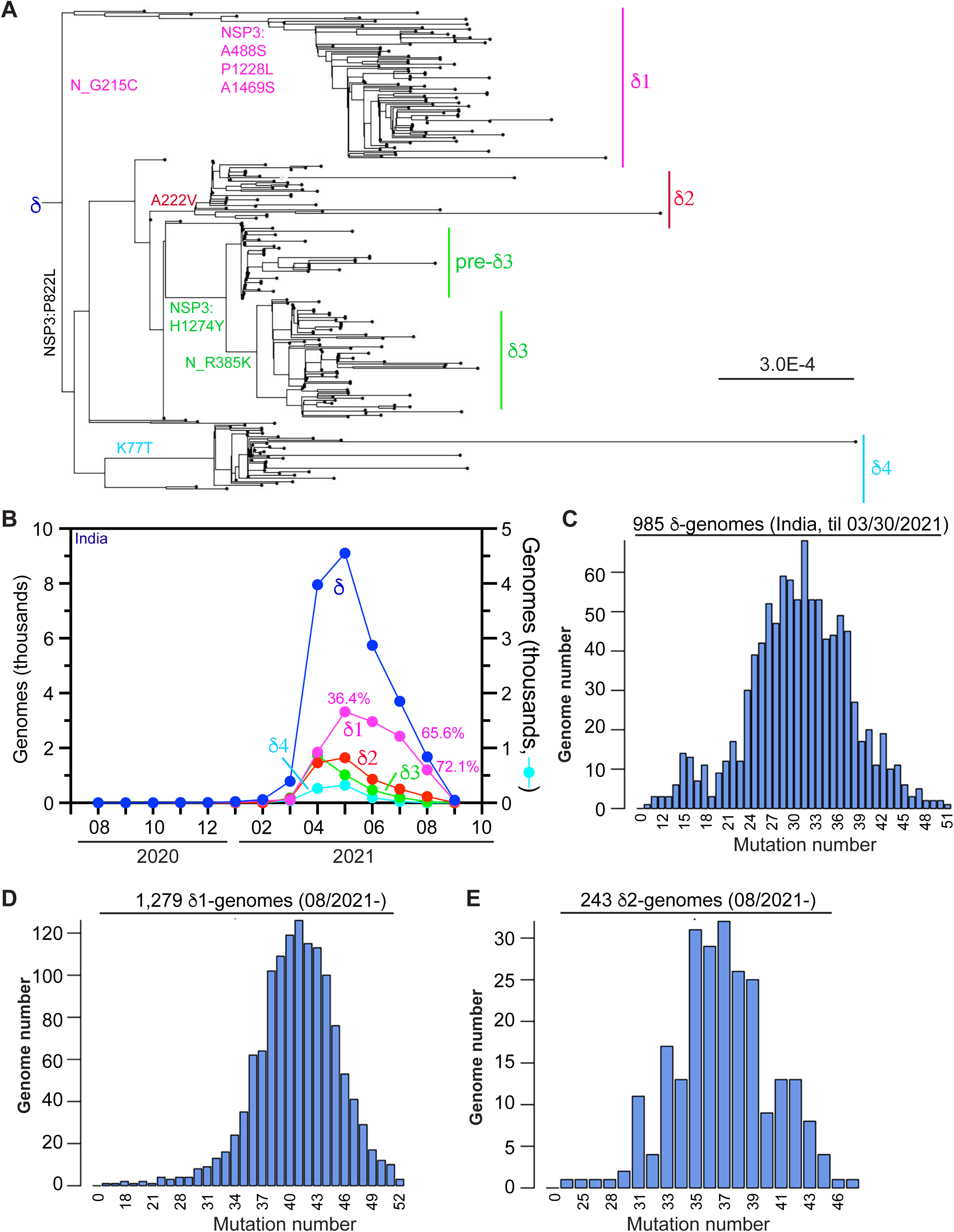
Analysis of δ subvariants in India. **(A)** Phylogenetic analysis of 291 δ-genomes identified in India by March 21, 2021. The genomes were downloaded from the GISAID SARS-COV-2 genome sequence database on September 14, 2021 for phylogenetic analysis. Only high-coverage genomes with complete date information on sample collection were used. The package RAxML-NG was used to generate 20 maximum likelihood trees and the bestTree for presentation via Figtree as in Fig. 3D. The strain names and GISAID accession numbers of the genomes are detailed in Figure S1. (**B**) Monthly distribution of δ subvariants in India. The analysis was done on the GISAID website on September 28, 2021. (**C**) Mutation load of 985 δ genomes identified in India by March 2021. The genomes were obtained and analyzed as in Fig. 2, but distribution of the mutation numbers in the genomes is shown here. (**D-E**) Mutation load of δ1 or δ2 genomes identified in India after August 01, 2021. The genomes were downloaded from the GISAID database on September 28, 2021 for mutation profiling via Coronapp. See the Materials and Methods section, as well as Table 1, for markers of different δ subvariants.

This raises the question how these subvariants have grown since the beginning of 2021. As shown in Fig. 3B, the number of genomes encoding δ1-δ4 subvariants was very low by January 2021. After that, the number has changed dynamically. Among the four subvariants, δ4 is minor. In April 2021, δ1, δ2 and δ3 genome numbers were comparable, but δ1 became dominant afterwards. In August 2021, it corresponded to >70% genomes sequenced (Fig. 3B). Thus, δ1 rapidly became a major pandemic driver in India. Among the remaining three subvariants, δ2 played a less important role than δ1 in August and September 2021 (Fig. 3B). For example, in August 2021, the number of δ2 genomes was ∼20% of that for δ1 (Fig. 3B). By comparison, δ3 and δ4 became almost insignificant in August and September 2021 (Fig. 3B). These observations also suggest that the distinct substitutions in the four subvariants make them players of different importance in driving the pandemic in India.

Shown in Fig. 3C is distribution of the mutation numbers of the 985 δ genomes identified in India by March 2021. The average load is about 33 mutations per genome. The average mutation load reached 41 and 37 for δ1 and δ2 genomes, respectively (Fig. 3D-E). Notably, δ1 and δ2 genomes identified in August 2021 were 1,279 and 243, respectively, supporting that δ1 and δ2 subvariants were responsible for ∼80% and ∼20% COVID-19 cases, respectively (Fig. 3D-E). Because δ1 subvariant possesses two extra NSP3 substitutions than δ2, the evolutionary speed or mutation rate of δ1 subvariant is slightly faster than δ2. This raises the question what makes δ1 a much more prominent pandemic driver than δ2. This also highlights the importance of the δ1 substitutions not shared by δ2 (Table 1). Compared to δ1, δ2 possesses an extra spike substitution, A222V (Fig. S3 & Table 1). Thus, it is not spike substitutions but rather extra nucleocapsid and NSP3 substitutions that make δ1 much more powerful in driving the pandemic than δ2 subvariant.

### Spread of δ subvariants around the world

Shown in Fig. 4A is phylogenetic analysis of 264 δ-genomes identified by March 09, 2021. This analysis revealed that similar to what was observed with δ-genomes from India, δ genomes identified around the world by March 09, 202 also formed 5 different clusters. They can be assigned using the same designation system: δ1, δ2, pre-δ3, δ3 and δ4 (Fig. 4A & Table 1). However, different from what was observed in India (Fig. 3A), the δ1 cluster is the largest, with many of its genomes identified in Europe (Fig. 4A), suggesting that δ1 was already the most prominent subvariant in the rest of the world by March 09, 2021. Monthly distribution of δ subvariants confirmed this and indicated that δ1 rapidly became a major δ subvariant in Europe and around the world after March 2021 (Fig. 4B-C).

**Figure 4.**
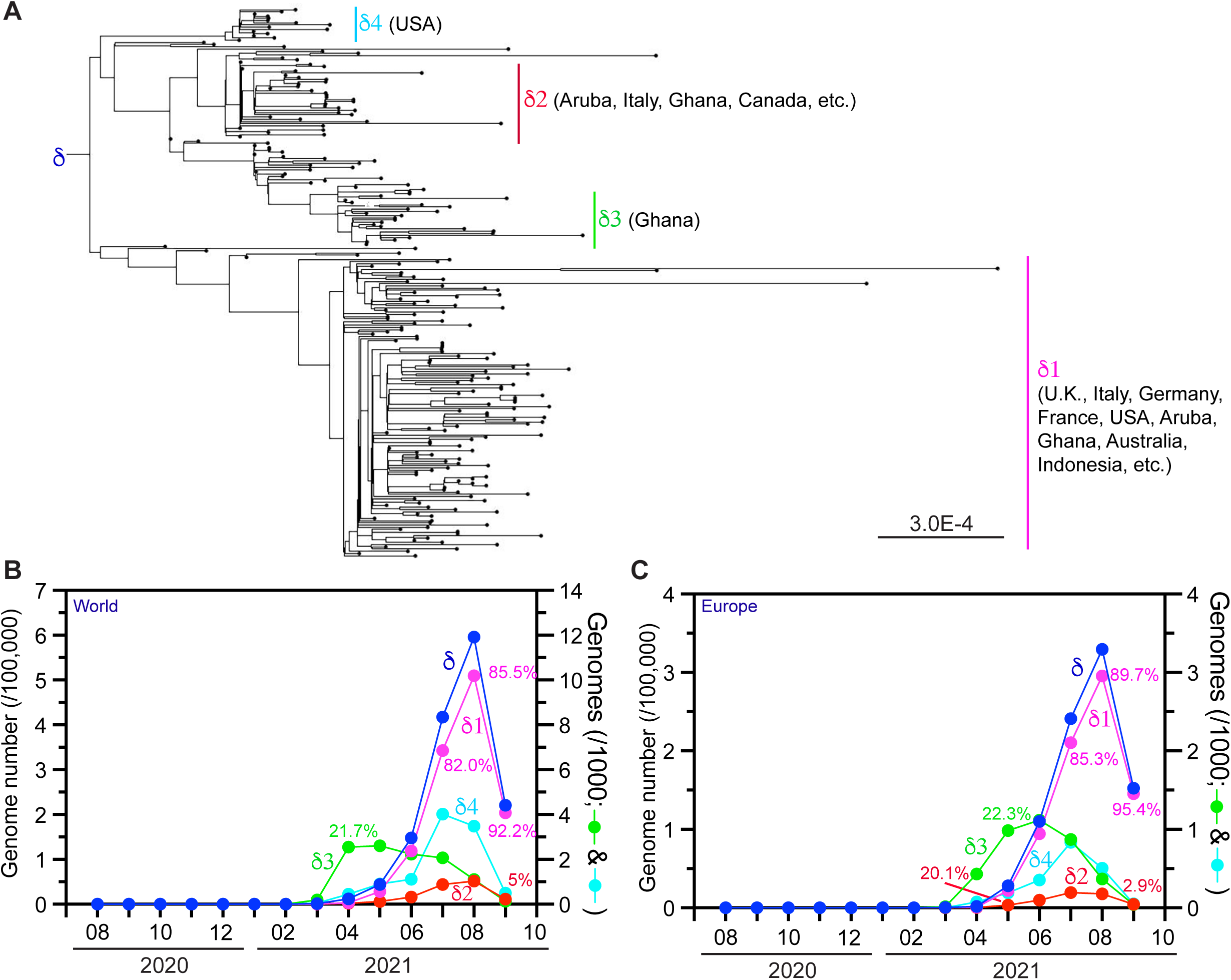
Analysis of δ variant genomes identified in the world and Europe. **(A)** Phylogenetic analysis of 264 δ-genomes identified by March 09, 2021. The genomes were downloaded from the GISAID database on September 14, 2021 for phylogenetic analysis. Only high-coverage genomes with complete sample collection date information were used. The package RAxML-NG was used to generate 20 maximum likelihood trees and the bestTree for presentation via Figtree as in Fig. 3D. The strain names and GISAID accession numbers of the genomes are detailed in Figure S2. (**B-C**) Monthly distribution of δ1 subvariants in the world and Europe. The analysis was done on the GISAID website on September 30, 2021.

### Emergence of δ1 subvariant as a major pandemic driver in the United Kingdom

As shown in Fig. 5A, there have been 4-5 waves of COVID-19 cases in the United Kingdom, with α variant being the major driver behind the third wave, occurring from December 2020 through March 2021. Vaccination campaign initiated in December 2020 and other public health measures may all have contributed to the rapid decline of α variant cases after the peak. Starting in April and May 2021, δ variant gradually became the major driver (Fig. 5B). As this variant has yielded δ1, δ2, δ3 and δ4 (Figs 3A-B & 4), I analyzed each of them. As shown in Fig. 5B, δ1 variant gradually became the major driver, but the other three δ subvariants played much less important roles. In March and April 2021, δ2 subvariant was still present in 21.5% and 11.1% in δ1 genomes (Fig. 5B), but the percentage rapidly declined to 1.0% in September 2021. Notably, this decline is much more dramatic than what was observed in India (Fig. 3B), raising the intriguing possibility that vaccination might have contributed to this rapid decline of δ2 cases in the U.K. If so, this subvariant may be more sensitive to vaccination than δ1 subvariant. Thus, it is possible that vaccination has helped make δ1 subvariant almost the sole pandemic driver in the country.

**Figure 5.**
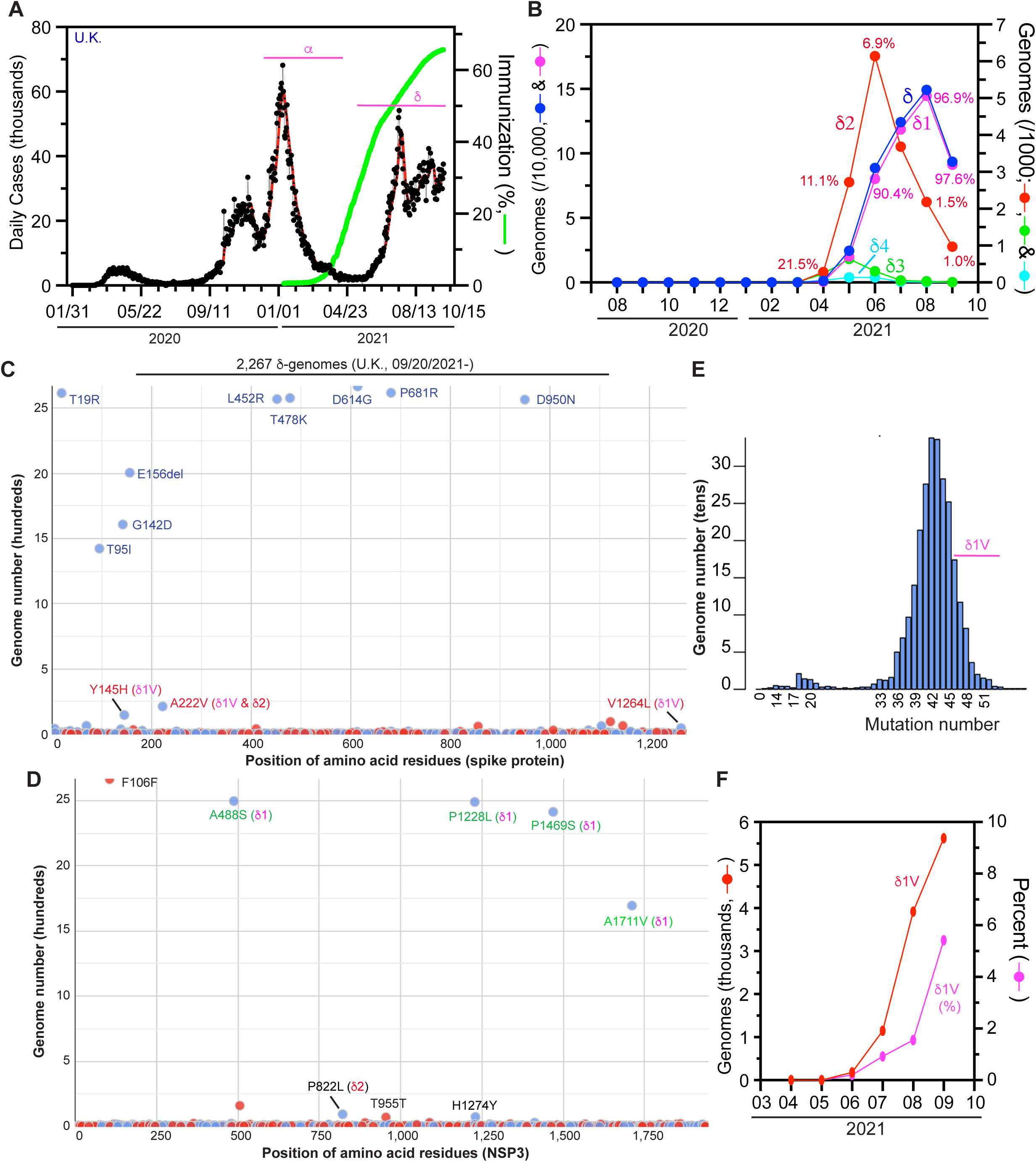
Analysis of δ variant in the United Kingdom. (**A**) Epidemiological curve and immunization progress in the country. For preparation of this panel, the Our World in Data website (https://ourworldindata.org/) was accessed on September 30, 2021. (**B**) Monthly distribution of δ1 subvariants in the country. The analysis was done on the GISAID website as for Fig. 3C on September 29, 2021. Because some δ1 genomes also encode spike A222V, the NSP3 substitution P822L was used along with spike A222V to identify δ2 genomes in the GISAID database. (**C-D**) Mutation profile of 2,267 δ1-genomes from in the country. The genomes were downloaded from the GISAID SARS-COV-2 genome sequence database on September 28, 2021 for mutation profiling via Coronapp. Shown in (C) and (D) are substitutions in spike and NSP3 proteins, respectively. (**E**) Mutation load in δ1 genomes. The distribution was generated via Coronapp as in (B-C), but the mutation load distribution among the genomes is shown here. (**F**) Monthly δ1 genomes that encode Y145H and A222V. This sublineage is referred to as δ1V. See Fig. S4 for additional information.

An important question is how δ variant has evolved in the U.K. To map the evolutionary trajectory, I utilized Coronapp [16,17] to analyze the mutation profile of 2,267 δgenomes identified in the country during September 2021. As expected, the predominant majority encodes δ1 subvariant (Fig. 5C-D1). The average mutation load is 42-43 per genome (Fig. 5E). This is 1-2 mutations more than δ1 subvariant genomes identified in India around the same time (Fig. 3D). Related to this difference, about 60% of the δ1 genomes from the U.K. encode A1711V of NSP3 as an extra substitution (Fig. 5D). Among this subgroup, there is a sublineage encoding two extra spike substitutions, Y145H and A222V (Fig. 5C). This is alarming as A222V is a signature substitution of δ2 subvariant (Table 1). Moreover, A222V is a signature substitution of the SARS-COV-2 GV clade, a major pandemic driver in Europe during the summer of 2020. Thus, A222V may make a variant more virulent. For convenience, this A222V-encoding δ1 sublineage is referred to as δ1V, where the letter V denotes V222. This sublineage carries at least three extra substitutions: spike Y145H, A222V and NSP3 A1711V (Table 1). This sublineage was first identified in May 2021 and since then, its genome number has increased rapidly, reaching almost 6% in the U.K. in September 2021 (Fig. 5F). The dramatic increase also supports that A222V makes δ1V more virulent than δ1 variant itself.

While analyzing V1264L-encoding genomes in another study [10], I found this substitution in a subset of δ1V genomes. As shown in Fig. S4A, V1264L is present in 20% δ1V genomes. Moreover, a subset of these genomes encodes spike S13T (Fig. S4A). S13 is in the signal peptide and its replacement by threonine may affect spike protein processing. While the average mutation load is 44 mutations per δ1V genome, this increases to 48 in the V1264L-encoding δ1V genomes [10]. V1264L is present in a δ1 sublineage that drives the pandemic in Indonesia, Singapore and Malaysia, and confers an acidic di-leucine motif in the cytoplasmic tail of spike protein for its cytoplasmic traffic and processing [10]. Thus, this V1264L-encoding δ1V sublineage (referred to as δ1V1) may be more virulent than δ1V itself.

Alarmingly, spike V36F is present in 10% δ1V genomes (Fig. S4A). This substitution is mutually exclusive with V1264L, suggestive of a second δ1V sublineage. This second sublineage encodes V36F as an extra substitution and is thus referred to as δ1V2. The first δ1V2 genome was identified in the U.K. on July 23, 2021 (Table 1). There are now 991 such genomes in the GISAID database. Coronapp analysis of these genomes revealed that almost all of them encodes an extra nucleocapid substitution, S202N (Fig. S4B). This is adjacent to R203M, a signature substitution shared by κ and δ variants. In agreement with the extra V36F and S202N substitution, the average mutation load is 44 mutations per δ1V2 genome (Fig. S4C). V36 is adjacent to A222 (Fig. S4D). Because both are part of an extensive hydrophobic interaction network (Fig. S4D), V36F may synergize with A222V to improve this network, suggesting that δ1V2 may be even more virulent than δ1V itself. Therefore, δ1 subvariant has not only driven the pandemic in the U.K. since April 2021 but also yielded different sublineages with potential to drive this pandemic even further.

### δ1 subvariant drives the pandemic in other European countries

What has happened in the U.K. raised the question whether similar events have also occurred in the rest of Europe. To address this question, I analyzed how δ subvariants drive the pandemic in Italy, Spain, France and Germany. Shown in Fig 6A-D is temporal distribution of the four δ subvariants in these countries. As occurred in the U.K., δ1 subvariant became the dominant pandemic driver in all four countries. Different from what has happened in the U.K., δ2 subvariant still played a role in Italy (Fig. 6A), France (Fig. 6C) and Germany (Fig. 6D) in September 2021. The situation in Spain (Fig. 6B) is more similar to what has happened in the U.K., with δ1 being predominant and δ2 subvariant playing an insignificant role.

**Figure 6.**
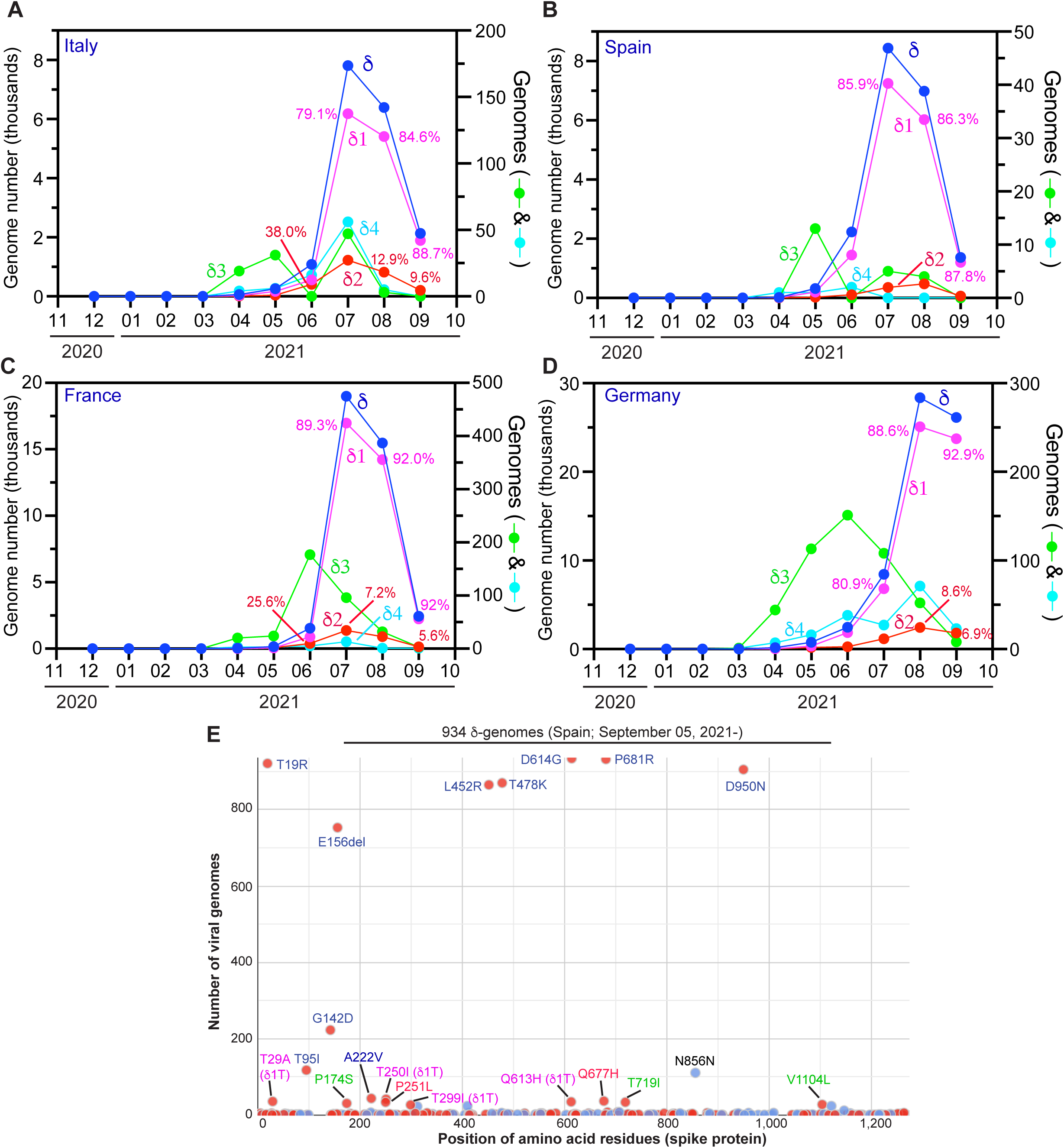
Analysis of δ subvariants identified in Europe. (**A-D**) Monthly distribution of the four δ subvariants in Italy (A), Spain (B), France (C) and Germany (D). The genomes were identified as in Fig. 5B. (**E**) Mutation profile of 934 δ1 genomes identified in Spain after September 05, 2021. The genomes were downloaded from the GISAID database on September 30, 2021 for mutation profiling via Coronapp. Shown here are substitutions in spike protein. See Fig. S5 for additional information.

Mutation profiling of 934 δ1 genomes identified in Spain after September 05, 2021 revealed a sublineage encoding spike T29A, T250A and Q613H (Fig. 6E). This sublineage is referred to as δ1T, where the letter T refers to T29 and T250, two spike residues to be replaced. A subset of the genomes also encode spike T299A and form an offspring strain, referred to as δ1T1 (denoting a derivative of δ1T). δ1T is also present in genomes from Italy, France and Germany even though the genome number is very low in Italy (Fig. S4). According to GISAID, the first δ1T genome sequence was identified in Japan on June 11, 2021 (Table 1), but this case has travel history to Morocoo. The second case was reported from Morocco on June 20, 2021 and additional cases with travel history to the country were identified in Japan, Singapore and South Korea. Among 62 δ genomes reported from Morocco (GISAID, accessed on October 03, 2021), 28 (45.2%) correspond to δ1T. Thus, δ1T sublineage is a key driver behind the latest wave of COVID-19 cases (from July to September 2021; https://ourworldindata.org/) in the country.

What occurred in Morocco suggests that δ1T is more virulent than δ1 itself. To investigate how δ1T has fared in Europe, I analyzed the number of δ1T genomes in different countries there. As shown in Fig. 7A, δ1T sublineage has grown steadily in Spain, France, Belgium and Demark. Notably, the δ1T genome number reached ∼10% of all δ genomes identified in Belgium in August 2021. In the rest of the world, this sublineage accounted for ∼8% of all δ genomes identified in Quebec, Canada in August 2021. δ1T sublineage has evolved dynamically and yielded new strains with extra spike, nucleocapsid and NSP3 substitutions (Figs 7B-C & S6A). One such strain (referred to as δ1T2) possesses an extra nucleocapsid substitution, Q229H. δ1T2 genomes are mainly from Denmark. This is alarming and suggests that δ1T has yielded a major new strain in the country. This new strain exhibits an average of 49 mutations per genome, making it one of the most mutated SARS-COV-2 variants identified so far. It encodes one NSP3 substitution (K1879R) and three extra silent mutations (nucleocapsid D571D and Q762Q, along with Q1756Q of NSP3). Thus, as in the U.K., δ1 subvariant has not only driven the pandemic in the rest of Europe but also yielded different sublineages with additional substitutions since April 2021.

**Figure 7.**
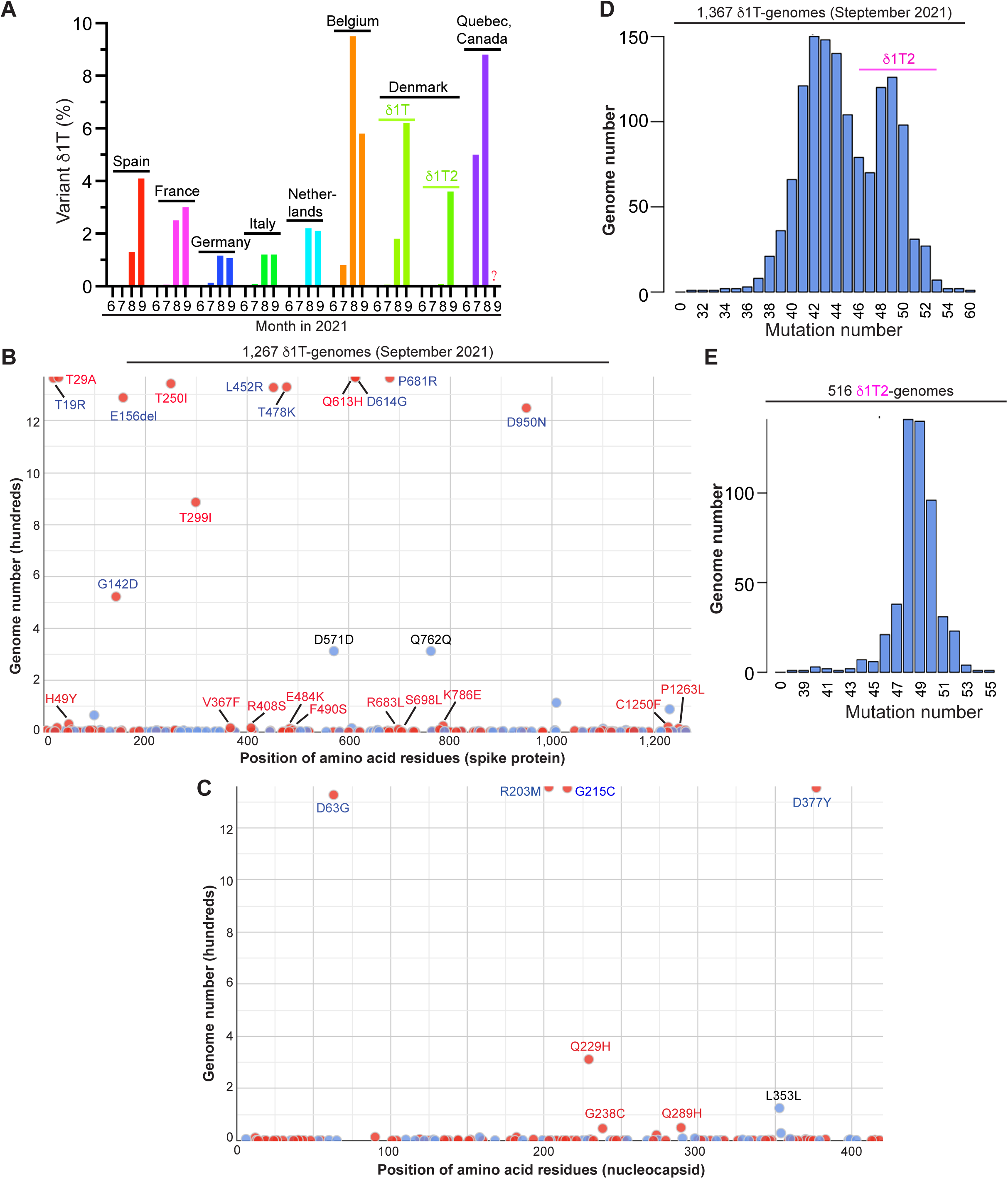
Emergence and spread of δ1T sublineage that encodes spike T29A, T250I and Q613H. (**A**) Monthly number of δ1T genomes identified in seven European countries and Quebec, Canada from June to September 2021. One subset of δ1T genomes (referred to as δ1T1) also encode T299I and another subset carries the nucleocapsid substitution Q229H (referred to as δ1T2, only identified in Denmark). The first δ1T genome sequence was first submitted to the GISAID database on June 11, 2021 in Japan, but the case has travel history to Morocoo (Table 1). δ1T sublineage is a key driver behind the latest wave of COVID-19 cases in the country. (**B-C**) Mutation profile of 1,267 δ1T genomes identified in different countries around the world. The genomes were downloaded from the GISAID database on October 01, 2021 for mutation profiling via Coronapp. Shown in (B) and (C) are substitutions in spike and NSP3 proteins, respectively. (**D-E**) Mutation load in δ1T sublineage (D) and a subgroup (referred to as δ1T2) that also encodes the nucleocapsid substitution Q229H (E). The distribution in panel D was generated via Coronapp as in (B-C), but the mutation load distribution among different genomes is shown here. For panel E, 516 δ1T2 genomes were downloaded from the GISAID SARS-COV-2 genome sequence database on October 01, 2021 for mutation profiling via Coronapp. The substitutions present in spike, nucleocapsid and NSP3 are shown Fig. S6.

### Mechanistic impact of spike, nucleocapsid and NSP3 substitutions in δ subvariants

Having identified different δ subvariants and their sublineages (Table 1), I next considered mechanistic impact of the associated new substitutions. Many of them are located at the N-terminal domain of spike protein (Fig. 8A) [18]. T29, K77, Y145 and T250 are within a super antigenic site in the N-terminal domain (Fig. 8A-B) [19]. Among them, T250 is located within an unstructured loop (Fig. 8B). Thus, T29A, K77T, Y145H and T250I may confer immune evasion. Both V36 and A222 are part of an extensive hydrophobic interaction network (Fig. S4D). In addition to L54, L560 and I285, there are 5 aromatic residues (F562, Y38, F275, F85 and F200). The side chain of V36 is 3.4 Å away from that of A222, supporting interaction between these two hydrophobic residues. V36F and A222V should facilitate the hydrophobic interaction. As shown in Fig. 8D, the methyl group of the T299 side chain is 3.7 Å away from that of T315, indicating hydrophobic interaction between the side chains. The methyl group of T299 is 6.6 Å away from that of T599, so T299I may facilitate hydrophobic interaction of I299 with the methyl groups of T513 and T599.

**Figure 8.**
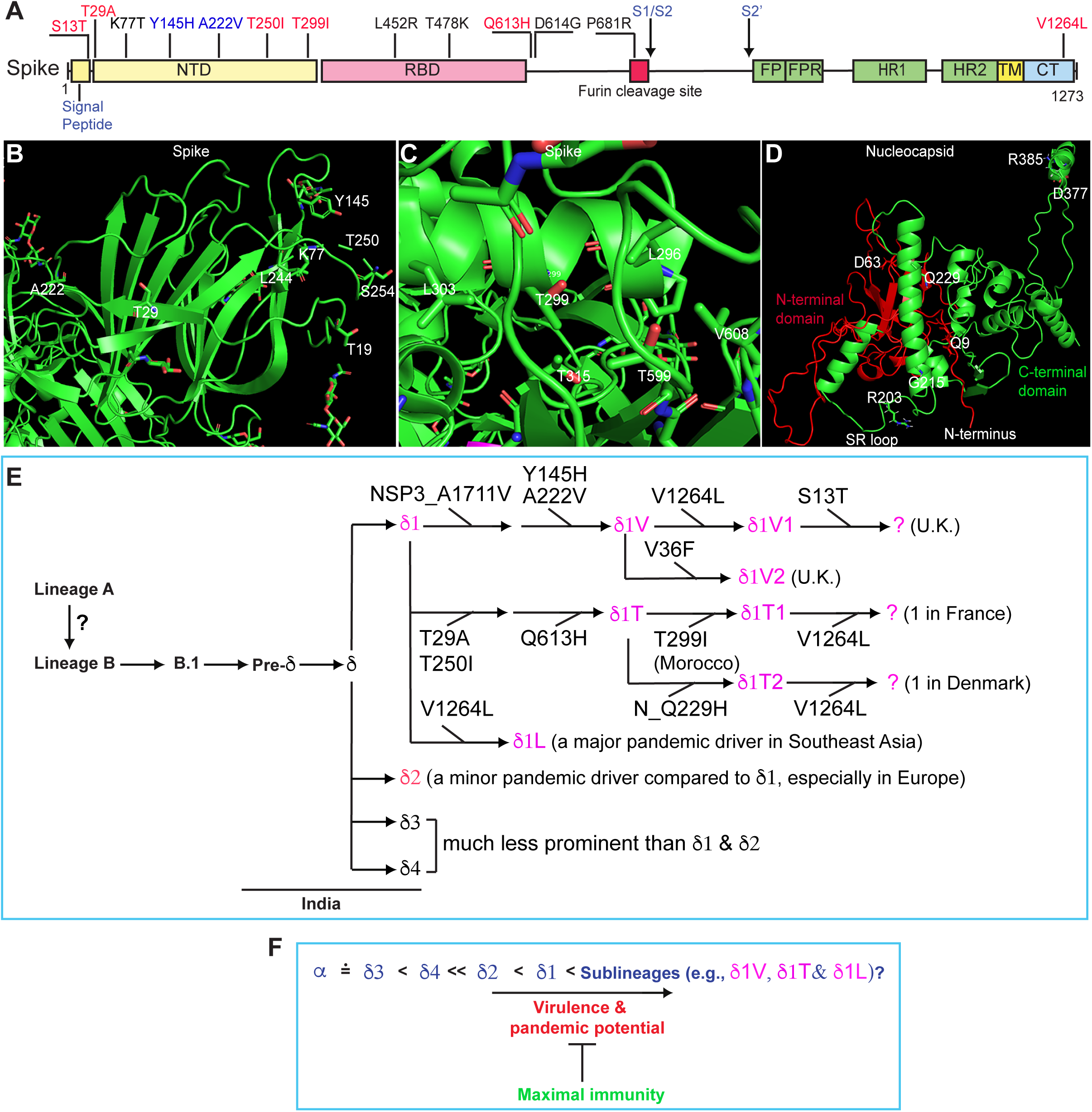
Mechanistic impact of different substitutions detected in δ subvariants and a hypothetical model. (**A**) Domain organization of spike protein. Some key substitutions are shown in black and new substitutions described herein are in red. NTD, N-terminal domain; RBD, receptor-binding domain; S1/S2, boundary of S1 and S2 domains after furin cleavage between residues R685 and S686; FP, fusion peptide; FPR, fusion peptide C-terminal proximal region; HR1 and HR2, heptad-repeat regions 1 and 2, respectively; S2’, cleavage site between residues R815 and S816 in S2 domain; TM, transmembrane motif; CT, C-terminal cytoplasmic tail. The domain organization was adapted from a published study [18]. (**B**) Structural details of spike T19, T29, K77, Y145, A222 and T250. These residues are within a super antigenic site in the N-terminal domain (Fig. 8A) [19]. T250 is located within an unstructured loop marked with broken lines at the right. (**C**) Structural details of spike T299 and its neighboring residues. The structural models (B-C) were adapted from PyMol presentation of the spike protein structure 6XR8 from the PDB database. (**D**) Structure of nucleocapsid protein of SARS-COV-2. The entire nucleocapsid structure is based on PyMol presentation of a structural model, QHD43423.pdb (https://zhanglab.ccmb.med.umich.edu/COVID-19/), built from crystal structures of the N- and C-terminal RNA-binding domains of nucleocapsid protein (6M3M and 6YUN from the PDP database, respectively) by Dr. Yang Zhang’s group at University of Michigan. Structure of the SR (serine/arginine-rich) domain is hypothetical. (**E**) Cartoon summarizing different subvariants described herein. (**F**) Relative virulence of different δ subvariants in comparison to α variant. Alarmingly, multiple δ1 subvariants appear to be even more virulent than itself.

Q613 is adjacent to D614 (Fig. 8A). D614G is a key substitution that defines the entire course of the pandemic [20-22]. This substitution alters conformational state of the spike protein [12], so Q613H may synergize with D614G. V1264 is located at the C-terminal cytoplasmic tail of spike protein (Fig. 8A) and V1264L confers an acidic di-leucine motif in the cytoplasmic tail of spike protein for its cytoplasmic traffic and processing [10]. Alarmingly, V1264L is a signature substitution of a δ1 sublineage that drives the pandemic in Indonesia, Singapore and Malaysia [10].

As shown in Fig. 8D, nucleocapsid protein is composed of an N-terminal tail, an N-terminal RNA-binding domain, a middle SR (serine/arginine-rich) loop, a C-terminal RNA-binding domain and a C-terminal tail. The N-terminal tail is a known immune epitope, so Q9L in a subset of δ1 variant may confer immune evasion. D63G is a signature substitution of δ1 variant. D63 is located within the N-terminal domain (Fig. 8D) and directly involved in RNA binding. D63G should affect this binding [23], so it is puzzling why δ1 variant has acquired this substitution. It is possible maximal RNA binding is not optimal for viral fitness.

The SR loop is highly phosphorylated [24] and may regulate the nucleocapsid protein level and function. In addition to δ variant, this loop is altered in the three other variants of concern. R203K and G204R are present in α and β variants, whereas T205I is associated with γ variant [5-7]. Similarly, R203M, a signature substitution of δ variant, alters the SR loop (Fig. 8D). Moreover, δ1V2 possesses S202N (Fig. S4D), recurrent in other SARS-COV-2 variants. S202 is phosphorylated [24], so S202N blocks phosphorylation. Thus, S202N may synergize with R203M in altering phosphorylation of the SR loop. In addition to this new substitution, δ1V2 has acquired spike V36F and A222V, both of which are alarming (Fig. S4C). Thus, it is important to watch out this new δ1 sublineage.

Nucleocapsid protein also possesses sequence elements for subcellular trafficking. Residues 219-235 are leucine-rich and may serve as a nuclear export signal [25]. This segment is part of a long α-helix and G215 is at the N-terminal end of this helix (Fig. 8D). Because glycine is often a helix breaker, G215C in δ1 subvariant (Table 1) should improve the structure of this helix. Moreover, Q229H from δ1V1 sublineage (Table 1) is also located in this helix and may thus synergize with G215C in improving the stability of this helix. In addition to its nuclear export signal, nucleocapsid protein possesses a bipartite nuclear localization signal spanning residues 369-389. Related to this, the substitutions D377Y and R385K are within this region (Fig. 8D). Thus, alteration of nucleocapsid trafficking in a common strategy that δ1 adopts to improve its viral fitness.

In addition to spike and nucleocapsid proteins, NSP3 is a frequent target for alteration in different δ subvariants (Table 1). NSP3 is a large multidomain protein known to interact with nucleocapsid during viral replication [26]. Moreover, a recent study revealed that NSP3 forms molecular pore spanning the double membrane of the coronavirus replication organelle [27]. A488S may improve interaction with K486, P822L may enhance interaction in a hydrophobic core and P1469S may facilitate interaction with M1441 and Y1574 (according to QHD43415; Dr. Yang Zhang’s lab, University of Michigan, USA). P1228 and A1711 are on the potential antigenic surface of the protein, so their alterations may confer immune evasion. Therefore, in addition to modifying spike protein, δ variant has taken diverse strategies to finetune functions of nucleocapsid and NSP3 for improving its viral fitness.

### A hypothetical model on SARS-COV-2 evolution and relative virulence of δ subvariants

Shown in Fig. 8E is a continuously branching model illustrating evolutionary relationship among δ subvariants described herein. According to this model, δ variant yields different subvariants, which in turn evolve and generate additional lineages continuously. Host environment (including natural immunity, vaccination and experimental drug treatment) then select a specific set of such lineages, which then spread and drive the pandemic. Such a model can also explain evolution of SARS-COV-2 in general. This model also suggests that there are simple rules this virus adopts to improve its fitness during evolution. Identification and elucidation of such rules will help us predict its evolutionary trajectory, which will in turn give us a winning chance to get ahead of the virus (instead of tailgating it) in this evolutionary game. This may be the most effective way to control and end this pandemic.

A companion study of SARS-COV-2 genomes from the USA supports a similar model in which δ variant drives the pandemic there mainly through δ1 and δ2 subvariants [9]. The subvariant δ1L encodes spike V1264L as an extra substitution (Fig. 8A) and is based on a third study about this sublineage in Indonesia, Singapore, Malaysia and East Timor [10]. The continuously branching model also appears true in many other countries, including Canada, Mexico, Israel, Singapore, Australia, New Zealand, Japan, South Korea, Vietnam and China, although the subvariant composition varies from country to country. As it is in Europe (Figs 5-6), δ1 is now predominant in Singapore [10], Israel, Australia, New Zealand and Japan, but δ2 is more dominant than δ1 subvariant in South Korea, Vietnam and some Canadian provinces (unpublished observations). Thus, it is necessary to systematically track δ subvariants and their evolutionary trajectory in different countries around the world.

As summarized in Fig. 8F, δ1 and δ2 subvariants are much more virulent than α variant, which explains why δ variant has overtaken all other variants and become almost the sole pandemic driver around the world. Alarmingly, δ1 sublineages such as δ1L, δ1V1, δ1V2, δ1T1 and δ1T2 (Fig. 8E) appear to be even more virulent than δ1 itself. Therefore, we will need to track SARS-COV-2 evolution very closely and map its evolutionary trajectory in a precise and timely fashion, which will help us adopt the best public health measures and develop the new generation of vaccines. Dynamic evolution of δ variant (Fig. 8E-F) also reiterates the notion that to end this pandemic swiftly, it will be important to take preventive measures (such as immunity maximization through full vaccination and subsequent boosters) to block further evolution of δ subvariants.

## Supporting information

Acknowledgement table on the GISAID genomes used in this study

Acknowledgement table on the GISAID genomes used in this study

Acknowledgement table on the GISAID genomes used in this study

Acknowledgement table on the GISAID genomes used in this study

Acknowledgement table on the GISAID genomes used in this study

Acknowledgement table on the GISAID genomes used in this study

Acknowledgement table on the GISAID genomes used in this study

Acknowledgement table on the GISAID genomes used in this study

Acknowledgement table on the GISAID genomes used in this study

Acknowledgement table on the GISAID genomes used in this study

Acknowledgement table on the GISAID genomes used in this study

Acknowledgement table on the GISAID genomes used in this study

Acknowledgement table on the GISAID genomes used in this study

Acknowledgement table on the GISAID genomes used in this study

Acknowledgement table on the GISAID genomes used in this study

Acknowledgement table on the GISAID genomes used in this study

## Data Availability

All data produced in the present study are available upon reasonable request to the authors

## ACKNOWLEDGEMENT

I gratefully acknowledge the GISAID database for diligent maintenance SARS-COV-2 genomes and numerous investigators for the valuable genome sequences used in this work (see the supplementary section for details). I am also grateful to Professor Federico M. Giorgi at University of Bologna, Italy, for the generosity to allow timely access to the Coronapp server. This work was supported by funds from Canadian Institutes of Health Research (CIHR), Natural Sciences and Engineering Research Council of Canada (NSERC) and Compute Canada (to X.J.Y.).

## MATERIALS AND METHODS

### SARS-COV-2 sequence files, mutation profiling and phylogenetic analysis

SARS-COV-2 genome sequences were downloaded the GISAID database (https://www.gisaid.org/) on the dates specified in the figure legends. CoVsurver (https://www.gisaid.org/epiflu-applications/covsurver-mutations-app/) was used to analyze mutations in representative genomes. Fasta files containing specific groups of genomes were downloaded from the GISAID database. During downloading, empty spaces in the fasta headers were replaced by underscores because such spaces make the files incompatible for subsequent sequence alignment and phylogenetic analysis. To shorten the Fasta headers, the Find/Replace_All function of TextEdit (version 1.16) was used to delete “Cov19/” and “/2021” at the beginning and middle of the headers, respectively. In addition, “/” and “|” symbols in the headers are incompatible for sequence alignment and phylogenetic analysis, so they were also replaced with underscores via the Find/Replace_All function of TextEdit. The cleaned Fasta files were used for mutation profiling via Coronapp (http://giorgilab.unibo.it/coronannotator/), a web-based mutation annotation application [16,17].

Some cleaned Fasta files were also uploaded to SnapGene (version 5.3.2) for multisequence analysis via the MAFFT tool. For this, only high-coverage genomes with complete sample collection date information were used. After the alignment, the 5’ and 3’ were manually trimmed to make >95% genomes possess the same length. The sequence alignment was exported out as Fasta files. Each Fasta header in the files contains “(0 bp)” along with some adjacent empty spaces, which are incompatible for subsequent phylogenetic analysis and were thus deleted via the Find/Replace_All function of TextEdit. Each file was transferred to a folder for RAxML-NG version 0.9.0 [28] for analysis via the Terminal mode on the Mac computer. The file was further cleaned via the command line: raxml-ng --check --msa file_name.fa --model GTR+G. This generated a file (file_name.fa.raxml.reduced.phy) for phylogenetic analysis via the following command line: raxml-ng --msa file_name.fa.raxml.reduced.phy --model GTR+G –prefix file_name --threads 2 --seed 2. In the command lines, no spaces, hyphens, special symbols or punctuation marks (except for underscores) are allowed. The phylogenetic analysis generated 20 maximum likelihood trees and one bestTree. FigTree V1.4.4 was used to open the 21 tree files for manual inspection, analysis and annotation. To guide the inspection, representative genomes were analyzed in batches to display mutations via CoVsurver (https://www.gisaid.org/epiflu-applications/covsurver-mutations-app/). In addition, results from mutation profiling via Coronapp [16] also served as a guide in manual inspection of the 21 trees. Furthermore, the time of case identification was also considered to choose a potential root. With these factors all considered, an optimal tree was selected from the 21 trees for re-rooting via FigTree (https://github.com/rambaut/figtree/releases/tag/v1.4.4). The resulting tree was exported for image processing via Adobe Photoshop and subsequent presentation through Illustrator.

### Defining different variant genomes using various markers

α, β, γ and δ variant genomes were downloaded from the GISAID database as defined by the server. To identify δ subvariant genomes in the database, nucleocapsid substitutions G215C and R385K (Table 1) were used as markers for δ1 or δ3 genomes, respectively. Spike substitutions A222V and K77T were used as markers for δ2 or δ4 genomes, respectively. In Europe, there are many δ1V genomes that also encode spike A222V, so the NSP3 substitution P822L was used together with spike A222V to identify δ2 genomes for analysis or downloading. There are several limitations with these markers. The first limitation is that some genomes do not have the right substitutions due to poor sequence quality. Another limitation is that some genomes may have incorrect date information on sample collection due to typos and other errors (for example, a genome collected in 2021 was submitted as one from 2020). A third limitation is that a few δ1 or δ2 genomes also encode K77T or R395K, so the number of δ3 or δ3 may be overestimated in some cases. However, these limitations should not affect the overall conclusions.

### PyMol structural modeling

The PyMol molecular graphics system (version 2.4.2, https://pymol.org/2/) from Schrödinger, Inc. was used for downloading structure files from the PDB database for further analysis and image export. The images were cropped via Adobe Photoshop and further presentation using Illustrator.

### Pandemic and vaccination data

Pandemic and vaccination data were downloaded from the Our World in Data website (https://ourworldindata.org/explorers/coronavirus-data-explorer?zoomToSelection=true&time=2020-03-01..latest&facet=none&pickerSort=desc&pickerMetric=total_cases&Metric=Confirmed+cases&Interval=Cumulative&Relative+to+Population=false&Align+outbreaks=false&country=~OWID_WRL) as a .csv file for further processing via Excel and figure generation through Prism 9.0 and Adobe Illustrator.

## SUPPLEMENTAL INFORMATION

This section includes 6 Supplementary Figures and 16 acknowledgement tables for the GISAID genomes used in this work.

## SUPPLEMENTAL FIGURE LEGENDS

**Figure S1.**
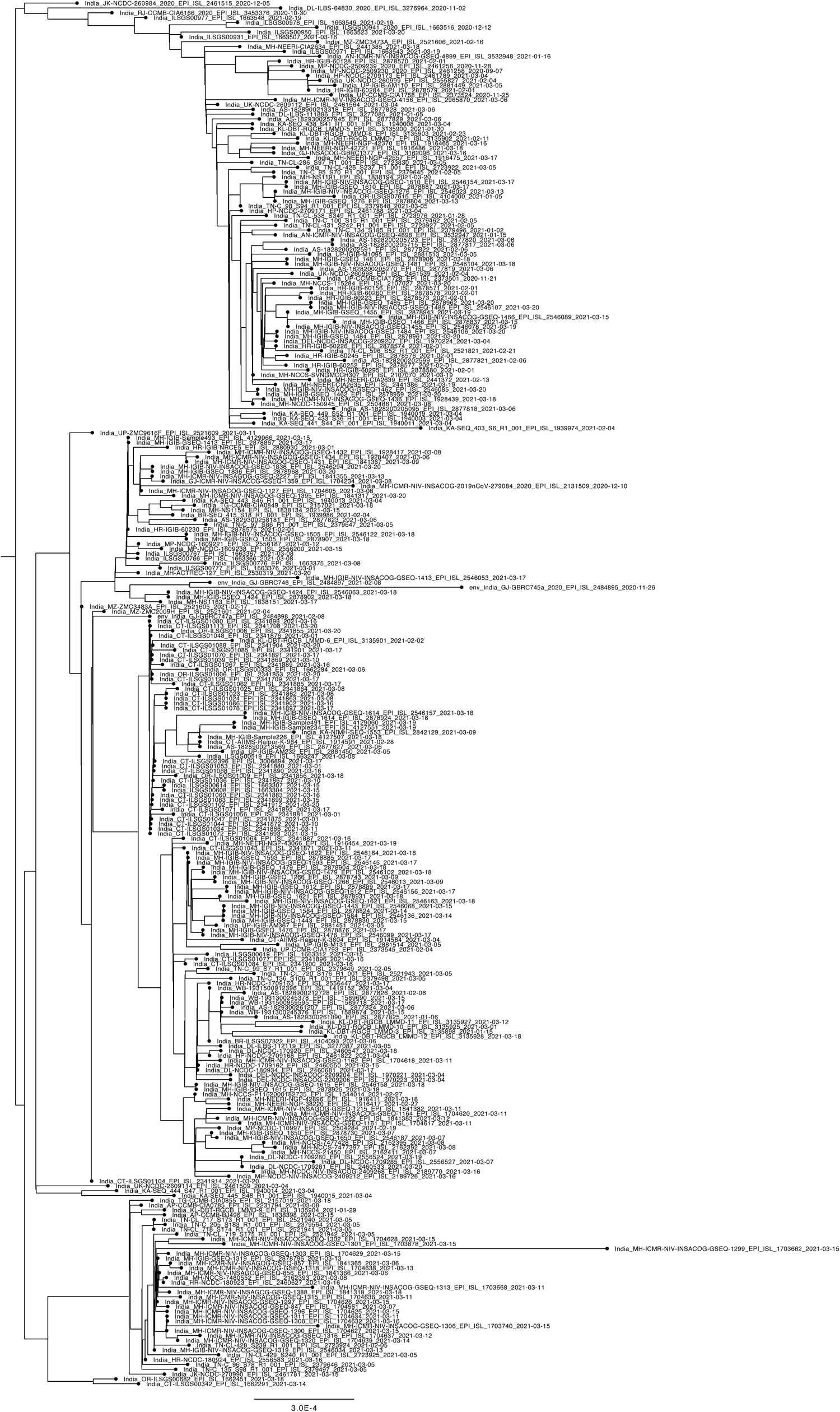
Phylogenetic analysis of 291 δ-genomes identified in India by March 21, 2021. The genomes were downloaded from the GISAID SARS-COV-2 genome sequence database on September 14, 2021 for phylogenetic analysis. Only high-coverage genomes with complete date information on sample collection were used. The package RAxML-NG was used to generate 20 maximum likelihood trees and the bestTree for presentation via Figtree as in Fig. 3B.

**Figure S2.**
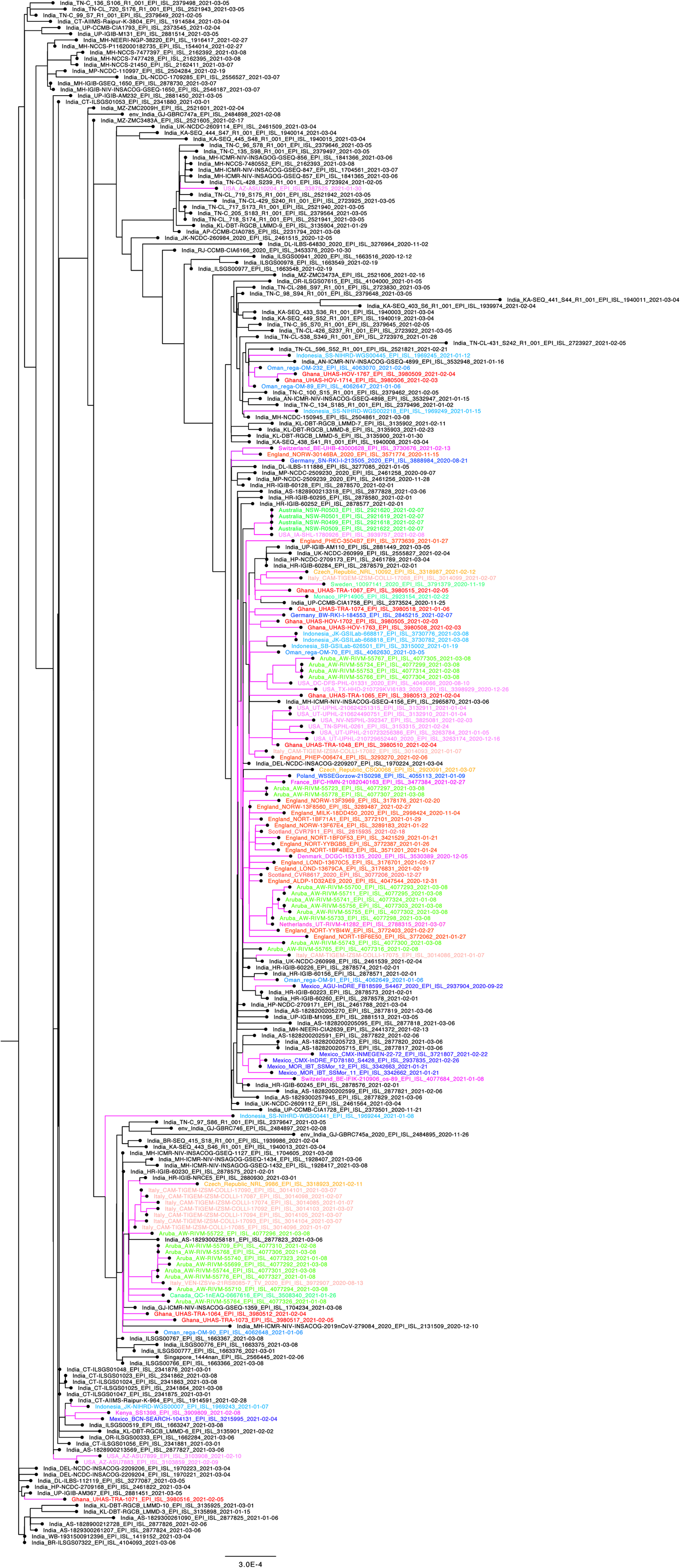
Phylogenetic analysis of 264 δ-genomes identified by March 09, 2021. The genomes were downloaded from the GISAID SARS-COV-2 genome sequence database on September 14, 2021 for phylogenetic analysis. Only high-coverage genomes with complete sample collection date information were used. The package RAxML-NG was used to generate 20 maximum likelihood trees and the bestTree for presentation via Figtree as in Fig. 4A.

**Figure S3.**
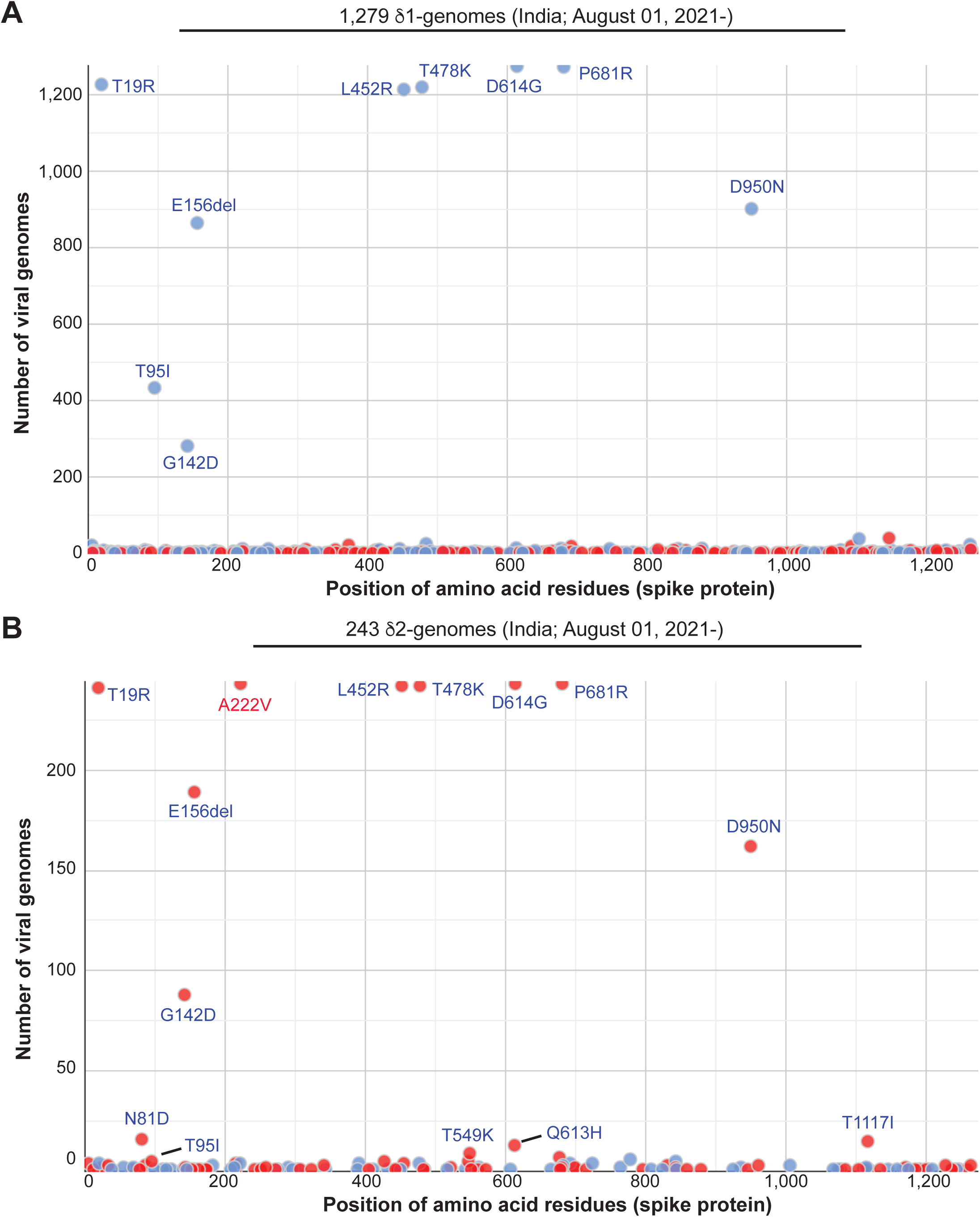
Mutation profile of δ genomes from India. δ1 (A) and δ2 (B) genomes were downloaded from the GISAID database on September 28, 2021 for mutation profiling via Coronapp as in Fig. 3E-F. Shown here are substitutions in spike protein. One major difference is that δ2 but not δ1 genomes encode A222V. Another difference is that ∼40% δ1 genomes from India encode T95I, but this percentage is very low among δ2 genomes. The percentage of T95I-encoding δ1 genomes varies from country to country.

**Figure S4.**
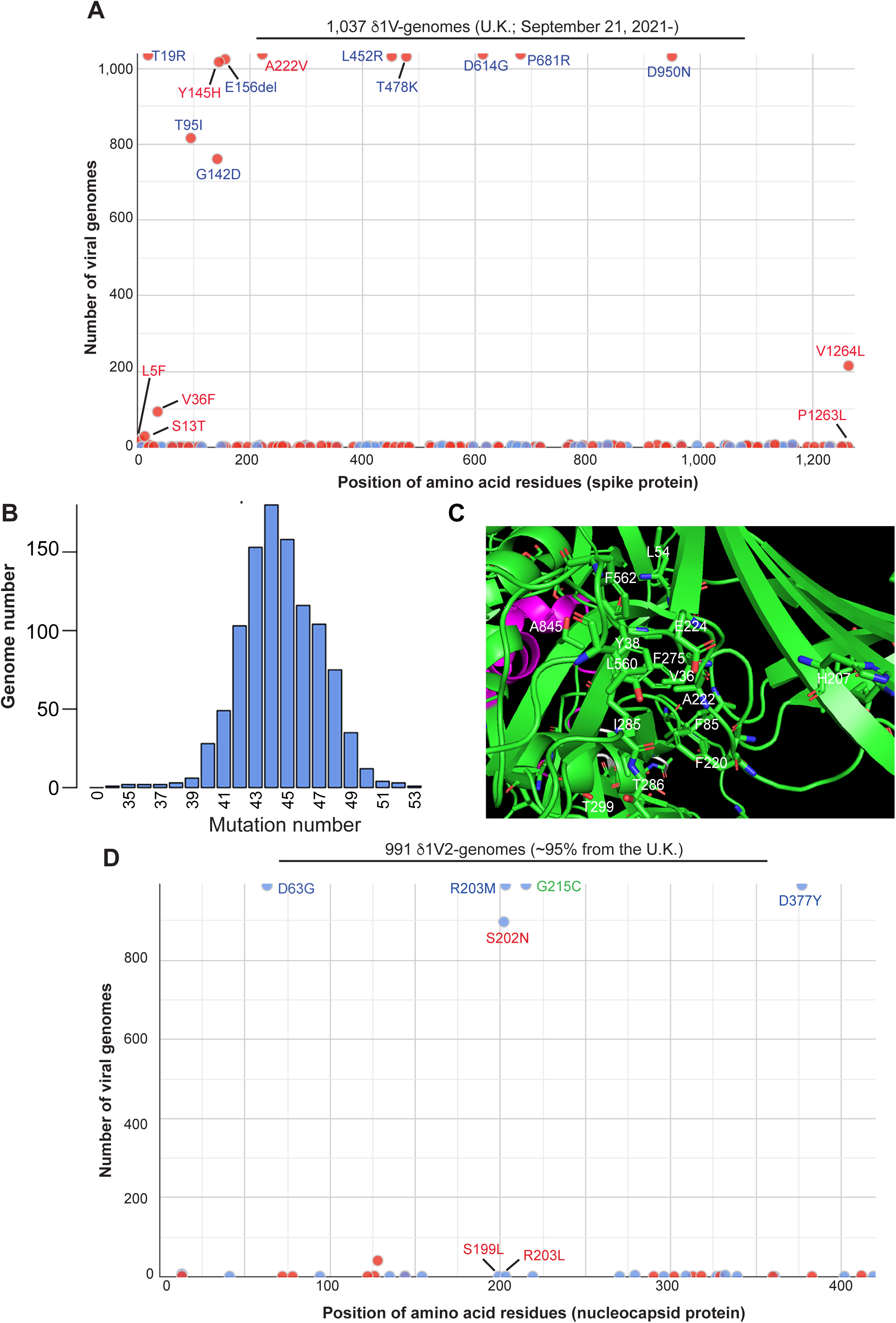
Analysis of δ1V genomes. (**A-B**) Mutation profile of 1,037 δ1V-genomes from the U.K. The genomes were downloaded from the GISAID database on October 02, 2021 for mutation profiling via Coronapp. Shown here are substitutions in spike protein (A) and the mutation load in the genomes (B). The substitution L5F is present in a group of genomes different from δ1V1 and δ1V2, whereas S13T is associated with a subset of δ1V1 genomes. (**C**) Structural details of spike V36, A222 and their neighboring residues. The side chain of V36 is 3.4 Å away from that of A222. They are part of a cleft with an extensive hydrophobic interaction network. In addition to L54, L560 and I285, there are 5 aromatic residues (F562, Y38, F275, F85 and F200) from the top to the bottom of the panel. At the bottom left is T286, which is 20.9 Å away from T299 outside the cleft. T299 is replaced by isoleucine in δ1T1 (Fig. 8E). This structural model was adapted from PyMol presentation of the spike protein structure 6XR8 from the PDB database. (**D**) Mutation profile of 991 δ1V2 genomes. The genomes were downloaded from the GISAID database in the morning of October 04, 2021 for mutation profiling via Coronapp. Shown here are substitutions in nucleocapsid protein. The average mutation load is 45 per genome (data not shown).

**Figure S5.**
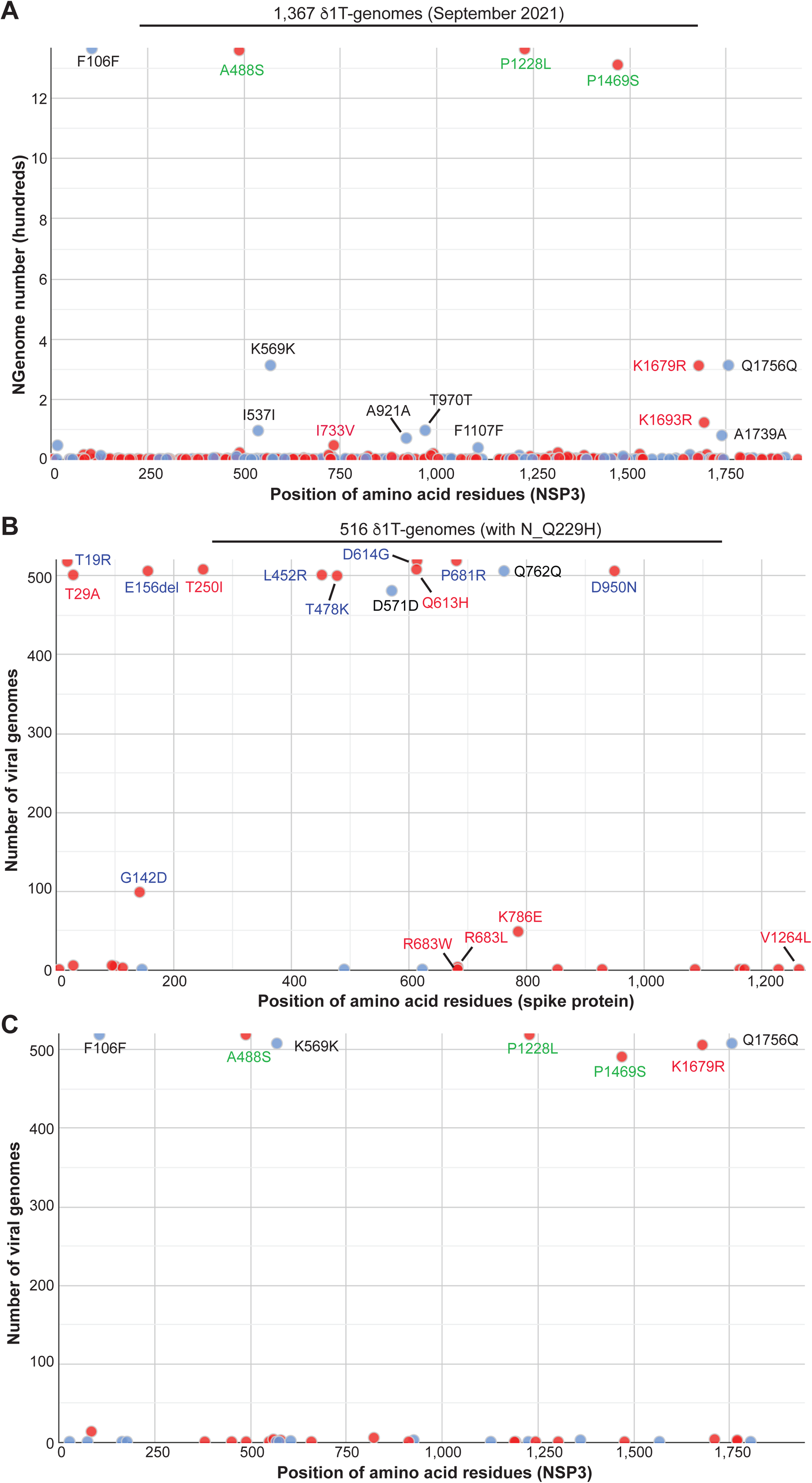
Mutation profile of δ1-genomes from Italy (A), France (B) and Germany (C). The genomes were downloaded from the GISAID SARS-COV-2 genome sequence database on September 30, 2021 for mutation profiling via Coronapp. Shown here are substitutions in spike protein.

**Figure S6.**
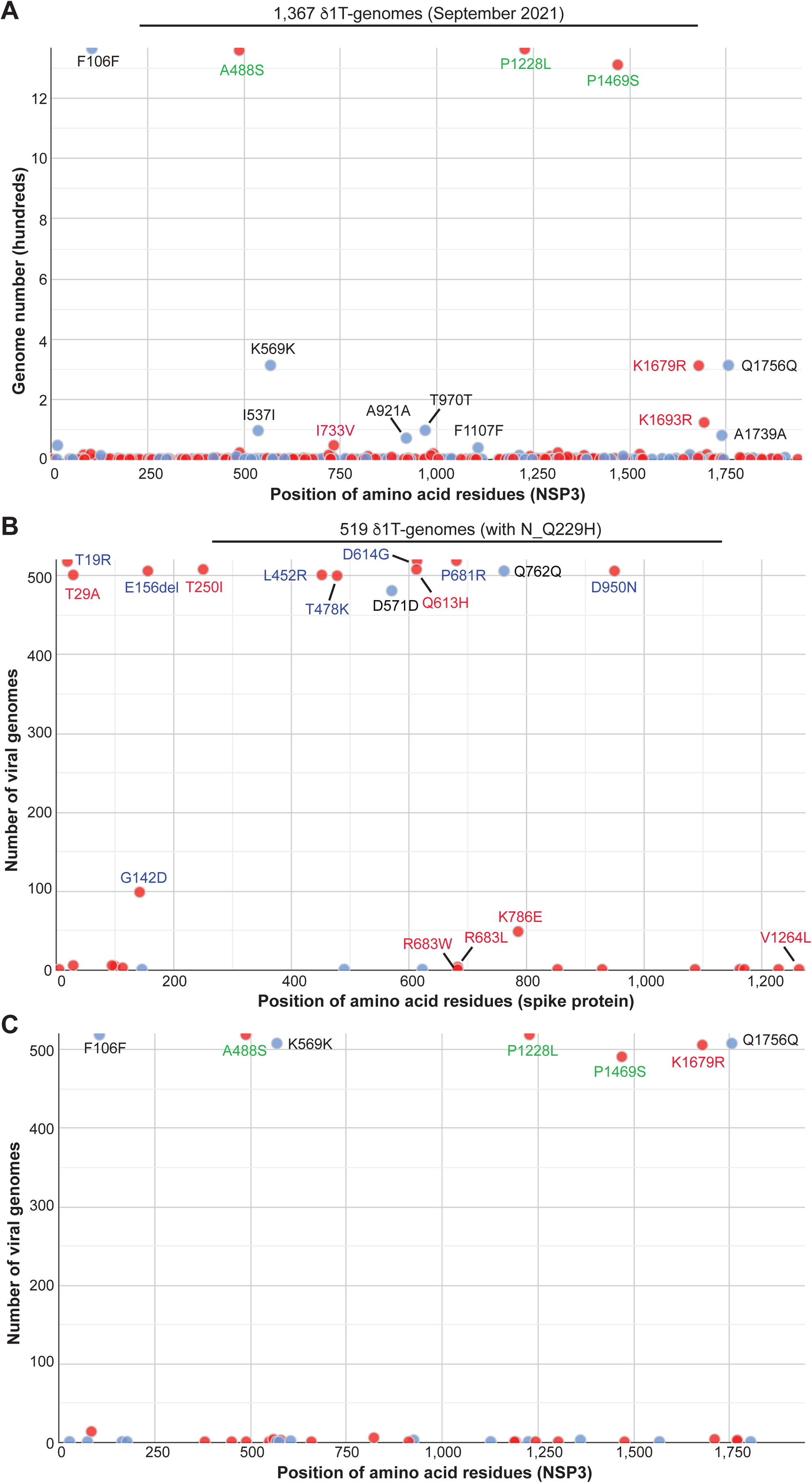
Mutational analysis of δ1T and δ1T1 genomes. (**A**) Mutation profile of 1,367 δ1T genomes from different countries around the world. The genomes were downloaded from the GISAID database on October 01, 2021 for mutation profiling via Coronapp as in Fig. 7B-C. NSP3 substitutions are shown here. (**B-C**) Mutation profile of 519 δ1T1 genomes. The genomes, mainly identified in Denmark, were downloaded from the GISAID SARS-COV-2 genome sequence database on October 01, 2021 for mutation profiling via Coronapp. Shown in (B) and (C) are substitutions in spike and NSP3 proteins, respectively.

