## Supplementary material for "SARS-COV-2 δ variant drives the pandemic in India and Europe via two subvariants": Acknowledgement table on the GISAID genomes used in this study

We gratefully acknowledge the following Authors from the Originating laboratories responsible for obtaining the specimens, as well as the Submitting laboratories where the genome data were generated and shared via GISAID, on which this research is based.

All Submitters of data may be contacted directly via www.gisaid.org

Authors are sorted alphabetically.

| Accession ID | Originating Laboratory | Submitting Laboratory | Authors |
|---|---|---|---|
| EPI_ISL_2502639 | All India Institute of Medical Sciences Delhi Hospital | Virology Laboratory, AIIMS Delhi | Aashish Choudhary; Chitra Sarkar; Deepankar Srigyan; Dibyabhaba Pradhan; Jyoti Jethani; Lalit Dar; Lata Rani; Manish Soneja; Megha Brijwal; Nazneen Arif; Pooja Pandey; Puneet Kaur; Rakesh Lodha; Randeep Guleria; Ritu Gupta; Shivram Dhakad; Subrata Sinha; Sumedha Bagga |
| EPI_ISL_910112, EPI_ISL_910123, EPI_ISL_910124, EPI_ISL_910125, EPI_ISL_910126, EPI_ISL_910127, EPI_ISL_910128, EPI_ISL_910129, EPI_ISL_910130, EPI_ISL_910131, EPI_ISL_910132, EPI_ISL_910133, EPI_ISL_910134, EPI_ISL_910135, EPI_ISL_910136, EPI_ISL_910137, EPI_ISL_910138, EPI_ISL_910139, EPI_ISL_910140, EPI_ISL_910141, EPI_ISL_910142, EPI_ISL_910143, EPI_ISL_910144, EPI_ISL_910145, EPI_ISL_910146, EPI_ISL_910147, EPI_ISL_910148, EPI_ISL_910149, EPI_ISL_910150, EPI_ISL_910151, EPI_ISL_910152, EPI_ISL_910153, EPI_ISL_910154, EPI_ISL_910155, EPI_ISL_910156, EPI_ISL_910157, EPI_ISL_910158, EPI_ISL_910159, EPI_ISL_910160, EPI_ISL_910161, EPI_ISL_910162, EPI_ISL_910163, EPI_ISL_910164, EPI_ISL_910165, EPI_ISL_910166, EPI_ISL_910167, EPI_ISL_910168, EPI_ISL_910169, EPI_ISL_910170, EPI_ISL_910171, EPI_ISL_910172, EPI_ISL_910173, EPI_ISL_910174, EPI_ISL_910175, EPI_ISL_910176, EPI_ISL_910177, EPI_ISL_910178, EPI_ISL_910179, EPI_ISL_910180, EPI_ISL_910181, EPI_ISL_910182, EPI_ISL_910183, EPI_ISL_910184, EPI_ISL_910185, EPI_ISL_910186, EPI_ISL_910187, EPI_ISL_910188, EPI_ISL_910189, EPI_ISL_910190, EPI_ISL_910191, EPI_ISL_910192, EPI_ISL_910193, EPI_ISL_910194, EPI_ISL_910195, EPI_ISL_910196, EPI_ISL_910197, EPI_ISL_910198, EPI_ISL_910199, EPI_ISL_910200, EPI_ISL_910201, EPI_ISL_910202, EPI_ISL_910203, EPI_ISL_910204, EPI_ISL_910205, EPI_ISL_910206, EPI_ISL_910207, EPI_ISL_910208, EPI_ISL_910209, EPI_ISL_910210, EPI_ISL_910211, EPI_ISL_910212, EPI_ISL_910213, EPI_ISL_910214, EPI_ISL_910215, EPI_ISL_910216, EPI_ISL_910217, EPI_ISL_910218, EPI_ISL_910219, EPI_ISL_910220, EPI_ISL_910221, EPI_ISL_910223, EPI_ISL_910224, EPI_ISL_910225, EPI_ISL_910226, EPI_ISL_910227, EPI_ISL_910228, EPI_ISL_910229, EPI_ISL_910230, EPI_ISL_910231, EPI_ISL_910232, EPI_ISL_910233, EPI_ISL_910234, EPI_ISL_910235, EPI_ISL_910236, EPI_ISL_910237, EPI_ISL_910238, EPI_ISL_910239, EPI_ISL_910240, EPI_ISL_910241, EPI_ISL_910242, EPI_ISL_910243, EPI_ISL_910244, EPI_ISL_910245, EPI_ISL_910246, EPI_ISL_910247, EPI_ISL_910248, EPI_ISL_910249, EPI_ISL_910250, EPI_ISL_910251, EPI_ISL_910252, EPI_ISL_910253, EPI_ISL_910254, EPI_ISL_910255, EPI_ISL_910256, EPI_ISL_910257, EPI_ISL_910258, EPI_ISL_910259, EPI_ISL_910260, EPI_ISL_910261, EPI_ISL_910262, EPI_ISL_910263, EPI_ISL_910264, EPI_ISL_910265, EPI_ISL_910266, EPI_ISL_910267, EPI_ISL_910268, EPI_ISL_910269, EPI_ISL_910270, EPI_ISL_910271, EPI_ISL_910272, EPI_ISL_910273, EPI_ISL_910274, EPI_ISL_910275, EPI_ISL_910276, EPI_ISL_910277, EPI_ISL_910278, EPI_ISL_910279, EPI_ISL_910280, EPI_ISL_910281, EPI_ISL_910282, EPI_ISL_910283, EPI_ISL_910284, EPI_ISL_910285, EPI_ISL_910286, EPI_ISL_910287, EPI_ISL_910288, EPI_ISL_910289, EPI_ISL_910290, EPI_ISL_910291, EPI_ISL_910292, EPI_ISL_910293 | | | |
| see above | CSIR-Centre for Cellular and Molecular Biology | CSIR-Centre for Cellular and Molecular Biology | Archana Bharadwaj Siva; B Himasri; Blessy B John; Divya Tej Sowpati; Karthik Bharadwaj Tallapaka; Lamuk Zaveri; Namami Gaur; Payel Mukherjee; Pratheusa Maccha; Priya Singh; Purushotham Vodnala; Rakesh K Mishra; Sofia Banu; Tulasi Nagabandi; Viswagithe S L |
| EPI_ISL_569859, EPI_ISL_569860, EPI_ISL_569861, EPI_ISL_569862, EPI_ISL_578080, EPI_ISL_578081, EPI_ISL_578168, EPI_ISL_578175, EPI_ISL_578178, EPI_ISL_578179, EPI_ISL_578180, EPI_ISL_578181, EPI_ISL_578182, EPI_ISL_578183, EPI_ISL_578184 | | | |
| see above | CSIR-Indian Institute of Chemical Biology, MEDICA Supercpecialty Hospital Kolkata | CSIR-Indian Institute of Chemical Biology, MEDICA Supercpecialty Hospital Kolkata | Abhishake Lahiri; Debaleena Bhowmik; Dr. Partha Chakrabarti; Dr. Arpita Ghosh Mitra; Dr. Aviral Roy; Dr. Aviral Roy; Dr. AviralRoy; Dr. Partha Chakrabarti; Dr. Rajesh Pandey; Dr. Saikat Chakrabarti; Dr. Sandip Paul; Dr. Soumen Saha; Dr.Partha Chakrabarti; Priyanka Mallick; Sujay Krishna Maity |
| EPI_ISL_2714634, EPI_ISL_2714635, EPI_ISL_2714636, EPI_ISL_2714637, EPI_ISL_2714638, EPI_ISL_2714639, EPI_ISL_2714640, EPI_ISL_2714641, EPI_ISL_2714642, EPI_ISL_2714643, EPI_ISL_2714699, EPI_ISL_2714700 | | | |
| see above | Communicable Diseases, Interactive Research School for Health Affairs (IRSHA) | Communicable Diseases, Interactive Research School for Health Affairs (IRSHA) | A.C.; Arankalle; Mishra; Modak, M.; Pisal; S.R.; Shrivastava, S.; V.A.; Virkar, R. |
| EPI_ISL_4392854 | Dr. Ishaan Gupta lab, Department of Biochemical Engineering and Biotechnology, Block I, Indian Institute of Technology Delhi | Dr. Ishaan Gupta lab, Department of Biochemical Engineering and Biotechnology, Block I, Indian Institute of Technology Delhi | Animesh Ray; Anjan Trikha; Anshul Budhraja; Anubhav Basu; Aruna Nambirajan; Atish Gheware; Chitra Sarkar; Dasari Abhilash; Deepali Jain; Ishaan Gupta; Madhuresh Sumit; Naveet Wig; Purva Mathur; Randeep Guleria; Ritu Gupta; S Arulselvi; Sachin Kumar; Seesandra Rajagopala; Suman Pakala |
| EPI_ISL_4392851 | Dr. Ishaan Gupta lab, Department of Biochemical Engineering and Biotechnology, Block I, Indian Institute of Technology Delhi | Dr. Ishaan gupta lab, Department of Biochemical Engineering and Biotechnology, Block I, Indian Institute of Technology Delhi | Animesh Ray; Anjan Trikha; Anshul Budhraja; Anubhav Basu; Aruna Nambirajan; Atish Gheware; Chitra Sarkar; Dasari Abhilash; Deepali Jain; Ishaan Gupta; Madhuresh Sumit; Naveet Wig; Purva Mathur; Randeep Guleria; Ritu Gupta; S Arulselvi; Sachin Kumar; Seesandra Rajagopala; Suman Pakala |
| EPI_ISL_4392853 | Dr. Ishaan gupta lab, Department of Biochemical Engineering and Biotechnology, Block I, Indian Institute of Technology Delhi | Dr. Ishaan gupta lab, Department of Biochemical Engineering and Biotechnology, Block I, Indian Institute of Technology Delhi | Animesh Ray; Anjan Trikha; Anshul Budhraja; Anubhav Basu; Aruna Nambirajan; Atish Gheware; Chitra Sarkar; Dasari Abhilash; Deepali Jain; Ishaan Gupta; Madhuresh Sumit; Naveet Wig; Purva Mathur; Randeep Guleria; Ritu Gupta; S Arulselvi; Sachin Kumar; Seesandra Rajagopala; Suman Pakala |
| EPI_ISL_586569, EPI_ISL_586570 | GCRI,Ahmedabad | Gujarat Biotechnology Research Centre | A M Kadri; Afzal Ansari; Apurvasinh Puvar; Chaitanya Joshi; Dinesh Kumar; Harsh Bakshi; Harsha Panchal; Janvi Raval; Komal Patel; Labdhi Pandya; Madhvi Joshi; Maharshi Pandya; Monika Gandhi; Nidhi Patel; Nikha Trivedi; Nitin Savaliya; Pinal Trivedi; R D Dixit; Raghawendra Kumar; Shashank Pandya; Zarna Patel; Zuber Saiyed |
| EPI_ISL_2674194 | Government Institute of Medical Sciences, Greater Noida | CSIR Institute of Genomics and Integrative Biology | Abhinav Jain; Bani Jolly; Disha Sharma; Gyan Ranjan; Mohamed Imran; Mohit Kumar Divakar; Paras Sehgal; Rahul C Bhoyar; Rakesh Gupta; Rashmi Upadhayay; Saurabh Srivastava; Sridhar Sivasubbu; Vinod Scaria; Vivek Gupta |
| EPI_ISL_700218, EPI_ISL_700219, EPI_ISL_700220, EPI_ISL_700221, EPI_ISL_700222, EPI_ISL_700223, EPI_ISL_700224, EPI_ISL_700225, EPI_ISL_700226, EPI_ISL_700227, EPI_ISL_700228, EPI_ISL_700229, EPI_ISL_700230, EPI_ISL_700231, EPI_ISL_700232, EPI_ISL_700233, EPI_ISL_700234, EPI_ISL_700235, EPI_ISL_700236, EPI_ISL_700237, EPI_ISL_700238, EPI_ISL_700239, EPI_ISL_700240, EPI_ISL_700241, EPI_ISL_700242, EPI_ISL_700243, EPI_ISL_700244, EPI_ISL_700245, EPI_ISL_700246, EPI_ISL_700247, EPI_ISL_700248, EPI_ISL_700249, EPI_ISL_700250, EPI_ISL_700251, EPI_ISL_700252, EPI_ISL_700253, EPI_ISL_700254, EPI_ISL_700255, EPI_ISL_700256, EPI_ISL_700257, EPI_ISL_700258, EPI_ISL_700259, EPI_ISL_700260, EPI_ISL_700261, EPI_ISL_700262, EPI_ISL_700263, EPI_ISL_700264, EPI_ISL_700265, EPI_ISL_700266, EPI_ISL_700267, EPI_ISL_700268, EPI_ISL_700269, EPI_ISL_700270, EPI_ISL_700271, EPI_ISL_700272, EPI_ISL_700273, EPI_ISL_700274, EPI_ISL_700275, EPI_ISL_700276, EPI_ISL_700277, EPI_ISL_700278, EPI_ISL_700279, EPI_ISL_700280, EPI_ISL_700281, EPI_ISL_700282, EPI_ISL_700283, EPI_ISL_700284, EPI_ISL_700285, EPI_ISL_700286, EPI_ISL_700287, EPI_ISL_700288, EPI_ISL_700289, EPI_ISL_700290, EPI_ISL_700291, EPI_ISL_700292, EPI_ISL_700293, EPI_ISL_700294, EPI_ISL_700295, EPI_ISL_700296, EPI_ISL_700297, EPI_ISL_700298, EPI_ISL_700299, EPI_ISL_700300, EPI_ISL_700301, EPI_ISL_700302, EPI_ISL_700303, EPI_ISL_700304, EPI_ISL_700305, EPI_ISL_700306, EPI_ISL_700307, EPI_ISL_700308, EPI_ISL_700309, EPI_ISL_700310, EPI_ISL_700311, EPI_ISL_700312, EPI_ISL_700313, EPI_ISL_700314, EPI_ISL_700315, EPI_ISL_700316, EPI_ISL_700317, EPI_ISL_700318, EPI_ISL_700319, EPI_ISL_700320, EPI_ISL_700321, EPI_ISL_700322, EPI_ISL_700323, EPI_ISL_700324, EPI_ISL_700325, EPI_ISL_723058, EPI_ISL_723059, EPI_ISL_723060, EPI_ISL_723061, EPI_ISL_723062, EPI_ISL_723063, EPI_ISL_723064, EPI_ISL_723065, EPI_ISL_723066, EPI_ISL_723067, EPI_ISL_723068, EPI_ISL_723069, EPI_ISL_723070, EPI_ISL_723071, EPI_ISL_723072, EPI_ISL_723073, EPI_ISL_723074, EPI_ISL_723075, EPI_ISL_723076 | | | |
| see above | Hematopathology Laboratory, ACTREC, TMC | Hematopathology Laboratory, ACTREC, TMC | ACTREC; Hematopathology Laboratory |
| EPI_ISL_1419077 | INSACOG-Mizoram | National Institute of Biomedical Genomics – INSACOG | Arindam Maitra; Gracy Laldinmawii; N Senthil Kumar; Nidhan Kumar Biswas; Saumitra Das; Sreedhar Chinnaswamy; Swagnik Roy |
| EPI_ISL_1419199, EPI_ISL_1419200, EPI_ISL_1419201, EPI_ISL_1419255, EPI_ISL_1419256, EPI_ISL_1419257, EPI_ISL_1419258, EPI_ISL_1419259, EPI_ISL_1419260, EPI_ISL_1419300, EPI_ISL_1419301, EPI_ISL_1419302, EPI_ISL_1419303, EPI_ISL_1419304, EPI_ISL_1419305, EPI_ISL_1419306, EPI_ISL_1419307, EPI_ISL_1419308, EPI_ISL_1419309, EPI_ISL_1419310, EPI_ISL_1419311, EPI_ISL_1419312, EPI_ISL_1419313, EPI_ISL_1419314, EPI_ISL_1419315, EPI_ISL_1419316, EPI_ISL_1419317, EPI_ISL_1419318, EPI_ISL_1419319, EPI_ISL_1419320, EPI_ISL_1419321, EPI_ISL_1419322, EPI_ISL_1419323, EPI_ISL_1419324, EPI_ISL_1419325, EPI_ISL_1419326, EPI_ISL_1419327, EPI_ISL_1419328, EPI_ISL_1419329, EPI_ISL_1419330, EPI_ISL_1419331, EPI_ISL_1419332, EPI_ISL_1419333, EPI_ISL_1419334, EPI_ISL_1419335, EPI_ISL_1419336, EPI_ISL_1419338, EPI_ISL_1419339, EPI_ISL_1419340, EPI_ISL_1419341, EPI_ISL_1419342, EPI_ISL_1419343, EPI_ISL_1419344, EPI_ISL_1419345, EPI_ISL_1419346, EPI_ISL_1419490, EPI_ISL_1419506, EPI_ISL_1419507, EPI_ISL_1419508, EPI_ISL_1419509, EPI_ISL_1419651, EPI_ISL_1419652, EPI_ISL_1419761, EPI_ISL_1419818, EPI_ISL_1419819, EPI_ISL_1419820, EPI_ISL_1419821, EPI_ISL_1419822, EPI_ISL_1419823, EPI_ISL_1419824, EPI_ISL_1419825, EPI_ISL_1419826, EPI_ISL_1419827, EPI_ISL_1419828, EPI_ISL_1419829, EPI_ISL_1419830, EPI_ISL_2132690, EPI_ISL_2132691 | | | |
| see above | INSACOG-WB | National Institute of Biomedical Genomics – INSACOG | Ajay Chakraborti; Arindam Maitra; Bhaswati Bandyopadhyay; Nidhan Kumar Biswas; Saumitra Das; Sreedhar Chinnaswamy; Tamal Ghosh |
| EPI_ISL_1063835 | Immunogenomics lab, Institute of Life Sciences, Bhubaneswar | Immunogenomics lab, Institute of Life Sciences, Bhubaneswar | Ajay Parida; Amol M. Kanampalliwar; Arup Ghosh; Atimukta Jha; INSACOG Consortium; Kirtal Hansdah; Punit Prasad; Rajeeb Swain; Rupesh Dash; Safal Walia; Shifu Aggarwal; Sunil K. Raghav |
| EPI_ISL_3473314, EPI_ISL_3473458, EPI_ISL_3473516, EPI_ISL_3473700, EPI_ISL_3473715 | Indian Council of Medical Research-National Institute of Virology, Microbial Containment Complex | Indian Council of Medical Research-National Institute of Virology, Microbial Containment Complex | Pragya Yadav |
| EPI_ISL_969365 | Indian Council of Medical Research-National Institute of Virology, Pune | Indian Council of Medical Research-National Institute of Virology, Maximum Containment Laboratory | Pragya D. Yadav |
| EPI_ISL_1708329, EPI_ISL_1708334, EPI_ISL_1708335, EPI_ISL_1708338, EPI_ISL_1708339, EPI_ISL_1708340, EPI_ISL_1708342, EPI_ISL_1708343, EPI_ISL_1708344, EPI_ISL_1708345, EPI_ISL_1708346, EPI_ISL_1708347, EPI_ISL_1708348, EPI_ISL_1708349, EPI_ISL_1708350, EPI_ISL_1708351, EPI_ISL_1708352, EPI_ISL_1708358 | | | |
| see above | Kodigehalli PHC | inStem NCBS - INSACOG | Uma Ramakrishnan Dasaradhi Palakodeti Aswin SaiNarain |
| EPI_ISL_1103557 | Kurnool Medical College (KMC) | CSIR Institute of Genomics and Integrative Biology | Abhinav Jain; Avileli Surekha; Bani Jolly; Damam Srinivasulu; Juturu Vijaya Lakshmi; Mohamed Imran; Mohit Kumar Divakar; PR Vanaja; Pallavali Roja Rani; Pulala Chandrasekhar; Rahul C Bhoyar; Sridhar Sivasubbu; Vigneshwar Senthivel; Vinod Scaria |
| EPI_ISL_862815 | Multidisciplinary Research Unit, DHR-ICMR, Institute of Medical Sciences, Banaras Hindu University, Varanasi | Multidisciplinary Research Unit, DHR-ICMR, Institute of Medical Sciences, Banaras Hindu University, Varanasi | Abhay Kumar Yadav; Ashish; Chetan Sahni; Deepa Devadas; Gulshan Kumar; Jay Prakash Maurya; Manpreet Kaur; Nitish Kumar Singh; Prashant Singh; Priyoneel Basu; Richa Arya; Royana Singh; Surendra Pratap Mishra; Tribhuwan Mohan Mohapatra |
| EPI_ISL_577639, EPI_ISL_577640, EPI_ISL_577641, EPI_ISL_577642, EPI_ISL_577643, EPI_ISL_577644, EPI_ISL_577645, EPI_ISL_577646, EPI_ISL_577647, EPI_ISL_577648, EPI_ISL_577649, EPI_ISL_577650, EPI_ISL_577651, EPI_ISL_577652, EPI_ISL_577653, EPI_ISL_577654, EPI_ISL_577655, EPI_ISL_577656, EPI_ISL_577657, EPI_ISL_577658, EPI_ISL_577659, EPI_ISL_577660, EPI_ISL_577661, EPI_ISL_577662, EPI_ISL_577663, EPI_ISL_577664, EPI_ISL_577665, EPI_ISL_577666, EPI_ISL_577667, EPI_ISL_577668, EPI_ISL_577669, EPI_ISL_577670, EPI_ISL_577672, EPI_ISL_577673, EPI_ISL_577674, EPI_ISL_577675, EPI_ISL_577676, EPI_ISL_577677, EPI_ISL_577678, EPI_ISL_577679, EPI_ISL_577680, EPI_ISL_577681, EPI_ISL_577682, EPI_ISL_577683, EPI_ISL_577684, EPI_ISL_577685, EPI_ISL_577686, EPI_ISL_577687, EPI_ISL_577688, EPI_ISL_577689, EPI_ISL_577690, EPI_ISL_577691, EPI_ISL_577692, EPI_ISL_577693, EPI_ISL_577694, EPI_ISL_577695, EPI_ISL_577696, EPI_ISL_577697, EPI_ISL_577698, EPI_ISL_577699, EPI_ISL_577700, EPI_ISL_577701, EPI_ISL_577702, EPI_ISL_577703, EPI_ISL_577704, EPI_ISL_577705, EPI_ISL_577706, EPI_ISL_577708, EPI_ISL_577709, EPI_ISL_577710, EPI_ISL_577711, EPI_ISL_577712, EPI_ISL_577713, EPI_ISL_577716, EPI_ISL_577717, EPI_ISL_577720, EPI_ISL_577721, EPI_ISL_577722, EPI_ISL_577723, EPI_ISL_577727, EPI_ISL_577728, EPI_ISL_577730, EPI_ISL_577731, EPI_ISL_582241 | | | |
| see above | NIV Influenza | NIV Influenza | Potdar V |
| EPI_ISL_1384836, EPI_ISL_1384837, EPI_ISL_1384838, EPI_ISL_1384839, EPI_ISL_1384840, EPI_ISL_1384841, EPI_ISL_1384842, EPI_ISL_1415115, EPI_ISL_1415142, EPI_ISL_1415146, EPI_ISL_1415151, EPI_ISL_1415154, EPI_ISL_1415185, EPI_ISL_1415187, EPI_ISL_1415230, EPI_ISL_1415325, EPI_ISL_1415330, EPI_ISL_1415352, EPI_ISL_1415375 | | | |
| see above | National Centre For Cell Science | National Centre For Cell Science – INSACOG | Ajay Pillai; Dhiraj Paul; INSACOG Consortium team; Manoj Kumar Bhat; Mitali Inamdar; Mohak P Gujare; Shivang P. Bhanushali; Sonal Manik Chavan; Yogesh Shouche. |
| EPI_ISL_2106595, EPI_ISL_2106596, EPI_ISL_2106600, EPI_ISL_2106601, EPI_ISL_2106604, EPI_ISL_2106610, EPI_ISL_2106613, EPI_ISL_2106614, EPI_ISL_2106616, EPI_ISL_2106618, EPI_ISL_2106619, EPI_ISL_2106626, EPI_ISL_2106628, EPI_ISL_2106629, EPI_ISL_2106630, EPI_ISL_2106631, EPI_ISL_2106635, EPI_ISL_2106637, EPI_ISL_2106638, EPI_ISL_2106639, EPI_ISL_2106641, EPI_ISL_2106642, EPI_ISL_2106644, EPI_ISL_2106652, EPI_ISL_2106653, EPI_ISL_2106654, EPI_ISL_2106655, EPI_ISL_2106656, EPI_ISL_2106657, EPI_ISL_2106658, EPI_ISL_2106659, EPI_ISL_2106660, EPI_ISL_2106661, EPI_ISL_2106662, EPI_ISL_2106663, EPI_ISL_2106664, EPI_ISL_2106665, EPI_ISL_2106666, EPI_ISL_2106667, EPI_ISL_2106668, EPI_ISL_2106669, EPI_ISL_2106670, EPI_ISL_2106671, EPI_ISL_2106672, EPI_ISL_2106673, EPI_ISL_2106674, EPI_ISL_2106675, EPI_ISL_2106676, EPI_ISL_2106677, EPI_ISL_2106678, EPI_ISL_2106679, EPI_ISL_2106680, EPI_ISL_2106681, EPI_ISL_2106682, EPI_ISL_2106683, EPI_ISL_2106684, EPI_ISL_2106685, EPI_ISL_2106686, EPI_ISL_2106687, EPI_ISL_2106688, EPI_ISL_2106689, EPI_ISL_2106690, EPI_ISL_2106691, EPI_ISL_2106692, EPI_ISL_2106693, EPI_ISL_2106694, EPI_ISL_2106695, EPI_ISL_2106696, EPI_ISL_2106697, EPI_ISL_2106698, EPI_ISL_2106699, EPI_ISL_2106700, EPI_ISL_2106701, EPI_ISL_2106702, EPI_ISL_2106703, EPI_ISL_2106704, EPI_ISL_2106705, EPI_ISL_2106706, EPI_ISL_2106707, EPI_ISL_2106708, EPI_ISL_2106709, EPI_ISL_2106710, EPI_ISL_2106711, EPI_ISL_2106712, EPI_ISL_2106713, EPI_ISL_2106714, EPI_ISL_2106715, EPI_ISL_2106716, EPI_ISL_2106717, EPI_ISL_2106718, EPI_ISL_2106719, EPI_ISL_2106720, EPI_ISL_2106721, EPI_ISL_2106722, EPI_ISL_2106723, EPI_ISL_2106724, EPI_ISL_2106725, EPI_ISL_2106726, EPI_ISL_2106727, EPI_ISL_2106728, EPI_ISL_2106729, EPI_ISL_2106730, EPI_ISL_2106731, EPI_ISL_2106732, EPI_ISL_2106733, EPI_ISL_2106734, EPI_ISL_2106735, EPI_ISL_2106736, EPI_ISL_2106737, EPI_ISL_2106738, EPI_ISL_2106739, EPI_ISL_2106740, EPI_ISL_2106741, EPI_ISL_2106742, EPI_ISL_2106743, EPI_ISL_2106744, EPI_ISL_2106745, EPI_ISL_2106746, EPI_ISL_2106747, EPI_ISL_2106748, EPI_ISL_2106749, EPI_ISL_2106750, EPI_ISL_2106751, EPI_ISL_2106752, EPI_ISL_2106753, EPI_ISL_2106754, EPI_ISL_2106755, EPI_ISL_2106756, EPI_ISL_2106757, EPI_ISL_2106758, EPI_ISL_2106759, EPI_ISL_2106760, EPI_ISL_2106761, EPI_ISL_2106762, EPI_ISL_2106763, EPI_ISL_2106764, EPI_ISL_2106765, EPI_ISL_2106766, EPI_ISL_2106767, EPI_ISL_2106768, EPI_ISL_2106769, EPI_ISL_2106770, EPI_ISL_2106771, EPI_ISL_2106772, EPI_ISL_2106773, EPI_ISL_2106774, EPI_ISL_2106775, EPI_ISL_2106776, EPI_ISL_2106777, EPI_ISL_2106778, EPI_ISL_2106779, EPI_ISL_2106780, EPI_ISL_2106781, EPI_ISL_2106782, EPI_ISL_2106783, EPI_ISL_2106784, EPI_ISL_2106785, EPI_ISL_2106786, EPI_ISL_2106787, EPI_ISL_2106788, EPI_ISL_2106789, EPI_ISL_2106790, EPI_ISL_2106791, EPI_ISL_2106792, EPI_ISL_2106793, EPI_ISL_2106794, EPI_ISL_2106795, EPI_ISL_2106796, EPI_ISL_2106797, EPI_ISL_2106798, EPI_ISL_2106800, EPI_ISL_2106801, EPI_ISL_2106802 | | | |
| see above | National Centre for Disease Control | CDFD-INSACOG | Ashwin Dalal; Asmita Gupta; Divya Vashisht; Murali Bashyam; Pratyusha Bala; Vinay Donipadi |
| EPI_ISL_2460613, EPI_ISL_2460620, EPI_ISL_2460628, EPI_ISL_2460630, EPI_ISL_2460635, EPI_ISL_2460639, EPI_ISL_2460668, EPI_ISL_2460679, EPI_ISL_2460687, EPI_ISL_2460688, EPI_ISL_2460689, EPI_ISL_2461258, EPI_ISL_2556669, EPI_ISL_2556670, EPI_ISL_2556671, EPI_ISL_2556672, EPI_ISL_2556673, EPI_ISL_2556674, EPI_ISL_2556675, EPI_ISL_2556676, EPI_ISL_2556677, EPI_ISL_2556678, EPI_ISL_2556679, EPI_ISL_2556680, EPI_ISL_2556681, EPI_ISL_2556682, EPI_ISL_2556683, EPI_ISL_2556684, EPI_ISL_2556685, EPI_ISL_2556686, EPI_ISL_2556687, EPI_ISL_2556688, EPI_ISL_2556689, EPI_ISL_2556690, EPI_ISL_2556691, EPI_ISL_2556692, EPI_ISL_2556693, EPI_ISL_2556694, EPI_ISL_2556695, EPI_ISL_2556696, EPI_ISL_2556697, EPI_ISL_2556698, EPI_ISL_2556699, EPI_ISL_2556700, EPI_ISL_2556701, EPI_ISL_2556702, EPI_ISL_2556703, EPI_ISL_2556704, EPI_ISL_2556705, EPI_ISL_2556706, EPI_ISL_2556707, EPI_ISL_2556708, EPI_ISL_2556709 | | | |
| see above | National Centre for Disease Control (NCDC) Biotechnology Division, Delhi | NCDC Delhi, Biotechnology Division INSACOG | Hema Gogia; Hemlata Lall; Kalaiarasan Ponnusamy; Mahesh S Dhar; Manoj K Singh; Meena Datta; Partha Rakshit; Preeti Madan; Priyanka Singh; Radhakrishnan V. S; Robin Marwal; Sandhya Kabra; Sujeet K Singh; Uma Sharma |
| EPI_ISL_541723, EPI_ISL_541724 | National Institute of Virology, NIV Influenza | National Institute of Virology, NIV Influenza | Potdar V |
| EPI_ISL_644715, EPI_ISL_644716, EPI_ISL_644717, EPI_ISL_644718, EPI_ISL_644719, EPI_ISL_644720, EPI_ISL_644721, EPI_ISL_644722, EPI_ISL_644723, EPI_ISL_644724 | | | |
| see above | Osmania Medical College | CSIR-Centre for Cellular and Molecular Biology | Archana Bharadwaj Siva; B Himasri; Blessy B John; Divya Tej Sowpati; Dr.P.Shashikala Reddy; Dr.S.Pavani; Dr.V.Sudha Rani; Karthik Bharadwaj Tallapaka; Lamuk Zaveri; M Soujanya Reddy; Namami Gaur; Nikhil Hajirnis; PPV Satya Prasad; Payel Mukherjee; Pratheusa Maccha; Priya Singh; Purushotham Vodnala; Rakesh K Mishra; Sakshi Shambhavi; Shagufta Khan; Sofia Banu; Tulasi Nagabandi; Viswagithe S L |
| EPI_ISL_2501528 | Regional VRDL,Dibrugarh | Regional VRDL,Dibrugarh | A Jakharia; B Borkakoty; C Bhattacharya; D Biswas; K Narain; Mandakini Das Sarma; R Hazarika |
| EPI_ISL_1708330, EPI_ISL_1708336, EPI_ISL_1708337, EPI_ISL_1708361 | Sahakara Nagar PHC | inStem NCBS - INSACOG | Uma Ramakrishnan Dasaradhi Palakodeti Aswin SaiNarain |
| EPI_ISL_676509 | TNMC & Nair ch. Hospital | CSIR-Institute of Genomics and Integrative Biology | Akshay Kanakan; Amol Borse; Janani Srinivasa Vasudevan; Jasmina Savak; Jayanthi Shastri; Mahesh Sangar; Manish Pathak; Mayuresh Vishwanathan; Nirjhar Chatterjee; Rajesh Pandey; Ranjeet Maurya; Sachee Agrawal; |

| | | | |
|---|---|---|---|
| | | | Shruthi Sachidanandan; Shrutika Pophale; Shweta Kawankar; Subrat Thanapati; Suresh Poojari; Swapneil Parikh; Utkarsha Yelve; Vasil Nachan |
| EPI_ISL_779705, EPI_ISL_779706, EPI_ISL_779708, EPI_ISL_779710 | The Foundation for Medical Research | The Foundation for Medical Research | Ambreen Shaikh; Ayan Mandal; Grishma Patel; Jayanthi Shastri; Kalpana Sriraman; Kayzad Nilgiriwala; Nerges Mistry; Nirjhar Chatterjee; Shreevatsa Udupa; Smriti Vaswani; Swapneil Parikh; Tejal Mestry |
| EPI_ISL_1708331, EPI_ISL_1708332, EPI_ISL_1708333, EPI_ISL_1708341, EPI_ISL_1708353, EPI_ISL_1708354, EPI_ISL_1708355, EPI_ISL_1708356, EPI_ISL_1708357, EPI_ISL_1708359, EPI_ISL_1708360, EPI_ISL_1708362, EPI_ISL_1708363, EPI_ISL_1708364, EPI_ISL_1708365, EPI_ISL_1708366, EPI_ISL_1708367, EPI_ISL_1708368, EPI_ISL_1708369, EPI_ISL_1708370, EPI_ISL_1708371, EPI_ISL_1708372, EPI_ISL_1708373, EPI_ISL_1708374, EPI_ISL_1708375, EPI_ISL_1708376, EPI_ISL_1708377, EPI_ISL_1708378, EPI_ISL_1708379, EPI_ISL_1708380, EPI_ISL_1708381 | | | |
| see above | Thindlu PHC | inStem NCBS - INSACOG | Uma Ramakrishnan Dasaradhi Palakodeti Aswin SaiNarain |
