## Supplementary material for "SARS-COV-2 δ variant drives the pandemic in India and Europe via two subvariants": Acknowledgement table on the GISAID genomes used in this study

We gratefully acknowledge the following Authors from the Originating laboratories responsible for obtaining the specimens, as well as the Submitting laboratories where the genome data were generated and shared via GISAID, on which this research is based.

All Submitters of data may be contacted directly via www.gisaid.org

Authors are sorted alphabetically.

| Accession ID | Originating Laboratory | Submitting Laboratory | Authors |
|---|---|---|---|
| EPI_ISL_1663507, EPI_ISL_1663516, EPI_ISL_1663522, EPI_ISL_1663523 | AIIMS, Patna | Institute of Life Sciences - INSACOG | Ajay Parida; Amol M. Kanampalliwar; Arup Ghosh; Atimukta Jha; INSACOG Consortium; Punit Prasad; Rajeeb Swain; Rupesh Dash; Safal Walia; Shifu Aggarwal; Sunil K. Raghav |
| EPI_ISL_2231794 | AP SSO | CSIR-Centre for Cellular and Molecular Biology-INSACOG | Amareshwar Vodapalli; Ara Sreenivas; Archana Bharadwaj Siva; B Himasri; Blessy B John; Divya Tej Sowpati; Karthik Bharadwaj Tallapaka; Lamuk Zaveri; Payel Mukherjee; Rakesh K Mishra; Sharath Chandra Thota; Shreekant Verma; Sofia Banu; Tulasi Nagabandi; Valli Nagalakshmi Undamatla; Viswagithe S L |
| EPI_ISL_2426214 | All India Institute of Medical Sciences, Delhi | CSIR Institute of Genomics and Integrative Biology | Abhinav Jain; Afra Shamnath; Anjali Bajaj; Arvinden VR; Bani Jolly; COVID CBNAAT CORE GROUP; Kiran Bala; Mercy Rophina; Mohamed Imran; Mohit Kumar Divakar; Nayer Jamshed; Praveen Aggarwal; Rahul C. Bhoyar; Rama Chaudhry; Randeep Guleria; Ritu Gupta; Sridhar Sivasubbu; Subrata Sinha; Urvashi B Singh; Vigneshwar Senthivel; Vinod Scaria |
| EPI_ISL_2484895, EPI_ISL_2484897, EPI_ISL_2484898 | Amdavad Municipal Corporation, Ahmedabad | Gujarat Biotechnology Research Centre | Chaitanya Joshi; Dalip Singh Rathore; Dinesh Kumar; Hiren Mandalia; Madhvi Joshi; Manish Kumar; Nitin Savaliya; Ramesh Pandit; Sonal Sharma; Twinkle Soni |
| EPI_ISL_2391488 | BJ Medical College and Civil Hospital, Ahmedabad | Gujarat Biotechnology Research Centre | Chaitanya Joshi; Dinesh Kumar; Janvi Raval; Madhvi Joshi; Nitesh Shah; Nitin Savaliya; Pranay Shah; Ramesh Pandit; Sonal Sharma; Twinkle Soni; Umang Mishra; Zarna Patel; Zuber Saiyed |
| EPI_ISL_4127507, EPI_ISL_4127551, EPI_ISL_4127751, EPI_ISL_4128439, EPI_ISL_4128445, EPI_ISL_4128459, EPI_ISL_4128988, EPI_ISL_4129060, EPI_ISL_4129066, EPI_ISL_4129374, EPI_ISL_4129602, EPI_ISL_4129749, EPI_ISL_4129855 | | | |
| see above | BJMC, Pune | IISER Pune | Aurnab Ghose; Joy Merwin Monteiro; Krishanpal Karmodiya; Rajesh Karyakarte; Suvarna Joshi |
| EPI_ISL_4415477, EPI_ISL_4415478, EPI_ISL_4415482, EPI_ISL_4415487, EPI_ISL_4415488, EPI_ISL_4415491, EPI_ISL_4415492, EPI_ISL_4415496, EPI_ISL_4415497, EPI_ISL_4415500, EPI_ISL_4415504, EPI_ISL_4415505, EPI_ISL_4415506, EPI_ISL_4415507, EPI_ISL_4415524, EPI_ISL_4415531, EPI_ISL_4415572, EPI_ISL_4415599, EPI_ISL_4415608, EPI_ISL_4415631, EPI_ISL_4415639, EPI_ISL_4415664, EPI_ISL_4415680, EPI_ISL_4415684, EPI_ISL_4415686, EPI_ISL_4415736, EPI_ISL_4415748, EPI_ISL_4415749, EPI_ISL_4415751, EPI_ISL_4415753 | | | |
| see above | BJMC, Pune | IISER Pune-INSACOG | Aurnab Ghose; Joy Merwin Monteiro; Krishanpal Karmodiya; Rajesh Karyakarte; Suvarna Joshi |
| EPI_ISL_1663247, EPI_ISL_2342659, EPI_ISL_2342868, EPI_ISL_2342873, EPI_ISL_2342878 | BRLSABVM Government Medical College, Rajnandgaon, Chhattisgarh | Institute of Life Sciences - INSACOG | Ajay Parida; Amol M. Kanampalliwar; Arup Ghosh; Atimukta Jha; INSACOG Consortium; Omprakash Shiriwas; Punit Prasad; Rajeeb Swain; Rupesh Dash; Safal Walia; Sana Fatma; Shifu Aggarwal; Sunil K. Raghav |
| EPI_ISL_2373501, EPI_ISL_2373524, EPI_ISL_2373545 | Banaras Hindu University | CSIR-Centre for Cellular and Molecular Biology-INSACOG | ; Abhay Kumar Yadav; Ajay Kumar Yadav; Amareshwar Vodapalli; Ara Sreenivas; Archana Bharadwaj Siva; Ashish; B Himasri; Blessy B John; Chetan Sahni; Deepa Devadas; Divya Tej Sowpati; Gunjan Rai; Gyaneshwer Chaubey; Jaya Chakraborty; Karthik Bharadwaj Tallapaka; Lamuk Zaveri; Maneesha Upadhyay; Manpreet Kaur; Nitish Kumar Singh; Payel Mukherjee; Priyoneel Basu; Rakesh K Mishra; Royana Singh; Saumya Singh; Saurabh Singh; Shani Vishwakarma; Sharath Chandra Thota; Shivam Tiwari; Shivani Mishra; Shreekant Verma; Sofia Banu; Surendra Pratap Mishra; Tulasi Nagabandi; Umesh Choudhary; Valli Nagalakshmi Undamatla; Viswagithe S L |
| EPI_ISL_1970221, EPI_ISL_1970223, EPI_ISL_1970224, EPI_ISL_1970286 | Biotechnology Division, NCDC Delhi | NCDC Delhi, Biotechnology Division | Hema Gogia; Hemlata Lall; Kalaiarasan Ponnusamy; Mahesh S Dhar; Manoj K Singh; Meena Datta; Partha Rakshit; Preeti Madan; Priyanka Singh; Radhakrishnan V. S; Robin Marwal; Sandhya Kabra; Sujeet K Singh; Uma Sharma |
| EPI_ISL_1663304, EPI_ISL_1663307, EPI_ISL_1663308, EPI_ISL_1663312 | CIIMS, Bilaspur, Chhattisgarh | Institute of Life Sciences - INSACOG | Ajay Parida; Amol M. Kanampalliwar; Arup Ghosh; Atimukta Jha; INSACOG Consortium; Punit Prasad; Rajeeb Swain; Rupesh Dash; Safal Walia; Shifu Aggarwal; Sunil K. Raghav |
| EPI_ISL_1838398, EPI_ISL_1838463, EPI_ISL_1838549, EPI_ISL_1838787, EPI_ISL_1838805, EPI_ISL_1838806, EPI_ISL_1838809, EPI_ISL_1838811, EPI_ISL_1838812, EPI_ISL_1838814, EPI_ISL_1838828, EPI_ISL_2660993, EPI_ISL_2661022 | | | |
| see above | CSIR-Centre for Cellular and Molecular Biology | CSIR-Centre for Cellular and Molecular Biology-INSACOG | ; Amareshwar Vodapalli; Ara Sreenivas; Archana Bharadwaj Siva; B Himasri; Blessy B John; Divya Tej Sowpati; Karthik Bharadwaj Tallapaka; Lamuk Zaveri; Onkar Kulkarni; Payel Mukherjee; Priya Nurkurthy; Rakesh K Mishra; Sharath Chandra Thota; Shreekant Verma; Sofia Banu; Sumedha Avadhanula; Tulasi Nagabandi; Valli Nagalakshmi Undamatla; Vidhyadhari Methuku; Viswagithe S L |
| EPI_ISL_4546215, EPI_ISL_4546225, EPI_ISL_4546243 | CSIR-Centre for Cellular and Molecular Biology - INSACOG | CSIR-Centre for Cellular and Molecular Biology - INSACOG | Amareshwar Vodapalli; Ara Sreenivas; Archana Bharadwaj Siva; B Himasri; Divya Tej Sowpati; Jandhyala Sai Krishna; Karthik Bharadwaj Tallapaka; Lamuk Zaveri; Onkar Kulkarni; Payel Mukherjee; Priya Nurkurthy; Rakesh K Mishra; Shreekant Verma; Sofia Banu; Sumedha Avadhanula; Tulasi Nagabandi; Valli Nagalakshmi Undamatla; Vidhyadhari Methuku |
| EPI_ISL_1916411, EPI_ISL_1916412, EPI_ISL_1916414, EPI_ISL_1916417, EPI_ISL_1916423, EPI_ISL_1916424, EPI_ISL_1916433, EPI_ISL_1916434, EPI_ISL_1916442, EPI_ISL_1916445, EPI_ISL_1916446, EPI_ISL_1916454, EPI_ISL_1916456, EPI_ISL_1916464, EPI_ISL_1916465, EPI_ISL_1916468, EPI_ISL_1916475, EPI_ISL_1916476, EPI_ISL_1916486, EPI_ISL_1916489 | | | |
| see above | CSIR-National Environmental Engineering Research Institute | CSIR-Centre for Cellular and Molecular Biology - INSACOG | Amareshwar Vodapalli; Ara Sreenivas; B Himasri; Divya Tej Sowpati; Karthik Bharadwaj Tallapaka; Krishna Khairnar; Lamuk Zaveri; Onkar Kulkarni; Rakesh K Mishra; Sharath Chandra Thota; Shreekant Verma; Sofia Banu; Viswagithe S L |
| EPI_ISL_2441372, EPI_ISL_2441385, EPI_ISL_2441386 | CSIR-National Environmental Engineering Research Institute | CSIR-Centre for Cellular and Molecular Biology-INSACOG | Amareshwar Vodapalli; Ara Sreenivas; Archana Bharadwaj Siva; B Himasri; Divya Tej Sowpati; Karthik Bharadwaj Tallapaka; Krishna Khairnar; Lamuk Zaveri; Onkar Kulkarni; Payel Mukherjee; Priya Nurkurthy; Rakesh K Mishra; Shreekant Verma; Sofia Banu; Sumedha Avadhanula; Tulasi Nagabandi; Valli Nagalakshmi Undamatla; Vidhyadhari Methuku |
| EPI_ISL_2106967 | Centre for DNA Fingerprinting and Diagnostics | CDFD -INSACOG | Ashwin Dalal; Asmita Gupta; Divya Vashisht; Murali Bashyam; Pratyusha Bala; Vinay Donipadi |
| EPI_ISL_3006894, EPI_ISL_3006917 | Chhattisgarh Institute of Medical Sciences (CIMS), Bilaspur | Institute of Life Sciences - INSACOG | Ajay Parida; Amol M. Kanampalliwar; Arup Ghosh; Atimukta Jha; INSACOG Consortium; Omprakash Shiriwas; Punit Prasad; Rajeeb Swain; Rupesh Dash; Safal Walia; Sana Fatma; Shifu Aggarwal; Sunil K. Raghav |
| EPI_ISL_3276964, EPI_ISL_3276972, EPI_ISL_3276984, EPI_ISL_3276988, EPI_ISL_3276995, EPI_ISL_3276998, EPI_ISL_3277001, EPI_ISL_3277085, EPI_ISL_3277087, EPI_ISL_3277136 | | | |
| see above | Clinical Virology Laboratory, Institute of Liver and Biliary Sciences | ILBS | Amit Pandey; Chhagan Bihari Sharma; Diptanu Paul; Ekta Gupta; Reshu Agarwal; Shiv Kumar Sarin; Varun Suroliya |
| EPI_ISL_4415410, EPI_ISL_4415411, EPI_ISL_4415412, EPI_ISL_4415413, EPI_ISL_4415414, EPI_ISL_4415415, EPI_ISL_4415416, EPI_ISL_4415417, EPI_ISL_4415418, EPI_ISL_4415419, EPI_ISL_4415420, EPI_ISL_4415421, EPI_ISL_4415422, EPI_ISL_4415423, EPI_ISL_4415424, EPI_ISL_4415425, EPI_ISL_4415426, EPI_ISL_4415427, EPI_ISL_4415428, EPI_ISL_4415429, EPI_ISL_4415430, EPI_ISL_4415431, EPI_ISL_4415432, EPI_ISL_4415433, EPI_ISL_4415434, EPI_ISL_4415435, EPI_ISL_4415436, EPI_ISL_4415437, EPI_ISL_4415438, EPI_ISL_4415439, EPI_ISL_4415440, EPI_ISL_4415441, EPI_ISL_4415527, EPI_ISL_4415528, EPI_ISL_4415533, EPI_ISL_4415604, EPI_ISL_4415605, EPI_ISL_4415610, EPI_ISL_4415613, EPI_ISL_4415614, EPI_ISL_4415616, EPI_ISL_4415621, EPI_ISL_4415623, EPI_ISL_4415634, EPI_ISL_4415636, EPI_ISL_4415641, EPI_ISL_4415642, EPI_ISL_4415646, EPI_ISL_4415647, EPI_ISL_4415649, EPI_ISL_4415650, EPI_ISL_4415652, EPI_ISL_4415653, EPI_ISL_4415666, EPI_ISL_4415667, EPI_ISL_4415672, EPI_ISL_4415673, EPI_ISL_4415676, EPI_ISL_4415678, EPI_ISL_4415679, EPI_ISL_4415681, EPI_ISL_4415685, EPI_ISL_4415704, EPI_ISL_4415721, EPI_ISL_4415733, EPI_ISL_4415739 | | | |
| see above | Department of Microbiology, Government Medical College, Baramati | IISER Pune-INSACOG | Aurnab Ghose; Joy Merwin Monteiro; Krishanpal Karmodiya |
| EPI_ISL_1662291 | Dept. Of Microbiology, Lt.. Baliram Kashyap Memorial Govt. Medical college, Dimrapal, Jagdalpur | Institute of Life Sciences - INSACOG | Ajay Parida; Amol M. Kanampalliwar; Arup Ghosh; Atimukta Jha; INSACOG Consortium; Punit Prasad; Rajeeb Swain; Rupesh Dash; Safal Walia; Shifu Aggarwal; Sunil K. Raghav |
| EPI_ISL_2379462, EPI_ISL_2379496, EPI_ISL_2379497, EPI_ISL_2379498, EPI_ISL_2379564, EPI_ISL_2379645, EPI_ISL_2379646, EPI_ISL_2379647, EPI_ISL_2379648, EPI_ISL_2379649 | | | |
| see above | Dr S Raju, Director of Public Heath and Preventive Medicine | inStem NCBS - INSACOG | Uma Ramakrishnan Dasaradhi Palakodeti Aswin SaiNarain |
| EPI_ISL_2521821, EPI_ISL_2521940, EPI_ISL_2521941, EPI_ISL_2521942, EPI_ISL_2521943 | Dr S raju, Director of public heath and preventive medicine | inStem NCBS - INSACOG | Uma Ramakrishnan Dasaradhi Palakodeti Aswin SaiNarain |
| EPI_ISL_2723800, EPI_ISL_2723801, EPI_ISL_2723830, EPI_ISL_2723831, EPI_ISL_2723917, EPI_ISL_2723922, EPI_ISL_2723923, EPI_ISL_2723924, EPI_ISL_2723925, EPI_ISL_2723926, EPI_ISL_2723927, EPI_ISL_2723976 | | | |
| see above | Dr S raju, Director of public heath and preventive medicine | inStem NCBS – INSACOG | Uma Ramakrishnan Dasaradhi Palakodeti Aswin SaiNarain |
| EPI_ISL_4106468 | Dr. B. Lal Institute of Biotechnology | Dr. B. Lal Institute of Biotechnology | Dr. Aditi Nag & Dr Sudipti Arora |
| EPI_ISL_4197532, EPI_ISL_4198119, EPI_ISL_4198392 | Dr. B. Lal Institute of Biotechnology, Jaipur | Dr. B. Lal Institute of Biotechnology, Jaipur | Aditi Nag and Sudipti Arora |
| EPI_ISL_2341931, EPI_ISL_2341932 | FM Medical College, Balasore | Institute of Life Sciences - INSACOG | Ajay Parida; Amol M. Kanampalliwar; Arup Ghosh; Atimukta Jha; INSACOG Consortium; Omprakash Shiriwas; Punit Prasad; Rajeeb Swain; Rupesh Dash; Safal Walia; Sana Fatma; Shifu Aggarwal; Sunil K. Raghav |
| EPI_ISL_2001139, EPI_ISL_2001143 | GMERS, Government Medical College, Gandhinagar | Gujarat Biotechnology Research Centre | Chaitanya Joshi; Dinesh Kumar; Gaurishankar Shrimali; Janvi Raval; Madhvi Joshi; Nitesh Shah; Nitin Savaliya; Ramesh Pandit; Sonal Sharma; Twinkle Soni; Umang Mishra; Zarna Patel; Zuber Saiyed |
| EPI_ISL_1940019 | GOVIMDRAJ NAGAR | inStem NCBS - INSACOG | Uma Ramakrishnan Dasaradhi Palakodeti Aswin SaiNarain |
| EPI_ISL_4125682, EPI_ISL_4125687, EPI_ISL_4125712 | Genepath, Pune | IISER Pune | Aurnab Ghose; Joy Merwin Monteiro; Krishanpal Karmodiya; Rajesh Karyakarte; Suvarna Joshi |
| EPI_ISL_3007890, EPI_ISL_3007902, EPI_ISL_3007905, EPI_ISL_3007906, EPI_ISL_3007916 | Government Medical College, Bettiah | Institute of Life Sciences - INSACOG | Ajay Parida; Amol M. Kanampalliwar; Arup Ghosh; Atimukta Jha; INSACOG Consortium; Omprakash Shiriwas; Punit Prasad; Rajeeb Swain; Rupesh Dash; Safal Walia; Sana Fatma; Shifu Aggarwal; Sunil K. Raghav |
| EPI_ISL_1940001, EPI_ISL_1940002, EPI_ISL_1940003, EPI_ISL_1940004, EPI_ISL_1940008, EPI_ISL_1940011, EPI_ISL_1940013, EPI_ISL_1940014, EPI_ISL_1940015 | | | |
| see above | HOSAHALLI | inStem NCBS - INSACOG | Uma Ramakrishnan Dasaradhi Palakodeti Aswin SaiNarain |
| EPI_ISL_2530319 | Hematopathology Laboratory, ACTREC, TMC | Hematopathology Laboratory, ACTREC, TMC | ACTREC; Hematopathology Laboratory |
| EPI_ISL_2189572, EPI_ISL_2189596, EPI_ISL_2189726, EPI_ISL_2189770, EPI_ISL_2555672 | ICMR-National Institute of Virology | NCDC Delhi, Biotechnology Division | A Walimbe; A.Awhale; A.Titkare; B.Apoorva; G.Divekar; H.Kengale; Hema Gogia; Hemlata Lall; K P.Shinde; K.Iyengar; K.Korabu; Kalaiarasan Ponnusamy; M L Choudhary; M.Das; Mahesh S Dhar; Manoj K Singh; Meena Datta; Partha Rakshit; Preeti Madan; Priyanka Singh; R.Verma; Radhakrishnan V. S; Robin Marwal; S Jadhav; S.Bhorekar; S.Jadhav; S.Shelkande; Sandhya Kabra; Sujeet K Singh; T.Sanjeevi; U.Saha; Uma Sharma; V Malik; V.Autude; V.Vipat; Z. Sayyed |
| EPI_ISL_1703662, EPI_ISL_1703668, EPI_ISL_1703740, EPI_ISL_1703878, EPI_ISL_1704234, EPI_ISL_1704561, EPI_ISL_1704605, EPI_ISL_1704617, EPI_ISL_1704618, EPI_ISL_1704620, EPI_ISL_1704625, EPI_ISL_1704626, EPI_ISL_1704627, EPI_ISL_1704628, EPI_ISL_1704629, EPI_ISL_1704630, EPI_ISL_1704632, EPI_ISL_1704634, EPI_ISL_1704635, EPI_ISL_1704636, EPI_ISL_1704637, EPI_ISL_1704638, EPI_ISL_1704639, EPI_ISL_1841293, EPI_ISL_1841317, EPI_ISL_1841318, EPI_ISL_1841355, EPI_ISL_1841356, EPI_ISL_1841357, EPI_ISL_1841365, EPI_ISL_1841366, EPI_ISL_1841367, EPI_ISL_1841382, EPI_ISL_1841383, EPI_ISL_1928407, EPI_ISL_1928417, EPI_ISL_1928439, EPI_ISL_1928457, EPI_ISL_1928462, EPI_ISL_1928477, EPI_ISL_1928484, EPI_ISL_1970422, EPI_ISL_2131509, EPI_ISL_2546013, | | | |

| | | | |
|---|---|---|---|
| EPI_ISL_2546023, EPI_ISL_2546034, EPI_ISL_2546053, EPI_ISL_2546063, EPI_ISL_2546068, EPI_ISL_2546078, EPI_ISL_2546085, EPI_ISL_2546089, EPI_ISL_2546099, EPI_ISL_2546102, EPI_ISL_2546104, EPI_ISL_2546106, EPI_ISL_2546107, EPI_ISL_2546122, EPI_ISL_2546136, EPI_ISL_2546145, EPI_ISL_2546154, EPI_ISL_2546156, EPI_ISL_2546157, EPI_ISL_2546158, EPI_ISL_2546163, EPI_ISL_2546164, EPI_ISL_2546187, EPI_ISL_2546228, EPI_ISL_2546230, EPI_ISL_2546233, EPI_ISL_2546275, EPI_ISL_2546276, EPI_ISL_2546278, EPI_ISL_2546279, EPI_ISL_2546281, EPI_ISL_2546289, EPI_ISL_2546291, EPI_ISL_2546294, EPI_ISL_2546295, EPI_ISL_2546297, EPI_ISL_2546298, EPI_ISL_2546304, EPI_ISL_2546308, EPI_ISL_2546321, EPI_ISL_2546327, EPI_ISL_2546330, EPI_ISL_2546336, EPI_ISL_2546337, EPI_ISL_2546349, EPI_ISL_2546350, EPI_ISL_2546356, EPI_ISL_2546357, EPI_ISL_2546359, EPI_ISL_2546360, EPI_ISL_2546362, EPI_ISL_2546364, EPI_ISL_2546365, EPI_ISL_2546366, EPI_ISL_2546367, EPI_ISL_2546368, EPI_ISL_2546374, EPI_ISL_2546375, EPI_ISL_2546381, EPI_ISL_2546387, EPI_ISL_2546388, EPI_ISL_2546389, EPI_ISL_2546396, EPI_ISL_2546397, EPI_ISL_2546401, EPI_ISL_2546403, EPI_ISL_2546411, EPI_ISL_2546414, EPI_ISL_2546416, EPI_ISL_2546417, EPI_ISL_2546418, EPI_ISL_2546419, EPI_ISL_2546422, EPI_ISL_2546425, EPI_ISL_2546431, EPI_ISL_2546432, EPI_ISL_2546434, EPI_ISL_2546435, EPI_ISL_2546437, EPI_ISL_2546438, EPI_ISL_2546443, EPI_ISL_2546444, EPI_ISL_2546447, EPI_ISL_2965870, EPI_ISL_3532947, EPI_ISL_3532948, EPI_ISL_3532961, EPI_ISL_3532963, EPI_ISL_3532964, EPI_ISL_3532965, EPI_ISL_3532966, EPI_ISL_3532967, EPI_ISL_3532968, EPI_ISL_3532969 | | | |
| see above | ICMR-National Institute of Virology - INSACOG | NIV Influenza | Dr. Varsha Potdar; Dr. Varsha Potdar and NIC Team |
| EPI_ISL_2878570, EPI_ISL_2878571, EPI_ISL_2878573, EPI_ISL_2878574, EPI_ISL_2878575, EPI_ISL_2878576, EPI_ISL_2878577, EPI_ISL_2878578, EPI_ISL_2878579, EPI_ISL_2878580, EPI_ISL_2878730, EPI_ISL_2878743, EPI_ISL_2878796, EPI_ISL_2878804, EPI_ISL_2878824, EPI_ISL_2878830, EPI_ISL_2878837, EPI_ISL_2878867, EPI_ISL_2878876, EPI_ISL_2878885, EPI_ISL_2878887, EPI_ISL_2878889, EPI_ISL_2878902, EPI_ISL_2878904, EPI_ISL_2878906, EPI_ISL_2878907, EPI_ISL_2878924, EPI_ISL_2878925, EPI_ISL_2878931, EPI_ISL_2878932, EPI_ISL_2878943, EPI_ISL_2878959, EPI_ISL_2878960, EPI_ISL_2878961, EPI_ISL_2878962, EPI_ISL_2878964, EPI_ISL_2878968, EPI_ISL_2878977, EPI_ISL_2878978, EPI_ISL_2878980, EPI_ISL_2878983, EPI_ISL_2878985, EPI_ISL_2878988, EPI_ISL_2878991, EPI_ISL_2878992, EPI_ISL_2878996, EPI_ISL_2878999, EPI_ISL_2879009, EPI_ISL_2879017, EPI_ISL_2879018, EPI_ISL_2879059, EPI_ISL_2879065, EPI_ISL_2879066, EPI_ISL_2879072, EPI_ISL_2879073, EPI_ISL_2879074, EPI_ISL_2879078, EPI_ISL_2879079, EPI_ISL_2879081, EPI_ISL_2879083, EPI_ISL_2879089, EPI_ISL_2879091, EPI_ISL_2879092, EPI_ISL_2879094, EPI_ISL_2879095, EPI_ISL_2879100, EPI_ISL_2879102, EPI_ISL_2879103, EPI_ISL_2879105, EPI_ISL_2879106, EPI_ISL_2879112, EPI_ISL_2879113, EPI_ISL_2879114, EPI_ISL_2879115, EPI_ISL_2879116, EPI_ISL_2879117, EPI_ISL_2879124, EPI_ISL_2879127, EPI_ISL_2879129, EPI_ISL_2879131, EPI_ISL_2879133, EPI_ISL_2879134, EPI_ISL_2879137, EPI_ISL_2879138, EPI_ISL_2879140, EPI_ISL_2879141, EPI_ISL_2879146, EPI_ISL_2879147, EPI_ISL_2879152, EPI_ISL_2879153, EPI_ISL_2879154, EPI_ISL_2879160, EPI_ISL_2879161, EPI_ISL_2879167, EPI_ISL_2880930, EPI_ISL_2881206, EPI_ISL_2881449, EPI_ISL_2881450, EPI_ISL_2881451, EPI_ISL_2881513, EPI_ISL_2881514 | | | |
| see above | INSACOG Surveillance | INSACOG at CSIR Institute of Genomics and Integrative Biology | INSACOG |
| EPI_ISL_2877817, EPI_ISL_2877818, EPI_ISL_2877819, EPI_ISL_2877820, EPI_ISL_2877821, EPI_ISL_2877822, EPI_ISL_2877823, EPI_ISL_2877824, EPI_ISL_2877825, EPI_ISL_2877826, EPI_ISL_2877827, EPI_ISL_2877828, EPI_ISL_2877829 | | | |
| see above | INSACOG-Assam | National Institute of Biomedical Genomics – INSACOG | Ajanta Sharma; Arindam Maitra; Kailash Chamuah; Lahari Saikia; Nidhan Kumar Biswas; Saumitra Das; Sreedhar Chinnaswamy |
| EPI_ISL_2521601, EPI_ISL_2521605, EPI_ISL_2521606, EPI_ISL_2521609 | INSACOG-Mizoram | National Institute of Biomedical Genomics – INSACOG | Arindam Maitra; Gracy Laldinmawii; N Senthil Kumar; Nidhan Kumar Biswas; Saumitra Das; Sreedhar Chinnaswamy; Swagnik Roy |
| EPI_ISL_4312739 | INSACOG-Mizoram | National Institute of Biomedical Genomics – INSACOG | Arindam Maitra; Gracy Laldinmawii; N Senthil Kumar; Nidhan Kumar Biswas; Saumitra Das; Sreedhar Chinnaswamy; Swagnik Roy |
| EPI_ISL_2151096, EPI_ISL_2151097, EPI_ISL_2151098 | INSACOG-Sikkim | National Institute of Biomedical Genomics – INSACOG | Arindam Maitra; Kaden Zangmu Bhutia; Nidhan Kumar Biswas; Saumitra Das; Sreedhar Chinnaswamy Tshering Ongmu Bhutia; Tara D Sharma |
| EPI_ISL_2433268, EPI_ISL_2433269 | INSACOG-Tripura | National Institute of Biomedical Genomics – INSACOG | Apurba Sarkar; Arindam Maitra; Nidhan Kumar Biswas; Saikat Majumder; Saumitra Das; Sreedhar Chinnaswamy; Tapan Majumdar |
| EPI_ISL_1419152, EPI_ISL_1589660, EPI_ISL_1589667, EPI_ISL_1589674, EPI_ISL_1589690, EPI_ISL_1589718, EPI_ISL_1589824, EPI_ISL_1589829, EPI_ISL_1589834, EPI_ISL_1589835, EPI_ISL_1589867, EPI_ISL_1589868, EPI_ISL_1589869, EPI_ISL_1589870, EPI_ISL_1589871, EPI_ISL_1589872, EPI_ISL_2503151, EPI_ISL_2503156, EPI_ISL_2503162, EPI_ISL_2503165, EPI_ISL_2503169, EPI_ISL_2503188, EPI_ISL_2503189, EPI_ISL_2503190, EPI_ISL_2503193, EPI_ISL_2503194, EPI_ISL_2503195, EPI_ISL_2503204, EPI_ISL_2503207, EPI_ISL_2503226, EPI_ISL_2503235, EPI_ISL_2503236, EPI_ISL_2503241, EPI_ISL_2503246, EPI_ISL_2503399, EPI_ISL_2503401, EPI_ISL_2503402, EPI_ISL_2503409, EPI_ISL_2503415, EPI_ISL_2503429, EPI_ISL_2503474, EPI_ISL_2503480, EPI_ISL_2503516, EPI_ISL_2503517, EPI_ISL_2503532, EPI_ISL_2503535, EPI_ISL_2503538, EPI_ISL_2503547, EPI_ISL_2548641, EPI_ISL_2548646, EPI_ISL_2548647, EPI_ISL_2548649, EPI_ISL_2548650, EPI_ISL_2548653, EPI_ISL_2548655, EPI_ISL_2548656, EPI_ISL_2548657, EPI_ISL_2548659, EPI_ISL_2548661, EPI_ISL_2548670, EPI_ISL_2548694, EPI_ISL_2548698, EPI_ISL_2548699, EPI_ISL_2548700, EPI_ISL_2548701, EPI_ISL_2548779, EPI_ISL_2548780 | | | |
| see above | INSACOG-WB | National Institute of Biomedical Genomics – INSACOG | Ajay Chakraborti; Arindam Maitra; Bhaswati Bandyopadhyay; Nidhan Kumar Biswas; Saumitra Das; Sreedhar Chinnaswamy; Tamal Ghosh |
| EPI_ISL_1662451, EPI_ISL_1663553, EPI_ISL_1663554, EPI_ISL_1663555, EPI_ISL_1663557, EPI_ISL_1663561, EPI_ISL_1663562, EPI_ISL_1663563, EPI_ISL_1663564 | | | |
| see above | Immunogenomics lab, Institute of Life Sciences, Bhubaneswar | Institute of Life Sciences - INSACOG | Ajay Parida; Amol M. Kanampalliwar; Arup Ghosh; Atimukta Jha; INSACOG Consortium; Punit Prasad; Rajeeb Swain; Rupesh Dash; Safal Walia; Shifu Aggarwal; Sunil K. Raghav |
| EPI_ISL_3473491 | Indian Council of Medical Research-National Institute of Virology, Microbial Containment Complex | Indian Council of Medical Research-National Institute of Virology, Microbial Containment Complex | Pragya Yadav |
| EPI_ISL_2341682, EPI_ISL_2341695, EPI_ISL_2341701 | Institute of Life Sciences, Bhubaneswar | Institute of Life Sciences - INSACOG | Ajay Parida; Amol M. Kanampalliwar; Arup Ghosh; Atimukta Jha; INSACOG Consortium; Omprakash Shiriwas; Punit Prasad; Rajeeb Swain; Rupesh Dash; Safal Walia; Sana Fatma; Shifu Aggarwal; Sunil K. Raghav |
| EPI_ISL_1939969 | KAMAKSHIPALYA | inStem NCBS - INSACOG | Uma Ramakrishnan Dasaradhi Palakodeti Aswin SaiNarain |
| EPI_ISL_1818617, EPI_ISL_1818618, EPI_ISL_2545211 | KEMPEGOWDA INTERNATIONAL AIRPORT | INSACOG-KA, NIMHANS | Ananthapadmanabha Kotambail; Anita S Desai; Anson Kunjumon George; Chetan G K; Chitra Pattabiraman; Darshan Sreenivas; Gautham Arunachal Udupi; Pramada Prasad; V Ravi |
| EPI_ISL_3135898, EPI_ISL_3135900, EPI_ISL_3135901, EPI_ISL_3135902, EPI_ISL_3135903, EPI_ISL_3135904, EPI_ISL_3135925, EPI_ISL_3135927, EPI_ISL_3135928, EPI_ISL_3135929, EPI_ISL_3135932, EPI_ISL_3136203 | | | |
| see above | Laboratory Medicine and Molecular Diagnostics | Laboratory Medicine and Molecular Diagnostics | Dayakar Seetha; Heera R Pillai; Radhakrishna R Nair; Radhakrishnan R Nair; Sanughosh Kalpothodi |
| EPI_ISL_1663541, EPI_ISL_1663543, EPI_ISL_1663548, EPI_ISL_1663549 | MGM Medical College, Jamshedpur | Institute of Life Sciences - INSACOG | Ajay Parida; Amol M. Kanampalliwar; Arup Ghosh; Atimukta Jha; INSACOG Consortium; Punit Prasad; Rajeeb Swain; Rupesh Dash; Safal Walia; Shifu Aggarwal; Sunil K. Raghav |
| EPI_ISL_2157019, EPI_ISL_2157021 | Mapmygenome | CSIR-Centre for Cellular and Molecular Biology-INSACOG | Amareshwar Vodapalli; Ara Sreenivas; Archana Bharadwaj Siva; B Himasri; Blessy B John; Divya Tej Sowpati; Karthik Bharadwaj Tallapaka; Lamuk Zaveri; Payel Mukherjee; Rakesh K Mishra; Sharath Chandra Thota; Shreekant Verma; Sofia Banu; Tulasi Nagabandi; Valli Nagalakshmi Undamatla; Viswagithe S L |
| EPI_ISL_3844261, EPI_ISL_3844269, EPI_ISL_3844402 | N.H.L Municipal Medical College, Ahmedabad | Gujarat Biotechnology Research Centre | Arpit Shukla; Bhadreshsinh Gohil; Chaitanya Joshi; Dinesh Kumar; Janvi Raval; Jayshri Pethani; Madhvi Joshi; Nimesh Patel; Nitin Shukla; Ramesh Pandit; Zarna Patel |
| EPI_ISL_1663366, EPI_ISL_1663367, EPI_ISL_1663375, EPI_ISL_1663376, EPI_ISL_2342642, EPI_ISL_2342644, EPI_ISL_2342656, EPI_ISL_2342722, EPI_ISL_2342919, EPI_ISL_2342930, EPI_ISL_2342931, EPI_ISL_2342932, EPI_ISL_2342937, EPI_ISL_2342938, EPI_ISL_2342946, EPI_ISL_2342947 | | | |
| see above | NCCS, Pune | Institute of Life Sciences - INSACOG | Ajay Parida; Amol M. Kanampalliwar; Arup Ghosh; Atimukta Jha; INSACOG Consortium; Omprakash Shiriwas; Punit Prasad; Rajeeb Swain; Rupesh Dash; Safal Walia; Sana Fatma; Shifu Aggarwal; Sunil K. Raghav |
| EPI_ISL_3162118 | NHL Municipal Medical College, Ahmedbad | Gujarat Biotechnology Research Centre | Chaitanya Joshi; Dinesh Kumar; Janvi Raval; Jayshri Pethani; Madhvi Joshi; Nitesh Shah; Nitin Savaliya; Nitin Shukla; Ramesh Pandit; Sonal Sharma; Twinkle Soni; Umang Mishra; Zarna Patel; Zuber Saiyed |
| EPI_ISL_2728444, EPI_ISL_2842129 | NIMHANS | INSACOG-KA, NIMHANS | Ananthapadmanabha Kotambail; Anita S Desai; Anson Kunjumon George; Chetan G K; Chitra Pattabiraman; Darshan Sreenivas; Ellango Ramasamy; Gautham Arunachal Udupi; Mahesh Kumar.C.S; Sony Sharma; V Ravi |
| EPI_ISL_1939986, EPI_ISL_1939999 | NIMHANS | inStem NCBS - INSACOG | Uma Ramakrishnan Dasaradhi Palakodeti Aswin SaiNarain |
| EPI_ISL_1544014, EPI_ISL_2162392, EPI_ISL_2162393, EPI_ISL_2162395, EPI_ISL_2162411, EPI_ISL_2162417, EPI_ISL_2162423, EPI_ISL_2162425, EPI_ISL_2162427, EPI_ISL_2162429, EPI_ISL_2162435, EPI_ISL_2162437, EPI_ISL_2162438, EPI_ISL_2162441, EPI_ISL_2162444, EPI_ISL_2162445, EPI_ISL_2162455, EPI_ISL_2162461, EPI_ISL_2304047, EPI_ISL_2304073 | | | |
| see above | National Centre For Cell Science | National Centre For Cell Science – INSACOG | Ajay Pillai; Dhiraj Paul; INSACOG Consortium team; Manoj Kumar Bhat; Mitali Inamdar; Mohak P Gujare; Shivang P. Bhanushali; Sonal Manik Chavan; Yogesh Shouche |
| EPI_ISL_2107009, EPI_ISL_2107010, EPI_ISL_2107017, EPI_ISL_2107020, EPI_ISL_2107022, EPI_ISL_2107023, EPI_ISL_2107025, EPI_ISL_2107027, EPI_ISL_2107033, EPI_ISL_2107034, EPI_ISL_2107060, EPI_ISL_2107062, EPI_ISL_2107063, EPI_ISL_2107066, EPI_ISL_2107070, EPI_ISL_2107071, EPI_ISL_2107072, EPI_ISL_2107075, EPI_ISL_2107077, EPI_ISL_2107080, EPI_ISL_2107081, EPI_ISL_2107083, EPI_ISL_2107085, EPI_ISL_2107087, EPI_ISL_2107088, EPI_ISL_2107089, EPI_ISL_2107090 | | | |
| see above | National Centre For Cell Science – INSACOG | National Centre For Cell Science | Ajay Pillai; Dhiraj Paul; INSACOG Consortium team; Manoj Kumar Bhat; Mitali Inamdar; Mohak P Gujare; Shivang P. Bhanushali; Yogesh Shouche |
| EPI_ISL_2099644, EPI_ISL_2233495, EPI_ISL_2233544, EPI_ISL_2233550, EPI_ISL_2233595 | National Centre for Disease Control | CDFD -INSACOG | Ashwin Dalal; Asmita Gupta; Divya Vashisht; Murali Bashyam; Pratyusha Bala; To be addded later; Vinay Donipadi |
| EPI_ISL_2460052, EPI_ISL_2460533, EPI_ISL_2460546, EPI_ISL_2460547, EPI_ISL_2460550, EPI_ISL_2460605, EPI_ISL_2460627, EPI_ISL_2460633, EPI_ISL_2460681, EPI_ISL_2460703, EPI_ISL_2460707, EPI_ISL_2460794, EPI_ISL_2460868, EPI_ISL_2460869, EPI_ISL_2460870, EPI_ISL_2460872, EPI_ISL_2460874, EPI_ISL_2461170, EPI_ISL_2461256, EPI_ISL_2461258, EPI_ISL_2461418, EPI_ISL_2461419, EPI_ISL_2461509, EPI_ISL_2461515, EPI_ISL_2461539, EPI_ISL_2461564, EPI_ISL_2461780, EPI_ISL_2461781, EPI_ISL_2461788, EPI_ISL_2461789, EPI_ISL_2461822, EPI_ISL_2461922, EPI_ISL_2461929, EPI_ISL_2461945, EPI_ISL_2461948, EPI_ISL_2461949, EPI_ISL_2461954, EPI_ISL_2461955, EPI_ISL_2461956, EPI_ISL_2461957, EPI_ISL_2461958, EPI_ISL_2461959, EPI_ISL_2461960, EPI_ISL_2461961, EPI_ISL_2461962, EPI_ISL_2461963, EPI_ISL_2461965, EPI_ISL_2461966, EPI_ISL_2461979, EPI_ISL_2461982, EPI_ISL_2461991, EPI_ISL_2502030, EPI_ISL_2502031, EPI_ISL_2502059, EPI_ISL_2502060, EPI_ISL_2502071, EPI_ISL_2502088, EPI_ISL_2502095, EPI_ISL_2502098, EPI_ISL_2502106, EPI_ISL_2502109, EPI_ISL_2502120, EPI_ISL_2502126, EPI_ISL_2502129, EPI_ISL_2502171, EPI_ISL_2502172, EPI_ISL_2502178, EPI_ISL_2502180, EPI_ISL_2502185, EPI_ISL_2502196, EPI_ISL_2502218, EPI_ISL_2502222, EPI_ISL_2502231, EPI_ISL_2502232, EPI_ISL_2502237, EPI_ISL_2502247, EPI_ISL_2502248, EPI_ISL_2502268, EPI_ISL_2502272, EPI_ISL_2502273, EPI_ISL_2502275, EPI_ISL_2502279, EPI_ISL_2502291, EPI_ISL_2502296, EPI_ISL_2502299, EPI_ISL_2502304, EPI_ISL_2504284, EPI_ISL_2504861, EPI_ISL_2555827, EPI_ISL_2555829, EPI_ISL_2555830, EPI_ISL_2555909, EPI_ISL_2555911, EPI_ISL_2555929, EPI_ISL_2555931, EPI_ISL_2555941, EPI_ISL_2555943, EPI_ISL_2555947, EPI_ISL_2555950, EPI_ISL_2555951, EPI_ISL_2555966, EPI_ISL_2556187, EPI_ISL_2556200, EPI_ISL_2556447, EPI_ISL_2556448, EPI_ISL_2556524, EPI_ISL_2556527, EPI_ISL_2556566, EPI_ISL_2556582, EPI_ISL_2556583, EPI_ISL_2556605, EPI_ISL_2556857, EPI_ISL_2556971, EPI_ISL_2556976, EPI_ISL_2556979, EPI_ISL_2556999, EPI_ISL_2557000, EPI_ISL_2557002, EPI_ISL_2557021, EPI_ISL_2557032, EPI_ISL_2955863, EPI_ISL_2955924, EPI_ISL_2955931 | | | |
| see above | National Centre for Disease Control (NCDC) Biotechnology Division, Delhi | NCDC Delhi, Biotechnology Division INSACOG | Hema Gogia; Hemlata Lall; Kalaiarasan Ponnusamy; Mahesh S Dhar; Manoj K Singh; Meena Datta; Partha Rakshit; Preeti Madan; Priyanka Singh; Radhakrishnan V. S; Robin Marwal; Sandhya Kabra; Sujeet K Singh; Uma Sharma |
| EPI_ISL_4436392, EPI_ISL_4446961, EPI_ISL_4446962, EPI_ISL_4474101, EPI_ISL_4474162 | National Centre for Disease Control (NCDC) Biotechnology Division, Delhi - INSACOG | NCDC Delhi, Biotechnology Division - INSACOG | Hema Gogia; Hemlata Lall; Kalaiarasan Ponnusamy; Mahesh S Dhar; Manoj K Singh; Meena Datta; Partha Rakshit; Preeti Madan; Priyanka Singh; Radhakrishnan V. S; Robin Marwal; Sandhya Kabra; Sujeet K Singh; Uma Sharma |
| EPI_ISL_4533969, EPI_ISL_4533970, EPI_ISL_4533971 | Neuberg Supratech Refrence Laboratory, Surat | Gujarat Biotechnology Research Centre | Apurvasinh Puvar; Arpit Shukla; Bhadreshsinh Gohil; Chaitanya Joshi; Dinesh Kumar; Janvi Raval; Madhvi Joshi; Manthan Shah; Nimesh Patel; Nitin Savaliya; Nitin Shukla; Priyank Chavda; Ramesh Pandit; Sonal Sharma; Zarna Patel |
| EPI_ISL_2955810 | Northern Railway Central Hospital, New Delhi, India | NCDC Delhi, Biotechnology Division INSACOG | Hema Gogia; Hemlata Lall; Kalaiarasan Ponnusamy; Mahesh S Dhar; Manoj K Singh; Meena Datta; Partha Rakshit; Preeti Madan; Priyanka Singh; Radhakrishnan V. S; Robin Marwal; Sandhya Kabra; Sujeet K Singh; Uma Sharma |
| EPI_ISL_4104000 | Pandit Raghunath Murmu Medical College, Baripada | Institute of Life Sciences-INSACOG | Ajay Parida; Amol M. Kanampalliwar; Arup Ghosh; Atimukta Jha; INSACOG Consortium; Punit Prasad; Rajeeb Swain; Rupesh Dash; Safal Walia; Sana Fatma; Shifu Aggarwal; Sunil K. Raghav |
| EPI_ISL_2341679, EPI_ISL_2341692, EPI_ISL_2341693, EPI_ISL_2341708, EPI_ISL_2341709, EPI_ISL_2341721, EPI_ISL_2341722, EPI_ISL_2341724, EPI_ISL_2341790, EPI_ISL_2341862, EPI_ISL_2341863, EPI_ISL_2341864, EPI_ISL_2341866, EPI_ISL_2341867, EPI_ISL_2341869, EPI_ISL_2341871, EPI_ISL_2341872, EPI_ISL_2341875, EPI_ISL_2341876, EPI_ISL_2341880, EPI_ISL_2341881, EPI_ISL_2341883, EPI_ISL_2341885, EPI_ISL_2341887, EPI_ISL_2341889, EPI_ISL_2341890, EPI_ISL_2341891, EPI_ISL_2341892, EPI_ISL_2341896, EPI_ISL_2341897, EPI_ISL_2341898, EPI_ISL_2341899, EPI_ISL_2341900, EPI_ISL_2341901, EPI_ISL_2341902, EPI_ISL_2341904, EPI_ISL_2341905, EPI_ISL_2341906, EPI_ISL_2341907, EPI_ISL_2341909, EPI_ISL_2341910, EPI_ISL_2341911, EPI_ISL_2341912, EPI_ISL_2341913, EPI_ISL_2341914, EPI_ISL_2341920, EPI_ISL_2341921, EPI_ISL_3009631, EPI_ISL_3009635, EPI_ISL_3009636, EPI_ISL_3009638, EPI_ISL_3009642, EPI_ISL_3009643, EPI_ISL_3009645, EPI_ISL_3009647, EPI_ISL_3009648, EPI_ISL_3009650, EPI_ISL_3009651, EPI_ISL_3009654, EPI_ISL_3009655, EPI_ISL_3009656, EPI_ISL_3009657, EPI_ISL_3009658, EPI_ISL_3009660, EPI_ISL_3009664, EPI_ISL_3009665, EPI_ISL_3009666, EPI_ISL_3009667, EPI_ISL_3009668, EPI_ISL_3009669, EPI_ISL_3009672, EPI_ISL_3009673, EPI_ISL_3009680, EPI_ISL_3009685, EPI_ISL_3009686, EPI_ISL_3009688, EPI_ISL_3009689, EPI_ISL_3009691, EPI_ISL_3009693 | | | |
| see above | Pt. Jawahar Lal Nehru Memorial Medical College, Raipur | Institute of Life Sciences - INSACOG | Ajay Parida; Amol M. Kanampalliwar; Arup Ghosh; Atimukta Jha; INSACOG Consortium; Omprakash Shiriwas; Punit Prasad; Rajeeb Swain; Rupesh Dash; Safal Walia; Sana Fatma; Shifu Aggarwal; Sunil K. Raghav |
| EPI_ISL_1663529, EPI_ISL_1663534 | REGIONAL VRDL, ICMR-RMRC BBSR | Institute of Life Sciences - INSACOG | Ajay Parida; Amol M. Kanampalliwar; Arup Ghosh; Atimukta Jha; INSACOG Consortium; Punit Prasad; Rajeeb Swain; Rupesh Dash; Safal Walia; Shifu Aggarwal; Sunil K. Raghav |
| EPI_ISL_3453376, EPI_ISL_3453530, EPI_ISL_3453544, EPI_ISL_3453549, EPI_ISL_3453559, EPI_ISL_3453562, EPI_ISL_3453568, EPI_ISL_3453575, EPI_ISL_3453581, EPI_ISL_3453588, EPI_ISL_3453590, EPI_ISL_3453598, EPI_ISL_3453615, EPI_ISL_3453624, EPI_ISL_3453628, EPI_ISL_3453836, EPI_ISL_3453837 | | | |

| | | | |
|---|---|---|---|
| see above | RUHS College of Medical Sciences | CSIR-Centre for Cellular and Molecular Biology - INSACOG | Amareshwar Vodapalli; Ara Sreenivas; Archana Bharadwaj Siva; B Himasri; Divya Tej Sowpati; Dr. Gaurav Dalela; Dr. Jitendra Panda; Dr. Nilofer Khayyam; Dr. Raja Babu Panwar; Dr. Rajeev Gupta; Dr. Ramesh Sharma; Dr. Sonali Sharma; Dr. Sudhanshu Kacker; Dr. Vaseem Naheed Baig; Jandhyala Sai Krishna; Karthik Bharadwaj Tallapaka; Lamuk Zaveri; Onkar Kulkarni; Payel Mukherjee; Priya Nurkurthy; Rakesh K Mishra; Shreekant Verma; Sofia Banu; Sumedha Avadhanula; Tulasi Nagabandi; Valli Nagalakshmi Undamatla; Vidhyadhari Methuku |
| EPI_ISL_4104093 | Rajendra Memorial Research Institute of Medical Sciences | Institute of Life Sciences-INSACOG | Ajay Parida; Amol M. Kanampalliwar; Arup Ghosh; Atimukta Jha; INSACOG Consortium; Punit Prasad; Rajeeb Swain; Rupesh Dash; Safal Walia; Sana Fatma; Shifu Aggarwal; Sunil K. Raghav |
| EPI_ISL_2341852, EPI_ISL_2341853, EPI_ISL_2341855, EPI_ISL_2341856, EPI_ISL_2341859 | S.C.B. Medical College and Hospital, Cuttack | Institute of Life Sciences - INSACOG | Ajay Parida; Amol M. Kanampalliwar; Arup Ghosh; Atimukta Jha; INSACOG Consortium; Omprakash Shiriwas; Punit Prasad; Rajeeb Swain; Rupesh Dash; Safal Walia; Sana Fatma; Shifu Aggarwal; Sunil K. Raghav |
| EPI_ISL_2341727 | SLN Medical College and Hospital, Koraput | Institute of Life Sciences - INSACOG | Ajay Parida; Amol M. Kanampalliwar; Arup Ghosh; Atimukta Jha; INSACOG Consortium; Omprakash Shiriwas; Punit Prasad; Rajeeb Swain; Rupesh Dash; Safal Walia; Sana Fatma; Shifu Aggarwal; Sunil K. Raghav |
| EPI_ISL_3717689 | SMIMER Hospital & Medical College, Surat | Gujarat Biotechnology Research Centre | Arpit Shukla; Bhadreshsinh Gohil; Chaitanya Joshi; Dinesh Kumar; Janvi Raval; Madhvi Joshi; Manish Patel; Nimesh Patel; Nitin Shukla; Ramesh Pandit; Zarna Patel |
| EPI_ISL_1939974 | SUBRAMNAYA NAGAR | inStem NCBS - INSACOG | Uma Ramakrishnan Dasaradhi Palakodeti Aswin SaiNarain |
| EPI_ISL_1914584, EPI_ISL_1914588, EPI_ISL_1914591, EPI_ISL_1914592, EPI_ISL_1914593, EPI_ISL_1914594, EPI_ISL_1914595, EPI_ISL_1914596, EPI_ISL_1969753, EPI_ISL_2098718, EPI_ISL_2098719, EPI_ISL_2098720, EPI_ISL_2098722, EPI_ISL_2307108, EPI_ISL_2620740 | | | |
| see above | State Virus Research and Diagnostic Laboratory (VRDL), AIIMS Raipur | State Virus Research and Diagnostic Laboratory (VRDL), AIIMS Raipur | Anudita Bhargava; Jiyan Chandrawanshi; Kuldeep Sharma; MD Rafiiqullah Khan; Priyanka Singh; Pushpendra Singh; Sanjay Singh Negi; Somya Sharma |
| EPI_ISL_1838134, EPI_ISL_1838151, EPI_ISL_1838194, EPI_ISL_1838260, EPI_ISL_1838262, EPI_ISL_1838265, EPI_ISL_1838285, EPI_ISL_1838292, EPI_ISL_1838312, EPI_ISL_1838315, EPI_ISL_1838322, EPI_ISL_1838327, EPI_ISL_1838337 | | | |
| see above | The National Centre for Cell Science | CSIR-Centre for Cellular and Molecular Biology-INSACOG | Ajay Pillai; Amareshwar Vodapalli; Ara Sreenivas; Archana Bharadwaj Siva; B Himasri; Blessy B John; Dhiraj Paul; Divya Tej Sowpati; INSACOG Consortium team; Karthik Bharadwaj Tallapaka; Lamuk Zaveri; Manoj Kumar Bhat; Mitali Inamdar; Mohak P Gujare; Onkar Kulkarni; Payel Mukherjee; Rakesh K Mishra; Sharath Chandra Thota; Shivang P. Bhanushali; Shreekant Verma; Sofia Banu; Sonal Manik Chavan; Tulasi Nagabandi; Valli Nagalakshmi Undamatla; Viswagithe S L; Yogesh Shouche |
| EPI_ISL_2017748 | VRDL, Goverment Medical College (GMC) and Hospital, Valsad | Gujarat Biotechnology Research Centre | Chaitanya Joshi; Dinesh Kumar; Janvi Raval; Madhvi Joshi; Nitesh Shah; Nitin Savaliya; Ramesh Pandit; Sonal Sharma; Twinkle Soni; Umang Mishra; Vicky Gandhi; Zarna Patel; Zuber Saiyed |
| EPI_ISL_2001187, EPI_ISL_2001188, EPI_ISL_2001189, EPI_ISL_2001190, EPI_ISL_2001191, EPI_ISL_2001192, EPI_ISL_2001193, EPI_ISL_2001194, EPI_ISL_2001211, EPI_ISL_2001212, EPI_ISL_2017728, EPI_ISL_2017731, EPI_ISL_2017734, EPI_ISL_2017735, EPI_ISL_2017737, EPI_ISL_2017738 | | | |
| see above | VRDL, Goverment Medical College (GMC), Surat | Gujarat Biotechnology Research Centre | Chaitanya Joshi; Dinesh Kumar; Janvi Raval; Madhvi Joshi; Neeta Khandelwal; Nitesh Shah; Nitin Savaliya; Ramesh Pandit; Sonal Sharma; Twinkle Soni; Umang Mishra; Zarna Patel; Zuber Saiyed |
| EPI_ISL_3844272 | VRDL, P. D. U. Medical College, Rajkot | Gujarat Biotechnology Research Centre | Arpit Shukla; Bhadreshsinh Gohil; Chaitanya Joshi; Dinesh Kumar; Gauravi A Dhruva; Janvi Raval; Madhvi Joshi; Nimesh Patel; Nitin Shukla; Ramesh Pandit; Zarna Patel |
| EPI_ISL_3162096 | VRDL,Goverment Medical College Surat | Gujarat Biotechnology Research Centre | Chaitanya Joshi; Dinesh Kumar; Janvi Raval; Madhvi Joshi; Neeta Khandelwal; Nitesh Shah; Nitin Savaliya; Nitin Shukla; Ramesh Pandit; Sonal Sharma; Twinkle Soni; Umang Mishra; Zarna Patel; Zuber Saiyed |
| EPI_ISL_3844263, EPI_ISL_3844393, EPI_ISL_3844497 | VRDL,Goverment Medical College Surat (GMC) | Gujarat Biotechnology Research Centre | Arpit Shukla; Bhadreshsinh Gohil; Chaitanya Joshi; Dinesh Kumar; Janvi Raval; Madhvi Joshi; Neeta Khandelwal; Nimesh Patel; Nitin Shukla; Ramesh Pandit; Zarna Patel |
| EPI_ISL_3844265, EPI_ISL_3844268 | VRDL,Goverment Medical College Surat (SMIMER) | Gujarat Biotechnology Research Centre | Arpit Shukla; Bhadreshsinh Gohil; Chaitanya Joshi; Dinesh Kumar; Janvi Raval; Madhvi Joshi; Manish Patel; Nimesh Patel; Nitin Shukla; Ramesh Pandit; Zarna Patel |
| EPI_ISL_3162110, EPI_ISL_3162111 | VRDL,Goverment Medical College, Surat | Gujarat Biotechnology Research Centre | Chaitanya Joshi; Dinesh Kumar; Janvi Raval; Madhvi Joshi; Neeta Khandelwal; Nitesh Shah; Nitin Savaliya; Nitin Shukla; Ramesh Pandit; Sonal Sharma; Twinkle Soni; Umang Mishra; Zarna Patel; Zuber Saiyed |
| EPI_ISL_1662284, EPI_ISL_1663498, EPI_ISL_1663501, EPI_ISL_1663502 | Veer Surendra Sai Institute of Medical Sciences and Research, Burla, Sambalpur | Institute of Life Sciences - INSACOG | Ajay Parida; Amol M. Kanampalliwar; Arup Ghosh; Atimukta Jha; INSACOG Consortium; Punit Prasad; Rajeeb Swain; Rupesh Dash; Safal Walia; Shifu Aggarwal; Sunil K. Raghav |
