## Supplementary material for "SARS-COV-2 δ variant drives the pandemic in India and Europe via two subvariants": Acknowledgement table on the GISAID genomes used in this study

We gratefully acknowledge the following Authors from the Originating laboratories responsible for obtaining the specimens, as well as the Submitting laboratories where the genome data were generated and shared via GISAID, on which this research is based.

All Submitters of data may be contacted directly via www.gisaid.org

Authors are sorted alphabetically.

| Accession ID | Originating Laboratory | Submitting Laboratory | Authors |
|---|---|---|---|
| EPI_ISL_1663516 | AIIMS, Patna | Institute of Life Sciences - INSACOG | Ajay Parida; Amol M. Kanampalliwar; Arup Ghosh; Atimukta Jha; INSACOG Consortium; Punit Prasad; Rajeeb Swain; Rupesh Dash; Safal Walia; Shifu Aggarwal; Sunil K. Raghav |
| EPI_ISL_2231794 | AP SSO | CSIR-Centre for Cellular and Molecular Biology-INSACOG | Amareshwar Vodapalli; Ara Sreenivas; Archana Bharadwaj Siva; B Himasri; Blessy B John; Divya Tej Sowpati; Karthik Bharadwaj Tallapaka; Lamuk Zaveri; Payel Mukherjee; Rakesh K Mishra; Sharath Chandra Thota; Shreekant Verma; Sofia Banu; Tulasi Nagabandi; Valli Nagalakshmi Undamatla; Viswagithe S L |
| EPI_ISL_3972907 | AULSS 2 Marca Trevigiana | Istituto Zooprofilattico Sperimentale delle Venezie | Adelaide Milani; Alessia Schivo; Alice Fusaro; Ambra Pastori; Annalisa Salviato; Antonia Ricci; Calogero Terregino; Edoardo Giussani; Elisa Palumbo; Erika Giorgia Quaranta; Isabella Monne; Luca Tassoni |
| EPI_ISL_2484895, EPI_ISL_2484897, EPI_ISL_2484898 | Amdavad Municipal Corporation, Ahmedabad | Gujarat Biotechnology Research Centre | Chaitanya Joshi; Dalip Singh Rathore; Dinesh Kumar; Hiren Mandalia; Madhvi Joshi; Manish Kumar; Nitin Savaliya; Ramesh Pandit; Sonal Sharma; Twinkle Soni |
| EPI_ISL_3103859, EPI_ISL_3103908, EPI_ISL_3387525 | Arizona State University | Arizona State University | Ajeet Bains; Efrem S. Lim; Joshua LaBaer; Joy M. Blain; LaRinda A. Holland; Matthew F. Smith; Nathaniel Johnson; Nicholas J. Mellor; Peter T. Skidmore; Rabia Maqsood; Valerie Harris; Vel Murugan |
| EPI_ISL_1969249 | BBLK Palembang | National Institute of Health Research and Development | Arie Ardiansyah Nugraha; Hana Apsari Pawestri; Hartanti Dian Ikawati; Kartika Dewi Puspa; Krisna Pangesti; Nelly Puspandari; Subangkit; Triyani Soekarso; Vivi Setiawaty |
| EPI_ISL_1663247 | BRLSABVM Government Medical College, Rajnandgaon, Chhattisgarh | Institute of Life Sciences - INSACOG | Ajay Parida; Amol M. Kanampalliwar; Arup Ghosh; Atimukta Jha; INSACOG Consortium; Punit Prasad; Rajeeb Swain; Rupesh Dash; Safal Walia; Shifu Aggarwal; Sunil K. Raghav |
| EPI_ISL_2373501, EPI_ISL_2373524, EPI_ISL_2373545 | Banaras Hindu University | CSIR-Centre for Cellular and Molecular Biology-INSACOG | ; Abhay Kumar Yadav; Ajay Kumar Yadav; Amareshwar Vodapalli; Ara Sreenivas; Archana Bharadwaj Siva; Ashish; B Himasri; Blessy B John; Chetan Sahni; Deepa Devadas; Divya Tej Sowpati; Gunjan Rai; Gyaneshwer Chaubey; Jaya Chakraborty; Karthik Bharadwaj Tallapaka; Lamuk Zaveri; Maneesha Upadhyay; Manpreet Kaur; Nitish Kumar Singh; Payel Mukherjee; Priyoneel Basu; Rakesh K Mishra; Royana Singh; Saumya Singh; Saurabh Singh; Shani Vishwakarma; Sharath Chandra Thota; Shivam Tiwari; Shivani Mishra; Shreekant Verma; Sofia Banu; Surendra Pratap Mishra; Tulasi Nagabandi; Umesh Choudhary; Valli Nagalakshmi Undamatla; Viswagithe S L |
| EPI_ISL_3888984 | Bioscientia Labor Wermsdorf | Robert Koch Institute | |
| EPI_ISL_1970221, EPI_ISL_1970223, EPI_ISL_1970224 | Biotechnology Division, NCDC Delhi | NCDC Delhi, Biotechnology Division | Hema Gogia; Hemlata Lall; Kalaiarasan Ponnusamy; Mahesh S Dhar; Manoj K Singh; Meena Datta; Partha Rakshit; Preeti Madan; Priyanka Singh; Radhakrishnan V. S; Robin Marwal; Sandhya Kabra; Sujeet K Singh; Uma Sharma |
| EPI_ISL_1916417 | CSIR-National Environmental Engineering Research Institute | CSIR-Centre for Cellular and Molecular Biology - INSACOG | Amareshwar Vodapalli; Ara Sreenivas; B Himasri; Divya Tej Sowpati; Karthik Bharadwaj Tallapaka; Krishna Khairnar; Lamuk Zaveri; Onkar Kulkarni; Rakesh K Mishra; Sharath Chandra Thota; Shreekant Verma; Sofia Banu; Viswagithe S L |
| EPI_ISL_2441372 | CSIR-National Environmental Engineering Research Institute | CSIR-Centre for Cellular and Molecular Biology-INSACOG | Amareshwar Vodapalli; Ara Sreenivas; Archana Bharadwaj Siva; B Himasri; Divya Tej Sowpati; Karthik Bharadwaj Tallapaka; Krishna Khairnar; Lamuk Zaveri; Onkar Kulkarni; Payel Mukherjee; Priya Nurkurthy; Rakesh K Mishra; Shreekant Verma; Sofia Banu; Sumedha Avadhanula; Tulasi Nagabandi; Valli Nagalakshmi Undamatla; Vidhyadhari Methuku |
| EPI_ISL_3276964, EPI_ISL_3277085, EPI_ISL_3277087 | Clinical Virology Laboratory, Institute of Liver and Biliary Sciences | ILBS | Amit Pandey; Chhagan Bihari Sharma; Diptanu Paul; Ekta Gupta; Reshu Agarwal; Shiv Kumar Sarin; Varun Suroliya |
| EPI_ISL_3014085, EPI_ISL_3014086, EPI_ISL_3014093, EPI_ISL_3014096, EPI_ISL_3014098, EPI_ISL_3014099, EPI_ISL_3014101, EPI_ISL_3014102, EPI_ISL_3014103, EPI_ISL_3014104, EPI_ISL_3014105 | | | |
| see above | Cotugno | TIGEM | Antonio Grimaldi Patrizia Annunziata Francesco Panariello Biancamaria Pierri Claudia Tiberio Teresa Giuliano Valentina Bouche Chiara Colantuono Maria Concetta Cuomo Denise Di Concilio Lucio Di Filippo Anna Manfredi Marcello Salvi Antonio Limone Luigi Atripaldi Pellegrino Cerino Andrea Ballabio Davide Cacchiarelli |
| EPI_ISL_4049066 | DC Public Health Lab/ Dept. of Forensic Sciences | DC Public Health Lab/ Dept. of Forensic Sciences | Brittany Hamilton; Connie Maza; Elizabeth Zelaya; Janis Doss; Jocelyn Hauser; Monica Mann; Nathan Bruns; Sarah Scott; Scott Nguyen |
| EPI_ISL_3530389, EPI_ISL_3531252, EPI_ISL_3531589 | Department of Clinical Microbiology, Odense University Hospital, Odense, Denmark | Statens Serum Institut Bioinformatics and Microbial Genomics | Danish Covid-19 Genome Consortium |
| EPI_ISL_3315002 | Diagnostic and Research Center of Infectious Diseases, Medical Faculty, Andalas University | Genomik Solidaritas Indonesia Laboratorium / Diagnostic and Research Center of Infectious Diseases, Medical Faculty, Andalas University | Andani Eka Putra; Annisa Muthiah Sukirman; Anuraj Shankar; Ariel Pradipta; Carissa Sintca Wijaya; Dede Rahman Agustian; Dhahlia Agustina Cahyono; Gracia Felias Enos Korompis; Linosefa; Louisa Markus; Meutia Ayuputeri Kumaheri; Reinhart Gabriel; Vania Gavrila Wikasa |
| EPI_ISL_2379462, EPI_ISL_2379496, EPI_ISL_2379497, EPI_ISL_2379498, EPI_ISL_2379564, EPI_ISL_2379645, EPI_ISL_2379646, EPI_ISL_2379647, EPI_ISL_2379648, EPI_ISL_2379649 | | | |
| see above | Dr S Raju, Director of Public Heath and Preventive Medicine | inStem NCBS - INSACOG | Uma Ramakrishnan Dasaradhi Palakodeti Aswin SaiNarain |
| EPI_ISL_2521821, EPI_ISL_2521940, EPI_ISL_2521941, EPI_ISL_2521942, EPI_ISL_2521943 | Dr S raju, Director of public heath and preventive medicine | inStem NCBS - INSACOG | Uma Ramakrishnan Dasaradhi Palakodeti Aswin SaiNarain |
| EPI_ISL_2723830, EPI_ISL_2723922, EPI_ISL_2723924, EPI_ISL_2723925, EPI_ISL_2723927, EPI_ISL_2723976 | Dr S raju, Director of public heath and preventive medicine | inStem NCBS – INSACOG | Uma Ramakrishnan Dasaradhi Palakodeti Aswin SaiNarain |
| EPI_ISL_2788315, EPI_ISL_4077292, EPI_ISL_4077293, EPI_ISL_4077294, EPI_ISL_4077295, EPI_ISL_4077296, EPI_ISL_4077297, EPI_ISL_4077298, EPI_ISL_4077299, EPI_ISL_4077300, EPI_ISL_4077301, EPI_ISL_4077302, EPI_ISL_4077303, EPI_ISL_4077304, EPI_ISL_4077305, EPI_ISL_4077306, EPI_ISL_4077307, EPI_ISL_4077308, EPI_ISL_4077309, EPI_ISL_4077310, EPI_ISL_4077311, EPI_ISL_4077312, EPI_ISL_4077313, EPI_ISL_4077314, EPI_ISL_4077315, EPI_ISL_4077316, EPI_ISL_4077317, EPI_ISL_4077318, EPI_ISL_4077319, EPI_ISL_4077320, EPI_ISL_4077322, EPI_ISL_4077323, EPI_ISL_4077324, EPI_ISL_4077326, EPI_ISL_4077327 | | | |
| see above | Dutch COVID-19 response team | National Institute for Public Health and the Environment (RIVM) | Adam Meijer; AnneMarie van den Brandt; Annelies Kroneman; Bas van der Veer; Chantal Reusken; Dennis Schmitz; Dirk Eggink; Euníce Then; Florian Zwagemaker; Harry Vennema; Ivo van Walle; Jeroen Cremer; Karim Hajji; Kim Freriks; Lisa Wijsman; Lynn Aarts; Melissa van Tuil; Ryanne Jaarsma; Sanne Bos; Sharon van den Brink; Stijn van Rossum; on behalf of the national COVID-19 response team |
| EPI_ISL_1940019 | GOVIMDRAJ NAGAR | inStem NCBS - INSACOG | Uma Ramakrishnan Dasaradhi Palakodeti Aswin SaiNarain |
| EPI_ISL_3980510, EPI_ISL_3980512, EPI_ISL_3980513, EPI_ISL_3980515, EPI_ISL_3980516, EPI_ISL_3980517, EPI_ISL_3980518 | | | |
| see above | Ga East Hospital | University of Health and Allied Sciences (UHAS) COVID-19 Testing and Research Centre | John O. Gyapong and the UHAS COVID-19 Lab Team; Jones Gyamfi; Kwabena O. Duedu; Reuben Ayivor-Djanie |
| EPI_ISL_3215995 | Genomica Lab Molecular, Mexico | Andersen lab at Scripps Research | Jose Horacio Reyna Verdugo; Jose Roman Chavez Mendez; Luis Alberto Rangel Gonzalez; Martin Gonzalez Ibarra; SEARCH Alliance San Diego with Jonathan Gonzalez Garcia |
| EPI_ISL_1940003, EPI_ISL_1940008, EPI_ISL_1940011, EPI_ISL_1940013, EPI_ISL_1940014, EPI_ISL_1940015 | HOSAHALLI | inStem NCBS - INSACOG | Uma Ramakrishnan Dasaradhi Palakodeti Aswin SaiNarain |
| EPI_ISL_3398929 | Houston Health Dept. | Houston Health Dept. | Adolpho Lara; Pamela Brown; Ryker Penn; Yanlai Lai |

| | | | |
|---|---|---|---|
| EPI_ISL_1704234, EPI_ISL_1704561, EPI_ISL_1704605, EPI_ISL_1841365, EPI_ISL_1841366, EPI_ISL_1928407, EPI_ISL_1928417, EPI_ISL_2131509, EPI_ISL_2546187, EPI_ISL_2965870, EPI_ISL_3532947, EPI_ISL_3532948 | | | |
| see above | ICMR-National Institute of Virology - INSACOG | NIV Influenza | Dr. Varsha Potdar; Dr. Varsha Potdar and NIC Team |
| EPI_ISL_2878570, EPI_ISL_2878571, EPI_ISL_2878573, EPI_ISL_2878574, EPI_ISL_2878575, EPI_ISL_2878576, EPI_ISL_2878577, EPI_ISL_2878578, EPI_ISL_2878579, EPI_ISL_2878580, EPI_ISL_2878730, EPI_ISL_2880930, EPI_ISL_2881449, EPI_ISL_2881450, EPI_ISL_2881451, EPI_ISL_2881513, EPI_ISL_2881514 | | | |
| see above | INSACOG Surveillance | INSACOG at CSIR Institute of Genomics and Integrative Biology | INSACOG |
| EPI_ISL_2877817, EPI_ISL_2877818, EPI_ISL_2877819, EPI_ISL_2877820, EPI_ISL_2877821, EPI_ISL_2877822, EPI_ISL_2877823, EPI_ISL_2877824, EPI_ISL_2877825, EPI_ISL_2877826, EPI_ISL_2877827, EPI_ISL_2877828, EPI_ISL_2877829 | | | |
| see above | INSACOG-Assam | National Institute of Biomedical Genomics – INSACOG | Ajanta Sharma; Arindam Maitra; Kailash Chamuah; Lahari Saikia; Nidhan Kumar Biswas; Saumitra Das; Sreedhar Chinnaswamy |
| EPI_ISL_2521601, EPI_ISL_2521605, EPI_ISL_2521606 | INSACOG-Mizoram | National Institute of Biomedical Genomics – INSACOG | Arindam Maitra; Gracy Laldinmawii; N Senthil Kumar; Nidhan Kumar Biswas; Saumitra Das; Sreedhar Chinnaswamy; Swagnik Roy |
| EPI_ISL_1419152 | INSACOG-WB | National Institute of Biomedical Genomics – INSACOG | Ajay Chakraborti; Arindam Maitra; Bhaswati Bandyopadhyay; Nidhan Kumar Biswas; Saumitra Das; Sreedhar Chinnaswamy; Tamal Ghosh |
| EPI_ISL_4077684 | Institute for Infectious Diseases | Institute for Infectious Diseases | Alban Ramette; Christian Baumann; Cora Sägesser; Franziska Suter-Riniker; Loïc Borcard; Miguel A Terrazos Miani; Nicole Liechti; Pascal Bittel; Peter Keller; Sonja Gempeler; Stefan Neuenschwander; Stephen L Leib |
| EPI_ISL_3721807 | Instituto Nacional de Medicina Genomica | Instituto Nacional de Medicina Genomica | Cedro-Tanda A; Escobar-Arrazola MA; Herrera-Montalvo LA.; Hidalgo-Miranda A; Mendoza-Vargas A; Ramirez-Vega O; Rangel-DeLeon D; Reyes-Grajeda JP; Yair Alfaro-Mora |
| EPI_ISL_3909809 | KEMRI-Wellcome Trust Research Programme,Kilifi | KEMRI-Wellcome Trust Research Programme,Kilifi | Agoti C.; Githinji G.; Lambisia A.; Mburu M.W.; Mohamed K.S.; Morobe J.; Ndwiga L.; Ochola I; Ong'era M.Edidah; de Laurent Z. |
| EPI_ISL_2937904 | LESP Aguascalientes | Instituto de Diagnostico y Referencia Epidemiologicos (INDRE) | Abril Rodriguez-Maldonado; Ariadna Medina-Benitez; Claudia Wong-Arambula; Ernesto Ramirez-Gonzalez.; Gisela Barrera-Badillo; Irma Lopez-Martinez; Joaquin Quiroz-Mercado; Lucia Hernandez-Rivas; Maribel Gonzalez-Villa; Natividad Cruz-Ortiz; Sergio Rangel-Guerrero; Tatiana Nunez-Garcia; Vanessa Rivero-Arredondo |
| EPI_ISL_2937835 | LESP Ciudad de Mexico | Instituto de Diagnostico y Referencia Epidemiologicos (INDRE) | Abril Rodriguez-Maldonado; Ariadna Medina-Benitez; Claudia Wong-Arambula; Ernesto Ramirez-Gonzalez.; Gisela Barrera-Badillo; Irma Lopez-Martinez; Joaquin Quiroz-Mercado; Lucia Hernandez-Rivas; Maribel Gonzalez-Villa; Natividad Cruz-Ortiz; Sergio Rangel-Guerrero; Tatiana Nunez-Garcia; Vanessa Rivero-Arredondo |
| EPI_ISL_1969244, EPI_ISL_1969245 | Lab RSUP DR Mohammad Hoesin Palembang | National Institute of Health Research and Development | Arie Ardiansyah Nugraha; Hana Apsari Pawestri; Hartanti Dian Ikawati; Kartika Dewi Puspa; Krisna Pangesti; Nelly Puspandari; Subangkit; Triyani Soekarso; Vivi Setiawaty |
| EPI_ISL_2923154 | Labo Analyses Med | National Reference Center for Viruses of Respiratory Infections, Institut Pasteur, Paris | Angela Brisebarre; Camille Capel; Christophe Malabat; Corinne Maufrais; DURIVAULT Jérôme; Etienne Simon-Lorière; Frédéric Lemoine; Hub Bioinformatique Biostatistiques; Louise Lefrançois; Marion Barbet; Maud Vanpeene; Méline Bizard; Sylvie Behillil; Sylvie Van der Werf; Vincent Enouf |
| EPI_ISL_3477384 | Laboratoire Coste | Department of Virology, Henri Mondor University Hospital, Assistance Publique Hôpitaux de Paris, Université Paris-Est Créteil, INSERM U955 | Alexandre Soulier; Christophe Rodriguez; Elisabeth Trawinski; Guillaume Gricourt; Jean-Michel Pawlotsky; Melissa N'Debi; Slim Fourati; Vanessa Demontant |
| EPI_ISL_3508340 | Laboratoire de santé publique du Québec | Laboratoire de santé publique du Québec | Guillaume Bourque; Ioannis Ragoussis; Jesse Shapiro; Mark Lathrop and Michel Roger on behalf of the CoVSeQ research group; Sandrine Moreira |
| EPI_ISL_3342662, EPI_ISL_3342663 | Laboratorio Central de Epidemiología (LCE) | Instituto de Biotecnología de la UNAM | ; Alejandra García-Gasca; Alejandra Hernández-Terán; Alejandro Sánchez-Flores; Alfredo Herrera-Estrella; Alicia Ocaña-Mondragón; Andreu Comas-García; Angel Gustavo Salas-Lais; Antonio Loza Román; Bernardo Martínez-Miguel; Blanca Taboada; Brenda Irasema Maldonado-Meza; Bruno Gómez-Gil; Carla Ivón Herrera-Najera; Carlos F. Arias; Celia Boukadida; Clara Esperanza Santacruz-Tinoco; Concepción Grajales-Muñiz; Consorcio Mexicano de Vigilancia Genómica (CoViGen-Mex). Authors (in alphabetical order): Julio Elias Alvarado-Yaah; Cristóbal Cháidez-Quiróz; Célida Duque Molina; Célida Martínez- Rodríguez; Daniel Fregoso-Rueda; Daniel Lira Morales; Eduardo Becerril-Vargas; Fernando Fontove-Herrera; Fidencio Mejía-Nepomuceno; Francisco Pulido; Gloria Elena Espinosa-Ayala; Gloria María Molina-Salinas; Gloria Vazquez; Hector Esteban Paz-Juárez; Hector Montoya-Fuentes; Helen Haydee Fernanda Ramírez-Plascencia; Irvin González-López; Jean Pierre González; Jesús Hernández; Joel Armando Vázquez-Pérez).; Jorge Salas-Hernández; José Antonio Enciso-Moreno; José Arturo Martínez-Orozco; José Esteban Muñoz-Medina; José de Jesús Nuñez-Contreras; Juan Bautista Chale-Dzul; Julissa Enciso-Ibarra; Kathia Elizabeth Tapia-Diaz; Luis Alberto Ochoa-Carrera; Margarita Matías-Florentino; Mario Mújica-Sánchez; Marissa Perez-Garcia; María Guadalupe Santiago-Mauricio; María Guadalupe de Jesús Mireles-Rivera; Nelly Sélem-Mojica; Pavel Isa; Ricardo Ciria Merce; Ricardo Grande; Rosa María Gutiérrez Rios; Santiago Ávila-Ríos; Selene Zárate; Susana Lopez; Verónica Mata-Haro; Victor Eduardo García-Arias; Víctor Hugo Borja-Aburto |
| EPI_ISL_3135898, EPI_ISL_3135900, EPI_ISL_3135901, EPI_ISL_3135902, EPI_ISL_3135903, EPI_ISL_3135904, EPI_ISL_3135925 | | | |
| see above | Laboratory Medicine and Molecular Diagnostics | Laboratory Medicine and Molecular Diagnostics | Dayakar Seetha; Heera R Pillai; Radhakrishna R Nair; Radhakrishnan R Nair; Sanughosh Kalpothodi |
| EPI_ISL_4047544 | Lighthouse Lab in Alderley Park | Wellcome Sanger Institute for the COVID-19 Genomics UK (COG-UK) Consortium | Cordelia Langford; David K. Jackson; Dominic Kwiatkowski; Ewan Harrison; Ian Johnston; Jacquelyn Wynn; Jeffrey Barrett; John Sillitoe on behalf of the Wellcome Sanger Institute COVID-19 Surveillance Team; Mairead Hyland; Roberto Amato; Sonia Goncalves; The Lighthouse Lab in Alderley Park and Alex Alderton |
| EPI_ISL_2998424 | Lighthouse Lab in Milton Keynes | Wellcome Sanger Institute for the COVID-19 Genomics UK (COG-UK) Consortium | Cordelia Langford; David K. Jackson; Dominic Kwiatkowski; Ewan Harrison; Ian Johnston; Jeffrey Barrett; John Sillitoe on behalf of the Wellcome Sanger Institute COVID-19 Surveillance Team; Roberto Amato; Sonia Goncalves; The Lighthouse Lab in Milton Keynes and Alex Alderton |
| EPI_ISL_1663548, EPI_ISL_1663549 | MGM Medical College, Jamshedpur | Institute of Life Sciences - INSACOG | Ajay Parida; Amol M. Kanampalliwar; Arup Ghosh; Atimukta Jha; INSACOG Consortium; Punit Prasad; Rajeeb Swain; Rupesh Dash; Safal Walia; Shifu Aggarwal; Sunil K. Raghav |
| EPI_ISL_2845215 | MVZ Labor Dr. Limbach & Kollegen GbR | Robert Koch Institute | |
| EPI_ISL_3730676 | Medics Labor AG | Clinical Bacteriology | Adrian Egli; Alfredo Mari; Fanny Wegner; Hans Hirsch; Helena MB Seth-Smith; Julia Bielicki; Karoline Leuzinger; Manuel Battegay; Tim Roloff |
| EPI_ISL_1663366, EPI_ISL_1663367, EPI_ISL_1663375, EPI_ISL_1663376 | NCCS, Pune | Institute of Life Sciences - INSACOG | Ajay Parida; Amol M. Kanampalliwar; Arup Ghosh; Atimukta Jha; INSACOG Consortium; Punit Prasad; Rajeeb Swain; Rupesh Dash; Safal Walia; Shifu Aggarwal; Sunil K. Raghav |
| EPI_ISL_1939986 | NIMHANS | inStem NCBS - INSACOG | Uma Ramakrishnan Dasaradhi Palakodeti Aswin SaiNarain |
| EPI_ISL_1544014, EPI_ISL_2162392, EPI_ISL_2162393, EPI_ISL_2162395, EPI_ISL_2162411 | National Centre For Cell Science | National Centre For Cell Science – INSACOG | Ajay Pillai; Dhiraj Paul; INSACOG Consortium team; Manoj Kumar Bhat; Mitali Inamdar; Mohak P Gujare; Shivang P. Bhanushali; Sonal Manik Chavan; Yogesh Shouche |
| EPI_ISL_2461256, EPI_ISL_2461258, EPI_ISL_2461509, EPI_ISL_2461515, EPI_ISL_2461539, EPI_ISL_2461564, EPI_ISL_2461788, EPI_ISL_2461789, EPI_ISL_2461822, EPI_ISL_2504284, EPI_ISL_2504861, EPI_ISL_2555827, EPI_ISL_2556527 | | | |
| see above | National Centre for Disease Control (NCDC) Biotechnology Division, Delhi | NCDC Delhi, Biotechnology Division INSACOG | Hema Gogia; Hemlata Lall; Kalaiarasan Ponnusamy; Mahesh S Dhar; Manoj K Singh; Meena Datta; Partha Rakshit; Preeti Madan; Priyanka Singh; Radhakrishnan V. S; Robin Marwal; Sandhya Kabra; Sujeet K Singh; Uma Sharma |
| EPI_ISL_3318923, EPI_ISL_3318987 | National Institute of Public Health | National Institute of Public Health | Alexander Nagy; Dusan Trnka; Helena Jirincova; Jaromira Vecerova; Timotej Suri |
| EPI_ISL_3825081 | Nevada State Public Health Laboratory | Nevada State Public Health Laboratory | Andrew Gorzalski; Mark Pandori |
| EPI_ISL_2921618, EPI_ISL_2921619, EPI_ISL_2921620, EPI_ISL_2921622 | New South Wales Health Pathology Royal Prince Alfred Hospital | Microbiology RPAH | Au, J.; Bull, R.; Deveson, I.; Foster, C.; Rawlinson, W.; Ruiz Silva, M.; Van Hal, S. |
| EPI_ISL_3421529, EPI_ISL_3571201, EPI_ISL_3772062, EPI_ISL_3772101, EPI_ISL_3772387, EPI_ISL_3772403 | Northumbria University / South Tees Hospitals NHS Foundation Trust / North Cumbria Integrated Care NHS Foundation Trust / North Tees and Hartlepool NHS Foundation Trust / Newcastle Hospitals NHS Foundation Trust | COVID-19 Genomics UK (COG-UK) Consortium | Andrew Nelson; Brendan Payne; Clive Graham; Darren L Smith; Debra Padgett; Edward Barton; Emma Swindells; Garren Scott; Gary Black; Gary Eltringham; Giles S Holt; Greg R Young; Jane Greenaway; Jennifer Collins; John Allan; Joshua Loh; Lynn Dover; Matthew Bashton; Mohammad A Tariq; Paul Baker; Sarah Essex; Steve Liggett; Wen C Yew; Yusri Taha |

| | | | |
|---|---|---|---|
| EPI_ISL_2920091 | OKMI | CMBG | Jan Svaton; Martina Lengerova; Matej Bezdicek; Monika Dolejska |
| EPI_ISL_4104000 | Pandit Raghunath Murmu Medical College, Baripada | Institute of Life Sciences-INSACOG | Ajay Parida; Amol M. Kanampalliwar; Arup Ghosh; Atimukta Jha; INSACOG Consortium; Punit Prasad; Rajeeb Swain; Rupesh Dash; Safal Walia; Sana Fatma; Shifu Aggarwal; Sunil K. Raghav |
| EPI_ISL_2341862, EPI_ISL_2341863, EPI_ISL_2341864, EPI_ISL_2341875, EPI_ISL_2341876, EPI_ISL_2341880, EPI_ISL_2341881 | | | |
| see above | Pt. Jawahar Lal Nehru Memorial Medical College, Raipur | Institute of Life Sciences - INSACOG | Ajay Parida; Amol M. Kanampalliwar; Arup Ghosh; Atimukta Jha; INSACOG Consortium; Omprakash Shiriwas; Punit Prasad; Rajeeb Swain; Rupesh Dash; Safal Walia; Sana Fatma; Shifu Aggarwal; Sunil K. Raghav |
| EPI_ISL_3178176, EPI_ISL_3289183, EPI_ISL_3289487, EPI_ISL_3571774 | Quadram Institute Bioscience | COVID-19 Genomics UK (COG-UK) Consortium | Alexander J Trotter; Alison E. Mather; Alp Aydin; Ana P. Tedim; Anastasia Kolyva; Andrew Bell; Andrew J. Page; Christopher Jeanes; Claire Stuart; Dave J. Baker; Ebenezer Foster-Nyarko; Gemma L. Kay; John Wain; Justin O'Grady; Leonardo de Oliveira Martins; Lewis G. Spurgin; Lindsay Coupland; Lizzie Meadows; Luke Bedford; Maria Diaz; Mark Webber; Martin Lott; Muhammed Yasir; Nabil-Fareed Alikhan; Ngozi Elumogo; Nicholas M. Thomson; Rachael Stanley; Rachel Gilroy; Reenesh Prakash; Rose K Davidson; Samir Dervisevic; Samuel Bloomfield; Sophie J. Prosolek; Steven Rudder; Thanh Le-Viet |
| EPI_ISL_3730776, EPI_ISL_3730782 | RSAB HARAPAN KITA | Genomik Solidaritas Indonesia Laboratorium | Anna Christina Brazia; Annisa Muthiah Sukirman; Anuraj Shankar; Ariel Pradipta; Dhahlia Agustina Cahyono; Dwi Oktavia; Gracia Felias Enos Korompis; Louisa Markus; Meutia Ayuputeri Kumaheri; Ngabila Salama; Reinhart Gabriel; Tiranti Vindha; Vania Gavrila Wikasa |
| EPI_ISL_3453376 | RUHS College of Medical Sciences | CSIR-Centre for Cellular and Molecular Biology - INSACOG | Amareshwar Vodapalli; Ara Sreenivas; Archana Bharadwaj Siva; B Himasri; Divya Tej Sowpati; Dr. Gaurav Dalela; Dr. Jitendra Panda; Dr. Nilofer Khayyam; Dr. Raja Babu Panwar; Dr. Rajeev Gupta; Dr. Ramesh Sharma; Dr. Sonali Sharma; Dr. Sudhanshu Kacker; Dr. Vaseem Naheed Baig; Jandhyala Sai Krishna; Karthik Bharadwaj Tallapaka; Lamuk Zaveri; Onkar Kulkarni; Payel Mukherjee; Priya Nurkurthy; Rakesh K Mishra; Shreekant Verma; Sofia Banu; Sumedha Avadhanula; Tulasi Nagabandi; Valli Nagalakshmi Undamatla; Vidhyadhari Methuku |
| EPI_ISL_4104093 | Rajendra Memorial Research Institute of Medical Sciences | Institute of Life Sciences-INSACOG | Ajay Parida; Amol M. Kanampalliwar; Arup Ghosh; Atimukta Jha; INSACOG Consortium; Punit Prasad; Rajeeb Swain; Rupesh Dash; Safal Walia; Sana Fatma; Shifu Aggarwal; Sunil K. Raghav |
| EPI_ISL_3293270, EPI_ISL_3773639 | Respiratory Virus Unit, Microbiology Services Colindale, Public Health England | COVID-19 Genomics UK (COG-UK) Consortium | PHE Covid Sequencing Team |
| EPI_ISL_1939974 | SUBRAMNAYA NAGAR | inStem NCBS - INSACOG | Uma Ramakrishnan Dasaradhi Palakodeti Aswin SaiNarain |
| EPI_ISL_2566445 | Singapore General Hospital | Department of Microbiology | Chayaporn Suphavilai; James Sim Heng Chiak; Karrie Ko; Kenneth Xin Long Chan; Kern Rei Chng; Kian Sing Chan; Kun Lee Lim; Lynette Oon; Niranjan Nagarajan; Nurdyana Abdul Rahman; Sui Sin Goh |
| EPI_ISL_3939757 | State Hygienic Laboratory at the University of Iowa | State Hygienic Laboratory at the University of Iowa | Alankar Kampoowale; Anna Yakos; Cindy Toll; Davis Rieckenberg; Erik Twait; Jeff Benfer; Kris Eveland; Kristen Zanon; Mariah Knutson; Mohammed Allam; Valerie Reeb; Wes Hottel |
| EPI_ISL_1914584, EPI_ISL_1914591 | State Virus Research and Diagnostic Laboratory (VRDL), AIIMS Raipur | State Virus Research and Diagnostic Laboratory (VRDL), AIIMS Raipur | Anudita Bhargava; Kuldeep Sharma; Priyanka Singh; Pushpendra Singh; Sanjay Singh Negi; Somya Sharma |
| EPI_ISL_4062630, EPI_ISL_4062647, EPI_ISL_4062648, EPI_ISL_4062649, EPI_ISL_4063070 | Sultan Qaboos University Hospital, Department of Microbiology & Immunology, Molecular Biology Section | KU Leuven, Rega Institute, Clinical and Epidemiological Virology | Abdullah Balkhair; Azza Alqayoudhi; Faiza Alnamaani; Fatma BaAlawi; Ishraq Al Kindi; Khuloud Al Maamari; Omar Balkhair; Piet Maes; Tony Wawina-Bokalanga; Zakaryia Almuharrmi |
| EPI_ISL_3791379 | Swedish national genomic surveillance program of SARS-CoV-2 | The Public Health Agency of Sweden | Alma Brolund; Maria Lind Karlberg; Maximilian Riess; Swedish national genomic surveillance program of SARS-CoV-2 |
| EPI_ISL_1969243 | Swissbel Hotel Airport | National Institute of Health Research and Development | Arie Ardiansyah Nugraha; Hana Apsari Pawestri; Hartanti Dian Ikawati; Kartika Dewi Puspa; Krisna Pangesti; Nelly Puspandari; Subangkit; Triyani Soekarso; Vivi Setiawaty |
| EPI_ISL_3153315 | TN DOH Lab Services | TN DOH Lab Services | Brian Selinsky; Karen Beasley-Maynard |
| EPI_ISL_3176701, EPI_ISL_3176831 | University College London, Great Ormond Street Hospital for Children NHS Foundation Trust, Imperial College Healthcare NHS Trust | COVID-19 Genomics UK (COG-UK) Consortium | Charlotte Williams; Helena Tutill; Judith Breuer; Marius Cotic; Mark Kristiansen; Nadua Bayzid; Patricia Dyal; Rachel Williams; Sergi Castellano; Sunando Roy |
| EPI_ISL_3980505, EPI_ISL_3980506, EPI_ISL_3980508, EPI_ISL_3980509 | University of Health and Allied Sciences (UHAS) COVID-19 Testing and Research Centre | University of Health and Allied Sciences (UHAS) COVID-19 Testing and Research Centre | John O. Gyapong and the UHAS COVID-19 Lab Team; Jones Gyamfi; Kwabena O. Duedu; Reuben Ayivor-Djanie |
| EPI_ISL_3132910, EPI_ISL_3132911, EPI_ISL_3263174, EPI_ISL_3263784 | Utah Public Health Laboratory | Utah Public Health Laboratory | Erin L. Young; Kelly F. Oakeson; Olinto Linares-Perdomo |
| EPI_ISL_1662284 | Veer Surendra Sai Institute of Medical Sciences and Research, Burla, Sambalpur | Institute of Life Sciences - INSACOG | Ajay Parida; Amol M. Kanampalliwar; Arup Ghosh; Atimukta Jha; INSACOG Consortium; Punit Prasad; Rajeeb Swain; Rupesh Dash; Safal Walia; Shifu Aggarwal; Sunil K. Raghav |
| EPI_ISL_2815935, EPI_ISL_3077206 | West of Scotland Specialist Virology Centre, NHSGGC / MRC-University of Glasgow Centre for Virus Research | COVID-19 Genomics UK (COG-UK) Consortium | Alasdair MacLean; Alice Broos; Ana da Silva Filipe; Antonia Ho; Daniel Mair; David L Robertson; Emma Thomson; Guy Mollett; Ioulia Tsatsani; James Shepherd; Jenna Nichols; Joseph Hughes; Kathy Li; Kathy Smollett; Kyriaki Nomikou; Lily Tong; Matthew Holden; Natasha Johnson; Rachel Blacow; Richard Orton; Rory Gunson; Sarah McDonald; Sharif Shaaban; Sreenu Vattipally; Stephen Carmichael |
| EPI_ISL_4055113 | Wielospecjalistyczny Szpital Wojewódzki | Wojewodzka Stacja Sanitarno-Epidemiologiczna w Gorzowie Wielkopolskim | Elżbieta Justyńska; Klaudia Kobendza-Wlodarczak; Patrycja Faberska and Marek Magol; Renata Siegel |
