## Supplementary material for "SARS-COV-2 δ variant drives the pandemic in India and Europe via two subvariants": Acknowledgement table on the GISAID genomes used in this study

We gratefully acknowledge the following Authors from the Originating laboratories responsible for obtaining the specimens, as well as the Submitting laboratories where the genome data were generated and shared via GISAID, on which this research is based.

All Submitters of data may be contacted directly via www.gisaid.org

Authors are sorted alphabetically.

| Accession ID | Originating Laboratory | Submitting Laboratory | Authors |
|---|---|---|---|
| EPI_ISL_4523720, EPI_ISL_4523725, EPI_ISL_4523735, EPI_ISL_4523739, EPI_ISL_4523743, EPI_ISL_4523747, EPI_ISL_4523756, EPI_ISL_4523757, EPI_ISL_4523766, EPI_ISL_4523772, EPI_ISL_4523776, EPI_ISL_4523779, EPI_ISL_4523787, EPI_ISL_4523788, EPI_ISL_4523801, EPI_ISL_4523810, EPI_ISL_4523813, EPI_ISL_4523818, EPI_ISL_4523834, EPI_ISL_4523849, EPI_ISL_4523851, EPI_ISL_4523856, EPI_ISL_4523868, EPI_ISL_4523869, EPI_ISL_4523879, EPI_ISL_4523885, EPI_ISL_4523892, EPI_ISL_4523896, EPI_ISL_4523898, EPI_ISL_4523909, EPI_ISL_4523913, EPI_ISL_4523921, EPI_ISL_4523926, EPI_ISL_4523938, EPI_ISL_4523945, EPI_ISL_4523951, EPI_ISL_4523959, EPI_ISL_4523961, EPI_ISL_4523963, EPI_ISL_4523978, EPI_ISL_4523981, EPI_ISL_4523983, EPI_ISL_4524006, EPI_ISL_4524015, EPI_ISL_4524018, EPI_ISL_4526261, EPI_ISL_4526263, EPI_ISL_4526264, EPI_ISL_4526267, EPI_ISL_4526268, EPI_ISL_4526269, EPI_ISL_4526271, EPI_ISL_4526272, EPI_ISL_4526273, EPI_ISL_4526274, EPI_ISL_4526276, EPI_ISL_4526288, EPI_ISL_4526290, EPI_ISL_4526292, EPI_ISL_4526295, EPI_ISL_4526298, EPI_ISL_4526302, EPI_ISL_4526304, EPI_ISL_4526305, EPI_ISL_4526307, EPI_ISL_4526309, EPI_ISL_4526311, EPI_ISL_4526312, EPI_ISL_4526313, EPI_ISL_4526314, EPI_ISL_4526318, EPI_ISL_4526319, EPI_ISL_4526320, EPI_ISL_4526322, EPI_ISL_4526323, EPI_ISL_4526325, EPI_ISL_4526329, EPI_ISL_4526334, EPI_ISL_4526335, EPI_ISL_4526337, EPI_ISL_4526340, EPI_ISL_4526343, EPI_ISL_4526346, EPI_ISL_4526348, EPI_ISL_4526350, EPI_ISL_4526353, EPI_ISL_4526354, EPI_ISL_4526355, EPI_ISL_4526357, EPI_ISL_4526358, EPI_ISL_4526360, EPI_ISL_4526362, EPI_ISL_4526368, EPI_ISL_4526374, EPI_ISL_4526386, EPI_ISL_4526389, EPI_ISL_4526390, EPI_ISL_4526392, EPI_ISL_4526393, EPI_ISL_4526396, EPI_ISL_4526397, EPI_ISL_4526398, EPI_ISL_4526399, EPI_ISL_4526406, EPI_ISL_4526407, EPI_ISL_4526408, EPI_ISL_4526409, EPI_ISL_4526411, EPI_ISL_4526413, EPI_ISL_4526414, EPI_ISL_4526417, EPI_ISL_4526423, EPI_ISL_4526424, EPI_ISL_4526426, EPI_ISL_4526433, EPI_ISL_4526440, EPI_ISL_4526441, EPI_ISL_4526446, EPI_ISL_4526447, EPI_ISL_4526448, EPI_ISL_4526461, EPI_ISL_4526468, EPI_ISL_4526471, EPI_ISL_4526473, EPI_ISL_4526475, EPI_ISL_4526476, EPI_ISL_4526477, EPI_ISL_4526478, EPI_ISL_4526482, EPI_ISL_4526486, EPI_ISL_4526487, EPI_ISL_4526492, EPI_ISL_4526494, EPI_ISL_4526496, EPI_ISL_4526497, EPI_ISL_4526498, EPI_ISL_4526500, EPI_ISL_4526503, EPI_ISL_4526506, EPI_ISL_4526508, EPI_ISL_4526510, EPI_ISL_4526513, EPI_ISL_4526516, EPI_ISL_4526517, EPI_ISL_4526519, EPI_ISL_4526523, EPI_ISL_4526524, EPI_ISL_4526525, EPI_ISL_4526530, EPI_ISL_4526531, EPI_ISL_4526532, EPI_ISL_4526533, EPI_ISL_4526534, EPI_ISL_4526536, EPI_ISL_4526537, EPI_ISL_4526538, EPI_ISL_4526546, EPI_ISL_4526547, EPI_ISL_4526549, EPI_ISL_4526554, EPI_ISL_4526557, EPI_ISL_4526560, EPI_ISL_4526562, EPI_ISL_4526563, EPI_ISL_4526564, EPI_ISL_4526565, EPI_ISL_4526566, EPI_ISL_4526568, EPI_ISL_4526574, EPI_ISL_4526577, EPI_ISL_4526578, EPI_ISL_4526583, EPI_ISL_4526588, EPI_ISL_4526590, EPI_ISL_4526591, EPI_ISL_4526594, EPI_ISL_4526597, EPI_ISL_4526601, EPI_ISL_4526603, EPI_ISL_4526614, EPI_ISL_4526616, EPI_ISL_4526618, EPI_ISL_4526620, EPI_ISL_4526623, EPI_ISL_4526626, EPI_ISL_4526632, EPI_ISL_4526635, EPI_ISL_4526643, EPI_ISL_4526645, EPI_ISL_4526646, EPI_ISL_4526653, EPI_ISL_4526655, EPI_ISL_4526660, EPI_ISL_4526669, EPI_ISL_4526671, EPI_ISL_4526672, EPI_ISL_4526675, EPI_ISL_4526679, EPI_ISL_4526684, EPI_ISL_4526693, EPI_ISL_4526703, EPI_ISL_4526704, EPI_ISL_4526712, EPI_ISL_4526714, EPI_ISL_4526716, EPI_ISL_4526722, EPI_ISL_4526734, EPI_ISL_4526738, EPI_ISL_4526742, EPI_ISL_4526748, EPI_ISL_4526750, EPI_ISL_4526760, EPI_ISL_4526765, EPI_ISL_4526782, EPI_ISL_4526785, EPI_ISL_4526787, EPI_ISL_4526789, EPI_ISL_4526796, EPI_ISL_4526797, EPI_ISL_4526798, EPI_ISL_4526802, EPI_ISL_4526813, EPI_ISL_4526821, EPI_ISL_4526830, EPI_ISL_4526831, EPI_ISL_4526832, EPI_ISL_4526835, EPI_ISL_4526837, EPI_ISL_4526841, EPI_ISL_4526842, EPI_ISL_4526851, EPI_ISL_4526853, EPI_ISL_4526854, EPI_ISL_4526858, EPI_ISL_4526862, EPI_ISL_4526867, EPI_ISL_4526883, EPI_ISL_4526884, EPI_ISL_4526891, EPI_ISL_4526894, EPI_ISL_4526896, EPI_ISL_4526900, EPI_ISL_4527743, EPI_ISL_4527746 | | | |
| see above | Berkshire and Surrey Pathology Services Lighthouse Laboratory | Wellcome Sanger Institute for the COVID-19 Genomics UK (COG-UK) Consortium | Berkshire and Surrey Pathology Services Lighthouse Laboratory and Alex Alderton; Cordelia Langford; David K. Jackson; Dominic Kwiatkowski; Ewan Harrison; Ian Johnston; Jeffrey Barrett; John Sillitoe on behalf of the Wellcome Sanger Institute COVID-19 Surveillance Team; Roberto Amato; Sonia Goncalves |
| EPI_ISL_4523729, EPI_ISL_4523732, EPI_ISL_4523733, EPI_ISL_4523745, EPI_ISL_4523748, EPI_ISL_4523758, EPI_ISL_4523782, EPI_ISL_4523789, EPI_ISL_4523793, EPI_ISL_4523797, EPI_ISL_4523798, EPI_ISL_4523804, EPI_ISL_4523809, EPI_ISL_4523826, EPI_ISL_4523830, EPI_ISL_4523833, EPI_ISL_4523840, EPI_ISL_4523841, EPI_ISL_4523844, EPI_ISL_4523847, EPI_ISL_4523848, EPI_ISL_4523850, EPI_ISL_4523857, EPI_ISL_4523858, EPI_ISL_4523862, EPI_ISL_4523871, EPI_ISL_4523872, EPI_ISL_4523876, EPI_ISL_4523886, EPI_ISL_4523888, EPI_ISL_4523889, EPI_ISL_4523890, EPI_ISL_4523900, EPI_ISL_4523901, EPI_ISL_4523910, EPI_ISL_4523911, EPI_ISL_4523915, EPI_ISL_4523918, EPI_ISL_4523924, EPI_ISL_4523932, EPI_ISL_4523939, EPI_ISL_4523943, EPI_ISL_4523944, EPI_ISL_4523968, EPI_ISL_4523970, EPI_ISL_4523973, EPI_ISL_4523974, EPI_ISL_4523993, EPI_ISL_4523995, EPI_ISL_4523996, EPI_ISL_4523997, EPI_ISL_4523998, EPI_ISL_4523999, EPI_ISL_4524000, EPI_ISL_4524007, EPI_ISL_4524009, EPI_ISL_4524020, EPI_ISL_4524022, EPI_ISL_4524026, EPI_ISL_4527737, EPI_ISL_4527739, EPI_ISL_4527741, EPI_ISL_4527744, EPI_ISL_4527745, EPI_ISL_4527748, EPI_ISL_4527749, EPI_ISL_4527750, EPI_ISL_4527751, EPI_ISL_4527752, EPI_ISL_4527753, EPI_ISL_4527755, EPI_ISL_4527756, EPI_ISL_4527757, EPI_ISL_4527758, EPI_ISL_4527759, EPI_ISL_4527762 | | | |
| see above | Health Services Laboratories | Wellcome Sanger Institute for the COVID-19 Genomics UK (COG-UK) Consortium | Cordelia Langford; David K. Jackson; Dominic Kwiatkowski; Ewan Harrison; Health Services Laboratories and Alex Alderton; Ian Johnston; Jeffrey Barrett; John Sillitoe on behalf of the Wellcome Sanger Institute COVID-19 Surveillance Team; Roberto Amato; Sonia Goncalves |
| EPI_ISL_4501824, EPI_ISL_4501882, EPI_ISL_4501903, EPI_ISL_4502020, EPI_ISL_4502030, EPI_ISL_4522389, EPI_ISL_4522391, EPI_ISL_4522396, EPI_ISL_4522405, EPI_ISL_4522409, EPI_ISL_4522429, EPI_ISL_4522435, EPI_ISL_4522444, EPI_ISL_4522448, EPI_ISL_4522450, EPI_ISL_4522454, EPI_ISL_4522460, EPI_ISL_4522462, EPI_ISL_4522472, EPI_ISL_4522473, EPI_ISL_4522476, EPI_ISL_4522478, EPI_ISL_4522484, EPI_ISL_4522496, EPI_ISL_4522501, EPI_ISL_4522502, EPI_ISL_4522507, EPI_ISL_4522516, EPI_ISL_4522523, EPI_ISL_4522528, EPI_ISL_4522532, EPI_ISL_4522547, EPI_ISL_4522556, EPI_ISL_4522558, EPI_ISL_4522559, EPI_ISL_4522561, EPI_ISL_4522587, EPI_ISL_4522588, EPI_ISL_4522593, EPI_ISL_4522600, EPI_ISL_4522601, EPI_ISL_4522607, EPI_ISL_4522608, EPI_ISL_4522617, EPI_ISL_4522619, EPI_ISL_4522626, EPI_ISL_4522634, EPI_ISL_4522644, EPI_ISL_4522653, EPI_ISL_4522655, EPI_ISL_4522658, EPI_ISL_4522660, EPI_ISL_4522663, EPI_ISL_4522671, EPI_ISL_4522680, EPI_ISL_4522682, EPI_ISL_4522685, EPI_ISL_4522711, EPI_ISL_4522712, EPI_ISL_4522713, EPI_ISL_4522718, EPI_ISL_4522719, EPI_ISL_4522943, EPI_ISL_4523058, EPI_ISL_4523059, EPI_ISL_4523079, EPI_ISL_4523157, EPI_ISL_4523164, EPI_ISL_4523204, EPI_ISL_4523216, EPI_ISL_4523235, EPI_ISL_4523241, EPI_ISL_4523247, EPI_ISL_4523287, EPI_ISL_4523293, EPI_ISL_4523334, EPI_ISL_4523345, EPI_ISL_4523356, EPI_ISL_4523375 | | | |
| see above | Integrated Covid Hub North East | Wellcome Sanger Institute for the COVID-19 Genomics UK (COG-UK) Consortium | Cordelia Langford; David K. Jackson; Dominic Kwiatkowski; Ewan Harrison; Ian Johnston; Integrated Covid Hub North East and Alex Alderton; Jeffrey Barrett; John Sillitoe on behalf of the Wellcome Sanger Institute COVID-19 Surveillance Team; Roberto Amato; Sonia Goncalves |
| EPI_ISL_4480546, EPI_ISL_4521407, EPI_ISL_4521411, EPI_ISL_4521414, EPI_ISL_4521429, EPI_ISL_4521430, EPI_ISL_4521438, EPI_ISL_4521446, EPI_ISL_4521450, EPI_ISL_4521452, EPI_ISL_4521457, EPI_ISL_4521464, EPI_ISL_4521466, EPI_ISL_4521468, EPI_ISL_4521474, EPI_ISL_4521476, EPI_ISL_4521481, EPI_ISL_4521488, EPI_ISL_4521493, EPI_ISL_4521501, EPI_ISL_4521502, EPI_ISL_4521504, EPI_ISL_4521514, EPI_ISL_4521516, EPI_ISL_4521517, EPI_ISL_4521521, EPI_ISL_4521522, EPI_ISL_4521531, EPI_ISL_4521532, EPI_ISL_4521536, EPI_ISL_4521538, EPI_ISL_4521542, EPI_ISL_4521543, EPI_ISL_4521546, EPI_ISL_4521549, EPI_ISL_4521554, EPI_ISL_4521559, EPI_ISL_4521560, EPI_ISL_4521562, EPI_ISL_4521583, EPI_ISL_4521589, EPI_ISL_4521603, EPI_ISL_4521608, EPI_ISL_4521616, EPI_ISL_4521634, EPI_ISL_4521635, EPI_ISL_4521639, EPI_ISL_4521642, EPI_ISL_4521646, EPI_ISL_4521651, EPI_ISL_4521652, EPI_ISL_4521664, EPI_ISL_4521686, EPI_ISL_4521694, EPI_ISL_4521704, EPI_ISL_4521706, EPI_ISL_4521712, EPI_ISL_4521729, EPI_ISL_4521731, EPI_ISL_4521732, EPI_ISL_4522068, EPI_ISL_4522077, EPI_ISL_4522079, EPI_ISL_4522089, EPI_ISL_4522093, EPI_ISL_4522100, EPI_ISL_4522102, EPI_ISL_4522117, EPI_ISL_4522128, EPI_ISL_4522136, EPI_ISL_4522143, EPI_ISL_4522147, EPI_ISL_4522153, EPI_ISL_4522170, EPI_ISL_4522174, EPI_ISL_4522179, EPI_ISL_4522183, EPI_ISL_4522191, EPI_ISL_4522215, EPI_ISL_4522216, EPI_ISL_4522218, EPI_ISL_4522274, EPI_ISL_4522280, EPI_ISL_4522287, EPI_ISL_4522288, EPI_ISL_4522289, EPI_ISL_4522297, EPI_ISL_4522300, EPI_ISL_4522305, EPI_ISL_4522313, EPI_ISL_4522325, EPI_ISL_4522341, EPI_ISL_4522343, EPI_ISL_4522346, EPI_ISL_4522348, EPI_ISL_4522352, EPI_ISL_4522353, EPI_ISL_4522363, EPI_ISL_4522372, EPI_ISL_4522383, EPI_ISL_4522386, EPI_ISL_4522438, EPI_ISL_4523386, EPI_ISL_4523389, EPI_ISL_4523394, EPI_ISL_4523398, EPI_ISL_4523399, EPI_ISL_4523400, EPI_ISL_4523402, EPI_ISL_4523404, EPI_ISL_4523406, EPI_ISL_4523418, EPI_ISL_4523423, EPI_ISL_4523427, EPI_ISL_4523430, EPI_ISL_4523434, EPI_ISL_4523435, EPI_ISL_4523436, EPI_ISL_4523439, EPI_ISL_4523449, EPI_ISL_4523450, EPI_ISL_4523451, EPI_ISL_4523453, EPI_ISL_4523456, EPI_ISL_4523478, EPI_ISL_4523481, EPI_ISL_4523482, EPI_ISL_4523484, EPI_ISL_4523486, EPI_ISL_4523487, EPI_ISL_4523488, EPI_ISL_4523489, EPI_ISL_4523491, EPI_ISL_4523499, EPI_ISL_4523500, EPI_ISL_4523501, EPI_ISL_4523504, EPI_ISL_4523505, EPI_ISL_4523506, EPI_ISL_4523509, EPI_ISL_4523510, EPI_ISL_4523513, EPI_ISL_4523523, EPI_ISL_4523524, EPI_ISL_4523526, EPI_ISL_4523531, EPI_ISL_4523532, EPI_ISL_4523535, EPI_ISL_4523541, EPI_ISL_4523545, EPI_ISL_4523548, EPI_ISL_4523554, EPI_ISL_4523564, EPI_ISL_4523568, EPI_ISL_4523569, EPI_ISL_4523572, EPI_ISL_4523573, EPI_ISL_4523583, EPI_ISL_4523593, EPI_ISL_4523597, EPI_ISL_4523599, EPI_ISL_4523601, EPI_ISL_4523603, EPI_ISL_4523609, EPI_ISL_4523611, EPI_ISL_4523612, EPI_ISL_4523613, EPI_ISL_4523619, EPI_ISL_4523620, EPI_ISL_4523624, EPI_ISL_4523627, EPI_ISL_4523628, EPI_ISL_4523642, EPI_ISL_4523646, EPI_ISL_4523652, EPI_ISL_4523655, EPI_ISL_4523656, EPI_ISL_4523659, EPI_ISL_4523662, EPI_ISL_4523664, EPI_ISL_4523669, EPI_ISL_4523672, EPI_ISL_4523680, EPI_ISL_4523688, EPI_ISL_4523690, EPI_ISL_4523699, EPI_ISL_4523701, EPI_ISL_4523703, EPI_ISL_4523705, EPI_ISL_4524301, EPI_ISL_4524303, EPI_ISL_4524306, EPI_ISL_4524307, EPI_ISL_4524320, EPI_ISL_4524321, EPI_ISL_4524330, EPI_ISL_4524342, EPI_ISL_4524343, EPI_ISL_4524345, EPI_ISL_4524350, EPI_ISL_4524355, EPI_ISL_4524360, EPI_ISL_4524368, EPI_ISL_4524374, EPI_ISL_4524376, EPI_ISL_4524379, EPI_ISL_4524389, EPI_ISL_4524394, EPI_ISL_4524415, EPI_ISL_4524416, EPI_ISL_4524417, EPI_ISL_4524425, EPI_ISL_4524428, EPI_ISL_4524441, EPI_ISL_4524450, EPI_ISL_4524455, EPI_ISL_4524459, EPI_ISL_4524466, EPI_ISL_4524468, EPI_ISL_4524473, EPI_ISL_4524481, EPI_ISL_4524482, EPI_ISL_4524483, EPI_ISL_4524501, EPI_ISL_4524502, EPI_ISL_4524509, EPI_ISL_4524511, EPI_ISL_4524523, EPI_ISL_4524524, EPI_ISL_4524528, EPI_ISL_4524529, EPI_ISL_4524539, EPI_ISL_4524547, EPI_ISL_4524550, EPI_ISL_4524553, EPI_ISL_4525914, EPI_ISL_4526630, EPI_ISL_4526652, EPI_ISL_4526661, EPI_ISL_4526663, EPI_ISL_4526664, EPI_ISL_4526732, EPI_ISL_4526745, EPI_ISL_4526753, EPI_ISL_4526758, EPI_ISL_4526834, EPI_ISL_4526839, EPI_ISL_4526892, EPI_ISL_4526904, EPI_ISL_4527048, EPI_ISL_4527049, EPI_ISL_4527052, EPI_ISL_4527055, EPI_ISL_4527059, EPI_ISL_4527070, EPI_ISL_4527084, EPI_ISL_4527085, EPI_ISL_4527088, EPI_ISL_4527094, EPI_ISL_4527097, EPI_ISL_4527117, EPI_ISL_4527125, EPI_ISL_4527129, EPI_ISL_4527132, EPI_ISL_4527138, EPI_ISL_4527142, EPI_ISL_4527145, EPI_ISL_4527146, EPI_ISL_4527150, EPI_ISL_4527151, EPI_ISL_4527159, EPI_ISL_4527164, EPI_ISL_4527169, EPI_ISL_4527175, EPI_ISL_4527191, EPI_ISL_4527193, EPI_ISL_4527194, EPI_ISL_4527197, EPI_ISL_4527206, EPI_ISL_4527208, EPI_ISL_4527212, EPI_ISL_4527213, EPI_ISL_4527214, EPI_ISL_4527227, EPI_ISL_4527229, EPI_ISL_4527234, EPI_ISL_4527236, EPI_ISL_4527240, EPI_ISL_4527242, EPI_ISL_4527244, EPI_ISL_4527246, EPI_ISL_4527247, EPI_ISL_4527251, EPI_ISL_4527255, EPI_ISL_4527257, EPI_ISL_4527266, EPI_ISL_4527273, EPI_ISL_4527281, EPI_ISL_4527288, EPI_ISL_4527292, EPI_ISL_4527296, EPI_ISL_4527297, EPI_ISL_4527299, EPI_ISL_4527303, EPI_ISL_4527304, EPI_ISL_4527306, EPI_ISL_4527311, EPI_ISL_4527312, EPI_ISL_4527319, EPI_ISL_4527321, EPI_ISL_4527324, EPI_ISL_4527327, EPI_ISL_4527329, EPI_ISL_4527341, EPI_ISL_4527343, EPI_ISL_4527348, EPI_ISL_4527350, EPI_ISL_4527351, EPI_ISL_4527354, EPI_ISL_4527357, EPI_ISL_4527359, EPI_ISL_4527369, EPI_ISL_4527376, EPI_ISL_4527381, EPI_ISL_4527383, EPI_ISL_4527385, EPI_ISL_4527390, EPI_ISL_4527394, EPI_ISL_4527398, EPI_ISL_4527413, EPI_ISL_4527418, EPI_ISL_4527420, EPI_ISL_4527423, EPI_ISL_4527424, EPI_ISL_4527429, EPI_ISL_4527430, EPI_ISL_4527435, EPI_ISL_4527441, EPI_ISL_4527449, EPI_ISL_4527450, EPI_ISL_4527455, EPI_ISL_4527456, EPI_ISL_4527460, EPI_ISL_4527464, EPI_ISL_4527465, EPI_ISL_4527466, EPI_ISL_4527487, EPI_ISL_4527494, EPI_ISL_4527496, EPI_ISL_4527500, EPI_ISL_4527503, EPI_ISL_4527508, EPI_ISL_4527511, EPI_ISL_4527514, EPI_ISL_4527520, EPI_ISL_4527523, EPI_ISL_4527533, EPI_ISL_4527537, EPI_ISL_4527540, EPI_ISL_4527545, EPI_ISL_4527549, EPI_ISL_4527554, EPI_ISL_4527555, EPI_ISL_4527556, EPI_ISL_4527557, EPI_ISL_4527562, EPI_ISL_4527576, EPI_ISL_4527577, EPI_ISL_4527580, EPI_ISL_4527583, EPI_ISL_4527584, EPI_ISL_4527590, EPI_ISL_4527593, EPI_ISL_4527599, EPI_ISL_4527601, EPI_ISL_4527606, EPI_ISL_4527615, EPI_ISL_4527619, EPI_ISL_4527620, EPI_ISL_4527623, EPI_ISL_4527624, EPI_ISL_4527631, EPI_ISL_4527641, EPI_ISL_4527642, EPI_ISL_4527644, EPI_ISL_4527650, EPI_ISL_4527651, EPI_ISL_4527662, EPI_ISL_4527665, EPI_ISL_4527671, EPI_ISL_4527674, EPI_ISL_4527675, EPI_ISL_4527684, EPI_ISL_4527685, EPI_ISL_4527687, EPI_ISL_4527690, EPI_ISL_4527701, EPI_ISL_4527706, EPI_ISL_4527710, EPI_ISL_4527724, EPI_ISL_4527798 | | | |
| see above | Lighthouse Lab in Alderley Park | Wellcome Sanger Institute for the COVID-19 Genomics UK (COG-UK) Consortium | Cordelia Langford; David K. Jackson; Dominic Kwiatkowski; Ewan Harrison; Ian Johnston; Jacquelyn Wynn; Jeffrey Barrett; John Sillitoe on behalf of the Wellcome Sanger Institute COVID-19 Surveillance Team; Mairead Hyland; Roberto Amato; Sonia Goncalves; The Lighthouse Lab in Alderley Park and Alex Alderton |
| EPI_ISL_4524047, EPI_ISL_4524074, EPI_ISL_4524078, EPI_ISL_4524125, EPI_ISL_4524177, EPI_ISL_4524233, EPI_ISL_4524262 | | | |
| see above | Lighthouse Lab in Glasgow | Wellcome Sanger Institute for the COVID-19 Genomics UK (COG-UK) Consortium | Anna Dominiczak and Alex Alderton; Carol Clugston; Cordelia Langford; David Gray; David K. Jackson; Dominic Kwiatkowski; Ewan Harrison; Harper VanSteenhouse; Ian Johnston; Jeffrey Barrett; John Sillitoe on behalf of the Wellcome Sanger Institute COVID-19 Surveillance Team; Roberto Amato; Sonia Goncalves; Yumi Kasai |
| EPI_ISL_4404374, EPI_ISL_4451289, EPI_ISL_4454335, EPI_ISL_4499483, EPI_ISL_4499486, EPI_ISL_4499494, EPI_ISL_4499495, EPI_ISL_4499499, EPI_ISL_4499502, EPI_ISL_4499503, EPI_ISL_4499506, EPI_ISL_4499508, EPI_ISL_4499510, EPI_ISL_4499511, EPI_ISL_4499512, EPI_ISL_4499514, EPI_ISL_4499520, EPI_ISL_4499521, EPI_ISL_4499522, EPI_ISL_4499526, EPI_ISL_4499527, EPI_ISL_4499528, EPI_ISL_4499529, EPI_ISL_4499530, EPI_ISL_4499534, EPI_ISL_4499535, EPI_ISL_4499538, EPI_ISL_4499539, EPI_ISL_4499541, EPI_ISL_4499542, EPI_ISL_4499545, EPI_ISL_4499556, EPI_ISL_4499558, EPI_ISL_4499559, EPI_ISL_4499561, EPI_ISL_4499565, EPI_ISL_4499574, EPI_ISL_4499576, EPI_ISL_4499578, EPI_ISL_4499580, EPI_ISL_4499582, EPI_ISL_4499583, EPI_ISL_4499586, EPI_ISL_4499588, EPI_ISL_4499590, EPI_ISL_4499592, EPI_ISL_4499597, EPI_ISL_4499601, EPI_ISL_4499602, EPI_ISL_4499604, EPI_ISL_4499609, EPI_ISL_4499611, EPI_ISL_4499612, EPI_ISL_4499613, EPI_ISL_4499614, EPI_ISL_4499615, EPI_ISL_4499618, EPI_ISL_4499619, EPI_ISL_4499620, EPI_ISL_4499621, EPI_ISL_4499622, EPI_ISL_4499623, EPI_ISL_4499625, EPI_ISL_4499626, EPI_ISL_4499629, EPI_ISL_4499630, EPI_ISL_4499631, EPI_ISL_4499638, EPI_ISL_4499641, EPI_ISL_4499642, EPI_ISL_4499643, EPI_ISL_4499648, EPI_ISL_4499650, EPI_ISL_4499651, EPI_ISL_4499652, EPI_ISL_4499657, EPI_ISL_4499660, EPI_ISL_4499661, EPI_ISL_4499663, EPI_ISL_4499668, EPI_ISL_4499669, EPI_ISL_4499670, EPI_ISL_4499674, EPI_ISL_4499675, EPI_ISL_4499676, EPI_ISL_4499678, EPI_ISL_4499679, EPI_ISL_4499680, EPI_ISL_4499682, EPI_ISL_4499684, EPI_ISL_4499686, EPI_ISL_4499690, EPI_ISL_4499692, EPI_ISL_4499694, EPI_ISL_4499702, EPI_ISL_4499704, EPI_ISL_4499707, EPI_ISL_4499708, EPI_ISL_4499709, EPI_ISL_4521402, EPI_ISL_4521404, EPI_ISL_4521406, EPI_ISL_4521408, EPI_ISL_4521413, EPI_ISL_4521415, EPI_ISL_4521420, EPI_ISL_4521435, EPI_ISL_4521440, EPI_ISL_4521444, EPI_ISL_4521445, EPI_ISL_4521459, EPI_ISL_4521461, EPI_ISL_4521465, EPI_ISL_4521470, EPI_ISL_4521471, EPI_ISL_4521472, EPI_ISL_4521477, EPI_ISL_4521478, EPI_ISL_4521483, EPI_ISL_4521487, EPI_ISL_4521491, EPI_ISL_4521496, EPI_ISL_4521497, EPI_ISL_4521518, EPI_ISL_4521523, EPI_ISL_4521527, EPI_ISL_4521528, EPI_ISL_4521533, EPI_ISL_4521534, EPI_ISL_4521539, EPI_ISL_4521556, EPI_ISL_4521561, EPI_ISL_4521563, EPI_ISL_4521565, EPI_ISL_4521569, EPI_ISL_4521570, EPI_ISL_4521572, EPI_ISL_4521575, EPI_ISL_4521580, EPI_ISL_4521581, EPI_ISL_4521584, EPI_ISL_4521588, EPI_ISL_4521592, EPI_ISL_4521594, EPI_ISL_4521595, EPI_ISL_4521596, EPI_ISL_4521599, EPI_ISL_4521602, EPI_ISL_4521606, EPI_ISL_4521612, EPI_ISL_4521613, EPI_ISL_4521614, EPI_ISL_4521615, EPI_ISL_4521623, EPI_ISL_4521627, EPI_ISL_4521631, EPI_ISL_4521636, EPI_ISL_4521637, EPI_ISL_4521638, EPI_ISL_4521643, EPI_ISL_4521644, EPI_ISL_4521647, EPI_ISL_4521653, EPI_ISL_4521654, EPI_ISL_4521655, EPI_ISL_4521658, EPI_ISL_4521659, EPI_ISL_4521661, EPI_ISL_4521669, EPI_ISL_4521671, EPI_ISL_4521676, EPI_ISL_4521677, EPI_ISL_4521683, EPI_ISL_4521685, EPI_ISL_4521690, EPI_ISL_4521693, EPI_ISL_4521695, EPI_ISL_4521696, EPI_ISL_4521699, EPI_ISL_4521701, EPI_ISL_4521705, EPI_ISL_4521711, EPI_ISL_4521713, EPI_ISL_4521714, EPI_ISL_4521719, EPI_ISL_4521721, EPI_ISL_4521728, EPI_ISL_4521734, EPI_ISL_4521738, EPI_ISL_4521742, EPI_ISL_4521744, EPI_ISL_4521745, EPI_ISL_4521746, EPI_ISL_4521747, EPI_ISL_4521748, EPI_ISL_4521751, EPI_ISL_4521756, EPI_ISL_4521758, EPI_ISL_4521763, EPI_ISL_4521766, EPI_ISL_4521779, EPI_ISL_4521783, EPI_ISL_4521786, EPI_ISL_4521790, EPI_ISL_4521792, EPI_ISL_4521795, EPI_ISL_4521796, EPI_ISL_4521799, EPI_ISL_4521802, EPI_ISL_4521808, EPI_ISL_4521809, EPI_ISL_4521812, EPI_ISL_4521815, EPI_ISL_4521826, EPI_ISL_4521830, EPI_ISL_4521833, EPI_ISL_4521834, EPI_ISL_4521844, EPI_ISL_4521849, EPI_ISL_4521861, EPI_ISL_4521870, EPI_ISL_4521873, EPI_ISL_4521876, EPI_ISL_4521881, EPI_ISL_4521895, EPI_ISL_4521904, EPI_ISL_4521910, EPI_ISL_4521913, EPI_ISL_4521915, EPI_ISL_4521917, EPI_ISL_4521922, EPI_ISL_4521930, EPI_ISL_4521941, EPI_ISL_4521957, EPI_ISL_4521962, EPI_ISL_4521969, EPI_ISL_4521974, EPI_ISL_4521979, EPI_ISL_4521981, EPI_ISL_4521984, EPI_ISL_4521989, EPI_ISL_4521992, EPI_ISL_4521994, EPI_ISL_4522002, EPI_ISL_4522005, EPI_ISL_4522006, EPI_ISL_4522007, EPI_ISL_4522010, EPI_ISL_4522011, EPI_ISL_4522014, EPI_ISL_4522018, EPI_ISL_4522020, EPI_ISL_4522024, EPI_ISL_4522027, EPI_ISL_4522028, EPI_ISL_4522031, EPI_ISL_4522032, EPI_ISL_4522033, EPI_ISL_4522040, EPI_ISL_4522044, EPI_ISL_4522048, EPI_ISL_4522053, EPI_ISL_4522058, EPI_ISL_4522059, EPI_ISL_4522074, EPI_ISL_4522075, EPI_ISL_4522076, EPI_ISL_4522083, EPI_ISL_4522086, EPI_ISL_4522090, EPI_ISL_4522092, EPI_ISL_4522095, EPI_ISL_4522096, EPI_ISL_4522098, EPI_ISL_4522099, EPI_ISL_4522103, EPI_ISL_4522104, EPI_ISL_4522110, EPI_ISL_4522112, EPI_ISL_4522114, EPI_ISL_4522115, EPI_ISL_4522116, EPI_ISL_4522130, EPI_ISL_4522132, EPI_ISL_4522134, EPI_ISL_4522135, EPI_ISL_4522139, EPI_ISL_4522140, EPI_ISL_4522141, EPI_ISL_4522145, EPI_ISL_4522150, EPI_ISL_4522151, EPI_ISL_4522160, EPI_ISL_4522165, EPI_ISL_4522166, EPI_ISL_4522171, EPI_ISL_4522172, EPI_ISL_4522176, EPI_ISL_4522178, EPI_ISL_4522181, EPI_ISL_4522182, EPI_ISL_4522187, EPI_ISL_4522188, EPI_ISL_4522197, EPI_ISL_4522198, EPI_ISL_4522199, EPI_ISL_4522202, EPI_ISL_4522203, EPI_ISL_4522223, EPI_ISL_4522225, EPI_ISL_4522226, EPI_ISL_4522228, EPI_ISL_4522229, EPI_ISL_4522231, EPI_ISL_4522234, EPI_ISL_4522235, EPI_ISL_4522237, EPI_ISL_4522238, EPI_ISL_4522239, EPI_ISL_4522240, EPI_ISL_4522246, EPI_ISL_4522250, EPI_ISL_4522255, EPI_ISL_4522257, EPI_ISL_4522259, EPI_ISL_4522260, EPI_ISL_4522263, EPI_ISL_4522266, EPI_ISL_4522267, EPI_ISL_4522269, EPI_ISL_4522270, EPI_ISL_4522273, EPI_ISL_4522279, EPI_ISL_4522282, EPI_ISL_4522284, EPI_ISL_4522292, EPI_ISL_4522302, EPI_ISL_4522312, EPI_ISL_4522318, EPI_ISL_4522321, EPI_ISL_4522322, EPI_ISL_4522323, EPI_ISL_4522326, EPI_ISL_4522329, EPI_ISL_4522330, EPI_ISL_4522332, EPI_ISL_4522334, EPI_ISL_4522337, EPI_ISL_4522338, EPI_ISL_4522339, EPI_ISL_4522340, EPI_ISL_4522347, EPI_ISL_4522350, EPI_ISL_4522356, EPI_ISL_4522357, EPI_ISL_4522358, EPI_ISL_4522367, EPI_ISL_4522370, EPI_ISL_4522374, EPI_ISL_4522375, EPI_ISL_4522376, EPI_ISL_4522378, EPI_ISL_4522379, EPI_ISL_4522382, EPI_ISL_4522384, EPI_ISL_4522388, EPI_ISL_4523384, EPI_ISL_4523388, EPI_ISL_4523390, EPI_ISL_4523395, EPI_ISL_4523407, EPI_ISL_4523417, EPI_ISL_4523422, EPI_ISL_4523428, EPI_ISL_4523429, EPI_ISL_4523441, EPI_ISL_4523444, EPI_ISL_4523445, EPI_ISL_4523455, EPI_ISL_4523458, EPI_ISL_4523468, EPI_ISL_4523473, EPI_ISL_4523476, EPI_ISL_4523483, EPI_ISL_4523492, EPI_ISL_4523503, EPI_ISL_4523507, EPI_ISL_4523508, EPI_ISL_4523511, EPI_ISL_4523515, EPI_ISL_4523516, EPI_ISL_4523517, EPI_ISL_4523521, EPI_ISL_4523527, EPI_ISL_4523528, EPI_ISL_4523540, EPI_ISL_4523550, EPI_ISL_4523552, EPI_ISL_4523555, EPI_ISL_4523560, EPI_ISL_4523562, EPI_ISL_4523570, EPI_ISL_4523575, EPI_ISL_4523576, EPI_ISL_4523589, EPI_ISL_4523591, EPI_ISL_4523604, EPI_ISL_4523610, EPI_ISL_4523615, EPI_ISL_4523641, EPI_ISL_4523658, EPI_ISL_4523660, EPI_ISL_4523663, EPI_ISL_4523679, EPI_ISL_4523684, EPI_ISL_4523687, EPI_ISL_4523695, EPI_ISL_4523697, EPI_ISL_4523698, EPI_ISL_4523704, EPI_ISL_4523707, EPI_ISL_4523714, EPI_ISL_4524033, EPI_ISL_4524035, EPI_ISL_4524037, EPI_ISL_4524042, EPI_ISL_4524044, EPI_ISL_4524050, EPI_ISL_4524053, EPI_ISL_4524054, EPI_ISL_4524055, EPI_ISL_4524056, EPI_ISL_4524057, EPI_ISL_4524059, EPI_ISL_4524067, EPI_ISL_4524071, EPI_ISL_4524075, EPI_ISL_4524076, EPI_ISL_4524077, EPI_ISL_4524082, EPI_ISL_4524083, EPI_ISL_4524090, EPI_ISL_4524095, EPI_ISL_4524102, EPI_ISL_4524106, EPI_ISL_4524114, EPI_ISL_4524115, EPI_ISL_4524116, EPI_ISL_4524118, EPI_ISL_4524121, EPI_ISL_4524124, EPI_ISL_4524126, EPI_ISL_4524127, EPI_ISL_4524133, EPI_ISL_4524144, EPI_ISL_4524147, EPI_ISL_4524151, EPI_ISL_4524155, EPI_ISL_4524156, EPI_ISL_4524159, EPI_ISL_4524169, EPI_ISL_4524170, EPI_ISL_4524174, EPI_ISL_4524175, EPI_ISL_4524176, EPI_ISL_4524181, EPI_ISL_4524185, EPI_ISL_4524186, EPI_ISL_4524187, EPI_ISL_4524188, EPI_ISL_4524189, EPI_ISL_4524190, EPI_ISL_4524194, EPI_ISL_4524195, EPI_ISL_4524197, EPI_ISL_4524198, EPI_ISL_4524210, EPI_ISL_4524213, EPI_ISL_4524218, EPI_ISL_4524220, EPI_ISL_4524222, EPI_ISL_4524223, EPI_ISL_4524228, EPI_ISL_4524232, EPI_ISL_4524239, EPI_ISL_4524242, EPI_ISL_4524253, EPI_ISL_4524254, EPI_ISL_4524260, EPI_ISL_4524261, EPI_ISL_4524275, EPI_ISL_4524278, EPI_ISL_4524292, EPI_ISL_4524293, EPI_ISL_4524296, EPI_ISL_4524298, EPI_ISL_4524302, EPI_ISL_4524304, EPI_ISL_4524309, EPI_ISL_4524312, EPI_ISL_4524315, EPI_ISL_4524318, EPI_ISL_4524322, EPI_ISL_4524327, EPI_ISL_4524338, EPI_ISL_4524341, EPI_ISL_4524344, EPI_ISL_4524347, EPI_ISL_4524348, EPI_ISL_4524351, EPI_ISL_4524354, EPI_ISL_4524365, EPI_ISL_4524366, EPI_ISL_4524370, EPI_ISL_4524371, EPI_ISL_4524375, EPI_ISL_4524382, EPI_ISL_4524383, EPI_ISL_4524384, EPI_ISL_4524388, EPI_ISL_4524390, EPI_ISL_4524391, EPI_ISL_4524395, EPI_ISL_4524399, EPI_ISL_4524403, EPI_ISL_4524406, EPI_ISL_4524413, EPI_ISL_4524418, EPI_ISL_4524421, EPI_ISL_4524426, EPI_ISL_4524429, EPI_ISL_4524432, EPI_ISL_4524433, EPI_ISL_4524434, EPI_ISL_4524435, EPI_ISL_4524437, EPI_ISL_4524442, EPI_ISL_4524449, EPI_ISL_4524451, EPI_ISL_4524457, EPI_ISL_4524460, EPI_ISL_4524463, EPI_ISL_4524464, EPI_ISL_4524469, EPI_ISL_4524470, EPI_ISL_4524471, EPI_ISL_4524472, EPI_ISL_4524475, EPI_ISL_4524476, EPI_ISL_4524477, EPI_ISL_4524478, EPI_ISL_4524479, EPI_ISL_4524484, EPI_ISL_4524486, EPI_ISL_4524488, EPI_ISL_4524489, EPI_ISL_4524490, EPI_ISL_4524491, EPI_ISL_4524492, EPI_ISL_4524495, EPI_ISL_4524496, EPI_ISL_4524498, EPI_ISL_4524499, EPI_ISL_4524500, EPI_ISL_4524504, EPI_ISL_4524505, EPI_ISL_4524506, EPI_ISL_4524507, EPI_ISL_4524512, EPI_ISL_4524513, EPI_ISL_4524514, EPI_ISL_4524515, EPI_ISL_4524518, EPI_ISL_4524519, EPI_ISL_4524522, EPI_ISL_4524525, EPI_ISL_4524526, EPI_ISL_4524530, EPI_ISL_4524532, EPI_ISL_4524534, EPI_ISL_4524538, EPI_ISL_4524540, EPI_ISL_4524543, EPI_ISL_4524546, EPI_ISL_4524549, EPI_ISL_4524551, EPI_ISL_4524552, EPI_ISL_4524853, EPI_ISL_4524854, EPI_ISL_4524859, EPI_ISL_4524864, EPI_ISL_4524872, EPI_ISL_4524874, EPI_ISL_4524880, EPI_ISL_4524881, EPI_ISL_4524884, EPI_ISL_4524885, EPI_ISL_4524886, EPI_ISL_4524888, EPI_ISL_4524890, EPI_ISL_4524891, EPI_ISL_4524892, EPI_ISL_4524893, EPI_ISL_4524904, EPI_ISL_4524905, EPI_ISL_4524906, EPI_ISL_4524909, EPI_ISL_4524919, EPI_ISL_4524921, EPI_ISL_4524924, EPI_ISL_4524927, EPI_ISL_4524929, EPI_ISL_4524931, EPI_ISL_4524932, EPI_ISL_4524933, EPI_ISL_4524935, EPI_ISL_4524938, EPI_ISL_4524941, EPI_ISL_4524943, EPI_ISL_4524945, EPI_ISL_4524947, EPI_ISL_4524948, EPI_ISL_4524949, EPI_ISL_4524950, EPI_ISL_4524955, EPI_ISL_4524956, EPI_ISL_4524958, EPI_ISL_4524965, EPI_ISL_4524970, EPI_ISL_4524971, EPI_ISL_4524972, EPI_ISL_4524976, EPI_ISL_4524977, EPI_ISL_4524978, EPI_ISL_4524979, EPI_ISL_4524980, EPI_ISL_4524981, EPI_ISL_4524982, EPI_ISL_4524986, EPI_ISL_4524987, EPI_ISL_4524990, EPI_ISL_4524991, EPI_ISL_4524992, EPI_ISL_4524994, EPI_ISL_4524995, EPI_ISL_4524996, EPI_ISL_4525000, EPI_ISL_4525001, EPI_ISL_4525002, EPI_ISL_4525005, EPI_ISL_4525006, EPI_ISL_4525010, EPI_ISL_4525012, EPI_ISL_4525013, EPI_ISL_4525015, EPI_ISL_4525018, EPI_ISL_4525023, EPI_ISL_4525026, EPI_ISL_4525028, EPI_ISL_4525029, EPI_ISL_4525031, EPI_ISL_4525032, EPI_ISL_4525034, EPI_ISL_4525036, EPI_ISL_4525040, EPI_ISL_4525042, EPI_ISL_4525044, EPI_ISL_4525046, EPI_ISL_4525925, EPI_ISL_4525929, EPI_ISL_4525931, EPI_ISL_4525938, EPI_ISL_4525948, EPI_ISL_4525949, EPI_ISL_4525956, EPI_ISL_4525963, EPI_ISL_4525965, EPI_ISL_4525973, EPI_ISL_4525974, EPI_ISL_4525975, EPI_ISL_4525977, EPI_ISL_4525981, EPI_ISL_4525994, EPI_ISL_4525997, EPI_ISL_4526007, EPI_ISL_4526011, EPI_ISL_4526014, EPI_ISL_4526017, EPI_ISL_4526019, EPI_ISL_4526021, EPI_ISL_4526032, EPI_ISL_4526035, EPI_ISL_4526040, EPI_ISL_4526042, EPI_ISL_4526044, EPI_ISL_4526048, EPI_ISL_4526066, EPI_ISL_4526068, EPI_ISL_4526075, EPI_ISL_4526076, EPI_ISL_4526077, EPI_ISL_4526079, EPI_ISL_4526084, EPI_ISL_4526086, EPI_ISL_4526090, EPI_ISL_4526095, EPI_ISL_4526103, EPI_ISL_4526104, EPI_ISL_4526111, EPI_ISL_4526114, EPI_ISL_4526126, EPI_ISL_4526127, EPI_ISL_4526131, EPI_ISL_4526134, EPI_ISL_4526137, EPI_ISL_4526144, EPI_ISL_4526147, EPI_ISL_4526150, EPI_ISL_4526151, EPI_ISL_4526156, EPI_ISL_4526157, EPI_ISL_4526159, EPI_ISL_4526162, EPI_ISL_4526165, EPI_ISL_4526168, EPI_ISL_4526179, EPI_ISL_4526182, EPI_ISL_4526184, EPI_ISL_4526194, EPI_ISL_4526195, EPI_ISL_4526198, EPI_ISL_4526204, EPI_ISL_4526205, EPI_ISL_4526214, EPI_ISL_4526215, EPI_ISL_4526219, EPI_ISL_4526223, EPI_ISL_4526228, EPI_ISL_4526230, EPI_ISL_4526232, EPI_ISL_4526233, EPI_ISL_4526234, EPI_ISL_4526235, EPI_ISL_4526236, EPI_ISL_4526239, EPI_ISL_4526253, EPI_ISL_4526257, EPI_ISL_4526260, EPI_ISL_4526270, EPI_ISL_4526278, EPI_ISL_4526281, EPI_ISL_4526282, EPI_ISL_4526286, EPI_ISL_4526291, EPI_ISL_4526299, EPI_ISL_4526300, EPI_ISL_4526306, EPI_ISL_4526308, EPI_ISL_4526315, EPI_ISL_4526321, EPI_ISL_4526326, EPI_ISL_4526327, EPI_ISL_4526330, EPI_ISL_4526338, EPI_ISL_4526342, EPI_ISL_4526344, EPI_ISL_4526352, EPI_ISL_4526359, EPI_ISL_4526365, EPI_ISL_4526366, EPI_ISL_4526367, EPI_ISL_4526376, EPI_ISL_4526378, EPI_ISL_4526379, EPI_ISL_4526381, EPI_ISL_4526382, EPI_ISL_4526383, EPI_ISL_4526384, EPI_ISL_4526385, EPI_ISL_4526387, EPI_ISL_4526400, EPI_ISL_4526403, EPI_ISL_4526404, EPI_ISL_4526419, EPI_ISL_4526420, EPI_ISL_4526421, EPI_ISL_4526427, EPI_ISL_4526437, EPI_ISL_4526438, EPI_ISL_4526439, EPI_ISL_4526442, EPI_ISL_4526443, EPI_ISL_4526444, EPI_ISL_4526445, EPI_ISL_4526450, EPI_ISL_4526456, EPI_ISL_4526457, EPI_ISL_4526460, EPI_ISL_4526463, EPI_ISL_4526467, EPI_ISL_4526470, EPI_ISL_4526472, EPI_ISL_4526474, EPI_ISL_4526479, EPI_ISL_4526481, EPI_ISL_4526485, EPI_ISL_4526488, EPI_ISL_4526489, EPI_ISL_4526490, EPI_ISL_4526493, EPI_ISL_4526499, EPI_ISL_4526502, EPI_ISL_4526515, EPI_ISL_4526521, EPI_ISL_4526522, EPI_ISL_4526526, EPI_ISL_4526527, EPI_ISL_4526535, EPI_ISL_4526544, EPI_ISL_4526545, EPI_ISL_4526551, EPI_ISL_4526555, EPI_ISL_4526556, EPI_ISL_4526558, EPI_ISL_4526561, EPI_ISL_4526567, EPI_ISL_4526570, EPI_ISL_4526573, EPI_ISL_4526575, EPI_ISL_4526576, EPI_ISL_4526579, EPI_ISL_4526582, EPI_ISL_4526584, EPI_ISL_4526585, EPI_ISL_4526593, EPI_ISL_4526604, EPI_ISL_4526608, EPI_ISL_4526617, EPI_ISL_4526619, EPI_ISL_4526622, EPI_ISL_4526637, EPI_ISL_4526640, EPI_ISL_4526641, EPI_ISL_4526642, EPI_ISL_4526649, EPI_ISL_4526657, EPI_ISL_4526658, EPI_ISL_4526659, EPI_ISL_4526662, EPI_ISL_4526670, EPI_ISL_4526678, EPI_ISL_4526680, EPI_ISL_4526682, EPI_ISL_4526683, EPI_ISL_4526686, EPI_ISL_4526691, EPI_ISL_4526694, EPI_ISL_4526697, EPI_ISL_4526698, EPI_ISL_4526700, EPI_ISL_4526702, EPI_ISL_4526709, EPI_ISL_4526717, EPI_ISL_4526723, EPI_ISL_4526728, EPI_ISL_4526729, EPI_ISL_4526730, EPI_ISL_4526744, EPI_ISL_4526746, EPI_ISL_4526747, EPI_ISL_4526756, EPI_ISL_4526759, EPI_ISL_4526762, EPI_ISL_4526764, EPI_ISL_4526769, EPI_ISL_4526770, EPI_ISL_4526773, EPI_ISL_4526774, EPI_ISL_4526783, EPI_ISL_4526790, EPI_ISL_4526803, EPI_ISL_4526804, EPI_ISL_4526806, EPI_ISL_4526810, EPI_ISL_4526812, EPI_ISL_4526818, EPI_ISL_4526819, EPI_ISL_4526822, EPI_ISL_4526823, EPI_ISL_4526829, EPI_ISL_4526844, EPI_ISL_4526852, EPI_ISL_4526856, EPI_ISL_4526859, EPI_ISL_4526860, EPI_ISL_4526861, EPI_ISL_4526863, EPI_ISL_4526864, EPI_ISL_4526866, EPI_ISL_4526871, EPI_ISL_4526873, EPI_ISL_4526877, EPI_ISL_4526880, EPI_ISL_4526881, EPI_ISL_4526888, EPI_ISL_4526889, EPI_ISL_4526899, EPI_ISL_4526906, EPI_ISL_4527051, EPI_ISL_4527054, EPI_ISL_4527073, EPI_ISL_4527079, EPI_ISL_4527082, EPI_ISL_4527086, EPI_ISL_4527091, EPI_ISL_4527092, EPI_ISL_4527099, EPI_ISL_4527101, EPI_ISL_4527104, EPI_ISL_4527107, EPI_ISL_4527114, EPI_ISL_4527119, EPI_ISL_4527127, EPI_ISL_4527133, EPI_ISL_4527135, EPI_ISL_4527143, EPI_ISL_4527156, EPI_ISL_4527163, EPI_ISL_4527166, EPI_ISL_4527173, EPI_ISL_4527178, EPI_ISL_4527181, EPI_ISL_4527198, EPI_ISL_4527201, EPI_ISL_4527211, EPI_ISL_4527225, EPI_ISL_4527226, EPI_ISL_4527232, EPI_ISL_4527243, EPI_ISL_4527250, EPI_ISL_4527253, EPI_ISL_4527256, EPI_ISL_4527259, EPI_ISL_4527260, EPI_ISL_4527277, EPI_ISL_4527278, EPI_ISL_4527294, EPI_ISL_4527298, EPI_ISL_4527301, EPI_ISL_4527308, EPI_ISL_4527313, EPI_ISL_4527316, EPI_ISL_4527323, EPI_ISL_4527326, EPI_ISL_4527330, EPI_ISL_4527333, EPI_ISL_4527337, | | | |

EPI_ISL_4527342, EPI_ISL_4527344, EPI_ISL_4527345, EPI_ISL_4527346, EPI_ISL_4527353, EPI_ISL_4527358, EPI_ISL_4527360, EPI_ISL_4527363, EPI_ISL_4527364, EPI_ISL_4527370, EPI_ISL_4527372, EPI_ISL_4527380, EPI_ISL_4527382, EPI_ISL_4527387, EPI_ISL_4527388, EPI_ISL_4527392, EPI_ISL_4527393, EPI_ISL_4527395, EPI_ISL_4527397, EPI_ISL_4527401, EPI_ISL_4527419, EPI_ISL_4527425, EPI_ISL_4527427, EPI_ISL_4527436, EPI_ISL_4527438, EPI_ISL_4527439, EPI_ISL_4527446, EPI_ISL_4527454, EPI_ISL_4527457, EPI_ISL_4527459, EPI_ISL_4527462, EPI_ISL_4527468, EPI_ISL_4527472, EPI_ISL_4527485, EPI_ISL_4527498, EPI_ISL_4527506, EPI_ISL_4527512, EPI_ISL_4527513, EPI_ISL_4527516, EPI_ISL_4527525, EPI_ISL_4527526, EPI_ISL_4527529, EPI_ISL_4527535, EPI_ISL_4527538, EPI_ISL_4527542, EPI_ISL_4527551, EPI_ISL_4527553, EPI_ISL_4527561, EPI_ISL_4527565, EPI_ISL_4527573, EPI_ISL_4527574, EPI_ISL_4527578, EPI_ISL_4527579, EPI_ISL_4527582, EPI_ISL_4527587, EPI_ISL_4527589, EPI_ISL_4527596, EPI_ISL_4527597, EPI_ISL_4527604, EPI_ISL_4527608, EPI_ISL_4527609, EPI_ISL_4527611, EPI_ISL_4527614, EPI_ISL_4527617, EPI_ISL_4527634, EPI_ISL_4527635, EPI_ISL_4527636, EPI_ISL_4527639, EPI_ISL_4527647, EPI_ISL_4527653, EPI_ISL_4527656, EPI_ISL_4527663, EPI_ISL_4527666, EPI_ISL_4527667, EPI_ISL_4527677, EPI_ISL_4527678, EPI_ISL_4527679, EPI_ISL_4527681, EPI_ISL_4527692, EPI_ISL_4527698, EPI_ISL_4527700, EPI_ISL_4527707, EPI_ISL_4527711, EPI_ISL_4527714, EPI_ISL_4527717, EPI_ISL_4527719, EPI_ISL_4527720, EPI_ISL_4527721, EPI_ISL_4527727, EPI_ISL_4527729, EPI_ISL_4527735, EPI_ISL_4527792, EPI_ISL_4527794, EPI_ISL_4527799, EPI_ISL_4551094, EPI_ISL_4551097, EPI_ISL_4551098, EPI_ISL_4551102

| | | | |
|---|---|---|---|
| see above | Lighthouse Lab in Milton Keynes | Wellcome Sanger Institute for the COVID-19 Genomics UK (COG-UK) Consortium | Cordelia Langford; David K. Jackson; Dominic Kwiatkowski; Ewan Harrison; Ian Johnston; Jeffrey Barrett; John Sillitoe on behalf of the Wellcome Sanger Institute COVID-19 Surveillance Team; Roberto Amato; Sonia Goncalves; The Lighthouse Lab in Milton Keynes and Alex Alderton |

EPI_ISL_4529752, EPI_ISL_4529753, EPI_ISL_4529754, EPI_ISL_4529755, EPI_ISL_4529756, EPI_ISL_4529757, EPI_ISL_4529758, EPI_ISL_4529759, EPI_ISL_4529761, EPI_ISL_4529762, EPI_ISL_4529763, EPI_ISL_4529764, EPI_ISL_4529765, EPI_ISL_4529766, EPI_ISL_4529767, EPI_ISL_4529768, EPI_ISL_4529769, EPI_ISL_4529770, EPI_ISL_4529771, EPI_ISL_4529772, EPI_ISL_4529773, EPI_ISL_4529774, EPI_ISL_4529775, EPI_ISL_4529777, EPI_ISL_4529778, EPI_ISL_4529779, EPI_ISL_4529780, EPI_ISL_4529781, EPI_ISL_4529782, EPI_ISL_4529783, EPI_ISL_4529784, EPI_ISL_4529785, EPI_ISL_4529786, EPI_ISL_4529787, EPI_ISL_4529788, EPI_ISL_4529789, EPI_ISL_4529790, EPI_ISL_4529791, EPI_ISL_4529792, EPI_ISL_4529793, EPI_ISL_4529794, EPI_ISL_4529795, EPI_ISL_4529796, EPI_ISL_4529797, EPI_ISL_4529798, EPI_ISL_4529799, EPI_ISL_4529800, EPI_ISL_4529801, EPI_ISL_4529802, EPI_ISL_4529803, EPI_ISL_4529804, EPI_ISL_4529805, EPI_ISL_4529806, EPI_ISL_4529807, EPI_ISL_4529808, EPI_ISL_4529809, EPI_ISL_4529810, EPI_ISL_4529811, EPI_ISL_4529812, EPI_ISL_4529813, EPI_ISL_4529814, EPI_ISL_4529815, EPI_ISL_4529816, EPI_ISL_4529817, EPI_ISL_4529818, EPI_ISL_4529819, EPI_ISL_4529820, EPI_ISL_4529821, EPI_ISL_4529822, EPI_ISL_4529823, EPI_ISL_4529824, EPI_ISL_4529825, EPI_ISL_4529826, EPI_ISL_4529827, EPI_ISL_4529828, EPI_ISL_4529829, EPI_ISL_4529830, EPI_ISL_4529831, EPI_ISL_4529832, EPI_ISL_4529833, EPI_ISL_4529834, EPI_ISL_4529835, EPI_ISL_4529836, EPI_ISL_4529837, EPI_ISL_4529838, EPI_ISL_4529839, EPI_ISL_4529840, EPI_ISL_4529841, EPI_ISL_4529842, EPI_ISL_4529843, EPI_ISL_4529844, EPI_ISL_4529845, EPI_ISL_4529846, EPI_ISL_4529847, EPI_ISL_4529848, EPI_ISL_4529849, EPI_ISL_4529850, EPI_ISL_4529851, EPI_ISL_4529852, EPI_ISL_4529853, EPI_ISL_4529854, EPI_ISL_4529855, EPI_ISL_4529856, EPI_ISL_4529857, EPI_ISL_4529858, EPI_ISL_4529859, EPI_ISL_4529860, EPI_ISL_4529861, EPI_ISL_4529862, EPI_ISL_4529863, EPI_ISL_4529865, EPI_ISL_4529866, EPI_ISL_4529867, EPI_ISL_4529868, EPI_ISL_4529869, EPI_ISL_4529870, EPI_ISL_4529871, EPI_ISL_4529872, EPI_ISL_4529873, EPI_ISL_4529874, EPI_ISL_4529875, EPI_ISL_4529876, EPI_ISL_4529878, EPI_ISL_4529879, EPI_ISL_4529880, EPI_ISL_4529881, EPI_ISL_4529882, EPI_ISL_4529883, EPI_ISL_4529884, EPI_ISL_4529885, EPI_ISL_4529886, EPI_ISL_4529887, EPI_ISL_4529888, EPI_ISL_4529889, EPI_ISL_4529890, EPI_ISL_4529891, EPI_ISL_4529892, EPI_ISL_4529893, EPI_ISL_4529894, EPI_ISL_4529895, EPI_ISL_4529896, EPI_ISL_4529897, EPI_ISL_4529898, EPI_ISL_4529899, EPI_ISL_4529900, EPI_ISL_4529901, EPI_ISL_4529902, EPI_ISL_4529903, EPI_ISL_4529904, EPI_ISL_4529905, EPI_ISL_4529906, EPI_ISL_4529907, EPI_ISL_4529908, EPI_ISL_4529910, EPI_ISL_4529911, EPI_ISL_4529912, EPI_ISL_4529913, EPI_ISL_4529914, EPI_ISL_4529915, EPI_ISL_4529916, EPI_ISL_4529917, EPI_ISL_4529918, EPI_ISL_4529919, EPI_ISL_4529920, EPI_ISL_4529921, EPI_ISL_4529922, EPI_ISL_4529923, EPI_ISL_4529924, EPI_ISL_4529925, EPI_ISL_4529926, EPI_ISL_4529927, EPI_ISL_4529928, EPI_ISL_4529929, EPI_ISL_4529930, EPI_ISL_4529931, EPI_ISL_4529932, EPI_ISL_4529933, EPI_ISL_4529934, EPI_ISL_4529935, EPI_ISL_4529936, EPI_ISL_4529937, EPI_ISL_4529938, EPI_ISL_4529939, EPI_ISL_4529940, EPI_ISL_4529941, EPI_ISL_4529942, EPI_ISL_4529943, EPI_ISL_4529944, EPI_ISL_4529945, EPI_ISL_4529946, EPI_ISL_4529947, EPI_ISL_4529948, EPI_ISL_4529949, EPI_ISL_4529950, EPI_ISL_4529951, EPI_ISL_4529952, EPI_ISL_4529953, EPI_ISL_4529954, EPI_ISL_4529956, EPI_ISL_4529957, EPI_ISL_4529958, EPI_ISL_4529959, EPI_ISL_4529960, EPI_ISL_4529961, EPI_ISL_4529962, EPI_ISL_4529963, EPI_ISL_4529964, EPI_ISL_4529965, EPI_ISL_4529966, EPI_ISL_4529967, EPI_ISL_4529968, EPI_ISL_4529969, EPI_ISL_4529970, EPI_ISL_4529971, EPI_ISL_4529972, EPI_ISL_4529973, EPI_ISL_4529974, EPI_ISL_4529975, EPI_ISL_4529976, EPI_ISL_4529977, EPI_ISL_4529978, EPI_ISL_4529979, EPI_ISL_4529980, EPI_ISL_4529981, EPI_ISL_4530103, EPI_ISL_4530105, EPI_ISL_4530107, EPI_ISL_4530108, EPI_ISL_4530114, EPI_ISL_4530116, EPI_ISL_4530122, EPI_ISL_4530125, EPI_ISL_4530127, EPI_ISL_4530129, EPI_ISL_4530130, EPI_ISL_4530131, EPI_ISL_4530132, EPI_ISL_4530133, EPI_ISL_4530137, EPI_ISL_4530140, EPI_ISL_4530141, EPI_ISL_4530147, EPI_ISL_4530157, EPI_ISL_4530162, EPI_ISL_4530163, EPI_ISL_4530171, EPI_ISL_4530173, EPI_ISL_4530181, EPI_ISL_4530184, EPI_ISL_4530189, EPI_ISL_4530195, EPI_ISL_4530196, EPI_ISL_4530205, EPI_ISL_4530206, EPI_ISL_4530208, EPI_ISL_4530218, EPI_ISL_4530282, EPI_ISL_4530283, EPI_ISL_4530285, EPI_ISL_4530286, EPI_ISL_4530287, EPI_ISL_4530288, EPI_ISL_4530289, EPI_ISL_4530290, EPI_ISL_4530291, EPI_ISL_4530292, EPI_ISL_4530293, EPI_ISL_4530294, EPI_ISL_4530295, EPI_ISL_4530296, EPI_ISL_4530297, EPI_ISL_4530298, EPI_ISL_4530299, EPI_ISL_4530300, EPI_ISL_4530301, EPI_ISL_4530302, EPI_ISL_4530303, EPI_ISL_4530304, EPI_ISL_4530305, EPI_ISL_4530306, EPI_ISL_4530307, EPI_ISL_4530308, EPI_ISL_4530309, EPI_ISL_4530310, EPI_ISL_4530311, EPI_ISL_4530312, EPI_ISL_4530313, EPI_ISL_4530314, EPI_ISL_4530315, EPI_ISL_4530316, EPI_ISL_4530317, EPI_ISL_4530318, EPI_ISL_4530319, EPI_ISL_4530320, EPI_ISL_4530321, EPI_ISL_4530322, EPI_ISL_4530323, EPI_ISL_4530324, EPI_ISL_4530325, EPI_ISL_4530326, EPI_ISL_4530327, EPI_ISL_4530328, EPI_ISL_4530329, EPI_ISL_4530330, EPI_ISL_4530331, EPI_ISL_4530332, EPI_ISL_4530333, EPI_ISL_4530334, EPI_ISL_4530335, EPI_ISL_4530336, EPI_ISL_4530337, EPI_ISL_4530338, EPI_ISL_4530339, EPI_ISL_4530340, EPI_ISL_4530341, EPI_ISL_4530342, EPI_ISL_4530343, EPI_ISL_4530344, EPI_ISL_4530345, EPI_ISL_4530346, EPI_ISL_4530347, EPI_ISL_4530348, EPI_ISL_4530349, EPI_ISL_4530350, EPI_ISL_4530351, EPI_ISL_4530352, EPI_ISL_4530353, EPI_ISL_4530354, EPI_ISL_4530355, EPI_ISL_4530356, EPI_ISL_4530357, EPI_ISL_4530358, EPI_ISL_4530359, EPI_ISL_4530360, EPI_ISL_4530361, EPI_ISL_4530362, EPI_ISL_4530403, EPI_ISL_4530404, EPI_ISL_4530405, EPI_ISL_4530407, EPI_ISL_4530408, EPI_ISL_4530409, EPI_ISL_4530410, EPI_ISL_4530411, EPI_ISL_4530412, EPI_ISL_4530413, EPI_ISL_4530414, EPI_ISL_4530415, EPI_ISL_4530416, EPI_ISL_4530417, EPI_ISL_4530418, EPI_ISL_4530419, EPI_ISL_4530420, EPI_ISL_4530421, EPI_ISL_4530422, EPI_ISL_4530423, EPI_ISL_4530424, EPI_ISL_4530425, EPI_ISL_4530426, EPI_ISL_4530427, EPI_ISL_4530428, EPI_ISL_4530429, EPI_ISL_4530430, EPI_ISL_4530431, EPI_ISL_4530432, EPI_ISL_4530433, EPI_ISL_4530434, EPI_ISL_4530435, EPI_ISL_4530436, EPI_ISL_4530437, EPI_ISL_4530438, EPI_ISL_4530439, EPI_ISL_4530440, EPI_ISL_4530441, EPI_ISL_4530442, EPI_ISL_4530443, EPI_ISL_4530445, EPI_ISL_4530446, EPI_ISL_4530448, EPI_ISL_4530449, EPI_ISL_4530450, EPI_ISL_4530451, EPI_ISL_4530452, EPI_ISL_4530454, EPI_ISL_4530455, EPI_ISL_4530456, EPI_ISL_4530457, EPI_ISL_4530458, EPI_ISL_4530459, EPI_ISL_4530460, EPI_ISL_4530461, EPI_ISL_4530462, EPI_ISL_4530463, EPI_ISL_4530464, EPI_ISL_4530465, EPI_ISL_4530466, EPI_ISL_4530467, EPI_ISL_4530468, EPI_ISL_4530469, EPI_ISL_4530470, EPI_ISL_4530471, EPI_ISL_4530472, EPI_ISL_4530473, EPI_ISL_4530474, EPI_ISL_4530476, EPI_ISL_4530477, EPI_ISL_4530478, EPI_ISL_4530479, EPI_ISL_4530480, EPI_ISL_4530481, EPI_ISL_4530482, EPI_ISL_4530483, EPI_ISL_4530484, EPI_ISL_4530485, EPI_ISL_4530486, EPI_ISL_4530487, EPI_ISL_4530489, EPI_ISL_4530490, EPI_ISL_4530492, EPI_ISL_4530493, EPI_ISL_4530496, EPI_ISL_4530497, EPI_ISL_4530498, EPI_ISL_4530499, EPI_ISL_4530500, EPI_ISL_4530501, EPI_ISL_4530502, EPI_ISL_4530503, EPI_ISL_4530504, EPI_ISL_4530505, EPI_ISL_4530506, EPI_ISL_4530507, EPI_ISL_4530508, EPI_ISL_4530509, EPI_ISL_4530510, EPI_ISL_4530512, EPI_ISL_4530513, EPI_ISL_4530514, EPI_ISL_4530515, EPI_ISL_4530516, EPI_ISL_4530517, EPI_ISL_4530518, EPI_ISL_4530519, EPI_ISL_4530520, EPI_ISL_4530521, EPI_ISL_4530523, EPI_ISL_4530524, EPI_ISL_4530525, EPI_ISL_4530526, EPI_ISL_4530528, EPI_ISL_4530529, EPI_ISL_4530530, EPI_ISL_4530531, EPI_ISL_4530532, EPI_ISL_4530533, EPI_ISL_4530534, EPI_ISL_4530535, EPI_ISL_4530536, EPI_ISL_4530537, EPI_ISL_4530538, EPI_ISL_4530539, EPI_ISL_4530540, EPI_ISL_4530541, EPI_ISL_4530542, EPI_ISL_4530543, EPI_ISL_4530544, EPI_ISL_4530545, EPI_ISL_4530546, EPI_ISL_4530548, EPI_ISL_4530549, EPI_ISL_4530550, EPI_ISL_4530551, EPI_ISL_4530553, EPI_ISL_4530554, EPI_ISL_4530555, EPI_ISL_4530556, EPI_ISL_4530557, EPI_ISL_4530558, EPI_ISL_4530559, EPI_ISL_4530560, EPI_ISL_4530561, EPI_ISL_4530563, EPI_ISL_4530564, EPI_ISL_4530565, EPI_ISL_4530566, EPI_ISL_4530567, EPI_ISL_4530568, EPI_ISL_4530569, EPI_ISL_4530570, EPI_ISL_4530571, EPI_ISL_4530572, EPI_ISL_4530573, EPI_ISL_4530574, EPI_ISL_4530575, EPI_ISL_4530576, EPI_ISL_4530577, EPI_ISL_4530578, EPI_ISL_4530579, EPI_ISL_4530580, EPI_ISL_4530581, EPI_ISL_4530582, EPI_ISL_4530583, EPI_ISL_4530584, EPI_ISL_4530585, EPI_ISL_4530587, EPI_ISL_4530588, EPI_ISL_4530589, EPI_ISL_4530590, EPI_ISL_4530592, EPI_ISL_4530593, EPI_ISL_4530594, EPI_ISL_4530595, EPI_ISL_4530596, EPI_ISL_4530597, EPI_ISL_4530598, EPI_ISL_4530599, EPI_ISL_4530600, EPI_ISL_4530601, EPI_ISL_4530603, EPI_ISL_4530604, EPI_ISL_4530605, EPI_ISL_4530606, EPI_ISL_4530607, EPI_ISL_4530608, EPI_ISL_4530609, EPI_ISL_4530610, EPI_ISL_4530611, EPI_ISL_4530612, EPI_ISL_4530613, EPI_ISL_4530614, EPI_ISL_4530615, EPI_ISL_4530616, EPI_ISL_4530617, EPI_ISL_4530619, EPI_ISL_4530620, EPI_ISL_4530621, EPI_ISL_4530622, EPI_ISL_4530623, EPI_ISL_4530624, EPI_ISL_4530625, EPI_ISL_4530626, EPI_ISL_4530627, EPI_ISL_4530628, EPI_ISL_4530629, EPI_ISL_4530630, EPI_ISL_4530631, EPI_ISL_4530632, EPI_ISL_4530633, EPI_ISL_4530634, EPI_ISL_4530635, EPI_ISL_4530636, EPI_ISL_4530638, EPI_ISL_4530639, EPI_ISL_4530640, EPI_ISL_4530641, EPI_ISL_4530642, EPI_ISL_4530643, EPI_ISL_4530644, EPI_ISL_4530645, EPI_ISL_4530646, EPI_ISL_4530647, EPI_ISL_4530648, EPI_ISL_4530649, EPI_ISL_4530650, EPI_ISL_4530651, EPI_ISL_4530652, EPI_ISL_4530653, EPI_ISL_4530654, EPI_ISL_4530655, EPI_ISL_4530656, EPI_ISL_4530658, EPI_ISL_4530659, EPI_ISL_4530661, EPI_ISL_4530662, EPI_ISL_4530663, EPI_ISL_4530664, EPI_ISL_4530665, EPI_ISL_4530666, EPI_ISL_4530667, EPI_ISL_4530668, EPI_ISL_4530669, EPI_ISL_4530670, EPI_ISL_4530671, EPI_ISL_4530672, EPI_ISL_4530673, EPI_ISL_4530674, EPI_ISL_4530675, EPI_ISL_4530676, EPI_ISL_4530677, EPI_ISL_4530678, EPI_ISL_4530679, EPI_ISL_4530680, EPI_ISL_4530681, EPI_ISL_4530682, EPI_ISL_4530683, EPI_ISL_4530684, EPI_ISL_4530685, EPI_ISL_4530686, EPI_ISL_4530687, EPI_ISL_4530688, EPI_ISL_4530689, EPI_ISL_4530690, EPI_ISL_4530691, EPI_ISL_4530692, EPI_ISL_4530693, EPI_ISL_4530694, EPI_ISL_4530695, EPI_ISL_4530696, EPI_ISL_4530698, EPI_ISL_4530699, EPI_ISL_4530700, EPI_ISL_4530701, EPI_ISL_4530702, EPI_ISL_4530703, EPI_ISL_4530704, EPI_ISL_4530705, EPI_ISL_4530706, EPI_ISL_4530707, EPI_ISL_4530708, EPI_ISL_4530709, EPI_ISL_4530710, EPI_ISL_4530711, EPI_ISL_4530712, EPI_ISL_4530713, EPI_ISL_4530714, EPI_ISL_4530715, EPI_ISL_4530716, EPI_ISL_4530717, EPI_ISL_4530718, EPI_ISL_4530719, EPI_ISL_4530720, EPI_ISL_4530721, EPI_ISL_4530722, EPI_ISL_4530723, EPI_ISL_4530724, EPI_ISL_4530725, EPI_ISL_4530726, EPI_ISL_4530727, EPI_ISL_4530728, EPI_ISL_4530729, EPI_ISL_4530730, EPI_ISL_4530731, EPI_ISL_4530732, EPI_ISL_4530733, EPI_ISL_4530734, EPI_ISL_4530735, EPI_ISL_4530736, EPI_ISL_4530737, EPI_ISL_4530738, EPI_ISL_4530739, EPI_ISL_4530741, EPI_ISL_4530742, EPI_ISL_4530743, EPI_ISL_4530744, EPI_ISL_4530745, EPI_ISL_4530746, EPI_ISL_4530747, EPI_ISL_4530748, EPI_ISL_4530749, EPI_ISL_4530750, EPI_ISL_4530751, EPI_ISL_4530752, EPI_ISL_4530753, EPI_ISL_4530754, EPI_ISL_4530755, EPI_ISL_4530756, EPI_ISL_4530757, EPI_ISL_4530758, EPI_ISL_4530759, EPI_ISL_4530760, EPI_ISL_4530761, EPI_ISL_4530762, EPI_ISL_4530763, EPI_ISL_4530764, EPI_ISL_4530765, EPI_ISL_4530766, EPI_ISL_4530767, EPI_ISL_4530768, EPI_ISL_4530769, EPI_ISL_4530770, EPI_ISL_4530771, EPI_ISL_4530772, EPI_ISL_4530773, EPI_ISL_4530774, EPI_ISL_4530775, EPI_ISL_4530776, EPI_ISL_4530777, EPI_ISL_4530778, EPI_ISL_4530779, EPI_ISL_4530780, EPI_ISL_4530781, EPI_ISL_4530782, EPI_ISL_4530783, EPI_ISL_4530784, EPI_ISL_4530785, EPI_ISL_4530786, EPI_ISL_4530787, EPI_ISL_4530788, EPI_ISL_4530789, EPI_ISL_4530790, EPI_ISL_4530791, EPI_ISL_4530792, EPI_ISL_4530793, EPI_ISL_4530794, EPI_ISL_4530795, EPI_ISL_4530796, EPI_ISL_4530797, EPI_ISL_4530798, EPI_ISL_4530799, EPI_ISL_4530800, EPI_ISL_4530801, EPI_ISL_4530802, EPI_ISL_4530803, EPI_ISL_4530804, EPI_ISL_4530805, EPI_ISL_4530807, EPI_ISL_4530808, EPI_ISL_4530809, EPI_ISL_4530810, EPI_ISL_4530811, EPI_ISL_4530812, EPI_ISL_4530813, EPI_ISL_4530814, EPI_ISL_4530815, EPI_ISL_4530817, EPI_ISL_4530818, EPI_ISL_4530819, EPI_ISL_4530820, EPI_ISL_4530821, EPI_ISL_4530822, EPI_ISL_4530823, EPI_ISL_4530824, EPI_ISL_4530825, EPI_ISL_4530826, EPI_ISL_4530827, EPI_ISL_4530828, EPI_ISL_4530829, EPI_ISL_4530830, EPI_ISL_4530831, EPI_ISL_4530833, EPI_ISL_4530834, EPI_ISL_4530835, EPI_ISL_4530836, EPI_ISL_4530837, EPI_ISL_4530838, EPI_ISL_4530839, EPI_ISL_4530840, EPI_ISL_4530841, EPI_ISL_4530842, EPI_ISL_4530843, EPI_ISL_4530844, EPI_ISL_4530845, EPI_ISL_4530847, EPI_ISL_4530848, EPI_ISL_4530849, EPI_ISL_4530850, EPI_ISL_4530851, EPI_ISL_4530852, EPI_ISL_4530853, EPI_ISL_4530854, EPI_ISL_4530855, EPI_ISL_4530856, EPI_ISL_4530857, EPI_ISL_4530858, EPI_ISL_4530859, EPI_ISL_4530860, EPI_ISL_4530861, EPI_ISL_4530862, EPI_ISL_4530863, EPI_ISL_4530864, EPI_ISL_4530865, EPI_ISL_4532010, EPI_ISL_4532011, EPI_ISL_4532012, EPI_ISL_4532013, EPI_ISL_4532014, EPI_ISL_4532015, EPI_ISL_4532016, EPI_ISL_4532017, EPI_ISL_4532018, EPI_ISL_4532019, EPI_ISL_4532020, EPI_ISL_4532021, EPI_ISL_4532022, EPI_ISL_4532023, EPI_ISL_4532024, EPI_ISL_4532025, EPI_ISL_4532026, EPI_ISL_4532027, EPI_ISL_4532028, EPI_ISL_4532029

| | | | |
|---|---|---|---|
| see above | Respiratory Virus Unit, Microbiology Services Colindale, Public Health England | COVID-19 Genomics UK (COG-UK) Consortium | PHE Covid Sequencing Team |
