## Supplementary material for "SARS-COV-2 δ variant drives the pandemic in India and Europe via two subvariants": Acknowledgement table on the GISAID genomes used in this study

We gratefully acknowledge the following Authors from the Originating laboratories responsible for obtaining the specimens, as well as the Submitting laboratories where the genome data were generated and shared via GISAID, on which this research is based.

All Submitters of data may be contacted directly via www.gisaid.org

Authors are sorted alphabetically.

| Accession ID | Originating Laboratory | Submitting Laboratory | Authors |
|---|---|---|---|
| EPI_ISL_4303856 | Althaia. Xarxa Assistencial Universitària de Manresa | IrsiCaixa | Antonia Flor; Bonaventura Clotet; Bonaventura Clotet Gloria Trujillo; Carolina Gonzalez-Fernandez; Eulalia Grau; Francesc Catala-Moll; Jaume Trapé; Marc Noguera-Julian; Maria Casadellà; Mariona Parera; Miquel Micó; Pilar Armengol; Rafel Perez Vidal; Roger Paredes |
| EPI_ISL_4414635, EPI_ISL_4414641, EPI_ISL_4414650, EPI_ISL_4414678, EPI_ISL_4414683, EPI_ISL_4543164, EPI_ISL_4543168 | | | |
| see above | CAP El Remei | Banc de Sang i Teixits | Carlos Hobeich; Francisco Vidal; Irene Corrales; Lorena Ramírez; Maria Glòria Soria; Natália Comes; Nina Borràs; Noemí Gonzalez; Silvia Sauleda |
| EPI_ISL_4414592, EPI_ISL_4543165, EPI_ISL_4543170 | CAP Manlleu | Banc de Sang i Teixits | Carlos Hobeich; Francisco Vidal; Irene Corrales; Lorena Ramírez; Maria Glòria Soria; Natália Comes; Nina Borràs; Noemí Gonzalez; Silvia Sauleda |
| EPI_ISL_4414626 | CAP Roda de Ter | Banc de Sang i Teixits | Carlos Hobeich; Francisco Vidal; Irene Corrales; Lorena Ramírez; Maria Glòria Soria; Natália Comes; Nina Borràs; Noemí Gonzalez; Silvia Sauleda |
| EPI_ISL_4543167, EPI_ISL_4543169, EPI_ISL_4543172 | CAP Taradell | Banc de Sang i Teixits | Carlos Hobeich; Francisco Vidal; Irene Corrales; Lorena Ramírez; Maria Glòria Soria; Natália Comes; Nina Borràs; Noemí Gonzalez; Silvia Sauleda |
| EPI_ISL_4414630, EPI_ISL_4543171, EPI_ISL_4543183 | CAP Tona | Banc de Sang i Teixits | Carlos Hobeich; Francisco Vidal; Irene Corrales; Lorena Ramírez; Maria Glòria Soria; Natália Comes; Nina Borràs; Noemí Gonzalez; Silvia Sauleda |
| EPI_ISL_4414667, EPI_ISL_4414668, EPI_ISL_4543175 | CAP Vic Nord | Banc de Sang i Teixits | Carlos Hobeich; Francisco Vidal; Irene Corrales; Lorena Ramírez; Maria Glòria Soria; Natália Comes; Nina Borràs; Noemí Gonzalez; Silvia Sauleda |
| EPI_ISL_4414598, EPI_ISL_4414599, EPI_ISL_4414607, EPI_ISL_4414610, EPI_ISL_4414611, EPI_ISL_4414613, EPI_ISL_4414622, EPI_ISL_4414623, EPI_ISL_4414624, EPI_ISL_4543166, EPI_ISL_4543173, EPI_ISL_4543174, EPI_ISL_4543176, EPI_ISL_4543177, EPI_ISL_4543178, EPI_ISL_4543179, EPI_ISL_4543180, EPI_ISL_4543181, EPI_ISL_4543182 | | | |
| see above | Fundacio Althaia-Manresa | Banc de Sang i Teixits | Carlos Hobeich; Francisco Vidal; Irene Corrales; Lorena Ramírez; Maria Glòria Soria; Natália Comes; Nina Borràs; Noemí Gonzalez; Silvia Sauleda |
| EPI_ISL_4350104, EPI_ISL_4350113, EPI_ISL_4350114, EPI_ISL_4350140, EPI_ISL_4350210, EPI_ISL_4350237, EPI_ISL_4350241, EPI_ISL_4350269, EPI_ISL_4350302, EPI_ISL_4550834, EPI_ISL_4550835, EPI_ISL_4550845, EPI_ISL_4550853, EPI_ISL_4550869, EPI_ISL_4550878, EPI_ISL_4550879, EPI_ISL_4550887, EPI_ISL_4550888, EPI_ISL_4550895, EPI_ISL_4550896 | | | |
| see above | HOSPITAL CAN MISSES | HOSPITAL UNIVERSITARIO SON ESPASES | Dr. Antonio Oliver; Dr. Carla López-Causapé; Dr. Gabriel Cabot; Hospital Universitario Son Espases; on behalf of Servicio de Microbiología |
| EPI_ISL_4350106, EPI_ISL_4350119, EPI_ISL_4350156, EPI_ISL_4350187, EPI_ISL_4350223, EPI_ISL_4350251, EPI_ISL_4550838, EPI_ISL_4550847, EPI_ISL_4550848, EPI_ISL_4550856, EPI_ISL_4550872, EPI_ISL_4550880 | | | |
| see above | HOSPITAL MANACOR | HOSPITAL UNIVERSITARIO SON ESPASES | Dr. Antonio Oliver; Dr. Carla López-Causapé; Dr. Gabriel Cabot; Hospital Universitario Son Espases; on behalf of Servicio de Microbiología |
| EPI_ISL_4550830, EPI_ISL_4550865, EPI_ISL_4550873 | HOSPITAL MATEU ORFILA | HOSPITAL UNIVERSITARIO SON ESPASES | Dr. Antonio Oliver; Dr. Carla López-Causapé; Dr. Gabriel Cabot; Hospital Universitario Son Espases; on behalf of Servicio de Microbiología |
| EPI_ISL_4547946 | HOSPITAL UNIVERSITARIO CENTRAL DE ASTURIAS | Laboratorio de Virología HUCA | ; Alba L; Alvarez-Arguelles ME; Boga JA; Costales I; Coto E; González-Alba JM; Gómez de Oña J; Martín-Rodríguez G; Melón S; Perez-Martínez Z; Rojo S; Sandoval M |
| EPI_ISL_4350099, EPI_ISL_4350100, EPI_ISL_4350101, EPI_ISL_4350102, EPI_ISL_4350103, EPI_ISL_4350105, EPI_ISL_4350107, EPI_ISL_4350108, EPI_ISL_4350109, EPI_ISL_4350110, EPI_ISL_4350111, EPI_ISL_4350112, EPI_ISL_4350116, EPI_ISL_4350123, EPI_ISL_4350126, EPI_ISL_4350130, EPI_ISL_4350137, EPI_ISL_4350147, EPI_ISL_4350150, EPI_ISL_4350153, EPI_ISL_4350160, EPI_ISL_4350163, EPI_ISL_4350167, EPI_ISL_4350170, EPI_ISL_4350173, EPI_ISL_4350177, EPI_ISL_4350184, EPI_ISL_4350190, EPI_ISL_4350193, EPI_ISL_4350197, EPI_ISL_4350200, EPI_ISL_4350203, EPI_ISL_4350220, EPI_ISL_4350226, EPI_ISL_4350229, EPI_ISL_4350233, EPI_ISL_4350244, EPI_ISL_4350247, EPI_ISL_4350254, EPI_ISL_4350258, EPI_ISL_4350262, EPI_ISL_4350265, EPI_ISL_4350273, EPI_ISL_4350280, EPI_ISL_4350283, EPI_ISL_4350288, EPI_ISL_4350294, EPI_ISL_4350298, EPI_ISL_4350305, EPI_ISL_4350309, EPI_ISL_4350313, EPI_ISL_4550828, EPI_ISL_4550831, EPI_ISL_4550832, EPI_ISL_4550833, EPI_ISL_4550836, EPI_ISL_4550837, EPI_ISL_4550839, EPI_ISL_4550840, EPI_ISL_4550841, EPI_ISL_4550843, EPI_ISL_4550844, EPI_ISL_4550846, EPI_ISL_4550849, EPI_ISL_4550850, EPI_ISL_4550851, EPI_ISL_4550852, EPI_ISL_4550854, EPI_ISL_4550855, EPI_ISL_4550859, EPI_ISL_4550860, EPI_ISL_4550861, EPI_ISL_4550862, EPI_ISL_4550863, EPI_ISL_4550864, EPI_ISL_4550866, EPI_ISL_4550867, EPI_ISL_4550868, EPI_ISL_4550871, EPI_ISL_4550874, EPI_ISL_4550875, EPI_ISL_4550876, EPI_ISL_4550877, EPI_ISL_4550881, EPI_ISL_4550882, EPI_ISL_4550884, EPI_ISL_4550885, EPI_ISL_4550886, EPI_ISL_4550889, EPI_ISL_4550890, EPI_ISL_4550891, EPI_ISL_4550892, EPI_ISL_4550893, EPI_ISL_4550894, EPI_ISL_4550897 | | | |
| see above | HOSPITAL UNIVERSITARIO SON ESPASES | HOSPITAL UNIVERSITARIO SON ESPASES | Dr. Antonio Oliver; Dr. Carla López-Causapé; Dr. Gabriel Cabot; Hospital Universitario Son Espases; on behalf of Servicio de Microbiología |
| EPI_ISL_4350277, EPI_ISL_4550857 | HOSPITAL UNIVERSITARIO SON LLATZER | HOSPITAL UNIVERSITARIO SON ESPASES | Dr. Antonio Oliver; Dr. Carla López-Causapé; Dr. Gabriel Cabot; Hospital Universitario Son Espases; on behalf of Servicio de Microbiología |
| EPI_ISL_4352055, EPI_ISL_4395972, EPI_ISL_4395973, EPI_ISL_4395974, EPI_ISL_4395975, EPI_ISL_4395976, EPI_ISL_4395977, EPI_ISL_4395978, EPI_ISL_4395979, EPI_ISL_4395980, EPI_ISL_4395981, EPI_ISL_4395982, EPI_ISL_4395983, EPI_ISL_4395984, EPI_ISL_4395985, EPI_ISL_4395986, EPI_ISL_4395987, EPI_ISL_4395988, EPI_ISL_4395989, EPI_ISL_4395990, EPI_ISL_4395991, EPI_ISL_4395992, EPI_ISL_4395993, EPI_ISL_4395994, EPI_ISL_4395995, EPI_ISL_4395996, EPI_ISL_4395997, EPI_ISL_4395998, EPI_ISL_4395999, EPI_ISL_4396000, EPI_ISL_4396001, EPI_ISL_4396002, EPI_ISL_4396003, EPI_ISL_4396004, EPI_ISL_4396005, EPI_ISL_4396006, EPI_ISL_4396007, EPI_ISL_4396008, EPI_ISL_4396009, EPI_ISL_4396010, EPI_ISL_4396011, EPI_ISL_4396012, EPI_ISL_4396013, EPI_ISL_4396014, EPI_ISL_4396015, EPI_ISL_4396016, EPI_ISL_4396017, EPI_ISL_4396018, EPI_ISL_4396019, EPI_ISL_4396020, EPI_ISL_4396021, EPI_ISL_4396022, EPI_ISL_4396023, EPI_ISL_4396024, EPI_ISL_4396025, EPI_ISL_4396026, EPI_ISL_4396027, EPI_ISL_4396028, EPI_ISL_4396029, EPI_ISL_4396030, EPI_ISL_4396031, EPI_ISL_4396032, EPI_ISL_4396033, EPI_ISL_4396034, EPI_ISL_4396035, EPI_ISL_4396036, EPI_ISL_4547811, EPI_ISL_4547812, EPI_ISL_4547813, EPI_ISL_4547814, EPI_ISL_4547815, EPI_ISL_4547816, EPI_ISL_4547817, EPI_ISL_4547818, EPI_ISL_4547819, EPI_ISL_4547820, EPI_ISL_4547821, EPI_ISL_4547822, EPI_ISL_4547823, EPI_ISL_4547824, EPI_ISL_4547825, EPI_ISL_4547826, EPI_ISL_4547827, EPI_ISL_4547828, EPI_ISL_4547829, EPI_ISL_4547830, EPI_ISL_4547831, EPI_ISL_4547832, EPI_ISL_4547833, EPI_ISL_4547834, EPI_ISL_4547835, EPI_ISL_4547836, EPI_ISL_4547837, EPI_ISL_4547838, EPI_ISL_4547839, EPI_ISL_4547840, EPI_ISL_4547841, EPI_ISL_4547842, EPI_ISL_4547843, EPI_ISL_4547844, EPI_ISL_4547845, EPI_ISL_4547846, EPI_ISL_4547847, EPI_ISL_4547848, EPI_ISL_4547849, EPI_ISL_4547850, EPI_ISL_4547851, EPI_ISL_4547852, EPI_ISL_4547853, EPI_ISL_4547854, EPI_ISL_4547855, EPI_ISL_4547856, EPI_ISL_4547857, EPI_ISL_4547858, EPI_ISL_4547859, EPI_ISL_4547860 | | | |
| see above | Hospital General Universitario Gregorio Marañón | Hospital General Universitario Gregorio Marañón | Cristina Rodriguez-Grande; Darío García de Viedma; Julia Suárez; Laura Pérez-Lago; Marta Herranz Martin; Patricia Muñoz; Pedro Sola Campoy; Pilar Catalán; Sergio Buenestado Serrano; Victor Manuel de la Cueva |
| EPI_ISL_4237860, EPI_ISL_4237867, EPI_ISL_4253776, EPI_ISL_4253777, EPI_ISL_4253778, EPI_ISL_4253779, EPI_ISL_4253780, EPI_ISL_4253781, EPI_ISL_4253782, EPI_ISL_4253784, EPI_ISL_4253785, EPI_ISL_4253786, EPI_ISL_4253787, EPI_ISL_4253788, EPI_ISL_4253789, EPI_ISL_4473254, EPI_ISL_4473255, EPI_ISL_4473256, EPI_ISL_4473257, EPI_ISL_4473259, EPI_ISL_4473262, EPI_ISL_4473263, EPI_ISL_4473264, EPI_ISL_4473266, EPI_ISL_4473267, EPI_ISL_4473268, EPI_ISL_4578036, EPI_ISL_4578038, EPI_ISL_4578041, EPI_ISL_4578042, EPI_ISL_4578044, EPI_ISL_4578046, EPI_ISL_4578049, EPI_ISL_4578051 | | | |
| see above | Hospital General Universitario de Ciudad Real | Hospital General Universitario de Ciudad Real | Cristina Colmenarejo; José Martínez-Alarcón; Lidia García-Agudo; Marta Torres-Narbona; Soledad Illescas Fernández-Bermejo |
| EPI_ISL_4473258, EPI_ISL_4473260, EPI_ISL_4473261, EPI_ISL_4578039, EPI_ISL_4578040, EPI_ISL_4578045 | Hospital General de Valdepeñas | Hospital General Universitario de Ciudad Real | Cristina Colmenarejo; José Martínez-Alarcón; Lidia García-Agudo; Marta Torres-Narbona; Soledad Illescas Fernández-Bermejo |
| EPI_ISL_4578037, EPI_ISL_4578047 | Hospital Santa Bárbara | Hospital General Universitario de Ciudad Real | Cristina Colmenarejo; José Martínez-Alarcón; Lidia García-Agudo; Marta Torres-Narbona; Soledad Illescas Fernández-Bermejo |
| EPI_ISL_4112835, EPI_ISL_4112838, EPI_ISL_4112841, EPI_ISL_4112844, EPI_ISL_4112847, EPI_ISL_4112850, EPI_ISL_4112855, EPI_ISL_4112860, EPI_ISL_4112863, EPI_ISL_4112865, EPI_ISL_4112871, EPI_ISL_4112979, EPI_ISL_4112993, EPI_ISL_4112996, EPI_ISL_4113002, EPI_ISL_4113004, EPI_ISL_4113008, EPI_ISL_4113014, EPI_ISL_4113028, EPI_ISL_4348639, EPI_ISL_4348640, EPI_ISL_4348641, EPI_ISL_4348642, EPI_ISL_4348643, EPI_ISL_4348644, EPI_ISL_4348645, EPI_ISL_4348646, EPI_ISL_4348647, EPI_ISL_4348648, EPI_ISL_4348649, EPI_ISL_4348650, EPI_ISL_4348651, EPI_ISL_4348652, EPI_ISL_4348654, EPI_ISL_4348655, EPI_ISL_4348656, EPI_ISL_4348657, EPI_ISL_4548970, EPI_ISL_4548971, EPI_ISL_4548972, EPI_ISL_4548973, EPI_ISL_4548974, EPI_ISL_4548975, EPI_ISL_4548976 | | | |
| see above | Hospital Universitari Arnau de Vilanova | Hospital Universitari Vall d'Hebron - Vall d'Hebron Institut de Recerca | Alejandra González-Sánchez; Andrés Antón; Ariadna Rando; Carla Castillo; Cristina Andrés; Damir Garcia-Cehic; Josep Quer; Juliana Esperalba; Karen García; Maria Carmen Martin; Maria Gema Codina; Maria Piñana; Rodrigo Vásquez; Tomàs Pumarola |
| EPI_ISL_4036449, EPI_ISL_4036450, EPI_ISL_4036451, EPI_ISL_4036452, EPI_ISL_4036453, EPI_ISL_4036454, EPI_ISL_4036455, EPI_ISL_4080640, EPI_ISL_4080641, EPI_ISL_4080642, EPI_ISL_4080643, EPI_ISL_4080644, EPI_ISL_4080645, EPI_ISL_4080646, EPI_ISL_4080647, EPI_ISL_4170721, EPI_ISL_4170722, EPI_ISL_4170723, EPI_ISL_4170724, EPI_ISL_4170725, EPI_ISL_4170726, EPI_ISL_4170727, EPI_ISL_4170728, EPI_ISL_4170729, EPI_ISL_4170730, EPI_ISL_4170731, EPI_ISL_4444201, EPI_ISL_4444203, EPI_ISL_4444206, EPI_ISL_4444208, EPI_ISL_4444210, EPI_ISL_4444213, EPI_ISL_4444215, EPI_ISL_4444217, EPI_ISL_4444219, EPI_ISL_4444222, EPI_ISL_4444224, EPI_ISL_4444226, EPI_ISL_4444228, EPI_ISL_4444230, EPI_ISL_4444232, EPI_ISL_4444234, EPI_ISL_4444236, EPI_ISL_4444238, EPI_ISL_4444240, EPI_ISL_4600396, EPI_ISL_4600397, EPI_ISL_4600398, EPI_ISL_4600399, EPI_ISL_4600400 | | | |
| see above | Hospital Universitari Joan XXIII | Hospital Universitari Joan XXIII | Carla Martín; Clara Benavent; Cristina Gutiérrez; Ester Picó; Gemma Recio; Margarida Terrón |
| EPI_ISL_4112930, EPI_ISL_4348580, EPI_ISL_4348581, EPI_ISL_4348582, EPI_ISL_4348583, EPI_ISL_4348584, EPI_ISL_4348588, EPI_ISL_4348590, EPI_ISL_4348591, EPI_ISL_4348592, EPI_ISL_4348593, EPI_ISL_4348598, EPI_ISL_4348599, EPI_ISL_4348600, EPI_ISL_4348606, EPI_ISL_4348607, EPI_ISL_4348616, EPI_ISL_4348619, EPI_ISL_4348620, EPI_ISL_4348621, EPI_ISL_4348622, EPI_ISL_4348625, EPI_ISL_4348626, EPI_ISL_4348628, EPI_ISL_4348630, EPI_ISL_4348631, EPI_ISL_4348633, EPI_ISL_4348636, EPI_ISL_4348638, EPI_ISL_4548901, EPI_ISL_4548902, EPI_ISL_4548903, EPI_ISL_4548904, EPI_ISL_4548905, EPI_ISL_4548906, EPI_ISL_4548907, EPI_ISL_4548908, EPI_ISL_4548909, EPI_ISL_4548910, EPI_ISL_4548911, EPI_ISL_4548912, EPI_ISL_4548913, EPI_ISL_4548917, EPI_ISL_4548918, EPI_ISL_4548920, EPI_ISL_4548922, EPI_ISL_4548927, EPI_ISL_4548928, EPI_ISL_4548929, EPI_ISL_4548933, EPI_ISL_4548936, EPI_ISL_4548939, EPI_ISL_4548940, EPI_ISL_4548941, EPI_ISL_4548942, EPI_ISL_4548945, EPI_ISL_4548947, EPI_ISL_4548953, EPI_ISL_4548954, EPI_ISL_4548955, EPI_ISL_4548956, EPI_ISL_4548957, EPI_ISL_4548958, EPI_ISL_4548959, EPI_ISL_4548962, EPI_ISL_4548963, EPI_ISL_4548964, EPI_ISL_4548965, EPI_ISL_4548966, EPI_ISL_4548967 | | | |
| see above | Hospital Universitari Vall d'Hebron - Vall d'Hebron Institut de Recerca | Hospital Universitari Vall d'Hebron - Vall d'Hebron Institut de Recerca | Alejandra González-Sánchez; Andrés Antón; Ariadna Rando; Carla Castillo; Cristina Andrés; Damir Garcia-Cehic; Josep Quer; Juliana Esperalba; Karen García; Maria Carmen Martin; Maria Gema Codina; Maria Piñana; Rodrigo Vásquez; Tomàs Pumarola |
| EPI_ISL_4253783, EPI_ISL_4473265, EPI_ISL_4473269, EPI_ISL_4578048 | Hospital Virgen De Altagracia | Hospital General Universitario de Ciudad Real | Cristina Colmenarejo; José Martínez-Alarcón; Lidia García-Agudo; Marta Torres-Narbona; Soledad Illescas Fernández-Bermejo |
| EPI_ISL_4533903 | Microbiologia CATLAB | Can Ruti SARS-CoV-2 Sequencing Hub (HUGTiP/IrsiCaixa/IGTP) | Alexia París; Anna Not; Antoni E Bordoy; Bonaventura Clotet; Cristina Casañ; David Panisello; Francesc Catala-Moll; Gemma Clara; Ignacio Blanco; Laia Soler; Lauro Sumoy; Marc Noguera-Julian; Maria Casadellà; Mariona Parera; Mercedes Guerrero; Montserrat Giménez; Pere-Joan Cardona; Pilar Armengol; Roger Paredes; Verónica Saludes; and Elisa Martró on behalf of the Can Ruti SARS-CoV-2 Sequencing Hub |
| EPI_ISL_4171988, EPI_ISL_4171989, EPI_ISL_4171990, EPI_ISL_4172001, EPI_ISL_4172002, EPI_ISL_4172012, EPI_ISL_4172013, EPI_ISL_4172014, EPI_ISL_4172023, EPI_ISL_4172024, EPI_ISL_4172025, EPI_ISL_4172035, EPI_ISL_4172036, EPI_ISL_4172037, EPI_ISL_4172050, EPI_ISL_4172054, EPI_ISL_4172067, EPI_ISL_4172069, EPI_ISL_4172071, EPI_ISL_4172084, EPI_ISL_4172086, EPI_ISL_4172088, EPI_ISL_4303761, EPI_ISL_4303763, EPI_ISL_4303764, EPI_ISL_4303765, EPI_ISL_4303766, EPI_ISL_4303767, EPI_ISL_4303771, EPI_ISL_4303775, EPI_ISL_4303776, EPI_ISL_4303777, EPI_ISL_4303778, EPI_ISL_4303779, EPI_ISL_4303783, EPI_ISL_4303787, EPI_ISL_4303788, EPI_ISL_4303789, EPI_ISL_4303790, EPI_ISL_4303791, EPI_ISL_4303795, EPI_ISL_4303799, EPI_ISL_4303800, EPI_ISL_4303801, EPI_ISL_4303802, EPI_ISL_4303803, EPI_ISL_4303807, EPI_ISL_4303811, EPI_ISL_4303812, EPI_ISL_4303813, EPI_ISL_4303815, EPI_ISL_4303819, EPI_ISL_4303820, EPI_ISL_4303822, EPI_ISL_4303823, EPI_ISL_4303824, EPI_ISL_4303826, EPI_ISL_4303830, EPI_ISL_4303831, EPI_ISL_4303833, EPI_ISL_4303834, EPI_ISL_4303835, EPI_ISL_4303836, EPI_ISL_4303841, EPI_ISL_4303845, EPI_ISL_4303846, EPI_ISL_4303847, EPI_ISL_4303848, EPI_ISL_4303849, EPI_ISL_4359735, EPI_ISL_4359739, EPI_ISL_4359740, EPI_ISL_4359741, EPI_ISL_4359742, EPI_ISL_4359743, EPI_ISL_4359744, EPI_ISL_4359745, EPI_ISL_4359746, EPI_ISL_4359747, EPI_ISL_4359759, EPI_ISL_4359765, EPI_ISL_4359768, EPI_ISL_4359770, EPI_ISL_4359773, EPI_ISL_4359775, EPI_ISL_4359777, EPI_ISL_4359779, EPI_ISL_4359789, EPI_ISL_4359792, EPI_ISL_4359795, EPI_ISL_4359797, EPI_ISL_4359799, EPI_ISL_4359802, EPI_ISL_4359805, EPI_ISL_4359808, EPI_ISL_4359811, EPI_ISL_4359825, EPI_ISL_4359828, EPI_ISL_4359836, EPI_ISL_4359841, EPI_ISL_4359845, EPI_ISL_4359850, EPI_ISL_4359854, EPI_ISL_4359872, EPI_ISL_4359876, EPI_ISL_4359880, EPI_ISL_4359884, EPI_ISL_4359887, EPI_ISL_4359891, EPI_ISL_4359896, EPI_ISL_4359904, EPI_ISL_4359913, EPI_ISL_4359915, EPI_ISL_4359918, EPI_ISL_4359920, EPI_ISL_4359924, EPI_ISL_4359926, EPI_ISL_4359929, EPI_ISL_4359935, EPI_ISL_4359942, EPI_ISL_4359943, EPI_ISL_4359946, EPI_ISL_4359950, EPI_ISL_4359953, EPI_ISL_4359955, EPI_ISL_4359963, EPI_ISL_4359971, EPI_ISL_4359973, EPI_ISL_4359976, EPI_ISL_4359978, EPI_ISL_4359980, EPI_ISL_4359982, EPI_ISL_4359985, EPI_ISL_4533561, EPI_ISL_4533565, EPI_ISL_4533567, EPI_ISL_4533568, EPI_ISL_4533570, EPI_ISL_4533573, EPI_ISL_4533575, EPI_ISL_4533577, EPI_ISL_4533578, EPI_ISL_4533579, EPI_ISL_4533580, EPI_ISL_4533581, EPI_ISL_4533584, EPI_ISL_4533586, EPI_ISL_4533588, EPI_ISL_4533589, EPI_ISL_4533590, EPI_ISL_4533591, EPI_ISL_4533592, EPI_ISL_4533595, EPI_ISL_4533597, EPI_ISL_4533599, EPI_ISL_4533600, EPI_ISL_4533601, EPI_ISL_4533602, EPI_ISL_4533603, EPI_ISL_4533604, EPI_ISL_4533607, EPI_ISL_4533610, EPI_ISL_4533611, EPI_ISL_4533612, EPI_ISL_4533613, EPI_ISL_4533614, EPI_ISL_4533617, EPI_ISL_4533621, EPI_ISL_4533622, EPI_ISL_4533623, EPI_ISL_4533624, EPI_ISL_4533628, EPI_ISL_4533631, EPI_ISL_4533632, EPI_ISL_4533633, EPI_ISL_4533634, EPI_ISL_4533635, EPI_ISL_4533639, EPI_ISL_4533640, EPI_ISL_4533643, EPI_ISL_4533644, EPI_ISL_4533646, EPI_ISL_4533647, EPI_ISL_4533772, EPI_ISL_4533776, EPI_ISL_4533777, EPI_ISL_4533779, EPI_ISL_4533780, EPI_ISL_4533781, EPI_ISL_4533783, EPI_ISL_4533784, EPI_ISL_4533786, EPI_ISL_4533788, EPI_ISL_4533789, EPI_ISL_4533790, EPI_ISL_4533791, EPI_ISL_4533792, EPI_ISL_4533793, EPI_ISL_4533794, EPI_ISL_4533795, EPI_ISL_4533801, EPI_ISL_4533802, EPI_ISL_4533803, EPI_ISL_4533804, EPI_ISL_4533805, EPI_ISL_4533806, EPI_ISL_4533807, EPI_ISL_4533808, EPI_ISL_4533812, EPI_ISL_4533813, EPI_ISL_4533814, EPI_ISL_4533815, EPI_ISL_4533816, EPI_ISL_4533817, EPI_ISL_4533818, EPI_ISL_4533819, EPI_ISL_4533820, EPI_ISL_4533825, EPI_ISL_4533826, EPI_ISL_4533827, EPI_ISL_4533828, EPI_ISL_4533829, EPI_ISL_4533830, EPI_ISL_4533831, EPI_ISL_4533836, EPI_ISL_4533837, EPI_ISL_4533838, EPI_ISL_4533839, EPI_ISL_4533840, EPI_ISL_4533841, EPI_ISL_4533842, EPI_ISL_4533844, EPI_ISL_4533847, EPI_ISL_4533848, EPI_ISL_4533849, EPI_ISL_4533850, EPI_ISL_4533851, EPI_ISL_4533852, EPI_ISL_4533855, EPI_ISL_4533858, EPI_ISL_4533859, EPI_ISL_4533860, EPI_ISL_4533861, EPI_ISL_4533862, EPI_ISL_4533863, EPI_ISL_4533864, EPI_ISL_4533866, EPI_ISL_4533876, EPI_ISL_4533881, EPI_ISL_4533882, EPI_ISL_4533885, EPI_ISL_4533890, EPI_ISL_4533895, EPI_ISL_4533900, EPI_ISL_4533905 | | | |
| see above | Microbiology Department, Laboratori Clínic Metropolitana Nord. Hospital Universitari Germans Trias i Pujol | Can Ruti SARS-CoV-2 Sequencing Hub (HUGTiP/IrsiCaixa/IGTP) | Alexia París; Anna Not; Antoni E Bordoy; Bonaventura Clotet; Cristina Casañ; David Panisello; Francesc Catala-Moll; Gemma Clara; Ignacio Blanco; Laia Soler; Lauro Sumoy; Marc Noguera-Julian; Maria Casadellà; Mariona Parera; Mercedes Guerrero; Montserrat Giménez; Pere-Joan Cardona; Pilar Armengol; Roger Paredes; Verónica Saludes; and Elisa Martró on behalf of the Can Ruti SARS-CoV-2 Sequencing Hub |
| EPI_ISL_4458501, EPI_ISL_4506756, EPI_ISL_4506757, EPI_ISL_4562723, EPI_ISL_4562726, EPI_ISL_4562728, EPI_ISL_4562731, EPI_ISL_4562734, EPI_ISL_4562737, EPI_ISL_4562742, EPI_ISL_4562746, EPI_ISL_4562749, EPI_ISL_4562751, EPI_ISL_4562757, EPI_ISL_4562760, EPI_ISL_4562763, EPI_ISL_4562766, EPI_ISL_4562769, EPI_ISL_4562810, EPI_ISL_4562821, EPI_ISL_4562832 | | | |
| see above | Microbiology Department. Complexo Hospitalario Universitario de Vigo | Microbiology Department. Complexo Hospitalario Universitario de Vigo | Alvarez M; Cabrera JJ; Carballo R; Cores O; Cortizo S; Davina C; Martinez L; Mediero G; Pena I; Perez S; Potel C; Regueiro B; Rey S; Vasallo FJ; del-Campo V |
| EPI_ISL_4506277, EPI_ISL_4506279, EPI_ISL_4506284, EPI_ISL_4506285, EPI_ISL_4506286, EPI_ISL_4506324, EPI_ISL_4506325, EPI_ISL_4506326, EPI_ISL_4506327, EPI_ISL_4506328, EPI_ISL_4506329, EPI_ISL_4506330, EPI_ISL_4506331, EPI_ISL_4506332, EPI_ISL_4506333, EPI_ISL_4506334, EPI_ISL_4506335, EPI_ISL_4506336, EPI_ISL_4506337, EPI_ISL_4506338, EPI_ISL_4506339, EPI_ISL_4506340, EPI_ISL_4506341, EPI_ISL_4506342, EPI_ISL_4506343, EPI_ISL_4506344, EPI_ISL_4506345, EPI_ISL_4506346, EPI_ISL_4506347, EPI_ISL_4506348, EPI_ISL_4506349, EPI_ISL_4506350, EPI_ISL_4506351, EPI_ISL_4506352, EPI_ISL_4506353, EPI_ISL_4506354, EPI_ISL_4506355, EPI_ISL_4506356, EPI_ISL_4506357, EPI_ISL_4506358, EPI_ISL_4506359, EPI_ISL_4506360, EPI_ISL_4506361, EPI_ISL_4506362, EPI_ISL_4506363, EPI_ISL_4506364, EPI_ISL_4506365, EPI_ISL_4506366, EPI_ISL_4506367, EPI_ISL_4506368, EPI_ISL_4506369, EPI_ISL_4506370, EPI_ISL_4506371, EPI_ISL_4506372, EPI_ISL_4506373, EPI_ISL_4506374, EPI_ISL_4506375, EPI_ISL_4506430, EPI_ISL_4506431, EPI_ISL_4506437, EPI_ISL_4506439, EPI_ISL_4506440, EPI_ISL_4506445, EPI_ISL_4506446, EPI_ISL_4506447, EPI_ISL_4506448, EPI_ISL_4506449, EPI_ISL_4506450, EPI_ISL_4506451, EPI_ISL_4506452, EPI_ISL_4506453, EPI_ISL_4506454, EPI_ISL_4506455, EPI_ISL_4506456, EPI_ISL_4506484, EPI_ISL_4506485, EPI_ISL_4506486, EPI_ISL_4506513, EPI_ISL_4506514, EPI_ISL_4506517, EPI_ISL_4506518, EPI_ISL_4506519, EPI_ISL_4506520, EPI_ISL_4506523, EPI_ISL_4506524, EPI_ISL_4506525, EPI_ISL_4506526, EPI_ISL_4506529, EPI_ISL_4506533, EPI_ISL_4506534, EPI_ISL_4506535, EPI_ISL_4506537, EPI_ISL_4506538, EPI_ISL_4506539, EPI_ISL_4506540, EPI_ISL_4506541, EPI_ISL_4506542, EPI_ISL_4506591, EPI_ISL_4506592, EPI_ISL_4506593, EPI_ISL_4506594, EPI_ISL_4506595, EPI_ISL_4506596, EPI_ISL_4506597, EPI_ISL_4506598, EPI_ISL_4506599, EPI_ISL_4506600, EPI_ISL_4506601, EPI_ISL_4506602, EPI_ISL_4506603, EPI_ISL_4506604, EPI_ISL_4506605, EPI_ISL_4506606, EPI_ISL_4506607, EPI_ISL_4506608, EPI_ISL_4506609, EPI_ISL_4506610, EPI_ISL_4506611, EPI_ISL_4506612, EPI_ISL_4506613, EPI_ISL_4506614, EPI_ISL_4506615, EPI_ISL_4506617, EPI_ISL_4506618, EPI_ISL_4506619, EPI_ISL_4506620, EPI_ISL_4506621, EPI_ISL_4506622, EPI_ISL_4506623, EPI_ISL_4506624, EPI_ISL_4506625, EPI_ISL_4506626, EPI_ISL_4506627, EPI_ISL_4506628, EPI_ISL_4506629, EPI_ISL_4506630, EPI_ISL_4506631, EPI_ISL_4506632, EPI_ISL_4506633, EPI_ISL_4506634, EPI_ISL_4506635, EPI_ISL_4506636 | | | |

| | | | |
|---|---|---|---|
| see above | SARS-CoV-2 Sequencing Castilla y Leon-Spain Consortium | SARS-CoV-2 Sequencing Castilla y Leon-Spain Consortium | Antonio Orduña-Domingo; Carlos Fuster Foz; Carmen Aldea-Mansilla; Carmen Gimeno Crespo; David Abad; Gabriel March Rosello; Gregoria Megías Lobón; Jose María Eiros Bouza; M. Isabel Fernandez-Natal; Marta Dominguez-Gil; Marta Hernandez; María Antonia García Castro; Mª Fe Brezmes-Valdivieso; Noelia Arenal Andrés; Silvia Rojo; Sonsoles Garcinuño Pérez |
| EPI_ISL_4391362, EPI_ISL_4391368, EPI_ISL_4391374, EPI_ISL_4391380, EPI_ISL_4391382, EPI_ISL_4391390, EPI_ISL_4391398, EPI_ISL_4391399, EPI_ISL_4391404, EPI_ISL_4391406, EPI_ISL_4391411, EPI_ISL_4391421, EPI_ISL_4391425, EPI_ISL_4391427, EPI_ISL_4391432, EPI_ISL_4391433 | | | |
| see above | Servicio de Microbiologia Hospital Ramon y Cajal | Servicio de Microbiologia Hospital Ramon y Cajal | Galan JC; Martinez L. Abreu M; Ponce M; y Gonzalez-Alba JM |
