## Supplementary material for "SARS-COV-2 δ variant drives the pandemic in India and Europe via two subvariants": Acknowledgement table on the GISAID genomes used in this study

We gratefully acknowledge the following Authors from the Originating laboratories responsible for obtaining the specimens, as well as the Submitting laboratories where the genome data were generated and shared via GISAID, on which this research is based.

All Submitters of data may be contacted directly via www.gisaid.org

Authors are sorted alphabetically.

| Accession ID | Originating Laboratory | Submitting Laboratory | Authors |
|---|---|---|---|
| EPI_ISL_4572088, EPI_ISL_4572089, EPI_ISL_4572220 | A.S.L. ALESSANDRIA | Fondazione del Piemonte per l'Oncologia IRCCS | Antonino Sottile; Giorgio Giardina; Paola Marino; Silvia Brossa |
| EPI_ISL_4256376, EPI_ISL_4256377, EPI_ISL_4256378, EPI_ISL_4256379, EPI_ISL_4256380, EPI_ISL_4256381, EPI_ISL_4256382, EPI_ISL_4256383, EPI_ISL_4256386, EPI_ISL_4256387, EPI_ISL_4436498, EPI_ISL_4436502, EPI_ISL_4436503, EPI_ISL_4436505, EPI_ISL_4572051, EPI_ISL_4572052, EPI_ISL_4572053, EPI_ISL_4572054, EPI_ISL_4572055, EPI_ISL_4572056, EPI_ISL_4572122, EPI_ISL_4572125, EPI_ISL_4572126 | | | |
| see above | A.S.L. ASTI | Fondazione del Piemonte per l'Oncologia IRCCS | Antonino Sottile; Giorgio Giardina; Paola Marino; Silvia Brossa |
| EPI_ISL_4436487, EPI_ISL_4436488, EPI_ISL_4436489, EPI_ISL_4436491, EPI_ISL_4436492, EPI_ISL_4436493, EPI_ISL_4436494, EPI_ISL_4436495, EPI_ISL_4436496, EPI_ISL_4436497, EPI_ISL_4436499, EPI_ISL_4436500, EPI_ISL_4436501, EPI_ISL_4436546, EPI_ISL_4436549, EPI_ISL_4436550, EPI_ISL_4436551, EPI_ISL_4436552, EPI_ISL_4436554 | | | |
| see above | A.S.L. CITTA DI TORINO - OSPEDALE MAURIZIANO | Fondazione del Piemonte per l'Oncologia IRCCS | Antonino Sottile; Giorgio Giardina; Paola Marino; Silvia Brossa |
| EPI_ISL_4256384, EPI_ISL_4256385, EPI_ISL_4256388, EPI_ISL_4256390, EPI_ISL_4256391, EPI_ISL_4256392, EPI_ISL_4256393, EPI_ISL_4256394, EPI_ISL_4256395, EPI_ISL_4256396, EPI_ISL_4256397, EPI_ISL_4256398, EPI_ISL_4256399, EPI_ISL_4256400, EPI_ISL_4256401, EPI_ISL_4256402, EPI_ISL_4256403, EPI_ISL_4256404, EPI_ISL_4256405, EPI_ISL_4256406, EPI_ISL_4256407, EPI_ISL_4256408, EPI_ISL_4256409, EPI_ISL_4256410, EPI_ISL_4256411, EPI_ISL_4256412, EPI_ISL_4256413, EPI_ISL_4256414, EPI_ISL_4256415, EPI_ISL_4256416, EPI_ISL_4256417, EPI_ISL_4256418, EPI_ISL_4256419, EPI_ISL_4256420, EPI_ISL_4256421, EPI_ISL_4256422, EPI_ISL_4256423, EPI_ISL_4256424, EPI_ISL_4256425, EPI_ISL_4256426, EPI_ISL_4256427, EPI_ISL_4256428, EPI_ISL_4436504, EPI_ISL_4436506, EPI_ISL_4436507, EPI_ISL_4436508, EPI_ISL_4436509, EPI_ISL_4436510, EPI_ISL_4436511, EPI_ISL_4436512, EPI_ISL_4436513, EPI_ISL_4436514, EPI_ISL_4436515, EPI_ISL_4436516, EPI_ISL_4436517, EPI_ISL_4436518, EPI_ISL_4436519, EPI_ISL_4436520, EPI_ISL_4436521, EPI_ISL_4436524, EPI_ISL_4436526, EPI_ISL_4436527, EPI_ISL_4436528, EPI_ISL_4436529, EPI_ISL_4436530, EPI_ISL_4436531, EPI_ISL_4436532, EPI_ISL_4436533, EPI_ISL_4436534, EPI_ISL_4436535, EPI_ISL_4436536, EPI_ISL_4436537, EPI_ISL_4436538, EPI_ISL_4436539, EPI_ISL_4436540, EPI_ISL_4436541, EPI_ISL_4436542, EPI_ISL_4436543, EPI_ISL_4436544, EPI_ISL_4436545, EPI_ISL_4436547, EPI_ISL_4572066, EPI_ISL_4572067, EPI_ISL_4572069, EPI_ISL_4572070, EPI_ISL_4572071, EPI_ISL_4572072, EPI_ISL_4572074, EPI_ISL_4572075, EPI_ISL_4572076, EPI_ISL_4572077, EPI_ISL_4572078, EPI_ISL_4572079, EPI_ISL_4572080, EPI_ISL_4572081, EPI_ISL_4572082, EPI_ISL_4572083, EPI_ISL_4572084, EPI_ISL_4572085, EPI_ISL_4572086, EPI_ISL_4572087, EPI_ISL_4572090, EPI_ISL_4572091, EPI_ISL_4572092, EPI_ISL_4572093, EPI_ISL_4572094, EPI_ISL_4572095, EPI_ISL_4572096, EPI_ISL_4572097, EPI_ISL_4572098, EPI_ISL_4572099, EPI_ISL_4572100, EPI_ISL_4572101, EPI_ISL_4572102, EPI_ISL_4572104, EPI_ISL_4572116, EPI_ISL_4572117, EPI_ISL_4572118, EPI_ISL_4572119, EPI_ISL_4572121, EPI_ISL_4572123, EPI_ISL_4572127, EPI_ISL_4572128, EPI_ISL_4572129, EPI_ISL_4572130, EPI_ISL_4572131, EPI_ISL_4572132, EPI_ISL_4572133, EPI_ISL_4572134, EPI_ISL_4572135, EPI_ISL_4572136, EPI_ISL_4572137, EPI_ISL_4572138, EPI_ISL_4572139, EPI_ISL_4572140, EPI_ISL_4572141, EPI_ISL_4572142, EPI_ISL_4572143, EPI_ISL_4572144, EPI_ISL_4572145, EPI_ISL_4572146, EPI_ISL_4572148, EPI_ISL_4572149, EPI_ISL_4572150, EPI_ISL_4572200, EPI_ISL_4572201, EPI_ISL_4572202, EPI_ISL_4572203, EPI_ISL_4572204, EPI_ISL_4572205, EPI_ISL_4572206, EPI_ISL_4572207, EPI_ISL_4572208, EPI_ISL_4572209, EPI_ISL_4572210, EPI_ISL_4572212, EPI_ISL_4572213, EPI_ISL_4572224 | | | |
| see above | A.S.L. CUNEO 1 | Fondazione del Piemonte per l'Oncologia IRCCS | Antonino Sottile; Giorgio Giardina; Paola Marino; Silvia Brossa |
| EPI_ISL_4572120, EPI_ISL_4572211, EPI_ISL_4572214, EPI_ISL_4572216, EPI_ISL_4572217, EPI_ISL_4572218, EPI_ISL_4572219 | | | |
| see above | A.S.L. NOVARA | Fondazione del Piemonte per l'Oncologia IRCCS | Antonino Sottile; Giorgio Giardina; Paola Marino; Silvia Brossa |
| EPI_ISL_4572057, EPI_ISL_4572058, EPI_ISL_4572059, EPI_ISL_4572060, EPI_ISL_4572061, EPI_ISL_4572062, EPI_ISL_4572063, EPI_ISL_4572114, EPI_ISL_4572115 | | | |
| see above | A.S.L. TO3 | Fondazione del Piemonte per l'Oncologia IRCCS | Antonino Sottile; Giorgio Giardina; Paola Marino; Silvia Brossa |
| EPI_ISL_4572215 | A.S.L. TO4 | Fondazione del Piemonte per l'Oncologia IRCCS | Antonino Sottile; Giorgio Giardina; Paola Marino; Silvia Brossa |
| EPI_ISL_4436522, EPI_ISL_4436523, EPI_ISL_4436525 | A.S.L. TO5 | Fondazione del Piemonte per l'Oncologia IRCCS | Antonino Sottile; Giorgio Giardina; Paola Marino; Silvia Brossa |
| EPI_ISL_4436553, EPI_ISL_4436555, EPI_ISL_4436556, EPI_ISL_4436557, EPI_ISL_4436558, EPI_ISL_4436559, EPI_ISL_4436560, EPI_ISL_4436561, EPI_ISL_4572065, EPI_ISL_4572068, EPI_ISL_4572105, EPI_ISL_4572106, EPI_ISL_4572107, EPI_ISL_4572108, EPI_ISL_4572109, EPI_ISL_4572110, EPI_ISL_4572111, EPI_ISL_4572112, EPI_ISL_4572113 | | | |
| see above | A.S.L. VERCELLI | Fondazione del Piemonte per l'Oncologia IRCCS | Antonino Sottile; Giorgio Giardina; Paola Marino; Silvia Brossa |
| EPI_ISL_4571178, EPI_ISL_4571179, EPI_ISL_4571180, EPI_ISL_4571181, EPI_ISL_4571182, EPI_ISL_4571183, EPI_ISL_4571184, EPI_ISL_4571185, EPI_ISL_4571186, EPI_ISL_4571187, EPI_ISL_4571188, EPI_ISL_4571189, EPI_ISL_4571190, EPI_ISL_4571191, EPI_ISL_4571192 | | | |
| see above | AOPD | Istituto Zooprofilattico Sperimentale delle Venezie | Adelaide Milani; Alessia Schivo; Alice Fusaro; Ambra Pastori; Annalisa Salviato; Antonia Ricci; Calogero Terregino; Edoardo Giussani; Elisa Palumbo; Erika Giorgia Quaranta; Isabella Monne; Luca Tassoni |
| EPI_ISL_4571140, EPI_ISL_4571141, EPI_ISL_4571142, EPI_ISL_4571143, EPI_ISL_4571144 | AOUIVR | Istituto Zooprofilattico Sperimentale delle Venezie | Adelaide Milani; Alessia Schivo; Alice Fusaro; Ambra Pastori; Annalisa Salviato; Antonia Ricci; Calogero Terregino; Edoardo Giussani; Elisa Palumbo; Erika Giorgia Quaranta; Isabella Monne; Luca Tassoni |
| EPI_ISL_4524666, EPI_ISL_4525910, EPI_ISL_4527801, EPI_ISL_4527895, EPI_ISL_4550599, EPI_ISL_4550898 | ASL BARI | University of Bari Biomedical Sciences and Human Oncology | Maria Chironna |
| EPI_ISL_4683989, EPI_ISL_4685074, EPI_ISL_4685786, EPI_ISL_4686040, EPI_ISL_4686748, EPI_ISL_4687460, EPI_ISL_4688487, EPI_ISL_4690264, EPI_ISL_4692336 | | | |
| see above | ASL GENZANO | University Campus Bio-Medico of Rome (UCBM) | Angeletti S; Angeletti S.; De Florio L.; Fogolari M.; Francesconi M.; Lintas C.; Riva E.; Veralli R. |
| EPI_ISL_4498712 | ASL Genzano | University Campus Bio-Medico of Rome (UCBM | Angeletti S.; De Florio L.; Fogolari M.; Francesconi M.; Lintas C.; Riva E.; Veralli R. |
| EPI_ISL_4498716, EPI_ISL_4498718, EPI_ISL_4498721, EPI_ISL_4498722, EPI_ISL_4498723, EPI_ISL_4498724, EPI_ISL_4498725, EPI_ISL_4498727, EPI_ISL_4498728, EPI_ISL_4498729, EPI_ISL_4546691, EPI_ISL_4546798, EPI_ISL_4546799, EPI_ISL_4546956, EPI_ISL_4547205, EPI_ISL_4547514, EPI_ISL_4547651, EPI_ISL_4547698 | | | |
| see above | ASL Genzano | University Campus Bio-Medico of Rome (UCBM) | Angeletti S.; De Florio L.; Fogolari M.; Francesconi M.; Lintas C.; Riva E.; University Campus Bio-Medico of Rome (UCBM); Veralli R. |
| EPI_ISL_4498714 | ASL genzano | University Campus Bio-Medico of Rome (UCBM) | Angeletti S.; De Florio L.; Fogolari M.; Francesconi M.; Lintas C.; Riva E.; Veralli R. |
| EPI_ISL_4571165, EPI_ISL_4571166, EPI_ISL_4571167, EPI_ISL_4571168, EPI_ISL_4571169, EPI_ISL_4571170, EPI_ISL_4571171, EPI_ISL_4571172, EPI_ISL_4571173, EPI_ISL_4571175, EPI_ISL_4571176 | | | |
| see above | AULSS 2 Marca Trevigiana | Istituto Zooprofilattico Sperimentale delle Venezie | Adelaide Milani; Alessia Schivo; Alice Fusaro; Ambra Pastori; Annalisa Salviato; Antonia Ricci; Calogero Terregino; Edoardo Giussani; Elisa Palumbo; Erika Giorgia Quaranta; Isabella Monne; Luca Tassoni |
| EPI_ISL_4571122, EPI_ISL_4571123, EPI_ISL_4571124, EPI_ISL_4571125, EPI_ISL_4571126, EPI_ISL_4571127 | AULSS 5 Polesana | Istituto Zooprofilattico Sperimentale delle Venezie | Adelaide Milani; Alessia Schivo; Alice Fusaro; Ambra Pastori; Annalisa Salviato; Antonia Ricci; Calogero Terregino; Edoardo Giussani; Elisa Palumbo; Erika Giorgia Quaranta; Isabella Monne; Luca Tassoni |
| EPI_ISL_4571128, EPI_ISL_4571129, EPI_ISL_4571130, EPI_ISL_4571131, EPI_ISL_4571132, EPI_ISL_4571133, EPI_ISL_4571134, EPI_ISL_4571135, EPI_ISL_4571136, EPI_ISL_4571137, EPI_ISL_4571139 | | | |
| see above | AULSS 6 Euganea | Istituto Zooprofilattico Sperimentale delle Venezie | Adelaide Milani; Alessia Schivo; Alice Fusaro; Ambra Pastori; Annalisa Salviato; Antonia Ricci; Calogero Terregino; Edoardo Giussani; Elisa Palumbo; Erika Giorgia Quaranta; Isabella Monne; Luca Tassoni |
| EPI_ISL_4571156, EPI_ISL_4571157, EPI_ISL_4571158, EPI_ISL_4571159, EPI_ISL_4571160, EPI_ISL_4571162, EPI_ISL_4571163, EPI_ISL_4571164 | | | |
| see above | AULSS 7 Pedemontana - Bassano | Istituto Zooprofilattico Sperimentale delle Venezie | Adelaide Milani; Alessia Schivo; Alice Fusaro; Ambra Pastori; Annalisa Salviato; Antonia Ricci; Calogero Terregino; Edoardo Giussani; Elisa Palumbo; Erika Giorgia Quaranta; Isabella Monne; Luca Tassoni |
| EPI_ISL_4571146, EPI_ISL_4571152, EPI_ISL_4571154 | AULSS 7 Pedemontana - Santorso | Istituto Zooprofilattico Sperimentale delle Venezie | Adelaide Milani; Alessia Schivo; Alice Fusaro; Ambra Pastori; Annalisa Salviato; Antonia Ricci; Calogero Terregino; Edoardo Giussani; Elisa Palumbo; Erika Giorgia Quaranta; Isabella Monne; Luca Tassoni |
| EPI_ISL_4571105, EPI_ISL_4571106, EPI_ISL_4571107, EPI_ISL_4571108, EPI_ISL_4571109, EPI_ISL_4571110, EPI_ISL_4571111, EPI_ISL_4571112, EPI_ISL_4571113, EPI_ISL_4571114, EPI_ISL_4603807 | | | |
| see above | AULSS 9 Scaligera - Legnago | Istituto Zooprofilattico Sperimentale delle Venezie | Adelaide Milani; Alessia Schivo; Alice Fusaro; Ambra Pastori; Annalisa Salviato; Antonia Ricci; Calogero Terregino; Edoardo Giussani; Elisa Palumbo; Erika Giorgia Quaranta; Isabella Monne; Luca Tassoni |
| EPI_ISL_4571001, EPI_ISL_4571002 | Azienda Ospedaliera Universitaria San Andrea | University Campus Bio-Medico of Rome (UCBM) | Angeletti S.; De Florio L.; Fogolari M.; Francesconi M.; Lintas C.; Riva E.; Veralli R. |
| EPI_ISL_4571197 | Azienda Sanitaria Locale Viterbo, Ospedale di Belcolle-Viterbo | University Campus Bio-Medico of Rome (UCBM) | Angeletti S.; De Florio L.; Fogolari M.; Francesconi M.; Lintas C.; Riva E.; Veralli R. |
| EPI_ISL_4498690, EPI_ISL_4498693, EPI_ISL_4498697, EPI_ISL_4498699 | Data Medica-Rome-Italy | University Campus Bio-Medico of Rome (UCBM) | Angeletti S.; De Florio L.; Fogolari M.; Francesconi M.; Lintas C.; Riva E.; Veralli R. |
| EPI_ISL_4270081, EPI_ISL_4270085, EPI_ISL_4270099, EPI_ISL_4270102, EPI_ISL_4305186, EPI_ISL_4305190, EPI_ISL_4443070, EPI_ISL_4443072, EPI_ISL_4443075, EPI_ISL_4443077, EPI_ISL_4443079, EPI_ISL_4443081, EPI_ISL_4443083, EPI_ISL_4443086, EPI_ISL_4443088, EPI_ISL_4443091, EPI_ISL_4443093, EPI_ISL_4443096, EPI_ISL_4443098, EPI_ISL_4443099, EPI_ISL_4443100, EPI_ISL_4443105, EPI_ISL_4443110, EPI_ISL_4443115, EPI_ISL_4443119, EPI_ISL_4443124, EPI_ISL_4443128, EPI_ISL_4443131, EPI_ISL_4443133, EPI_ISL_4443135, EPI_ISL_4443137, EPI_ISL_4443139, EPI_ISL_4443141, EPI_ISL_4443143, EPI_ISL_4443145, EPI_ISL_4443147, EPI_ISL_4508166, EPI_ISL_4508167, EPI_ISL_4508168, EPI_ISL_4508169, EPI_ISL_4508170, EPI_ISL_4508171, EPI_ISL_4508172, EPI_ISL_4508173, EPI_ISL_4533451, EPI_ISL_4533452, EPI_ISL_4533453, EPI_ISL_4533454, EPI_ISL_4533455, EPI_ISL_4533456, EPI_ISL_4533457, EPI_ISL_4533458, EPI_ISL_4533459, EPI_ISL_4533460, EPI_ISL_4533461, EPI_ISL_4533462, EPI_ISL_4533463, EPI_ISL_4533760, EPI_ISL_4533761, EPI_ISL_4533762, EPI_ISL_4533763, EPI_ISL_4533764, EPI_ISL_4533765, EPI_ISL_4533766, EPI_ISL_4533767, EPI_ISL_4533768, EPI_ISL_4533769, EPI_ISL_4533770, EPI_ISL_4533771, EPI_ISL_4541033, EPI_ISL_4541035, EPI_ISL_4541037, EPI_ISL_4541039, EPI_ISL_4541041, EPI_ISL_4541043, EPI_ISL_4541044, EPI_ISL_4541046, EPI_ISL_4541048, EPI_ISL_4541050, EPI_ISL_4541051, EPI_ISL_4541053, EPI_ISL_4541055, EPI_ISL_4553159, EPI_ISL_4553160, EPI_ISL_4553161, EPI_ISL_4553162, EPI_ISL_4553163, EPI_ISL_4553164, EPI_ISL_4553165, EPI_ISL_4563803, EPI_ISL_4563804, EPI_ISL_4563805, EPI_ISL_4563806, EPI_ISL_4563807, EPI_ISL_4563808, EPI_ISL_4563809, EPI_ISL_4563810, EPI_ISL_4563811, EPI_ISL_4563812, EPI_ISL_4563813, EPI_ISL_4563814, EPI_ISL_4563815, EPI_ISL_4563816, EPI_ISL_4563817, EPI_ISL_4563818, EPI_ISL_4563819, EPI_ISL_4563820, EPI_ISL_4563821, EPI_ISL_4563822, EPI_ISL_4563823, EPI_ISL_4563824, EPI_ISL_4563825, EPI_ISL_4563826, EPI_ISL_4563827, EPI_ISL_4563828, EPI_ISL_4563829, EPI_ISL_4563830, EPI_ISL_4563831, EPI_ISL_4563832, EPI_ISL_4563833, EPI_ISL_4563834, EPI_ISL_4563835, EPI_ISL_4563836, EPI_ISL_4563837, EPI_ISL_4563838, EPI_ISL_4563839, EPI_ISL_4629416, EPI_ISL_4633982, EPI_ISL_4633983, EPI_ISL_4633984, EPI_ISL_4633985, EPI_ISL_4633986, EPI_ISL_4633987, EPI_ISL_4633988, EPI_ISL_4633989, EPI_ISL_4633990, EPI_ISL_4633991, EPI_ISL_4633992, EPI_ISL_4633993, EPI_ISL_4633994, EPI_ISL_4633995, EPI_ISL_4633996, EPI_ISL_4633997, EPI_ISL_4633998, EPI_ISL_4633999, EPI_ISL_4634000, EPI_ISL_4634001, EPI_ISL_4634002, EPI_ISL_4634003, EPI_ISL_4634004, EPI_ISL_4634005, EPI_ISL_4634006, EPI_ISL_4634007, EPI_ISL_4634008, EPI_ISL_4634009, EPI_ISL_4634010, EPI_ISL_4634011, EPI_ISL_4634012, EPI_ISL_4634013, EPI_ISL_4634014, EPI_ISL_4634015, EPI_ISL_4634016, EPI_ISL_4634017, EPI_ISL_4672096, EPI_ISL_4672109, EPI_ISL_4672120, EPI_ISL_4672126, EPI_ISL_4672149, EPI_ISL_4672163, EPI_ISL_4672171, EPI_ISL_4672179 | | | |
| see above | Dipartimento di Medicina di Laboratorio, Azienda sanitaria universitaria Friuli Centrale (ASU FC) | Dipartimento di Medicina di Laboratorio, Azienda sanitaria universitaria Friuli Centrale (ASU FC) | Catia Mio; Chiara Dal Secco; Corrado Pipan; Federica Baldan; Francesco Curcio; Stefania Marzinotto |
| EPI_ISL_4545890, EPI_ISL_4545891, EPI_ISL_4545892, EPI_ISL_4545894, EPI_ISL_4546089 | Eurofins Genoma Group S.r.l. (Rome) | University Campus Bio-Medico of Rome (UCBM) | Angeletti S.; De Florio L.; Fogolari M.; Francesconi M.; Lintas C.; Riva E.; Veralli R. |
| EPI_ISL_4339180, EPI_ISL_4339182, EPI_ISL_4339185, EPI_ISL_4339188, EPI_ISL_4339191, EPI_ISL_4339193, EPI_ISL_4339197, EPI_ISL_4339200, EPI_ISL_4339203, EPI_ISL_4339205 | | | |
| see above | Fondazione IRCCS Ca' Granda Ospedale Maggiore Policlinico | Fondazione IRCCS Ca' Granda Ospedale Maggiore Policlinico | Ferruccio Ceriotti; Sara Uceda Renteria |
| EPI_ISL_4256817, EPI_ISL_4256818 | Fondazione del Piemonte per l'Oncologia IRCCS | Fondazione del Piemonte per l'Oncologia IRCCS | Antonino Sottile; Giorgio Giardina; Paola Marino; Silvia Brossa |
| EPI_ISL_4546368, EPI_ISL_4546546 | INMI Lazzaro Spallanzani - Istituto Nazionale Malattie Infettive | University Campus Bio-Medico of Rome (UCBM) | Angeletti S.; De Florio L.; Fogolari M.; Francesconi M.; Lintas C.; Riva E.; Veralli R. |
| EPI_ISL_4223134, EPI_ISL_4223136, EPI_ISL_4223138, EPI_ISL_4223140, EPI_ISL_4223142, EPI_ISL_4223144, EPI_ISL_4223146, EPI_ISL_4472532, EPI_ISL_4472533, EPI_ISL_4472535, EPI_ISL_4472536, EPI_ISL_4472537, EPI_ISL_4472538, EPI_ISL_4472539 | | | |
| see above | Istituto Auxologico Italiano | Istituto Auxologico Italiano | Erminio Torresani |
| EPI_ISL_4498835, EPI_ISL_4498836, EPI_ISL_4498837, EPI_ISL_4498838, EPI_ISL_4498839, EPI_ISL_4687991, EPI_ISL_4688321, EPI_ISL_4688718, EPI_ISL_4689095, EPI_ISL_4689521, EPI_ISL_4690060, EPI_ISL_4690814, EPI_ISL_4694398, EPI_ISL_4694823 | | | |

| | | | |
|---|---|---|---|
| see above | LIFEBRAIN SRL GUIDONIA | University Campus Bio-Medico of Rome (UCBM) | Angeletti S.; De Florio L.; Fogolari M.; Francesconi M.; Lintas C.; Riva E.; Veralli R. |
| EPI_ISL_4525723, EPI_ISL_4525724, EPI_ISL_4525725, EPI_ISL_4525726, EPI_ISL_4525727, EPI_ISL_4525728, EPI_ISL_4525729, EPI_ISL_4525730, EPI_ISL_4525731, EPI_ISL_4525732, EPI_ISL_4525733, EPI_ISL_4525734, EPI_ISL_4525735, EPI_ISL_4525736, EPI_ISL_4525737, EPI_ISL_4525738, EPI_ISL_4525739, EPI_ISL_4525740, EPI_ISL_4525741, EPI_ISL_4525742, EPI_ISL_4525743, EPI_ISL_4525744, EPI_ISL_4525745, EPI_ISL_4525746, EPI_ISL_4525747, EPI_ISL_4525748, EPI_ISL_4525749, EPI_ISL_4525750, EPI_ISL_4525751, EPI_ISL_4525752, EPI_ISL_4525753, EPI_ISL_4525754, EPI_ISL_4525755, EPI_ISL_4525756, EPI_ISL_4525757, EPI_ISL_4525758, EPI_ISL_4525759, EPI_ISL_4525760, EPI_ISL_4525761, EPI_ISL_4525779, EPI_ISL_4525821, EPI_ISL_4525822 | | | |
| see above | Lab. Microbiologia e Virologia Cotugno A.O. dei Colli | TIGEM | Antonio Grimaldi Patrizia Annunziata Francesco Panariello Claudia Tiberio Teresa Giuliano Valentina Bouche Chiara Colantuono Lucio Di Filippo Anna Manfredi Marcello Salvi Antonio Limone Luigi Atripaldi Andrea Ballabio Davide Cacchiarelli |
| EPI_ISL_4628049, EPI_ISL_4628057, EPI_ISL_4628058, EPI_ISL_4628067, EPI_ISL_4628077, EPI_ISL_4628078, EPI_ISL_4628086, EPI_ISL_4628092, EPI_ISL_4628099, EPI_ISL_4628100, EPI_ISL_4628106, EPI_ISL_4628111, EPI_ISL_4628112, EPI_ISL_4628118, EPI_ISL_4628121, EPI_ISL_4628122, EPI_ISL_4628124, EPI_ISL_4628129, EPI_ISL_4628130, EPI_ISL_4628135, EPI_ISL_4628142, EPI_ISL_4628143, EPI_ISL_4628148, EPI_ISL_4628154, EPI_ISL_4628160, EPI_ISL_4628161, EPI_ISL_4628171, EPI_ISL_4628181, EPI_ISL_4628187, EPI_ISL_4628196, EPI_ISL_4628197, EPI_ISL_4628201, EPI_ISL_4628202, EPI_ISL_4628209, EPI_ISL_4628213, EPI_ISL_4628216, EPI_ISL_4628217, EPI_ISL_4628221, EPI_ISL_4628228, EPI_ISL_4628229, EPI_ISL_4628232, EPI_ISL_4628235, EPI_ISL_4628239, EPI_ISL_4628245, EPI_ISL_4628252, EPI_ISL_4628253 | | | |
| see above | Laboratorio CQRC | CQRC_QUALITY CONTROL CHEMICAL BIOLOGICAL RISK_AOOR Villa Sofia Cervello Palermo | Brunacci G.; Contino F.; Di Gaudio F. |
| EPI_ISL_4498840, EPI_ISL_4498841 | Laboratorio GENOMA | University Campus Bio-Medico of Rome (UCBM) | Angeletti S.; De Florio L.; Fogolari M.; Francesconi M.; Lintas C.; Riva E.; Veralli R. |
| EPI_ISL_4571288, EPI_ISL_4571289, EPI_ISL_4571290, EPI_ISL_4571291, EPI_ISL_4571292, EPI_ISL_4571293, EPI_ISL_4572000, EPI_ISL_4572045, EPI_ISL_4572046, EPI_ISL_4572047, EPI_ISL_4572048, EPI_ISL_4572049, EPI_ISL_4572050, EPI_ISL_4572124, EPI_ISL_4572151, EPI_ISL_4572198, EPI_ISL_4572199, EPI_ISL_4666299, EPI_ISL_4666747, EPI_ISL_4666969, EPI_ISL_4667532, EPI_ISL_4668675, EPI_ISL_4669037, EPI_ISL_4669306, EPI_ISL_4670622, EPI_ISL_4672482, EPI_ISL_4674847, EPI_ISL_4675845, EPI_ISL_4677810, EPI_ISL_4681400 | | | |
| see above | Microbiology and Virology IFO, Rome, Italy | University Campus Bio-Medico of Rome (UCBM) | Angeletti S; Angeletti S.; De Florio L.; Fogolari M.; Francesconi M.; Lintas C.; Riva E.; Veralli R. |
| EPI_ISL_4506002, EPI_ISL_4506004, EPI_ISL_4506008, EPI_ISL_4506009, EPI_ISL_4506010, EPI_ISL_4506011, EPI_ISL_4506012, EPI_ISL_4506013, EPI_ISL_4506014 | | | |
| see above | Ospedale "San Francesco", ASSL Nuoro | Laboratorio Biologia Molecolare Sars Cov2 - UOC Laboratorio Analisi - Servizio Medicina di Laboratorio, Ospedale "San Francesco" - ATS-ASSL Nuoro and Laboratorio specialistico UOC Ematologia - Ospedale "San Francesco" - ATS-ASSL Nuoro | Asproni Rosanna; Carta Franco; Fancello Patrizia; Fiamma Maura; Garau Maria Cristina; Mameli Giuseppe; Palmas Angelo Domenico; Pira Giovanna; Piras Giovanna; Rosu Valentina |
| EPI_ISL_4570378 | Ospedale GB Grassi | University Campus Bio-Medico of Rome (UCBM) | Angeletti S.; De Florio L.; Fogolari M.; Francesconi M.; Lintas C.; Riva E.; Veralli R. |
| EPI_ISL_4454404, EPI_ISL_4454579, EPI_ISL_4454939, EPI_ISL_4455282, EPI_ISL_4455549, EPI_ISL_4456015, EPI_ISL_4456186, EPI_ISL_4544826, EPI_ISL_4544984, EPI_ISL_4545120, EPI_ISL_4545316, EPI_ISL_4545382, EPI_ISL_4545398, EPI_ISL_4545498, EPI_ISL_4545499, EPI_ISL_4545500, EPI_ISL_4545502, EPI_ISL_4545503, EPI_ISL_4545504, EPI_ISL_4545548, EPI_ISL_4545549, EPI_ISL_4545611, EPI_ISL_4545713, EPI_ISL_4545715, EPI_ISL_4545853, EPI_ISL_4545889 | | | |
| see above | Ospedale San Giovanni Evangelista (Tivoli) ASLRM5 | University Campus Bio-Medico of Rome (UCBM) | Angeletti S.; De Florio L.; Fogolari M.; Francesconi M.; Lintas C.; Riva E.; Veralli R. |
| EPI_ISL_4498688, EPI_ISL_4498689, EPI_ISL_4498694, EPI_ISL_4498695 | Ospedale Sant'Eugenio-Rome | University Campus Bio-Medico of Rome (UCBM) | Angeletti S.; De Florio L.; Fogolari M.; Francesconi M.; Lintas C.; Riva E.; Veralli R. |
| EPI_ISL_4455717, EPI_ISL_4456399, EPI_ISL_4456538, EPI_ISL_4456655 | Ospedale Santa Maria Goretti-Latina-Italy | University Campus Bio-Medico of Rome (UCBM) | Angeletti S.; De Florio L.; Fogolari M.; Francesconi M.; Lintas C.; Riva E.; Veralli R. |
| EPI_ISL_4471556, EPI_ISL_4471560, EPI_ISL_4471562, EPI_ISL_4471563, EPI_ISL_4471564, EPI_ISL_4471565, EPI_ISL_4571006, EPI_ISL_4571007, EPI_ISL_4571008, EPI_ISL_4571009, EPI_ISL_4571010, EPI_ISL_4571011 | | | |
| see above | PTV-Università di Tor vergata-Rome | University Campus Bio-Medico of Rome (UCBM) | Angeletti S.; De Florio L.; Fogolari M.; Francesconi M.; Lintas C.; Riva E.; Veralli R. |
| EPI_ISL_4470546, EPI_ISL_4470547, EPI_ISL_4470549, EPI_ISL_4470550, EPI_ISL_4470551, EPI_ISL_4470554 | Presidio Ospedaliero "Madonna delle Grazie" di Matera | ASM - P.O. Madonna delle Grazie - Matera | Massimo Dell'Edera |
| EPI_ISL_4471515, EPI_ISL_4471567, EPI_ISL_4471568, EPI_ISL_4471615 | SYNLAB | University Campus Bio-Medico of Rome (UCBM) | Angeletti S.; De Florio L.; Fogolari M.; Francesconi M.; Lintas C.; Riva E.; Veralli R. |
| EPI_ISL_4170822, EPI_ISL_4170823, EPI_ISL_4170824, EPI_ISL_4170825, EPI_ISL_4170826, EPI_ISL_4170827, EPI_ISL_4170828, EPI_ISL_4170829, EPI_ISL_4170830, EPI_ISL_4170831, EPI_ISL_4170832, EPI_ISL_4170833, EPI_ISL_4170834, EPI_ISL_4170835, EPI_ISL_4170836, EPI_ISL_4170837, EPI_ISL_4170838, EPI_ISL_4170839, EPI_ISL_4170840, EPI_ISL_4170841, EPI_ISL_4170842, EPI_ISL_4170843, EPI_ISL_4170844, EPI_ISL_4170845, EPI_ISL_4170846, EPI_ISL_4170847, EPI_ISL_4170848, EPI_ISL_4170849, EPI_ISL_4170850, EPI_ISL_4170851, EPI_ISL_4170852, EPI_ISL_4170853, EPI_ISL_4170854, EPI_ISL_4170855, EPI_ISL_4170856, EPI_ISL_4170857, EPI_ISL_4170858, EPI_ISL_4170859, EPI_ISL_4170860, EPI_ISL_4357198, EPI_ISL_4357200, EPI_ISL_4357202, EPI_ISL_4357203, EPI_ISL_4357205, EPI_ISL_4357207, EPI_ISL_4357209, EPI_ISL_4357210, EPI_ISL_4357212, EPI_ISL_4357214, EPI_ISL_4357216, EPI_ISL_4357218, EPI_ISL_4357219, EPI_ISL_4357221, EPI_ISL_4357223, EPI_ISL_4357225, EPI_ISL_4357226, EPI_ISL_4357228, EPI_ISL_4357230, EPI_ISL_4357232, EPI_ISL_4357233, EPI_ISL_4357235, EPI_ISL_4357237, EPI_ISL_4357239, EPI_ISL_4357241, EPI_ISL_4357242, EPI_ISL_4357244, EPI_ISL_4357246, EPI_ISL_4357248, EPI_ISL_4357250, EPI_ISL_4357251, EPI_ISL_4357891, EPI_ISL_4357893, EPI_ISL_4357894, EPI_ISL_4357895, EPI_ISL_4357896, EPI_ISL_4357897, EPI_ISL_4357899, EPI_ISL_4357900, EPI_ISL_4357902, EPI_ISL_4357904, EPI_ISL_4357906, EPI_ISL_4357907, EPI_ISL_4357909, EPI_ISL_4357911, EPI_ISL_4357912, EPI_ISL_4357914, EPI_ISL_4357916, EPI_ISL_4357917, EPI_ISL_4357919, EPI_ISL_4357920, EPI_ISL_4357922, EPI_ISL_4357924, EPI_ISL_4357926, EPI_ISL_4357927, EPI_ISL_4357929, EPI_ISL_4357931, EPI_ISL_4357933, EPI_ISL_4357934, EPI_ISL_4357936, EPI_ISL_4357938, EPI_ISL_4357940, EPI_ISL_4357942, EPI_ISL_4357943, EPI_ISL_4357945, EPI_ISL_4357946, EPI_ISL_4357948, EPI_ISL_4357949, EPI_ISL_4357951, EPI_ISL_4358522, EPI_ISL_4358524, EPI_ISL_4358526, EPI_ISL_4358528, EPI_ISL_4358529, EPI_ISL_4358531, EPI_ISL_4358533, EPI_ISL_4358534, EPI_ISL_4358536, EPI_ISL_4358538, EPI_ISL_4358540, EPI_ISL_4358542, EPI_ISL_4358543, EPI_ISL_4358545, EPI_ISL_4358547, EPI_ISL_4358548, EPI_ISL_4358550, EPI_ISL_4358552, EPI_ISL_4358553, EPI_ISL_4358555, EPI_ISL_4358557, EPI_ISL_4358558, EPI_ISL_4358560, EPI_ISL_4358561, EPI_ISL_4358563, EPI_ISL_4358565, EPI_ISL_4358566, EPI_ISL_4358568, EPI_ISL_4358570, EPI_ISL_4358571, EPI_ISL_4358573, EPI_ISL_4358575, EPI_ISL_4358577, EPI_ISL_4358578, EPI_ISL_4358580, EPI_ISL_4358582, EPI_ISL_4358583, EPI_ISL_4638340, EPI_ISL_4638341, EPI_ISL_4638342, EPI_ISL_4638343, EPI_ISL_4638344, EPI_ISL_4638345, EPI_ISL_4638346, EPI_ISL_4638347, EPI_ISL_4638348, EPI_ISL_4638349, EPI_ISL_4638350, EPI_ISL_4638351, EPI_ISL_4638352, EPI_ISL_4638353, EPI_ISL_4638354, EPI_ISL_4638355, EPI_ISL_4638356, EPI_ISL_4638357, EPI_ISL_4638358, EPI_ISL_4638359, EPI_ISL_4638360, EPI_ISL_4638361, EPI_ISL_4638362, EPI_ISL_4638363, EPI_ISL_4638364, EPI_ISL_4638365, EPI_ISL_4638366, EPI_ISL_4638367, EPI_ISL_4638368, EPI_ISL_4638369, EPI_ISL_4638370, EPI_ISL_4638371, EPI_ISL_4638372, EPI_ISL_4638373, EPI_ISL_4638374, EPI_ISL_4638375, EPI_ISL_4638376, EPI_ISL_4638377, EPI_ISL_4638378, EPI_ISL_4638381, EPI_ISL_4638382, EPI_ISL_4638383, EPI_ISL_4638384, EPI_ISL_4638385, EPI_ISL_4638386, EPI_ISL_4638387, EPI_ISL_4638388, EPI_ISL_4638389, EPI_ISL_4638390, EPI_ISL_4638391, EPI_ISL_4638392, EPI_ISL_4638393, EPI_ISL_4638394, EPI_ISL_4638395, EPI_ISL_4638396, EPI_ISL_4638397, EPI_ISL_4638398, EPI_ISL_4638399, EPI_ISL_4638400, EPI_ISL_4638401, EPI_ISL_4638402, EPI_ISL_4638403, EPI_ISL_4638404, EPI_ISL_4638405, EPI_ISL_4638406, EPI_ISL_4638407, EPI_ISL_4638408, EPI_ISL_4638409, EPI_ISL_4638410, EPI_ISL_4638411, EPI_ISL_4638412, EPI_ISL_4638413, EPI_ISL_4638414, EPI_ISL_4638415, EPI_ISL_4638416, EPI_ISL_4638417, EPI_ISL_4638418, EPI_ISL_4638451, EPI_ISL_4638452, EPI_ISL_4638453, EPI_ISL_4638454, EPI_ISL_4638455, EPI_ISL_4638456, EPI_ISL_4638457, EPI_ISL_4638458, EPI_ISL_4638459, EPI_ISL_4638460, EPI_ISL_4638461, EPI_ISL_4638462, EPI_ISL_4638463, EPI_ISL_4638464, EPI_ISL_4638466, EPI_ISL_4638467, EPI_ISL_4638468, EPI_ISL_4638469, EPI_ISL_4638470, EPI_ISL_4638471, EPI_ISL_4638472, EPI_ISL_4638474, EPI_ISL_4638475, EPI_ISL_4638476, EPI_ISL_4638477, EPI_ISL_4638478, EPI_ISL_4638479, EPI_ISL_4638480, EPI_ISL_4638481, EPI_ISL_4638482, EPI_ISL_4638483, EPI_ISL_4638484, EPI_ISL_4638485, EPI_ISL_4638486, EPI_ISL_4638487, EPI_ISL_4638488, EPI_ISL_4638489 | | | |
| see above | U.O. Microbiologia Laboratorio Unico Centro Servizi - AUSL della Romagna | U.O. Microbiologia, Laboratorio Unico Centro Servizi - AUSL della Romagna | Giorgio Dirani |
| EPI_ISL_4571012, EPI_ISL_4571013, EPI_ISL_4571014, EPI_ISL_4571015, EPI_ISL_4571016, EPI_ISL_4571017, EPI_ISL_4571018, EPI_ISL_4571019, EPI_ISL_4571020, EPI_ISL_4571021, EPI_ISL_4571096, EPI_ISL_4571098, EPI_ISL_4571099, EPI_ISL_4571100, EPI_ISL_4571102, EPI_ISL_4571103, EPI_ISL_4571193 | | | |
| see above | UOC Microbiologia e Virologia, Ospedale San Filippo Neri-Rome-Italy | University Campus Bio-Medico of Rome (UCBM) | Angeletti S.; De Florio L.; Fogolari M.; Francesconi M.; Lintas C.; Riva E.; Veralli R. |
| EPI_ISL_4470555, EPI_ISL_4470556, EPI_ISL_4470557, EPI_ISL_4470558, EPI_ISL_4470559, EPI_ISL_4470560, EPI_ISL_4470561, EPI_ISL_4470562, EPI_ISL_4470563, EPI_ISL_4470564, EPI_ISL_4470565, EPI_ISL_4470567, EPI_ISL_4470608, EPI_ISL_4470609 | | | |
| see above | UOC Microbiologia e Virologia, Ospedale San Filippo Neri-Rome-italy | University Campus Bio-Medico of Rome (UCBM) | Angeletti S.; De Florio L.; Fogolari M.; Francesconi M.; Lintas C.; Riva E.; Veralli R. |
| EPI_ISL_4471553, EPI_ISL_4471554, EPI_ISL_4491148, EPI_ISL_4491672, EPI_ISL_4491821, EPI_ISL_4492079, EPI_ISL_4492233, EPI_ISL_4493113, EPI_ISL_4493114, EPI_ISL_4493150, EPI_ISL_4498687, EPI_ISL_4498691, EPI_ISL_4498692 | | | |
| see above | UOC Microbiologia e Virologia, Ospedale Sandro Pertini-Rome-Italy | University Campus Bio-Medico of Rome (UCBM) | Angeletti S.; De Florio L.; Fogolari M.; Francesconi M.; Lintas C.; Riva E.; Veralli R. |
| EPI_ISL_4498686 | UOC Microbiologia e Virologia, Ospedale Sandro Pertini-Rome-Italy | University Campus Bio-Medico of Rome (UCBM) Via A. del Portillo, 200 - Rome - Italy | Angeletti S.; De Florio L.; Fogolari M.; Francesconi M.; Lintas C.; Riva E.; Veralli R. |
| EPI_ISL_4498842, EPI_ISL_4498843, EPI_ISL_4498844, EPI_ISL_4498845, EPI_ISL_4498846 | UOC Microbiology and Virology Policlinico Umberto I Rome, Italy | University Campus Bio-Medico of Rome (UCBM) | Angeletti S.; De Florio L.; Fogolari M.; Francesconi M.; Lintas C.; Riva E.; Veralli R. |
| EPI_ISL_4570515 | UOC Patologia Clinica AO San Giovanni Addolorata Hospital-Rome | University Campus Bio-Medico of Rome (UCBM) | Angeletti S.; De Florio L.; Fogolari M.; Francesconi M.; Lintas C.; Riva E.; Veralli R. |
| EPI_ISL_4498847, EPI_ISL_4498848, EPI_ISL_4498849, EPI_ISL_4498850, EPI_ISL_4498851, EPI_ISL_4571003, EPI_ISL_4571004, EPI_ISL_4571005 | | | |
| see above | UOC Patologia Clinica, Ospedale Fabrizio Spaziani, Frosinone, Italy | University Campus Bio-Medico of Rome (UCBM) | Angeletti S.; De Florio L.; Fogolari M.; Francesconi M.; Lintas C.; Riva E.; Veralli R. |
| EPI_ISL_4498696, EPI_ISL_4498698, EPI_ISL_4498700, EPI_ISL_4498701, EPI_ISL_4498702, EPI_ISL_4498703, EPI_ISL_4498708, EPI_ISL_4498710 | | | |
| see above | UOSD Laboratorio Analisi, Ospedale San Camillo de Lellis – Rieti-Italy | University Campus Bio-Medico of Rome (UCBM) | Angeletti S.; De Florio L.; Fogolari M.; Francesconi M.; Lintas C.; Riva E.; Veralli R. |
| EPI_ISL_4454352, EPI_ISL_4454353, EPI_ISL_4470813, EPI_ISL_4471033, EPI_ISL_4471123, EPI_ISL_4471201, EPI_ISL_4471351, EPI_ISL_4471516, EPI_ISL_4544048, EPI_ISL_4544327, EPI_ISL_4544560, EPI_ISL_4685131, EPI_ISL_4685826, EPI_ISL_4685928, EPI_ISL_4686209, EPI_ISL_4686826, EPI_ISL_4687223 | | | |
| see above | University Campus Bio-Medico of Rome (UCBM) | University Campus Bio-Medico of Rome (UCBM) | Angeletti S.; De Florio L.; Fogolari M.; Francesconi M.; Lintas C.; Riva E.; Veralli R. |
