## Supplementary material for "SARS-COV-2 δ variant drives the pandemic in India and Europe via two subvariants": Acknowledgement table on the GISAID genomes used in this study

We gratefully acknowledge the following Authors from the Originating laboratories responsible for obtaining the specimens, as well as the Submitting laboratories where the genome data were generated and shared via GISAID, on which this research is based.

All Submitters of data may be contacted directly via www.gisaid.org

Authors are sorted alphabetically.

| Accession ID | Originating Laboratory | Submitting Laboratory | Authors |
|---|---|---|---|
| EPI_ISL_4606197, EPI_ISL_4606198, EPI_ISL_4606199, EPI_ISL_4606200, EPI_ISL_4606201, EPI_ISL_4606202, EPI_ISL_4606203, EPI_ISL_4606204, EPI_ISL_4606205, EPI_ISL_4606206, EPI_ISL_4606207, EPI_ISL_4606208, EPI_ISL_4606209, EPI_ISL_4606210, EPI_ISL_4606211 | | | |
| see above | ASTRALAB TASSIGNY | CNR Virus des Infections Respiratoires - France SUD | Antonin Bal; Bruno Lina; Gregory Destras; Gwendolyne Burfin; Hadrien Regue; Laurence Josset; Martine Valette; Quentin Semanas |
| EPI_ISL_4606261, EPI_ISL_4606262, EPI_ISL_4606263, EPI_ISL_4606264, EPI_ISL_4606265, EPI_ISL_4606267, EPI_ISL_4606268, EPI_ISL_4606269, EPI_ISL_4606270, EPI_ISL_4606271, EPI_ISL_4606272, EPI_ISL_4606273, EPI_ISL_4606274, EPI_ISL_4606275, EPI_ISL_4606276, EPI_ISL_4606277, EPI_ISL_4606278, EPI_ISL_4606279, EPI_ISL_4606280, EPI_ISL_4606281, EPI_ISL_4606950, EPI_ISL_4606951, EPI_ISL_4606954, EPI_ISL_4606960, EPI_ISL_4606961 | | | |
| see above | AX BIO OCEAN | CNR Virus des Infections Respiratoires - France SUD | Antonin Bal; Bruno Lina; Gregory Destras; Gwendolyne Burfin; Hadrien Regue; Laurence Josset; Martine Valette; Quentin Semanas |
| EPI_ISL_4186460 | Arlon | Plateforme de testing Namuroise | Degosserie Jonathan; Demars Aurore; Denis Olivier; Maschietto Céline; Mullier François; Nobis Chloé; Nyinkeu Kemamen Lesly; Otto Gaetan |
| EPI_ISL_4503713, EPI_ISL_4503714, EPI_ISL_4503768, EPI_ISL_4600727, EPI_ISL_4600728 | Armies | National Reference Center for Viruses of Respiratory Infections, Institut Pasteur, Paris | Angela Brisebarre; Camille Capel; Christophe Malabat; Corinne Maufrais; Etienne Simon-Lorière; Frédéric Lemoine; Julien Fumey; Louise Lefrançois; Marine Desroches; Marion Barbet; Maud Vanpeene; Méline Bizard; Slim El Khiari; Sylvie Behillil; Sylvie Van der Werf; Vincent Enouf |
| EPI_ISL_4607026, EPI_ISL_4607027, EPI_ISL_4607028, EPI_ISL_4607029, EPI_ISL_4607030, EPI_ISL_4607034, EPI_ISL_4607044, EPI_ISL_4607046, EPI_ISL_4607070, EPI_ISL_4607071, EPI_ISL_4607072, EPI_ISL_4607073, EPI_ISL_4607074 | | | |
| see above | BIOFFICE - LBM RAVEZIES | CNR Virus des Infections Respiratoires - France SUD | Antonin Bal; Bruno Lina; Gregory Destras; Gwendolyne Burfin; Hadrien Regue; Laurence Josset; Martine Valette; Quentin Semanas |
| EPI_ISL_4605280, EPI_ISL_4605284, EPI_ISL_4605293, EPI_ISL_4605306, EPI_ISL_4605315, EPI_ISL_4605318, EPI_ISL_4605320, EPI_ISL_4605330, EPI_ISL_4605331, EPI_ISL_4605332, EPI_ISL_4605337, EPI_ISL_4605338, EPI_ISL_4605340, EPI_ISL_4605357, EPI_ISL_4605362, EPI_ISL_4605369, EPI_ISL_4605371, EPI_ISL_4605372, EPI_ISL_4605376, EPI_ISL_4605383, EPI_ISL_4605391, EPI_ISL_4606987, EPI_ISL_4606989, EPI_ISL_4606992, EPI_ISL_4606995, EPI_ISL_4607001, EPI_ISL_4607004, EPI_ISL_4607008, EPI_ISL_4607012, EPI_ISL_4607015, EPI_ISL_4607019, EPI_ISL_4607021, EPI_ISL_4607022, EPI_ISL_4607023, EPI_ISL_4607024, EPI_ISL_4607025 | | | |
| see above | BIOLAB 33 | CNR Virus des Infections Respiratoires - France SUD | Antonin Bal; Bruno Lina; Gregory Destras; Gwendolyne Burfin; Hadrien Regue; Laurence Josset; Martine Valette; Quentin Semanas |
| EPI_ISL_4605947, EPI_ISL_4605948, EPI_ISL_4605949, EPI_ISL_4605950, EPI_ISL_4605951, EPI_ISL_4605952, EPI_ISL_4605953, EPI_ISL_4605954, EPI_ISL_4606017, EPI_ISL_4606020, EPI_ISL_4606023, EPI_ISL_4606025, EPI_ISL_4606027, EPI_ISL_4606029, EPI_ISL_4606032, EPI_ISL_4606034, EPI_ISL_4606036, EPI_ISL_4606039, EPI_ISL_4606041, EPI_ISL_4606044, EPI_ISL_4606047, EPI_ISL_4606104, EPI_ISL_4606105, EPI_ISL_4606107, EPI_ISL_4606108, EPI_ISL_4606109, EPI_ISL_4606110, EPI_ISL_4606111, EPI_ISL_4606113, EPI_ISL_4606114, EPI_ISL_4606115, EPI_ISL_4606116, EPI_ISL_4606117, EPI_ISL_4606118, EPI_ISL_4606119, EPI_ISL_4606120, EPI_ISL_4606121, EPI_ISL_4606122, EPI_ISL_4606123, EPI_ISL_4606124, EPI_ISL_4606125, EPI_ISL_4606126, EPI_ISL_4606127 | | | |
| see above | BIOLITTORAL BIOGROUP PLATEAU TECH | CNR Virus des Infections Respiratoires - France SUD | Antonin Bal; Bruno Lina; Gregory Destras; Gwendolyne Burfin; Hadrien Regue; Laurence Josset; Martine Valette; Quentin Semanas |
| EPI_ISL_4606554, EPI_ISL_4606560, EPI_ISL_4606565, EPI_ISL_4606567, EPI_ISL_4606568, EPI_ISL_4606571 | BIOMER LABORATOIRE ST DIE | CNR Virus des Infections Respiratoires - France SUD | Antonin Bal; Bruno Lina; Gregory Destras; Gwendolyne Burfin; Hadrien Regue; Laurence Josset; Martine Valette; Quentin Semanas |
| EPI_ISL_4605936, EPI_ISL_4605937, EPI_ISL_4605938, EPI_ISL_4605939, EPI_ISL_4605941, EPI_ISL_4605942, EPI_ISL_4605943, EPI_ISL_4605944 | | | |
| see above | BIOPTIMA - VEZERONCE CURTIN | CNR Virus des Infections Respiratoires - France SUD | Antonin Bal; Bruno Lina; Gregory Destras; Gwendolyne Burfin; Hadrien Regue; Laurence Josset; Martine Valette; Quentin Semanas |
| EPI_ISL_4539760, EPI_ISL_4539761, EPI_ISL_4539762 | C.H. LE DAMANY LANNION-TRESTEL | CHU Pontchaillou | DE TAYRAC Marie; DENOUAL Florent; DUFOUR Marie Jose; ETCHEVERRY Amandine; FEBREAU Christine; GALIBERT Marie Dominique; GROLHIER Claire; JAGLINE Steven; PRONIER Charlotte; QUENET Benjamin; SASSI Mohamed; THIBAULT Vincent |
| EPI_ISL_4606362, EPI_ISL_4606363, EPI_ISL_4606366, EPI_ISL_4606368, EPI_ISL_4606369, EPI_ISL_4606371, EPI_ISL_4606373, EPI_ISL_4606374, EPI_ISL_4606977 | | | |
| see above | C.H.R.U. MONTPIED | CNR Virus des Infections Respiratoires - France SUD | Antonin Bal; Bruno Lina; Gregory Destras; Gwendolyne Burfin; Hadrien Regue; Laurence Josset; Martine Valette; Quentin Semanas |
| EPI_ISL_4606310, EPI_ISL_4606313, EPI_ISL_4606638 | CENTRE HOSPITALIER | CNR Virus des Infections Respiratoires - France SUD | Antonin Bal; Bruno Lina; Gregory Destras; Gwendolyne Burfin; Hadrien Regue; Laurence Josset; Martine Valette; Quentin Semanas |
| EPI_ISL_4606417, EPI_ISL_4606418, EPI_ISL_4606420 | CENTRE HOSPITALIER DE LA COTE BASQUE | CNR Virus des Infections Respiratoires - France SUD | Antonin Bal; Bruno Lina; Gregory Destras; Gwendolyne Burfin; Hadrien Regue; Laurence Josset; Martine Valette; Quentin Semanas |
| EPI_ISL_4605654, EPI_ISL_4606132, EPI_ISL_4606133, EPI_ISL_4606134, EPI_ISL_4606139, EPI_ISL_4606290, EPI_ISL_4606292, EPI_ISL_4606294, EPI_ISL_4606295, EPI_ISL_4606422, EPI_ISL_4606976 | | | |
| see above | CENTRE HOSPITALIER DE VALENCE | CNR Virus des Infections Respiratoires - France SUD | Antonin Bal; Bruno Lina; Gregory Destras; Gwendolyne Burfin; Hadrien Regue; Laurence Josset; Martine Valette; Quentin Semanas |
| EPI_ISL_4606321, EPI_ISL_4606323, EPI_ISL_4606324 | CENTRE HOSPITALIER DUPUYTREN | CNR Virus des Infections Respiratoires - France SUD | Antonin Bal; Bruno Lina; Gregory Destras; Gwendolyne Burfin; Hadrien Regue; Laurence Josset; Martine Valette; Quentin Semanas |
| EPI_ISL_4607114 | CENTRE HOSPITALIER EMILE ROUX | CNR Virus des Infections Respiratoires - France SUD | Antonin Bal; Bruno Lina; Gregory Destras; Gwendolyne Burfin; Hadrien Regue; Laurence Josset; Martine Valette; Quentin Semanas |
| EPI_ISL_4326321 | CENTRE HOSPITALIER LUCIEN HUSSEL | CNR Virus des Infections Respiratoires - France SUD | Antonin Bal; Bruno Lina; Gregory Destras; Gwendolyne Burfin; Hadrien Regue; Laurence Josset; Martine Valette; Quentin Semanas |
| EPI_ISL_4607077 | CENTRE HOSPITALIER NIORT | CNR Virus des Infections Respiratoires - France SUD | Antonin Bal; Bruno Lina; Gregory Destras; Gwendolyne Burfin; Hadrien Regue; Laurence Josset; Martine Valette; Quentin Semanas |
| EPI_ISL_4606347, EPI_ISL_4606351, EPI_ISL_4606353, EPI_ISL_4606354, EPI_ISL_4606356, EPI_ISL_4606358, EPI_ISL_4606360 | | | |
| see above | CENTRE HOSPITALIER PELLEGRIN | CNR Virus des Infections Respiratoires - France SUD | Antonin Bal; Bruno Lina; Gregory Destras; Gwendolyne Burfin; Hadrien Regue; Laurence Josset; Martine Valette; Quentin Semanas |
| EPI_ISL_4605716, EPI_ISL_4605978 | CENTRE HOSPITALIER PIERRE OUDOT | CNR Virus des Infections Respiratoires - France SUD | Antonin Bal; Bruno Lina; Gregory Destras; Gwendolyne Burfin; Hadrien Regue; Laurence Josset; Martine Valette; Quentin Semanas |
| EPI_ISL_4605940, EPI_ISL_4606054, EPI_ISL_4606063, EPI_ISL_4606066, EPI_ISL_4606072, EPI_ISL_4606074, EPI_ISL_4606076 | | | |
| see above | CENTRE HOSPITALIER POITIERS | CNR Virus des Infections Respiratoires - France SUD | Antonin Bal; Bruno Lina; Gregory Destras; Gwendolyne Burfin; Hadrien Regue; Laurence Josset; Martine Valette; Quentin Semanas |
| EPI_ISL_4326479, EPI_ISL_4326516, EPI_ISL_4605611, | CENTRE HOSPITALIER ST JOSEPH ST LUC | CNR Virus des Infections Respiratoires - France SUD | Antonin Bal; Bruno Lina; Gregory Destras; Gwendolyne Burfin; Hadrien Regue; Laurence Josset; Martine Valette; Quentin Semanas |

| | | | |
|---|---|---|---|
| EPI_ISL_4606130, EPI_ISL_4606131 | | | |
| EPI_ISL_4606316, EPI_ISL_4606318, EPI_ISL_4606319 | CENTRE HOSPITALIER VAL D'ARIEGE | CNR Virus des Infections Respiratoires - France SUD | Antonin Bal; Bruno Lina; Gregory Destras; Gwendolyne Burfin; Hadrien Regue; Laurence Josset; Martine Valette; Quentin Semanas |
| EPI_ISL_4605855, EPI_ISL_4605858, EPI_ISL_4605862, EPI_ISL_4605867, EPI_ISL_4605868, EPI_ISL_4605871, EPI_ISL_4605873 | | | |
| see above | CERBALLIANCE CHARENTES SAINTES | CNR Virus des Infections Respiratoires - France SUD | Antonin Bal; Bruno Lina; Gregory Destras; Gwendolyne Burfin; Hadrien Regue; Laurence Josset; Martine Valette; Quentin Semanas |
| EPI_ISL_4605409, EPI_ISL_4605418, EPI_ISL_4605419, EPI_ISL_4605427, EPI_ISL_4605435, EPI_ISL_4605442, EPI_ISL_4605451, EPI_ISL_4605454, EPI_ISL_4605457, EPI_ISL_4605458, EPI_ISL_4605459, EPI_ISL_4605461, EPI_ISL_4605465, EPI_ISL_4605467, EPI_ISL_4605468, EPI_ISL_4605480, EPI_ISL_4605484, EPI_ISL_4605492, EPI_ISL_4605500, EPI_ISL_4605509, EPI_ISL_4605516, EPI_ISL_4605520, EPI_ISL_4605523, EPI_ISL_4605524, EPI_ISL_4605529, EPI_ISL_4605531, EPI_ISL_4605537, EPI_ISL_4605540, EPI_ISL_4605550, EPI_ISL_4605557, EPI_ISL_4605561, EPI_ISL_4605568, EPI_ISL_4606739, EPI_ISL_4606740, EPI_ISL_4606741, EPI_ISL_4606743, EPI_ISL_4606748, EPI_ISL_4606772, EPI_ISL_4606773, EPI_ISL_4606776, EPI_ISL_4606778, EPI_ISL_4606785, EPI_ISL_4606816, EPI_ISL_4606817, EPI_ISL_4606818, EPI_ISL_4606819, EPI_ISL_4606822, EPI_ISL_4606823, EPI_ISL_4606824 | | | |
| see above | CERBALLIANCE OCCITANIE | CNR Virus des Infections Respiratoires - France SUD | Antonin Bal; Bruno Lina; Gregory Destras; Gwendolyne Burfin; Hadrien Regue; Laurence Josset; Martine Valette; Quentin Semanas |
| EPI_ISL_4606632 | CH ANTIBES - JUAN LES PINS | CNR Virus des Infections Respiratoires - France SUD | Antonin Bal; Bruno Lina; Gregory Destras; Gwendolyne Burfin; Hadrien Regue; Laurence Josset; Martine Valette; Quentin Semanas |
| EPI_ISL_4606005 | CH GIVORS | CNR Virus des Infections Respiratoires - France SUD | Antonin Bal; Bruno Lina; Gregory Destras; Gwendolyne Burfin; Hadrien Regue; Laurence Josset; Martine Valette; Quentin Semanas |
| EPI_ISL_4607020, EPI_ISL_4607031, EPI_ISL_4607035, EPI_ISL_4607043, EPI_ISL_4607045, EPI_ISL_4607075 | CHU GRENOBLE | CNR Virus des Infections Respiratoires - France SUD | Antonin Bal; Bruno Lina; Gregory Destras; Gwendolyne Burfin; Hadrien Regue; Laurence Josset; Martine Valette; Quentin Semanas |
| EPI_ISL_4548669, EPI_ISL_4548670, EPI_ISL_4548671, EPI_ISL_4548672, EPI_ISL_4548673, EPI_ISL_4558227, EPI_ISL_4558228, EPI_ISL_4558229, EPI_ISL_4558230, EPI_ISL_4558231, EPI_ISL_4558232, EPI_ISL_4558233, EPI_ISL_4558234 | | | |
| see above | CHU NIMES | CHU NIMES | Agathe Boudet; Marie-Josée Carles; Sophie Bravo; Stephan Robin |
| EPI_ISL_4261654, EPI_ISL_4539757, EPI_ISL_4539758, EPI_ISL_4539759 | CHU Pontchaillou | CHU Pontchaillou | DE TAYRAC Marie; DENOUAL Florent; ETCHEVERRY Amandine; FEBREAU Christine; GALIBERT Marie Dominique; GROLHIER Claire; JAGLINE Steven; PRONIER Charlotte; QUENET Benjamin; SASSI Mohamed; THIBAULT Vincent |
| EPI_ISL_4604994, EPI_ISL_4604996, EPI_ISL_4604997, EPI_ISL_4604998, EPI_ISL_4604999, EPI_ISL_4605006, EPI_ISL_4605008, EPI_ISL_4605194, EPI_ISL_4605206, EPI_ISL_4605210, EPI_ISL_4605213, EPI_ISL_4605217, EPI_ISL_4605233 | | | |
| see above | CHU ST ETIENNE HOPITAL NORD | CNR Virus des Infections Respiratoires - France SUD | Antonin Bal; Bruno Lina; Gregory Destras; Gwendolyne Burfin; Hadrien Regue; Laurence Josset; Martine Valette; Quentin Semanas |
| EPI_ISL_4606784 | CHU TOULOUSE | CNR Virus des Infections Respiratoires - France SUD | Antonin Bal; Bruno Lina; Gregory Destras; Gwendolyne Burfin; Hadrien Regue; Laurence Josset; Martine Valette; Quentin Semanas |
| EPI_ISL_4606820 | CLINIQUE ESQUIROL-ST HILAIRE | CNR Virus des Infections Respiratoires - France SUD | Antonin Bal; Bruno Lina; Gregory Destras; Gwendolyne Burfin; Hadrien Regue; Laurence Josset; Martine Valette; Quentin Semanas |
| EPI_ISL_4326128, EPI_ISL_4326129, EPI_ISL_4326131, EPI_ISL_4326306, EPI_ISL_4326318, EPI_ISL_4326335, EPI_ISL_4326341, EPI_ISL_4326352, EPI_ISL_4326470, EPI_ISL_4326473, EPI_ISL_4326476, EPI_ISL_4326482, EPI_ISL_4326499, EPI_ISL_4326629, EPI_ISL_4326643, EPI_ISL_4326646, EPI_ISL_4326649, EPI_ISL_4326655, EPI_ISL_4326658, EPI_ISL_4326677, EPI_ISL_4326680, EPI_ISL_4326683, EPI_ISL_4326685, EPI_ISL_4326737, EPI_ISL_4326740, EPI_ISL_4326744, EPI_ISL_4326747, EPI_ISL_4326750, EPI_ISL_4605037, EPI_ISL_4605577, EPI_ISL_4605578, EPI_ISL_4605676, EPI_ISL_4605802, EPI_ISL_4605955, EPI_ISL_4605956, EPI_ISL_4605957, EPI_ISL_4605958, EPI_ISL_4605959, EPI_ISL_4605962, EPI_ISL_4605970, EPI_ISL_4605977, EPI_ISL_4605994, EPI_ISL_4606001, EPI_ISL_4606002, EPI_ISL_4606003, EPI_ISL_4606004, EPI_ISL_4606006, EPI_ISL_4606007, EPI_ISL_4606008, EPI_ISL_4606009, EPI_ISL_4606010, EPI_ISL_4606014, EPI_ISL_4606015, EPI_ISL_4606064, EPI_ISL_4606112, EPI_ISL_4606161, EPI_ISL_4606181, EPI_ISL_4606212, EPI_ISL_4606217, EPI_ISL_4606289, EPI_ISL_4606411, EPI_ISL_4606416, EPI_ISL_4606439, EPI_ISL_4606444, EPI_ISL_4606445, EPI_ISL_4606505, EPI_ISL_4606646, EPI_ISL_4606653, EPI_ISL_4606935, EPI_ISL_4607142, EPI_ISL_4607143, EPI_ISL_4607144, EPI_ISL_4607145, EPI_ISL_4607146, EPI_ISL_4607147, EPI_ISL_4607148, EPI_ISL_4607149, EPI_ISL_4607150, EPI_ISL_4607151, EPI_ISL_4607152, EPI_ISL_4607153 | | | |
| see above | CNR Virus des Infections Respiratoires - France SUD | CNR Virus des Infections Respiratoires - France SUD | Antonin Bal; Bruno Lina; Gregory Destras; Gwendolyne Burfin; Hadrien Regue; Laurence Josset; Martine Valette; Quentin Semanas |
| EPI_ISL_4606129, EPI_ISL_4606141, EPI_ISL_4606143, EPI_ISL_4606144, EPI_ISL_4606145, EPI_ISL_4606146, EPI_ISL_4606147, EPI_ISL_4606148, EPI_ISL_4606149, EPI_ISL_4606150, EPI_ISL_4606151, EPI_ISL_4606152, EPI_ISL_4606153, EPI_ISL_4606155, EPI_ISL_4606156, EPI_ISL_4606157, EPI_ISL_4606158, EPI_ISL_4606159, EPI_ISL_4606160, EPI_ISL_4606162, EPI_ISL_4606601, EPI_ISL_4606603, EPI_ISL_4606607, EPI_ISL_4606609, EPI_ISL_4606612, EPI_ISL_4606614, EPI_ISL_4606617, EPI_ISL_4606619, EPI_ISL_4606634, EPI_ISL_4606636, EPI_ISL_4606637, EPI_ISL_4606639, EPI_ISL_4606640, EPI_ISL_4606642, EPI_ISL_4606643, EPI_ISL_4606644, EPI_ISL_4606645, EPI_ISL_4606651 | | | |
| see above | DYOMEDEA-LABORATOIRE DE LA SAUVEGARDE | CNR Virus des Infections Respiratoires - France SUD | Antonin Bal; Bruno Lina; Gregory Destras; Gwendolyne Burfin; Hadrien Regue; Laurence Josset; Martine Valette; Quentin Semanas |
| EPI_ISL_4606972 | EUROFINS LABAZUR RUMILLY | CNR Virus des Infections Respiratoires - France SUD | Antonin Bal; Bruno Lina; Gregory Destras; Gwendolyne Burfin; Hadrien Regue; Laurence Josset; Martine Valette; Quentin Semanas |
| EPI_ISL_4607088, EPI_ISL_4607090, EPI_ISL_4607092, EPI_ISL_4607095, EPI_ISL_4607101, EPI_ISL_4607102, EPI_ISL_4607103, EPI_ISL_4607104, EPI_ISL_4607107, EPI_ISL_4607108, EPI_ISL_4607110, EPI_ISL_4607113, EPI_ISL_4607115, EPI_ISL_4607116 | | | |
| see above | EXALAB LE HAILLAN | CNR Virus des Infections Respiratoires - France SUD | Antonin Bal; Bruno Lina; Gregory Destras; Gwendolyne Burfin; Hadrien Regue; Laurence Josset; Martine Valette; Quentin Semanas |
| EPI_ISL_4606379, EPI_ISL_4606380, EPI_ISL_4606382, EPI_ISL_4606384 | HOPITAL ALBI | CNR Virus des Infections Respiratoires - France SUD | Antonin Bal; Bruno Lina; Gregory Destras; Gwendolyne Burfin; Hadrien Regue; Laurence Josset; Martine Valette; Quentin Semanas |
| EPI_ISL_4606767, EPI_ISL_4606768, EPI_ISL_4606769, EPI_ISL_4606770, EPI_ISL_4606771 | HOPITAL DU PAYS D'AUTAN | CNR Virus des Infections Respiratoires - France SUD | Antonin Bal; Bruno Lina; Gregory Destras; Gwendolyne Burfin; Hadrien Regue; Laurence Josset; Martine Valette; Quentin Semanas |
| EPI_ISL_4606304, EPI_ISL_4606305 | HOPITAL NORD OUEST - CH VILLEFRANCHE | CNR Virus des Infections Respiratoires - France SUD | Antonin Bal; Bruno Lina; Gregory Destras; Gwendolyne Burfin; Hadrien Regue; Laurence Josset; Martine Valette; Quentin Semanas |
| EPI_ISL_4607016 | HOPITAL ROBERT BOULIN | CNR Virus des Infections Respiratoires - France SUD | Antonin Bal; Bruno Lina; Gregory Destras; Gwendolyne Burfin; Hadrien Regue; Laurence Josset; Martine Valette; Quentin Semanas |
| EPI_ISL_4606944 | HOPITAUX DROME NORD | CNR Virus des Infections Respiratoires - France SUD | Antonin Bal; Bruno Lina; Gregory Destras; Gwendolyne Burfin; Hadrien Regue; Laurence Josset; Martine Valette; Quentin Semanas |
| EPI_ISL_4461359, EPI_ISL_4461360, EPI_ISL_4461361, EPI_ISL_4461425, EPI_ISL_4461502, EPI_ISL_4461503, EPI_ISL_4461514, EPI_ISL_4461525, EPI_ISL_4461528, EPI_ISL_4461529, EPI_ISL_4461629, EPI_ISL_4461645, EPI_ISL_4461646, EPI_ISL_4461647, EPI_ISL_4461648, EPI_ISL_4461649, EPI_ISL_4461691, EPI_ISL_4461728, EPI_ISL_4461735, EPI_ISL_4461799, EPI_ISL_4503773, EPI_ISL_4503774, EPI_ISL_4503775, EPI_ISL_4503776, EPI_ISL_4503804, EPI_ISL_4503805, EPI_ISL_4503806, EPI_ISL_4503855, EPI_ISL_4503856, EPI_ISL_4503857, EPI_ISL_4503858, EPI_ISL_4503860, EPI_ISL_4503912, EPI_ISL_4503913, EPI_ISL_4503914, EPI_ISL_4503915, EPI_ISL_4503916, EPI_ISL_4503917, EPI_ISL_4503918, EPI_ISL_4503919, EPI_ISL_4503920, EPI_ISL_4545352, EPI_ISL_4545361, EPI_ISL_4545362, EPI_ISL_4545363, EPI_ISL_4545364, EPI_ISL_4545365, EPI_ISL_4545366, EPI_ISL_4600584, EPI_ISL_4600585, EPI_ISL_4600586, EPI_ISL_4600587, EPI_ISL_4600588, EPI_ISL_4600633, EPI_ISL_4600634, EPI_ISL_4600635, EPI_ISL_4600636, EPI_ISL_4600637, EPI_ISL_4600638, EPI_ISL_4600639, EPI_ISL_4600640, EPI_ISL_4600642, EPI_ISL_4600651, EPI_ISL_4600652, EPI_ISL_4600684, EPI_ISL_4600716, EPI_ISL_4600717, EPI_ISL_4600729, EPI_ISL_4600730, EPI_ISL_4600731, EPI_ISL_4600732, EPI_ISL_4600748, EPI_ISL_4600749, EPI_ISL_4600750, EPI_ISL_4600764, EPI_ISL_4600765, EPI_ISL_4600767, EPI_ISL_4600768, EPI_ISL_4600769, EPI_ISL_4600772, EPI_ISL_4600785, EPI_ISL_4600786, EPI_ISL_4600787, EPI_ISL_4600788, EPI_ISL_4600789, EPI_ISL_4600790, EPI_ISL_4600792, EPI_ISL_4600811, EPI_ISL_4600812, EPI_ISL_4600941, EPI_ISL_4600951, EPI_ISL_4600958, EPI_ISL_4600959, EPI_ISL_4600960, EPI_ISL_4600986, EPI_ISL_4601008, EPI_ISL_4601009, EPI_ISL_4601068, EPI_ISL_4601074, EPI_ISL_4601113 | | | |
| see above | Hospital | National Reference Center for Viruses of Respiratory Infections, Institut Pasteur, Paris | Alexandra Ducancelle; Angela Brisebarre; Anne Cady; Axelle Paquin; Benjamin Maneglie; Camille Capel; Christine Febreau; Christophe Malabat; CléMence Guillaume; Corinne Maufrais; Cyril Hoche; CéLine Bressollette; Elena Guillotel; Emeline Riverain; Eric Farfour; Etienne Simon-Lorière; Facon; Frédéric Lemoine; Ghu Paris Psychiatrie; Haciba Moudjahed; Isabelle Gros; Isabelle Joly; José-Manuel Ramirez; Julien Fumey; L. Courdavault; Laura Djamdjian; Louise Lefrançois; LéA Pilorge; Mama Imene Fodil Pacha; Marianne Burgard; Marie-Christine Legrand-Quillien; Marion Barbet; Martine Escure; Maud Vanpeene; Méline Bizard; Pierre Alain Billy; Pierre Patoz; Sandrine Castelain; Slim El Khiari; Slim El-Khiari; Sylvie Behillil; Sylvie Van der Werf; Thibault Guinoiseau; ValéRie Serazin; Vincent Enouf; Vincent Thibault; Zehaira Benseddick |
| EPI_ISL_4600774 | Hôpital | National Reference Center for Viruses of Respiratory Infections, Institut Pasteur, Paris | Angela Brisebarre; Camille Capel; Christophe Malabat; Corinne Maufrais; Etienne Simon-Lorière; Frédéric Lemoine; Julien Fumey; Louise Lefrançois; Marianne Burgard; Marion Barbet; Maud Vanpeene; Méline Bizard; Slim El Khiari; Sylvie Behillil; Sylvie Van der Werf; Vincent Enouf |
| EPI_ISL_4606019, | LABM CH | CNR Virus des Infections | Antonin Bal; Bruno Lina; Gregory Destras; Gwendolyne Burfin; Hadrien Regue; Laurence Josset; Martine Valette; Quentin Semanas |

| | | | |
|---|---|---|---|
| EPI_ISL_4606022, EPI_ISL_4606024 | MONTAUBAN | Respiratoires - France SUD | |
| EPI_ISL_4605908, EPI_ISL_4605912, EPI_ISL_4605915, EPI_ISL_4605917, EPI_ISL_4605918, EPI_ISL_4605919, EPI_ISL_4605920, EPI_ISL_4605921, EPI_ISL_4605922, EPI_ISL_4605923, EPI_ISL_4605927, EPI_ISL_4606933, EPI_ISL_4606936, EPI_ISL_4606940, EPI_ISL_4606941, EPI_ISL_4606942, EPI_ISL_4606943 | | | |
| see above | LABO BIOSEVRES BRESSUIRE | CNR Virus des Infections Respiratoires - France SUD | Antonin Bal; Bruno Lina; Gregory Destras; Gwendolyne Burfin; Hadrien Regue; Laurence Josset; Martine Valette; Quentin Semanas |
| EPI_ISL_4607010, EPI_ISL_4607013 | LABO BIOSEVRES THOUARS | CNR Virus des Infections Respiratoires - France SUD | Antonin Bal; Bruno Lina; Gregory Destras; Gwendolyne Burfin; Hadrien Regue; Laurence Josset; Martine Valette; Quentin Semanas |
| EPI_ISL_4606298, EPI_ISL_4606299, EPI_ISL_4606300, EPI_ISL_4606302 | LABO HOSPILAB 47 SITE D AGEN | CNR Virus des Infections Respiratoires - France SUD | Antonin Bal; Bruno Lina; Gregory Destras; Gwendolyne Burfin; Hadrien Regue; Laurence Josset; Martine Valette; Quentin Semanas |
| EPI_ISL_4606599, EPI_ISL_4606602, EPI_ISL_4606605, EPI_ISL_4606608, EPI_ISL_4606611, EPI_ISL_4606613, EPI_ISL_4606615, EPI_ISL_4606618, EPI_ISL_4606620, EPI_ISL_4606622, EPI_ISL_4606623, EPI_ISL_4606624, EPI_ISL_4606626 | | | |
| see above | LABORATOIRE ACCOLAB SUD OUEST | CNR Virus des Infections Respiratoires - France SUD | Antonin Bal; Bruno Lina; Gregory Destras; Gwendolyne Burfin; Hadrien Regue; Laurence Josset; Martine Valette; Quentin Semanas |
| EPI_ISL_4605251, EPI_ISL_4605253, EPI_ISL_4605259, EPI_ISL_4605268, EPI_ISL_4605271, EPI_ISL_4606831, EPI_ISL_4606833, EPI_ISL_4606834, EPI_ISL_4606835, EPI_ISL_4606836, EPI_ISL_4606838, EPI_ISL_4606839, EPI_ISL_4606847, EPI_ISL_4606849, EPI_ISL_4606851, EPI_ISL_4606852, EPI_ISL_4606853, EPI_ISL_4606866, EPI_ISL_4606875, EPI_ISL_4606894, EPI_ISL_4606900, EPI_ISL_4606909 | | | |
| see above | LABORATOIRE ANABIO BERGSON | CNR Virus des Infections Respiratoires - France SUD | Antonin Bal; Bruno Lina; Gregory Destras; Gwendolyne Burfin; Hadrien Regue; Laurence Josset; Martine Valette; Quentin Semanas |
| EPI_ISL_4607105, EPI_ISL_4607106, EPI_ISL_4607109, EPI_ISL_4607111, EPI_ISL_4607112, EPI_ISL_4607117, EPI_ISL_4607119 | | | |
| see above | LABORATOIRE BIO-VAL | CNR Virus des Infections Respiratoires - France SUD | Antonin Bal; Bruno Lina; Gregory Destras; Gwendolyne Burfin; Hadrien Regue; Laurence Josset; Martine Valette; Quentin Semanas |
| EPI_ISL_4605756, EPI_ISL_4605757, EPI_ISL_4605759, EPI_ISL_4605760, EPI_ISL_4605761, EPI_ISL_4605763, EPI_ISL_4605765, EPI_ISL_4605767, EPI_ISL_4605769, EPI_ISL_4605771, EPI_ISL_4605772, EPI_ISL_4605777, EPI_ISL_4605778, EPI_ISL_4605783, EPI_ISL_4605786, EPI_ISL_4605789, EPI_ISL_4605792, EPI_ISL_4605795, EPI_ISL_4605798, EPI_ISL_4605804, EPI_ISL_4605806, EPI_ISL_4605808, EPI_ISL_4605811, EPI_ISL_4605815, EPI_ISL_4605817, EPI_ISL_4605819, EPI_ISL_4605821, EPI_ISL_4605822, EPI_ISL_4605824, EPI_ISL_4605826, EPI_ISL_4605829, EPI_ISL_4605847, EPI_ISL_4605852 | | | |
| see above | LABORATOIRE BIOESTEREL | CNR Virus des Infections Respiratoires - France SUD | Antonin Bal; Bruno Lina; Gregory Destras; Gwendolyne Burfin; Hadrien Regue; Laurence Josset; Martine Valette; Quentin Semanas |
| EPI_ISL_4606394, EPI_ISL_4606396, EPI_ISL_4606398, EPI_ISL_4606401, EPI_ISL_4606402, EPI_ISL_4606404, EPI_ISL_4606406, EPI_ISL_4606408, EPI_ISL_4606409, EPI_ISL_4606410, EPI_ISL_4606413, EPI_ISL_4606415 | | | |
| see above | LABORATOIRE BIOFUSION | CNR Virus des Infections Respiratoires - France SUD | Antonin Bal; Bruno Lina; Gregory Destras; Gwendolyne Burfin; Hadrien Regue; Laurence Josset; Martine Valette; Quentin Semanas |
| EPI_ISL_4606535, EPI_ISL_4606536, EPI_ISL_4606537, EPI_ISL_4606538, EPI_ISL_4606539, EPI_ISL_4606547, EPI_ISL_4606550, EPI_ISL_4606551, EPI_ISL_4606552, EPI_ISL_4606555, EPI_ISL_4606557 | | | |
| see above | LABORATOIRE BIOLAB AVENIR | CNR Virus des Infections Respiratoires - France SUD | Antonin Bal; Bruno Lina; Gregory Destras; Gwendolyne Burfin; Hadrien Regue; Laurence Josset; Martine Valette; Quentin Semanas |
| EPI_ISL_4606742, EPI_ISL_4606744, EPI_ISL_4606745, EPI_ISL_4606746, EPI_ISL_4606747, EPI_ISL_4606749, EPI_ISL_4606750, EPI_ISL_4606751 | | | |
| see above | LABORATOIRE BIOLYSS | CNR Virus des Infections Respiratoires - France SUD | Antonin Bal; Bruno Lina; Gregory Destras; Gwendolyne Burfin; Hadrien Regue; Laurence Josset; Martine Valette; Quentin Semanas |
| EPI_ISL_4607137 | LABORATOIRE BIOMEDICA | CNR Virus des Infections Respiratoires - France SUD | Antonin Bal; Bruno Lina; Gregory Destras; Gwendolyne Burfin; Hadrien Regue; Laurence Josset; Martine Valette; Quentin Semanas |
| EPI_ISL_4606970, EPI_ISL_4606973 | LABORATOIRE BIOPTEAM - BOURG EN BRESSE (LALANDE) | CNR Virus des Infections Respiratoires - France SUD | Antonin Bal; Bruno Lina; Gregory Destras; Gwendolyne Burfin; Hadrien Regue; Laurence Josset; Martine Valette; Quentin Semanas |
| EPI_ISL_4606342 | LABORATOIRE BOUVIER | CNR Virus des Infections Respiratoires - France SUD | Antonin Bal; Bruno Lina; Gregory Destras; Gwendolyne Burfin; Hadrien Regue; Laurence Josset; Martine Valette; Quentin Semanas |
| EPI_ISL_4605846, EPI_ISL_4605850, EPI_ISL_4605854, EPI_ISL_4605857, EPI_ISL_4605860, EPI_ISL_4605863, EPI_ISL_4605865, EPI_ISL_4605869 | | | |
| see above | LABORATOIRE CANARELLI-FERNANDEZ | CNR Virus des Infections Respiratoires - France SUD | Antonin Bal; Bruno Lina; Gregory Destras; Gwendolyne Burfin; Hadrien Regue; Laurence Josset; Martine Valette; Quentin Semanas |
| EPI_ISL_4606081, EPI_ISL_4606082, EPI_ISL_4606083, EPI_ISL_4606084, EPI_ISL_4606085, EPI_ISL_4606086, EPI_ISL_4606087, EPI_ISL_4606088, EPI_ISL_4606089, EPI_ISL_4606090, EPI_ISL_4606091, EPI_ISL_4606092, EPI_ISL_4606093, EPI_ISL_4606094, EPI_ISL_4606095, EPI_ISL_4606096, EPI_ISL_4606097, EPI_ISL_4606098, EPI_ISL_4606099, EPI_ISL_4606100, EPI_ISL_4606101, EPI_ISL_4606102, EPI_ISL_4606103 | | | |
| see above | LABORATOIRE CBM DE MURET | CNR Virus des Infections Respiratoires - France SUD | Antonin Bal; Bruno Lina; Gregory Destras; Gwendolyne Burfin; Hadrien Regue; Laurence Josset; Martine Valette; Quentin Semanas |
| EPI_ISL_4605762, EPI_ISL_4605766, EPI_ISL_4605773, EPI_ISL_4605774, EPI_ISL_4605776, EPI_ISL_4605779, EPI_ISL_4605780, EPI_ISL_4605782, EPI_ISL_4605784, EPI_ISL_4605787, EPI_ISL_4605788, EPI_ISL_4605791, EPI_ISL_4605794, EPI_ISL_4605796, EPI_ISL_4605797, EPI_ISL_4605799, EPI_ISL_4605801, EPI_ISL_4605809, EPI_ISL_4605812, EPI_ISL_4605814, EPI_ISL_4605818, EPI_ISL_4605820, EPI_ISL_4605827, EPI_ISL_4605830, EPI_ISL_4605832, EPI_ISL_4605833, EPI_ISL_4605834, EPI_ISL_4605836, EPI_ISL_4605839, EPI_ISL_4605840, EPI_ISL_4605841, EPI_ISL_4605842, EPI_ISL_4605843, EPI_ISL_4605844, EPI_ISL_4605845, EPI_ISL_4605848, EPI_ISL_4605849, EPI_ISL_4605851, EPI_ISL_4605853, EPI_ISL_4605856, EPI_ISL_4605859, EPI_ISL_4605861, EPI_ISL_4605864, EPI_ISL_4605866, EPI_ISL_4605870, EPI_ISL_4605872, EPI_ISL_4605874, EPI_ISL_4605876, EPI_ISL_4605877, EPI_ISL_4605878, EPI_ISL_4605880, EPI_ISL_4605882, EPI_ISL_4605883, EPI_ISL_4605884, EPI_ISL_4605886, EPI_ISL_4605887, EPI_ISL_4605888, EPI_ISL_4605889, EPI_ISL_4606170, EPI_ISL_4606328, EPI_ISL_4606330, EPI_ISL_4606332, EPI_ISL_4606335, EPI_ISL_4606336, EPI_ISL_4606338, EPI_ISL_4606399, EPI_ISL_4606423, EPI_ISL_4606424, EPI_ISL_4606426, EPI_ISL_4606428, EPI_ISL_4606429, EPI_ISL_4606430, EPI_ISL_4606431, EPI_ISL_4606432, EPI_ISL_4606433, EPI_ISL_4606434, EPI_ISL_4606435, EPI_ISL_4606436, EPI_ISL_4606437, EPI_ISL_4606438, EPI_ISL_4606633, EPI_ISL_4606654, EPI_ISL_4606655, EPI_ISL_4606656, EPI_ISL_4606657, EPI_ISL_4606658, EPI_ISL_4606660, EPI_ISL_4606661, EPI_ISL_4606662, EPI_ISL_4606663, EPI_ISL_4606665, EPI_ISL_4606667, EPI_ISL_4606668, EPI_ISL_4606669, EPI_ISL_4606671, EPI_ISL_4606672, EPI_ISL_4606674, EPI_ISL_4606675, EPI_ISL_4606676, EPI_ISL_4606678, EPI_ISL_4606680, EPI_ISL_4606681, EPI_ISL_4606682, EPI_ISL_4606684, EPI_ISL_4606685, EPI_ISL_4606686, EPI_ISL_4606687, EPI_ISL_4606688, EPI_ISL_4606689, EPI_ISL_4606690, EPI_ISL_4606691, EPI_ISL_4606692, EPI_ISL_4606693, EPI_ISL_4606694, EPI_ISL_4606695, EPI_ISL_4606696, EPI_ISL_4606697, EPI_ISL_4606698, EPI_ISL_4606699, EPI_ISL_4606700, EPI_ISL_4606701, EPI_ISL_4606702, EPI_ISL_4606703, EPI_ISL_4606704, EPI_ISL_4606705, EPI_ISL_4606706, EPI_ISL_4606708, EPI_ISL_4606709, EPI_ISL_4606710, EPI_ISL_4606711 | | | |
| see above | LABORATOIRE CERBALLIANCE PLT VILLON | CNR Virus des Infections Respiratoires - France SUD | Antonin Bal; Bruno Lina; Gregory Destras; Gwendolyne Burfin; Hadrien Regue; Laurence Josset; Martine Valette; Quentin Semanas |
| EPI_ISL_4607085, EPI_ISL_4607086, EPI_ISL_4607089, EPI_ISL_4607091, EPI_ISL_4607094, EPI_ISL_4607096, EPI_ISL_4607097, EPI_ISL_4607098, EPI_ISL_4607099, EPI_ISL_4607100 | | | |
| see above | LABORATOIRE CREAVALLEE | CNR Virus des Infections Respiratoires - France SUD | Antonin Bal; Bruno Lina; Gregory Destras; Gwendolyne Burfin; Hadrien Regue; Laurence Josset; Martine Valette; Quentin Semanas |
| EPI_ISL_4606752, EPI_ISL_4606753, EPI_ISL_4606756, EPI_ISL_4606757, EPI_ISL_4606758, EPI_ISL_4606759, EPI_ISL_4606761, EPI_ISL_4606762, EPI_ISL_4606764 | | | |
| see above | LABORATOIRE FORTE BIO DAX | CNR Virus des Infections Respiratoires - France SUD | Antonin Bal; Bruno Lina; Gregory Destras; Gwendolyne Burfin; Hadrien Regue; Laurence Josset; Martine Valette; Quentin Semanas |
| EPI_ISL_4606990, EPI_ISL_4606991, EPI_ISL_4606993, EPI_ISL_4606994, EPI_ISL_4606996, EPI_ISL_4606997, EPI_ISL_4606998, EPI_ISL_4606999, EPI_ISL_4607000, EPI_ISL_4607002, EPI_ISL_4607003, EPI_ISL_4607005, EPI_ISL_4607006, EPI_ISL_4607007, EPI_ISL_4607009, EPI_ISL_4607011, EPI_ISL_4607014, EPI_ISL_4607017, EPI_ISL_4607018 | | | |
| see above | LABORATOIRE LABAZUR | CNR Virus des Infections Respiratoires - France SUD | Antonin Bal; Bruno Lina; Gregory Destras; Gwendolyne Burfin; Hadrien Regue; Laurence Josset; Martine Valette; Quentin Semanas |
| EPI_ISL_4326397, EPI_ISL_4606051, EPI_ISL_4606052, EPI_ISL_4606053, EPI_ISL_4606055, EPI_ISL_4606056, EPI_ISL_4606057, EPI_ISL_4606058, EPI_ISL_4606059, EPI_ISL_4606060, EPI_ISL_4606061, EPI_ISL_4606062, EPI_ISL_4606065, EPI_ISL_4606067, EPI_ISL_4606069, EPI_ISL_4606071, EPI_ISL_4606073, EPI_ISL_4606075, EPI_ISL_4606077, EPI_ISL_4606079 | | | |
| see above | LABORATOIRE LBA JAYAN AGEN | CNR Virus des Infections Respiratoires - France SUD | Antonin Bal; Bruno Lina; Gregory Destras; Gwendolyne Burfin; Hadrien Regue; Laurence Josset; Martine Valette; Quentin Semanas |
| EPI_ISL_4605875, EPI_ISL_4606026, EPI_ISL_4606028, EPI_ISL_4606030, EPI_ISL_4606031, EPI_ISL_4606033, EPI_ISL_4606035, EPI_ISL_4606037, EPI_ISL_4606038, EPI_ISL_4606040, EPI_ISL_4606042, EPI_ISL_4606043, EPI_ISL_4606045 | | | |
| see above | LABORATOIRE MAYMAT | CNR Virus des Infections Respiratoires - France SUD | Antonin Bal; Bruno Lina; Gregory Destras; Gwendolyne Burfin; Hadrien Regue; Laurence Josset; Martine Valette; Quentin Semanas |
| EPI_ISL_4606213, EPI_ISL_4606214, EPI_ISL_4606216, EPI_ISL_4606219, EPI_ISL_4606221, EPI_ISL_4606223, EPI_ISL_4606225, EPI_ISL_4606227, EPI_ISL_4606229, EPI_ISL_4606231, EPI_ISL_4606233, EPI_ISL_4606236, EPI_ISL_4606238, EPI_ISL_4606240, EPI_ISL_4606241, EPI_ISL_4606242, EPI_ISL_4606243, EPI_ISL_4606245, EPI_ISL_4606247, EPI_ISL_4606248, EPI_ISL_4606250, EPI_ISL_4606251, EPI_ISL_4606253 | | | |

| | | | |
|---|---|---|---|
| see above | LABORATOIRE NOVELAB | CNR Virus des Infections Respiratoires - France SUD | Antonin Bal; Bruno Lina; Gregory Destras; Gwendolyne Burfin; Hadrien Regue; Laurence Josset; Martine Valette; Quentin Semanas |
| EPI_ISL_4605616, EPI_ISL_4605618, EPI_ISL_4605619, EPI_ISL_4605620, EPI_ISL_4605622, EPI_ISL_4605624, EPI_ISL_4605625, EPI_ISL_4605626, EPI_ISL_4605628, EPI_ISL_4605629, EPI_ISL_4605630, EPI_ISL_4605632, EPI_ISL_4605633, EPI_ISL_4605634, EPI_ISL_4605636, EPI_ISL_4605637, EPI_ISL_4605638, EPI_ISL_4605639, EPI_ISL_4605640, EPI_ISL_4605642, EPI_ISL_4605643, EPI_ISL_4605645, EPI_ISL_4605646, EPI_ISL_4605648, EPI_ISL_4605649, EPI_ISL_4605650, EPI_ISL_4605651, EPI_ISL_4605652, EPI_ISL_4605655, EPI_ISL_4605656, EPI_ISL_4605657, EPI_ISL_4605658, EPI_ISL_4605659, EPI_ISL_4605660, EPI_ISL_4605661, EPI_ISL_4605662, EPI_ISL_4605663, EPI_ISL_4605664, EPI_ISL_4605665, EPI_ISL_4605666, EPI_ISL_4605667, EPI_ISL_4605668, EPI_ISL_4605669, EPI_ISL_4605670, EPI_ISL_4605671, EPI_ISL_4605672, EPI_ISL_4605673, EPI_ISL_4605674, EPI_ISL_4605675, EPI_ISL_4605678, EPI_ISL_4605683, EPI_ISL_4605684, EPI_ISL_4605685, EPI_ISL_4605691, EPI_ISL_4605692, EPI_ISL_4605693, EPI_ISL_4605698, EPI_ISL_4605705, EPI_ISL_4605714, EPI_ISL_4605715, EPI_ISL_4605717, EPI_ISL_4605718, EPI_ISL_4605719, EPI_ISL_4605720, EPI_ISL_4605721, EPI_ISL_4605722, EPI_ISL_4605723, EPI_ISL_4605724, EPI_ISL_4605725, EPI_ISL_4605726, EPI_ISL_4605727, EPI_ISL_4605728, EPI_ISL_4605729, EPI_ISL_4605734, EPI_ISL_4605735, EPI_ISL_4605737, EPI_ISL_4605738, EPI_ISL_4605740, EPI_ISL_4605741, EPI_ISL_4605742, EPI_ISL_4605744, EPI_ISL_4605746, EPI_ISL_4605748 | | | |
| see above | LABORATOIRE UNILIANS DECINES | CNR Virus des Infections Respiratoires - France SUD | Antonin Bal; Bruno Lina; Gregory Destras; Gwendolyne Burfin; Hadrien Regue; Laurence Josset; Martine Valette; Quentin Semanas |
| EPI_ISL_4326434, EPI_ISL_4607118, EPI_ISL_4607120, EPI_ISL_4607121, EPI_ISL_4607126, EPI_ISL_4607127, EPI_ISL_4607128, EPI_ISL_4607129, EPI_ISL_4607130, EPI_ISL_4607131, EPI_ISL_4607132, EPI_ISL_4607133, EPI_ISL_4607134, EPI_ISL_4607135, EPI_ISL_4607136 | | | |
| see above | LAM BIO 86 SITE DE CHAUMONT | CNR Virus des Infections Respiratoires - France SUD | Antonin Bal; Bruno Lina; Gregory Destras; Gwendolyne Burfin; Hadrien Regue; Laurence Josset; Martine Valette; Quentin Semanas |
| EPI_ISL_4606628 | LAM CERBA | CNR Virus des Infections Respiratoires - France SUD | Antonin Bal; Bruno Lina; Gregory Destras; Gwendolyne Burfin; Hadrien Regue; Laurence Josset; Martine Valette; Quentin Semanas |
| EPI_ISL_4607138, EPI_ISL_4607139, EPI_ISL_4607140, EPI_ISL_4607141 | LAM EIMER LENYS | CNR Virus des Infections Respiratoires - France SUD | Antonin Bal; Bruno Lina; Gregory Destras; Gwendolyne Burfin; Hadrien Regue; Laurence Josset; Martine Valette; Quentin Semanas |
| EPI_ISL_4606540, EPI_ISL_4606541, EPI_ISL_4606542, EPI_ISL_4606543, EPI_ISL_4606544, EPI_ISL_4606545, EPI_ISL_4606546, EPI_ISL_4606548, EPI_ISL_4606549, EPI_ISL_4606553, EPI_ISL_4606556, EPI_ISL_4606561, EPI_ISL_4606564, EPI_ISL_4606569, EPI_ISL_4606573, EPI_ISL_4606576, EPI_ISL_4606580, EPI_ISL_4606582, EPI_ISL_4606586, EPI_ISL_4606589, EPI_ISL_4606594, EPI_ISL_4606600, EPI_ISL_4606604, EPI_ISL_4606610, EPI_ISL_4606616, EPI_ISL_4606621, EPI_ISL_4606625, EPI_ISL_4606627, EPI_ISL_4606629, EPI_ISL_4606630, EPI_ISL_4606631 | | | |
| see above | LAM GEN-BIO GRAVANCHES | CNR Virus des Infections Respiratoires - France SUD | Antonin Bal; Bruno Lina; Gregory Destras; Gwendolyne Burfin; Hadrien Regue; Laurence Josset; Martine Valette; Quentin Semanas |
| EPI_ISL_4606046, EPI_ISL_4606048, EPI_ISL_4606049, EPI_ISL_4606050 | LAM SELARL 2A-2B | CNR Virus des Infections Respiratoires - France SUD | Antonin Bal; Bruno Lina; Gregory Destras; Gwendolyne Burfin; Hadrien Regue; Laurence Josset; Martine Valette; Quentin Semanas |
| EPI_ISL_4605891, EPI_ISL_4605892, EPI_ISL_4605893, EPI_ISL_4605894, EPI_ISL_4605895, EPI_ISL_4605897, EPI_ISL_4605900, EPI_ISL_4605902, EPI_ISL_4605903 | | | |
| see above | LAM SYNLAB BORDEAUX ATLANTIQUE | CNR Virus des Infections Respiratoires - France SUD | Antonin Bal; Bruno Lina; Gregory Destras; Gwendolyne Burfin; Hadrien Regue; Laurence Josset; Martine Valette; Quentin Semanas |
| EPI_ISL_4606981 | LBIA SAVOIE KANTYSBIO | CNR Virus des Infections Respiratoires - France SUD | Antonin Bal; Bruno Lina; Gregory Destras; Gwendolyne Burfin; Hadrien Regue; Laurence Josset; Martine Valette; Quentin Semanas |
| EPI_ISL_4606513, EPI_ISL_4606523, EPI_ISL_4606525, EPI_ISL_4606531 | LBM DYNABIO CARREAU | CNR Virus des Infections Respiratoires - France SUD | Antonin Bal; Bruno Lina; Gregory Destras; Gwendolyne Burfin; Hadrien Regue; Laurence Josset; Martine Valette; Quentin Semanas |
| EPI_ISL_4607076, EPI_ISL_4607078, EPI_ISL_4607079, EPI_ISL_4607080, EPI_ISL_4607082, EPI_ISL_4607083, EPI_ISL_4607084, EPI_ISL_4607087 | | | |
| see above | LBM SEALAB DARRASSE ET ASSOCIES | CNR Virus des Infections Respiratoires - France SUD | Antonin Bal; Bruno Lina; Gregory Destras; Gwendolyne Burfin; Hadrien Regue; Laurence Josset; Martine Valette; Quentin Semanas |
| EPI_ISL_4606506, EPI_ISL_4606507, EPI_ISL_4606509, EPI_ISL_4606510, EPI_ISL_4606512, EPI_ISL_4606517, EPI_ISL_4606518, EPI_ISL_4606520, EPI_ISL_4606521, EPI_ISL_4606522, EPI_ISL_4606524, EPI_ISL_4606527, EPI_ISL_4606528, EPI_ISL_4606529, EPI_ISL_4606530, EPI_ISL_4606532, EPI_ISL_4606533, EPI_ISL_4606534 | | | |
| see above | LBM TRONQUIERES | CNR Virus des Infections Respiratoires - France SUD | Antonin Bal; Bruno Lina; Gregory Destras; Gwendolyne Burfin; Hadrien Regue; Laurence Josset; Martine Valette; Quentin Semanas |
| EPI_ISL_4326327, EPI_ISL_4604288, EPI_ISL_4605896, EPI_ISL_4605898, EPI_ISL_4605901, EPI_ISL_4605904, EPI_ISL_4605905, EPI_ISL_4605906, EPI_ISL_4605907, EPI_ISL_4605909, EPI_ISL_4605910, EPI_ISL_4605911, EPI_ISL_4605913, EPI_ISL_4605914, EPI_ISL_4605916 | | | |
| see above | LX BIO | CNR Virus des Infections Respiratoires - France SUD | Antonin Bal; Bruno Lina; Gregory Destras; Gwendolyne Burfin; Hadrien Regue; Laurence Josset; Martine Valette; Quentin Semanas |
| EPI_ISL_4461362, EPI_ISL_4461363, EPI_ISL_4461364, EPI_ISL_4461365, EPI_ISL_4461366, EPI_ISL_4461367, EPI_ISL_4461368, EPI_ISL_4461369, EPI_ISL_4461370, EPI_ISL_4461371, EPI_ISL_4461372, EPI_ISL_4461373, EPI_ISL_4461374, EPI_ISL_4461439, EPI_ISL_4461440, EPI_ISL_4461441, EPI_ISL_4461442, EPI_ISL_4461443, EPI_ISL_4461444, EPI_ISL_4461445, EPI_ISL_4461446, EPI_ISL_4461447, EPI_ISL_4461448, EPI_ISL_4461449, EPI_ISL_4461450, EPI_ISL_4461451, EPI_ISL_4461452, EPI_ISL_4461453, EPI_ISL_4461454, EPI_ISL_4461455, EPI_ISL_4461456, EPI_ISL_4461457, EPI_ISL_4461458, EPI_ISL_4461459, EPI_ISL_4461460, EPI_ISL_4461461, EPI_ISL_4461468, EPI_ISL_4461469, EPI_ISL_4461470, EPI_ISL_4461471, EPI_ISL_4461472, EPI_ISL_4461473, EPI_ISL_4461475, EPI_ISL_4461476, EPI_ISL_4461487, EPI_ISL_4461488, EPI_ISL_4461489, EPI_ISL_4461490, EPI_ISL_4461491, EPI_ISL_4461492, EPI_ISL_4461493, EPI_ISL_4461494, EPI_ISL_4461495, EPI_ISL_4461496, EPI_ISL_4461497, EPI_ISL_4461498, EPI_ISL_4461499, EPI_ISL_4461500, EPI_ISL_4461501, EPI_ISL_4461566, EPI_ISL_4461567, EPI_ISL_4461568, EPI_ISL_4461569, EPI_ISL_4461570, EPI_ISL_4461571, EPI_ISL_4461572, EPI_ISL_4461573, EPI_ISL_4461574, EPI_ISL_4461575, EPI_ISL_4461576, EPI_ISL_4461577, EPI_ISL_4461578, EPI_ISL_4461579, EPI_ISL_4461580, EPI_ISL_4461581, EPI_ISL_4461582, EPI_ISL_4461583, EPI_ISL_4461584, EPI_ISL_4461585, EPI_ISL_4461586, EPI_ISL_4461587, EPI_ISL_4461588, EPI_ISL_4461589, EPI_ISL_4461590, EPI_ISL_4461591, EPI_ISL_4461592, EPI_ISL_4461624, EPI_ISL_4461625, EPI_ISL_4461635, EPI_ISL_4461636, EPI_ISL_4461637, EPI_ISL_4461638, EPI_ISL_4461639, EPI_ISL_4461640, EPI_ISL_4461641, EPI_ISL_4461642, EPI_ISL_4461643, EPI_ISL_4461644, EPI_ISL_4461651, EPI_ISL_4461652, EPI_ISL_4461653, EPI_ISL_4461654, EPI_ISL_4461655, EPI_ISL_4461656, EPI_ISL_4461657, EPI_ISL_4461658, EPI_ISL_4461659, EPI_ISL_4461660, EPI_ISL_4461661, EPI_ISL_4461688, EPI_ISL_4461692, EPI_ISL_4461699, EPI_ISL_4461700, EPI_ISL_4461702, EPI_ISL_4461704, EPI_ISL_4461706, EPI_ISL_4461707, EPI_ISL_4461709, EPI_ISL_4461711, EPI_ISL_4461712, EPI_ISL_4461717, EPI_ISL_4461719, EPI_ISL_4461720, EPI_ISL_4461725, EPI_ISL_4461726, EPI_ISL_4461727, EPI_ISL_4461731, EPI_ISL_4461732, EPI_ISL_4461733, EPI_ISL_4461736, EPI_ISL_4461781, EPI_ISL_4461782, EPI_ISL_4461797, EPI_ISL_4461801, EPI_ISL_4461803, EPI_ISL_4461806, EPI_ISL_4503706, EPI_ISL_4503740, EPI_ISL_4503741, EPI_ISL_4503742, EPI_ISL_4503743, EPI_ISL_4503744, EPI_ISL_4503745, EPI_ISL_4503746, EPI_ISL_4503747, EPI_ISL_4503748, EPI_ISL_4503749, EPI_ISL_4503750, EPI_ISL_4503751, EPI_ISL_4503752, EPI_ISL_4503753, EPI_ISL_4503754, EPI_ISL_4503755, EPI_ISL_4503756, EPI_ISL_4503757, EPI_ISL_4503758, EPI_ISL_4503759, EPI_ISL_4503760, EPI_ISL_4503761, EPI_ISL_4503762, EPI_ISL_4503763, EPI_ISL_4503764, EPI_ISL_4503765, EPI_ISL_4503766, EPI_ISL_4503778, EPI_ISL_4503779, EPI_ISL_4503780, EPI_ISL_4503781, EPI_ISL_4503782, EPI_ISL_4503783, EPI_ISL_4503784, EPI_ISL_4503785, EPI_ISL_4503786, EPI_ISL_4503787, EPI_ISL_4503788, EPI_ISL_4503790, EPI_ISL_4503791, EPI_ISL_4503792, EPI_ISL_4503793, EPI_ISL_4503794, EPI_ISL_4503795, EPI_ISL_4503796, EPI_ISL_4503797, EPI_ISL_4503798, EPI_ISL_4503799, EPI_ISL_4503800, EPI_ISL_4503801, EPI_ISL_4503802, EPI_ISL_4503803, EPI_ISL_4503808, EPI_ISL_4503809, EPI_ISL_4503810, EPI_ISL_4503811, EPI_ISL_4503812, EPI_ISL_4503813, EPI_ISL_4503814, EPI_ISL_4503817, EPI_ISL_4503818, EPI_ISL_4503819, EPI_ISL_4503820, EPI_ISL_4503822, EPI_ISL_4503823, EPI_ISL_4503824, EPI_ISL_4503825, EPI_ISL_4503826, EPI_ISL_4503827, EPI_ISL_4503828, EPI_ISL_4503829, EPI_ISL_4503830, EPI_ISL_4503831, EPI_ISL_4503832, EPI_ISL_4503833, EPI_ISL_4503834, EPI_ISL_4503835, EPI_ISL_4503836, EPI_ISL_4503837, EPI_ISL_4503838, EPI_ISL_4503839, EPI_ISL_4503840, EPI_ISL_4503841, EPI_ISL_4503842, EPI_ISL_4503843, EPI_ISL_4503844, EPI_ISL_4503845, EPI_ISL_4503846, EPI_ISL_4503847, EPI_ISL_4503848, EPI_ISL_4503849, EPI_ISL_4503850, EPI_ISL_4503851, EPI_ISL_4503852, EPI_ISL_4503853, EPI_ISL_4503854, EPI_ISL_4503861, EPI_ISL_4503862, EPI_ISL_4503863, EPI_ISL_4503864, EPI_ISL_4503865, EPI_ISL_4503866, EPI_ISL_4503867, EPI_ISL_4503868, EPI_ISL_4503869, EPI_ISL_4503871, EPI_ISL_4503872, EPI_ISL_4503873, EPI_ISL_4503874, EPI_ISL_4503875, EPI_ISL_4503876, EPI_ISL_4503877, EPI_ISL_4503878, EPI_ISL_4503879, EPI_ISL_4503880, EPI_ISL_4503881, EPI_ISL_4503882, EPI_ISL_4503883, EPI_ISL_4503884, EPI_ISL_4503885, EPI_ISL_4503886, EPI_ISL_4503887, EPI_ISL_4503888, EPI_ISL_4503889, EPI_ISL_4503890, EPI_ISL_4503891, EPI_ISL_4503892, EPI_ISL_4503893, EPI_ISL_4503894, EPI_ISL_4503895, EPI_ISL_4503896, EPI_ISL_4503897, EPI_ISL_4503898, EPI_ISL_4503899, EPI_ISL_4503900, EPI_ISL_4503901, EPI_ISL_4503902, EPI_ISL_4503903, EPI_ISL_4503904, EPI_ISL_4503905, EPI_ISL_4503906, EPI_ISL_4503907, EPI_ISL_4503927, EPI_ISL_4503928, EPI_ISL_4503929, EPI_ISL_4503930, EPI_ISL_4503931, EPI_ISL_4503932, EPI_ISL_4503933, EPI_ISL_4503934, EPI_ISL_4503935, EPI_ISL_4503936, EPI_ISL_4503937, EPI_ISL_4503938, EPI_ISL_4503939, EPI_ISL_4503940, EPI_ISL_4503941, EPI_ISL_4503942, EPI_ISL_4503943, EPI_ISL_4503944, EPI_ISL_4503945, EPI_ISL_4503946, EPI_ISL_4503947, EPI_ISL_4503948, EPI_ISL_4503949, EPI_ISL_4503950, EPI_ISL_4503951, EPI_ISL_4503952, EPI_ISL_4503953, EPI_ISL_4503954, EPI_ISL_4503955, EPI_ISL_4503956, EPI_ISL_4545317, EPI_ISL_4545318, EPI_ISL_4545332, EPI_ISL_4545333, EPI_ISL_4545334, EPI_ISL_4545335, EPI_ISL_4545336, EPI_ISL_4545337, EPI_ISL_4545338, EPI_ISL_4545339, EPI_ISL_4545340, EPI_ISL_4545341, EPI_ISL_4545342, EPI_ISL_4545343, EPI_ISL_4545344, EPI_ISL_4545345, EPI_ISL_4545346, EPI_ISL_4545347, EPI_ISL_4545348, EPI_ISL_4545349, EPI_ISL_4545350, EPI_ISL_4545351, EPI_ISL_4545353, EPI_ISL_4545354, EPI_ISL_4545355, EPI_ISL_4545356, EPI_ISL_4545357, EPI_ISL_4545358, EPI_ISL_4545371, EPI_ISL_4545372, EPI_ISL_4545373, EPI_ISL_4545374, EPI_ISL_4600583, EPI_ISL_4600589, EPI_ISL_4600590, EPI_ISL_4600592, EPI_ISL_4600593, EPI_ISL_4600594, EPI_ISL_4600595, EPI_ISL_4600596, EPI_ISL_4600597, EPI_ISL_4600598, EPI_ISL_4600599, EPI_ISL_4600600, EPI_ISL_4600601, EPI_ISL_4600602, EPI_ISL_4600603, EPI_ISL_4600607, EPI_ISL_4600608, EPI_ISL_4600609, EPI_ISL_4600610, EPI_ISL_4600611, EPI_ISL_4600612, EPI_ISL_4600613, EPI_ISL_4600614, EPI_ISL_4600615, EPI_ISL_4600616, EPI_ISL_4600617, EPI_ISL_4600618, EPI_ISL_4600619, EPI_ISL_4600620, EPI_ISL_4600621, EPI_ISL_4600622, EPI_ISL_4600623, EPI_ISL_4600624, EPI_ISL_4600625, EPI_ISL_4600626, EPI_ISL_4600627, EPI_ISL_4600631, EPI_ISL_4600641, EPI_ISL_4600643, EPI_ISL_4600644, EPI_ISL_4600645, EPI_ISL_4600646, EPI_ISL_4600647, EPI_ISL_4600648, EPI_ISL_4600649, EPI_ISL_4600650, EPI_ISL_4600653, EPI_ISL_4600654, EPI_ISL_4600655, EPI_ISL_4600656, EPI_ISL_4600657, EPI_ISL_4600658, EPI_ISL_4600659, EPI_ISL_4600660, EPI_ISL_4600661, EPI_ISL_4600662, EPI_ISL_4600663, EPI_ISL_4600664, EPI_ISL_4600665, EPI_ISL_4600666, EPI_ISL_4600667, EPI_ISL_4600668, EPI_ISL_4600669, EPI_ISL_4600670, EPI_ISL_4600671, EPI_ISL_4600672, EPI_ISL_4600673, EPI_ISL_4600674, EPI_ISL_4600675, EPI_ISL_4600676, EPI_ISL_4600677, EPI_ISL_4600678, EPI_ISL_4600679, EPI_ISL_4600680, EPI_ISL_4600681, EPI_ISL_4600682, EPI_ISL_4600683, EPI_ISL_4600686, EPI_ISL_4600687, EPI_ISL_4600688, EPI_ISL_4600689, EPI_ISL_4600690, EPI_ISL_4600691, EPI_ISL_4600695, EPI_ISL_4600696, EPI_ISL_4600697, EPI_ISL_4600698, EPI_ISL_4600699, EPI_ISL_4600700, EPI_ISL_4600701, EPI_ISL_4600702, EPI_ISL_4600703, EPI_ISL_4600704, EPI_ISL_4600705, EPI_ISL_4600706, EPI_ISL_4600707, EPI_ISL_4600708, EPI_ISL_4600709, EPI_ISL_4600710, EPI_ISL_4600711, EPI_ISL_4600712, EPI_ISL_4600713, EPI_ISL_4600714, EPI_ISL_4600715, EPI_ISL_4600718, EPI_ISL_4600719, EPI_ISL_4600720, EPI_ISL_4600721, EPI_ISL_4600722, EPI_ISL_4600723, EPI_ISL_4600724, EPI_ISL_4600725, EPI_ISL_4600726, EPI_ISL_4600734, EPI_ISL_4600735, EPI_ISL_4600736, EPI_ISL_4600737, EPI_ISL_4600738, EPI_ISL_4600739, EPI_ISL_4600740, EPI_ISL_4600741, EPI_ISL_4600742, EPI_ISL_4600743, EPI_ISL_4600744, EPI_ISL_4600745, EPI_ISL_4600746, EPI_ISL_4600747, EPI_ISL_4600751, EPI_ISL_4600752, EPI_ISL_4600753, EPI_ISL_4600754, EPI_ISL_4600755, EPI_ISL_4600756, EPI_ISL_4600757, EPI_ISL_4600758, EPI_ISL_4600759, EPI_ISL_4600760, EPI_ISL_4600761, EPI_ISL_4600762, EPI_ISL_4600763, EPI_ISL_4600770, EPI_ISL_4600771, EPI_ISL_4600773, EPI_ISL_4600775, EPI_ISL_4600776, EPI_ISL_4600777, EPI_ISL_4600778, EPI_ISL_4600779, EPI_ISL_4600780, EPI_ISL_4600781, EPI_ISL_4600782, EPI_ISL_4600783, EPI_ISL_4600784, EPI_ISL_4600797, EPI_ISL_4600798, EPI_ISL_4600799, EPI_ISL_4600800, EPI_ISL_4600801, EPI_ISL_4600802, EPI_ISL_4600803, EPI_ISL_4600804, EPI_ISL_4600805, EPI_ISL_4600806, EPI_ISL_4600807, EPI_ISL_4600808, EPI_ISL_4600809, EPI_ISL_4600810, EPI_ISL_4600813, EPI_ISL_4600815, EPI_ISL_4600816, EPI_ISL_4600817, EPI_ISL_4600818, EPI_ISL_4600819, EPI_ISL_4600820, EPI_ISL_4600821, EPI_ISL_4600822, EPI_ISL_4600823, EPI_ISL_4600824, EPI_ISL_4600825, EPI_ISL_4600826, EPI_ISL_4600827, EPI_ISL_4600828, EPI_ISL_4600829, EPI_ISL_4600830, EPI_ISL_4600831, EPI_ISL_4600832, EPI_ISL_4600833, EPI_ISL_4600834, EPI_ISL_4600835, EPI_ISL_4600836, EPI_ISL_4600837, EPI_ISL_4600838, EPI_ISL_4600839, EPI_ISL_4600840, EPI_ISL_4600841, EPI_ISL_4600842, EPI_ISL_4600844, EPI_ISL_4600845, EPI_ISL_4600846, EPI_ISL_4600847, EPI_ISL_4600848, EPI_ISL_4600849, EPI_ISL_4600850, EPI_ISL_4600851, EPI_ISL_4600852, EPI_ISL_4600853, EPI_ISL_4600904, EPI_ISL_4600905, EPI_ISL_4600906, EPI_ISL_4600907, EPI_ISL_4600908, EPI_ISL_4600909, EPI_ISL_4600910, EPI_ISL_4600911, EPI_ISL_4600912, EPI_ISL_4600913, EPI_ISL_4600914, EPI_ISL_4600915, EPI_ISL_4600916, EPI_ISL_4600917, EPI_ISL_4600918, EPI_ISL_4600919, EPI_ISL_4600920, EPI_ISL_4600921, EPI_ISL_4600922, EPI_ISL_4600923, EPI_ISL_4600924, EPI_ISL_4600925, EPI_ISL_4600926, EPI_ISL_4600927, EPI_ISL_4600928, EPI_ISL_4600929, EPI_ISL_4600938, EPI_ISL_4600939, EPI_ISL_4600940, EPI_ISL_4600943, EPI_ISL_4600944, EPI_ISL_4600945, EPI_ISL_4600946, EPI_ISL_4600947, EPI_ISL_4600948, EPI_ISL_4600949, EPI_ISL_4600950, EPI_ISL_4600952, EPI_ISL_4600953, EPI_ISL_4600954, EPI_ISL_4600955, EPI_ISL_4600956, EPI_ISL_4600957, EPI_ISL_4600961, EPI_ISL_4600968, EPI_ISL_4600969, EPI_ISL_4600970, EPI_ISL_4600971, EPI_ISL_4600972, EPI_ISL_4600977, EPI_ISL_4600978, EPI_ISL_4600983, EPI_ISL_4600984, EPI_ISL_4600985, EPI_ISL_4600987, EPI_ISL_4600988, EPI_ISL_4600989, EPI_ISL_4600990, EPI_ISL_4600991, EPI_ISL_4600992, EPI_ISL_4600993, EPI_ISL_4600997, EPI_ISL_4600998, EPI_ISL_4600999, EPI_ISL_4601000, EPI_ISL_4601001, EPI_ISL_4601002, EPI_ISL_4601003, EPI_ISL_4601004, EPI_ISL_4601005, EPI_ISL_4601006, EPI_ISL_4601007, EPI_ISL_4601010, EPI_ISL_4601011, EPI_ISL_4601012, EPI_ISL_4601013, EPI_ISL_4601014, EPI_ISL_4601015, EPI_ISL_4601016, EPI_ISL_4601040, EPI_ISL_4601041, EPI_ISL_4601042, EPI_ISL_4601043, EPI_ISL_4601044, EPI_ISL_4601045, EPI_ISL_4601050, EPI_ISL_4601051, EPI_ISL_4601056, EPI_ISL_4601067, EPI_ISL_4601069, EPI_ISL_4601073, EPI_ISL_4601083, EPI_ISL_4601084, EPI_ISL_4601085, EPI_ISL_4601086, EPI_ISL_4601087, EPI_ISL_4601088, EPI_ISL_4601089, EPI_ISL_4601090, EPI_ISL_4601096, EPI_ISL_4601103, EPI_ISL_4601110, EPI_ISL_4601111, EPI_ISL_4601112, EPI_ISL_4601114 | | | |
| see above | Labo Analyses Med | National Reference Center for Viruses of Respiratory Infections, Institut Pasteur, Paris | Anais Inquel; Angela Brisebarre; Anne Duterrail; Anne Holstein; Arnaud Jean; Axelle Paquin; Baptiste Moreira; Beate Heym; Brieuc Gestin; Bruno Berdin; Camille Capel; Carine Lecorre; Caroline Malderet; Christophe Malabat; Claire Felloni; Corinne Maufrais; Dauriat; Domitille Leman; Dorine Bouvet; Emmanuel Pasquier; Eric Morin; Etienne Simon-Lorière; F. Kerdavid; FrançOis-Xavier Le Guillou; Frédéric Lemoine; Gilles Abs; GréGoire Potiron; Guillaume Aubin; GwéNolé Prigent; Jc Fiorina; Jean-FrançOis Comes; Julien Fumey; Karine Michez; L. Courdavault; Lbm Dubet; Leonard; Louise Lefrançois; Marc Duval-Arnould; Marie-Jeanne Blanc-Galiby; Marion Barbet; Maud Vanpeene; Murielle Chiron; Méline Bizard; Nabila Belhouachi; OphéLie Said-Delattre; Pascal Maillet; Patrice Nizard; Philippe Girard; Philippe Miara; S Perron; S. Cocco; Sandrine Simon; Slim El Khiari; Slim El-Khiari; Sophie Chalmin; Stephanie Hainos; StéPhane Romand; Sylvie Behillil; Sylvie Lamy; Sylvie Robert; Sylvie Van der Werf; Valentin Robbe; ValéRie Weber; Vanessa Magin; Vincent Enouf; Xavier Guettier |
| EPI_ISL_4326572, EPI_ISL_4606381, EPI_ISL_4606383, EPI_ISL_4606386, EPI_ISL_4606390, EPI_ISL_4606391, EPI_ISL_4606393, EPI_ISL_4606395, EPI_ISL_4606397, EPI_ISL_4606519, EPI_ISL_4606526, EPI_ISL_4607081 | | | |
| see above | MIRIALIS CLUSE | CNR Virus des Infections Respiratoires - France SUD | Antonin Bal; Bruno Lina; Gregory Destras; Gwendolyne Burfin; Hadrien Regue; Laurence Josset; Martine Valette; Quentin Semanas |
| EPI_ISL_4606215, EPI_ISL_4606327, EPI_ISL_4606329, EPI_ISL_4606331, EPI_ISL_4606333, EPI_ISL_4606334, EPI_ISL_4606337, EPI_ISL_4606339, EPI_ISL_4606340, EPI_ISL_4606365, EPI_ISL_4606370, EPI_ISL_4606372, EPI_ISL_4606375, EPI_ISL_4606377, EPI_ISL_4606378, EPI_ISL_4606392, EPI_ISL_4606400, EPI_ISL_4606403, EPI_ISL_4606405, EPI_ISL_4606407, EPI_ISL_4606412, EPI_ISL_4606419, EPI_ISL_4606421, EPI_ISL_4606559, EPI_ISL_4606563, EPI_ISL_4606566, EPI_ISL_4606570, EPI_ISL_4606572, EPI_ISL_4606574, EPI_ISL_4606575, EPI_ISL_4606578, EPI_ISL_4606579, EPI_ISL_4606581, EPI_ISL_4606583, EPI_ISL_4606584, EPI_ISL_4606585, EPI_ISL_4606587, EPI_ISL_4606588, EPI_ISL_4606590, EPI_ISL_4606591, EPI_ISL_4606592, EPI_ISL_4606593, EPI_ISL_4606595, EPI_ISL_4606596, EPI_ISL_4606597, EPI_ISL_4606598, EPI_ISL_4606821, EPI_ISL_4606850, EPI_ISL_4606854, EPI_ISL_4606855, EPI_ISL_4606862, EPI_ISL_4606885, EPI_ISL_4606895, EPI_ISL_4606901, EPI_ISL_4606945, EPI_ISL_4606946, EPI_ISL_4606947, EPI_ISL_4606948, EPI_ISL_4606949, EPI_ISL_4606957, EPI_ISL_4606959, EPI_ISL_4606971, EPI_ISL_4606974, EPI_ISL_4606975, EPI_ISL_4606982, EPI_ISL_4606983, EPI_ISL_4606984, EPI_ISL_4606986 | | | |
| see above | MIRIALIS CLUSES BECHET | CNR Virus des Infections Respiratoires - France SUD | Antonin Bal; Bruno Lina; Gregory Destras; Gwendolyne Burfin; Hadrien Regue; Laurence Josset; Martine Valette; Quentin Semanas |

| | | | |
|---|---|---|---|
| EPI_ISL_4604588, EPI_ISL_4606650, EPI_ISL_4606652, EPI_ISL_4606664, EPI_ISL_4606683, EPI_ISL_4606715, EPI_ISL_4606717, EPI_ISL_4606718, EPI_ISL_4606719, EPI_ISL_4606720, EPI_ISL_4606721, EPI_ISL_4606722, EPI_ISL_4606724, EPI_ISL_4606728, EPI_ISL_4606729, EPI_ISL_4606730, EPI_ISL_4606731, EPI_ISL_4606732, EPI_ISL_4606733, EPI_ISL_4606734, EPI_ISL_4606737, EPI_ISL_4606738, EPI_ISL_4606763, EPI_ISL_4606774, EPI_ISL_4606783, EPI_ISL_4606787, EPI_ISL_4606797, EPI_ISL_4606832, EPI_ISL_4606837, EPI_ISL_4606840, EPI_ISL_4606848, EPI_ISL_4606863, EPI_ISL_4606919, EPI_ISL_4606921, EPI_ISL_4606923, EPI_ISL_4606937, EPI_ISL_4606938 | | | |
| see above | ORIAPOLE | CNR Virus des Infections Respiratoires - France SUD | Antonin Bal; Bruno Lina; Gregory Destras; Gwendolyne Burfin; Hadrien Regue; Laurence Josset; Martine Valette; Quentin Semanas |
| EPI_ISL_4606175 | SCM B12 | CNR Virus des Infections Respiratoires - France SUD | Antonin Bal; Bruno Lina; Gregory Destras; Gwendolyne Burfin; Hadrien Regue; Laurence Josset; Martine Valette; Quentin Semanas |
| EPI_ISL_4605823, EPI_ISL_4605825, EPI_ISL_4605828, EPI_ISL_4605831, EPI_ISL_4605835, EPI_ISL_4605837, EPI_ISL_4605838 | | | |
| see above | SYNLAB GASCOGNE | CNR Virus des Infections Respiratoires - France SUD | Antonin Bal; Bruno Lina; Gregory Destras; Gwendolyne Burfin; Hadrien Regue; Laurence Josset; Martine Valette; Quentin Semanas |
| EPI_ISL_4605608 | SYNLAB PAYS DE SAVOIE | CNR Virus des Infections Respiratoires - France SUD | Antonin Bal; Bruno Lina; Gregory Destras; Gwendolyne Burfin; Hadrien Regue; Laurence Josset; Martine Valette; Quentin Semanas |
| EPI_ISL_4473234, EPI_ISL_4473235, EPI_ISL_4473236, EPI_ISL_4473237, EPI_ISL_4473238, EPI_ISL_4473239, EPI_ISL_4473240, EPI_ISL_4473241 | | | |
| see above | VIROLOGY LABORATORY-CHU NICE | VIROLOGY LABORATORY-CHU NICE | Aicha El Yakine; Geraldine Gonfrier; Jean Machowiak; Sebastien Vitale; Valerie Giordanengo; Virginie Flahou |
