## Supplementary material for "SARS-COV-2 δ variant drives the pandemic in India and Europe via two subvariants": Acknowledgement table on the GISAID genomes used in this study

We gratefully acknowledge the following Authors from the Originating laboratories responsible for obtaining the specimens, as well as the Submitting laboratories where the genome data were generated and shared via GISAID, on which this research is based.

All Submitters of data may be contacted directly via www.gisaid.org

Authors are sorted alphabetically.

| Accession ID | Originating Laboratory | Submitting Laboratory | Authors |
|---|---|---|---|
| EPI_ISL_4445123, EPI_ISL_4445124, EPI_ISL_4445125, EPI_ISL_4445126, EPI_ISL_4445127, EPI_ISL_4445128, EPI_ISL_4445129, EPI_ISL_4445130, EPI_ISL_4445131, EPI_ISL_4445132, EPI_ISL_4445133, EPI_ISL_4445134, EPI_ISL_4445135, EPI_ISL_4445136, EPI_ISL_4445137, EPI_ISL_4445138, EPI_ISL_4445139, EPI_ISL_4445140, EPI_ISL_4445141, EPI_ISL_4445142, EPI_ISL_4445143, EPI_ISL_4445144, EPI_ISL_4445145, EPI_ISL_4445146, EPI_ISL_4445147, EPI_ISL_4445148, EPI_ISL_4445149, EPI_ISL_4445150, EPI_ISL_4445151, EPI_ISL_4445152, EPI_ISL_4445153, EPI_ISL_4445154, EPI_ISL_4445155, EPI_ISL_4445156, EPI_ISL_4445157, EPI_ISL_4445158, EPI_ISL_4445159, EPI_ISL_4445160, EPI_ISL_4445161, EPI_ISL_4445162, EPI_ISL_4445163, EPI_ISL_4445164, EPI_ISL_4445165, EPI_ISL_4445166, EPI_ISL_4445167, EPI_ISL_4445168, EPI_ISL_4445169, EPI_ISL_4445170, EPI_ISL_4445171, EPI_ISL_4445172, EPI_ISL_4445173, EPI_ISL_4445174, EPI_ISL_4445175, EPI_ISL_4445176, EPI_ISL_4445177, EPI_ISL_4445178, EPI_ISL_4445179, EPI_ISL_4445180, EPI_ISL_4445181, EPI_ISL_4445182, EPI_ISL_4445183, EPI_ISL_4445184, EPI_ISL_4445185, EPI_ISL_4445186, EPI_ISL_4445187, EPI_ISL_4445188, EPI_ISL_4445189, EPI_ISL_4445190, EPI_ISL_4445191, EPI_ISL_4445192, EPI_ISL_4445193, EPI_ISL_4445194, EPI_ISL_4445195, EPI_ISL_4445196, EPI_ISL_4445197, EPI_ISL_4445198, EPI_ISL_4445199, EPI_ISL_4445200, EPI_ISL_4445201, EPI_ISL_4445202, EPI_ISL_4445203, EPI_ISL_4445204, EPI_ISL_4445205, EPI_ISL_4445206, EPI_ISL_4445207, EPI_ISL_4445208, EPI_ISL_4445209, EPI_ISL_4445210, EPI_ISL_4445211, EPI_ISL_4445212, EPI_ISL_4445213, EPI_ISL_4445214, EPI_ISL_4445215, EPI_ISL_4445216, EPI_ISL_4445217, EPI_ISL_4445218, EPI_ISL_4445219, EPI_ISL_4445220, EPI_ISL_4445221, EPI_ISL_4445222, EPI_ISL_4445223, EPI_ISL_4445224, EPI_ISL_4445225, EPI_ISL_4445226, EPI_ISL_4445227, EPI_ISL_4445228, EPI_ISL_4445229, EPI_ISL_4445230, EPI_ISL_4445231, EPI_ISL_4445232, EPI_ISL_4445233, EPI_ISL_4445234, EPI_ISL_4445235, EPI_ISL_4445236, EPI_ISL_4445237, EPI_ISL_4445238, EPI_ISL_4445239, EPI_ISL_4445240, EPI_ISL_4445241, EPI_ISL_4445242, EPI_ISL_4445243, EPI_ISL_4445244, EPI_ISL_4445245, EPI_ISL_4445246, EPI_ISL_4445247, EPI_ISL_4445248, EPI_ISL_4445249, EPI_ISL_4445250, EPI_ISL_4445251, EPI_ISL_4445252, EPI_ISL_4445253, EPI_ISL_4445254, EPI_ISL_4445255, EPI_ISL_4445256, EPI_ISL_4445257, EPI_ISL_4445258, EPI_ISL_4445259, EPI_ISL_4445260, EPI_ISL_4445261, EPI_ISL_4445262, EPI_ISL_4445263, EPI_ISL_4445264, EPI_ISL_4445265, EPI_ISL_4445266, EPI_ISL_4445267, EPI_ISL_4445268, EPI_ISL_4445269, EPI_ISL_4445270, EPI_ISL_4445271, EPI_ISL_4445272, EPI_ISL_4445273, EPI_ISL_4445274, EPI_ISL_4445275, EPI_ISL_4445276, EPI_ISL_4445277, EPI_ISL_4445278, EPI_ISL_4445279, EPI_ISL_4445280, EPI_ISL_4445281, EPI_ISL_4445282, EPI_ISL_4445283, EPI_ISL_4445284, EPI_ISL_4445285, EPI_ISL_4445286, EPI_ISL_4445287, EPI_ISL_4445288, EPI_ISL_4445289, EPI_ISL_4445290, EPI_ISL_4445291, EPI_ISL_4445292, EPI_ISL_4445293, EPI_ISL_4445294, EPI_ISL_4445295 | | | |
| see above | Eurofins MVZ Labor Gelsenkirchen | Robert Koch Institute | |
| EPI_ISL_4612734 | Kerckhoff-Klinik GmbH | Robert Koch Institute | |
| EPI_ISL_4609653, EPI_ISL_4614280, EPI_ISL_4614281, EPI_ISL_4614282, EPI_ISL_4614283, EPI_ISL_4614284, EPI_ISL_4614285, EPI_ISL_4614286, EPI_ISL_4614287, EPI_ISL_4614288, EPI_ISL_4614289, EPI_ISL_4614292, EPI_ISL_4614293, EPI_ISL_4614294, EPI_ISL_4614295, EPI_ISL_4614296, EPI_ISL_4614297, EPI_ISL_4614298, EPI_ISL_4614299 | | | |
| see above | Klinikum Ernst von Bergmann gemeinnützige GmbH - stationärer Bereich | Robert Koch Institute | |
| EPI_ISL_4518781, EPI_ISL_4518791, EPI_ISL_4518793, EPI_ISL_4518797, EPI_ISL_4518798, EPI_ISL_4518802, EPI_ISL_4518803 | | | |
| see above | Labor Berlin Charité Vivantes GmbH / Institut für Virologie | Charité Universitätsmedizin Berlin, Institut für Virologie/Labor Berlin | Barbara Mühlemann; Christian Drosten; Christine Stephan; Peter Menzel; Rolf Schwarzer; Terry Jones; Victor M Corman |
| EPI_ISL_4612254, EPI_ISL_4612269, EPI_ISL_4612284, EPI_ISL_4612310, EPI_ISL_4612313, EPI_ISL_4614261, EPI_ISL_4614262, EPI_ISL_4614263, EPI_ISL_4614264, EPI_ISL_4614265, EPI_ISL_4614266, EPI_ISL_4614267, EPI_ISL_4614268, EPI_ISL_4614269 | | | |
| see above | Labor Dr. Heidrich & Kollegen MVZ GmbH Hamburg | Robert Koch Institute | |
| EPI_ISL_4609278, EPI_ISL_4609279, EPI_ISL_4609284, EPI_ISL_4609292, EPI_ISL_4609294, EPI_ISL_4609300, EPI_ISL_4609305, EPI_ISL_4609314, EPI_ISL_4609317, EPI_ISL_4609329, EPI_ISL_4609352, EPI_ISL_4609367, EPI_ISL_4609371, EPI_ISL_4609374, EPI_ISL_4609377, EPI_ISL_4609386, EPI_ISL_4609390, EPI_ISL_4609392, EPI_ISL_4609398, EPI_ISL_4609400, EPI_ISL_4609426, EPI_ISL_4609427, EPI_ISL_4609430, EPI_ISL_4609447, EPI_ISL_4609460, EPI_ISL_4609467, EPI_ISL_4609470, EPI_ISL_4609479, EPI_ISL_4609482, EPI_ISL_4609483, EPI_ISL_4609487, EPI_ISL_4609490, EPI_ISL_4609491, EPI_ISL_4609493, EPI_ISL_4609500, EPI_ISL_4609503, EPI_ISL_4609518, EPI_ISL_4609521, EPI_ISL_4609525, EPI_ISL_4609533, EPI_ISL_4609534, EPI_ISL_4609535, EPI_ISL_4609540, EPI_ISL_4609546, EPI_ISL_4609553, EPI_ISL_4609555, EPI_ISL_4609557, EPI_ISL_4609561, EPI_ISL_4609562, EPI_ISL_4609564, EPI_ISL_4609565, EPI_ISL_4609566, EPI_ISL_4609568, EPI_ISL_4614006, EPI_ISL_4614007, EPI_ISL_4614008, EPI_ISL_4614009, EPI_ISL_4614010, EPI_ISL_4614011, EPI_ISL_4614012, EPI_ISL_4614013, EPI_ISL_4614014, EPI_ISL_4614015, EPI_ISL_4614017, EPI_ISL_4614018, EPI_ISL_4614020, EPI_ISL_4614021, EPI_ISL_4614022, EPI_ISL_4614023, EPI_ISL_4614024, EPI_ISL_4614025, EPI_ISL_4614026, EPI_ISL_4614027, EPI_ISL_4614028, EPI_ISL_4614029, EPI_ISL_4614030, EPI_ISL_4614032, EPI_ISL_4614033, EPI_ISL_4614034, EPI_ISL_4614036, EPI_ISL_4614037, EPI_ISL_4614038, EPI_ISL_4614039, EPI_ISL_4614040, EPI_ISL_4614041, EPI_ISL_4614043, EPI_ISL_4614044, EPI_ISL_4614048, EPI_ISL_4614050, EPI_ISL_4614051, EPI_ISL_4614052, EPI_ISL_4614053, EPI_ISL_4614054, EPI_ISL_4614055, EPI_ISL_4614056, EPI_ISL_4614057, EPI_ISL_4614058, EPI_ISL_4614060, EPI_ISL_4614061, EPI_ISL_4614062, EPI_ISL_4614064, EPI_ISL_4614065, EPI_ISL_4614066, EPI_ISL_4614067, EPI_ISL_4614068, EPI_ISL_4614069, EPI_ISL_4614070, EPI_ISL_4614071, EPI_ISL_4614072, EPI_ISL_4614073, EPI_ISL_4614075, EPI_ISL_4614076, EPI_ISL_4614077, EPI_ISL_4614078, EPI_ISL_4614079, EPI_ISL_4614080, EPI_ISL_4614081, EPI_ISL_4614082, EPI_ISL_4614083, EPI_ISL_4614084, EPI_ISL_4614085, EPI_ISL_4614086, EPI_ISL_4614087, EPI_ISL_4614088, EPI_ISL_4614089, EPI_ISL_4614090, EPI_ISL_4614092, EPI_ISL_4614093, EPI_ISL_4614097, EPI_ISL_4614098, EPI_ISL_4614099, EPI_ISL_4614100, EPI_ISL_4614101, EPI_ISL_4614102, EPI_ISL_4614104, EPI_ISL_4614105, EPI_ISL_4614106, EPI_ISL_4614107, EPI_ISL_4614108, EPI_ISL_4614109, EPI_ISL_4614110, EPI_ISL_4614113, EPI_ISL_4614115, EPI_ISL_4614116, EPI_ISL_4614117, EPI_ISL_4614118, EPI_ISL_4614119, EPI_ISL_4614120, EPI_ISL_4614122, EPI_ISL_4614123, EPI_ISL_4614124, EPI_ISL_4614125, EPI_ISL_4614126, EPI_ISL_4614127, EPI_ISL_4614128, EPI_ISL_4614129, EPI_ISL_4614130, EPI_ISL_4614131, EPI_ISL_4614133, EPI_ISL_4614134, EPI_ISL_4614135, EPI_ISL_4614136, EPI_ISL_4614137, EPI_ISL_4614138, EPI_ISL_4614139, EPI_ISL_4614140, EPI_ISL_4614141, EPI_ISL_4614142, EPI_ISL_4614143, EPI_ISL_4614144, EPI_ISL_4614145, EPI_ISL_4614146, EPI_ISL_4614147, EPI_ISL_4614148, EPI_ISL_4614161, EPI_ISL_4614163, EPI_ISL_4614166, EPI_ISL_4614167, EPI_ISL_4614168, EPI_ISL_4614170, EPI_ISL_4614172, EPI_ISL_4614173, EPI_ISL_4614174, EPI_ISL_4614175, EPI_ISL_4614176, EPI_ISL_4614177, EPI_ISL_4614178, EPI_ISL_4614179, EPI_ISL_4614220, EPI_ISL_4614221, EPI_ISL_4614222, EPI_ISL_4614224, EPI_ISL_4614225, EPI_ISL_4614226, EPI_ISL_4614227, EPI_ISL_4614228, EPI_ISL_4614229, EPI_ISL_4614230, EPI_ISL_4614232, EPI_ISL_4614233, EPI_ISL_4614234, EPI_ISL_4614235, EPI_ISL_4614236, EPI_ISL_4614238, EPI_ISL_4614240, EPI_ISL_4614241, EPI_ISL_4614242, EPI_ISL_4614243, EPI_ISL_4614244, EPI_ISL_4614245, EPI_ISL_4614246, EPI_ISL_4614247, EPI_ISL_4614248, EPI_ISL_4614249, EPI_ISL_4614250, EPI_ISL_4614251, EPI_ISL_4614252, EPI_ISL_4614253, EPI_ISL_4614254, EPI_ISL_4614255, EPI_ISL_4614256, EPI_ISL_4614257, EPI_ISL_4614258, EPI_ISL_4614259 | | | |
| see above | Labor Dr. Wisplinghoff - Köln | Robert Koch Institute | |
| EPI_ISL_4446556, EPI_ISL_4446558, EPI_ISL_4446559, EPI_ISL_4446560, EPI_ISL_4446563, EPI_ISL_4446564, EPI_ISL_4446565, EPI_ISL_4446566, EPI_ISL_4446568, EPI_ISL_4446571, EPI_ISL_4446572, EPI_ISL_4446575, EPI_ISL_4446692, EPI_ISL_4609745, EPI_ISL_4609746, EPI_ISL_4609747, EPI_ISL_4609750, EPI_ISL_4609752, EPI_ISL_4609753, EPI_ISL_4609756, EPI_ISL_4609757, EPI_ISL_4609758, EPI_ISL_4611771, EPI_ISL_4611774, EPI_ISL_4611775, EPI_ISL_4611776, EPI_ISL_4611778, EPI_ISL_4611779, EPI_ISL_4611783, EPI_ISL_4611784, EPI_ISL_4611787, EPI_ISL_4611788, EPI_ISL_4611789, EPI_ISL_4611790, EPI_ISL_4611791, EPI_ISL_4611792, EPI_ISL_4611793, EPI_ISL_4611794, EPI_ISL_4611795, EPI_ISL_4611796, EPI_ISL_4611797, EPI_ISL_4611798, EPI_ISL_4611799, EPI_ISL_4611800, EPI_ISL_4611801, EPI_ISL_4611802, EPI_ISL_4611803, EPI_ISL_4611804, EPI_ISL_4611805, EPI_ISL_4612177, EPI_ISL_4612178, EPI_ISL_4612179, EPI_ISL_4612180, EPI_ISL_4612181, EPI_ISL_4612182, EPI_ISL_4612183, EPI_ISL_4612184, EPI_ISL_4612185, EPI_ISL_4612186, EPI_ISL_4612187, EPI_ISL_4612188, EPI_ISL_4612189, EPI_ISL_4612190, EPI_ISL_4612191, EPI_ISL_4612192, EPI_ISL_4612193, EPI_ISL_4612194, EPI_ISL_4612195, EPI_ISL_4612196, EPI_ISL_4612197, EPI_ISL_4612198, EPI_ISL_4612407, EPI_ISL_4612416, EPI_ISL_4612442, EPI_ISL_4612450, EPI_ISL_4612458, EPI_ISL_4612464, EPI_ISL_4612473, EPI_ISL_4614000, EPI_ISL_4614002 | | | |
| see above | Labor Mönchengladbach MVZ Dr. Stein + Kollegen GbR | Robert Koch Institute | |
| EPI_ISL_4609707 | Limbach - MVZ Gemeinschaftslabor Suhl | Robert Koch Institute | |
| EPI_ISL_4611530, EPI_ISL_4611548, EPI_ISL_4611552, EPI_ISL_4611558, EPI_ISL_4611559, EPI_ISL_4611561, EPI_ISL_4611563, EPI_ISL_4611565, EPI_ISL_4611566, EPI_ISL_4611567, EPI_ISL_4611568, EPI_ISL_4611570, EPI_ISL_4611574, EPI_ISL_4611577, EPI_ISL_4611579, EPI_ISL_4611582, EPI_ISL_4611584, EPI_ISL_4611586, EPI_ISL_4611587, EPI_ISL_4611588, EPI_ISL_4611589, EPI_ISL_4611590, EPI_ISL_4611591, EPI_ISL_4611593, EPI_ISL_4611594, EPI_ISL_4611595, EPI_ISL_4611596, EPI_ISL_4611598, EPI_ISL_4611599, EPI_ISL_4611600, EPI_ISL_4611601, EPI_ISL_4611602, EPI_ISL_4611603, EPI_ISL_4611607, EPI_ISL_4611610, EPI_ISL_4611611, EPI_ISL_4611612, EPI_ISL_4611613, EPI_ISL_4611614, EPI_ISL_4611618, EPI_ISL_4611619, EPI_ISL_4611620, EPI_ISL_4611621, EPI_ISL_4611622, EPI_ISL_4611623, EPI_ISL_4611624, EPI_ISL_4611625, EPI_ISL_4611626, EPI_ISL_4611627, EPI_ISL_4611628, EPI_ISL_4611629, EPI_ISL_4611630, EPI_ISL_4611631, EPI_ISL_4611632, EPI_ISL_4611633, EPI_ISL_4611634, EPI_ISL_4611635, EPI_ISL_4611636, EPI_ISL_4611637, EPI_ISL_4611638, EPI_ISL_4611639, EPI_ISL_4611640, EPI_ISL_4611641, EPI_ISL_4611642, EPI_ISL_4611643, EPI_ISL_4611644, EPI_ISL_4611645, EPI_ISL_4611646, EPI_ISL_4611647, EPI_ISL_4611648, EPI_ISL_4611649, EPI_ISL_4611650, EPI_ISL_4611651, EPI_ISL_4611652, EPI_ISL_4611653, EPI_ISL_4611654, EPI_ISL_4611655, EPI_ISL_4611656 | | | |
| see above | Limbach - MVZ Humangenetik Ulm | Robert Koch Institute | |
| EPI_ISL_4614004 | Limbach - MVZ Labor Eveld & Kollegen Essen | Robert Koch Institute | |
| EPI_ISL_4446845, EPI_ISL_4446849, EPI_ISL_4446858, EPI_ISL_4446859, EPI_ISL_4446860, EPI_ISL_4446862, EPI_ISL_4446864, EPI_ISL_4446871, EPI_ISL_4446873, EPI_ISL_4446875, EPI_ISL_4446876, EPI_ISL_4609698, EPI_ISL_4609700, EPI_ISL_4609702, EPI_ISL_4609706, EPI_ISL_4609708, EPI_ISL_4609709, EPI_ISL_4609710, EPI_ISL_4609711, EPI_ISL_4609724, EPI_ISL_4609725, EPI_ISL_4609728, EPI_ISL_4609729, EPI_ISL_4609734, EPI_ISL_4609739, EPI_ISL_4609741, EPI_ISL_4610089, EPI_ISL_4610093, EPI_ISL_4610099, EPI_ISL_4610102, EPI_ISL_4610103, EPI_ISL_4610106, EPI_ISL_4610109, EPI_ISL_4610110, EPI_ISL_4610111, EPI_ISL_4610115, EPI_ISL_4610119, EPI_ISL_4610121, EPI_ISL_4610124, EPI_ISL_4610127, EPI_ISL_4612132, EPI_ISL_4612134, EPI_ISL_4612138, EPI_ISL_4612139, EPI_ISL_4612142, EPI_ISL_4612146, EPI_ISL_4612147, EPI_ISL_4612148, EPI_ISL_4612149, EPI_ISL_4612152, EPI_ISL_4612153, EPI_ISL_4612154, EPI_ISL_4612162, EPI_ISL_4612169, EPI_ISL_4612170, EPI_ISL_4612171, EPI_ISL_4612420, EPI_ISL_4612431, EPI_ISL_4612469, EPI_ISL_4612477, EPI_ISL_4612485, EPI_ISL_4612497, EPI_ISL_4612498, EPI_ISL_4612499, EPI_ISL_4612500, EPI_ISL_4612726, EPI_ISL_4612727, EPI_ISL_4612728, EPI_ISL_4612736, EPI_ISL_4612738, EPI_ISL_4612740, EPI_ISL_4612742, EPI_ISL_4612750, EPI_ISL_4612753, EPI_ISL_4612754, EPI_ISL_4612755, EPI_ISL_4612758, EPI_ISL_4612759 | | | |
| see above | MDI Limbach Berlin GmbH; MVZ Labor Berlin | Robert Koch Institute | |
| EPI_ISL_4610195, EPI_ISL_4610207 | MVZ Dr. Eberhard & Partner Dortmund | Robert Koch Institute | |
| EPI_ISL_4614341, EPI_ISL_4614342, EPI_ISL_4614343, EPI_ISL_4614344, EPI_ISL_4614345, EPI_ISL_4614347, EPI_ISL_4614348, EPI_ISL_4614349, EPI_ISL_4614351, EPI_ISL_4614353, EPI_ISL_4614354, EPI_ISL_4614355, EPI_ISL_4614356, EPI_ISL_4614357, EPI_ISL_4614358, EPI_ISL_4614359, EPI_ISL_4614360, EPI_ISL_4614362 | | | |
| see above | MVZ Labor Dr. Fenner und Kollegen (Standort Hamburg) | Robert Koch Institute | |
| EPI_ISL_4609633, EPI_ISL_4609643, EPI_ISL_4609644, EPI_ISL_4609646, EPI_ISL_4609685, EPI_ISL_4609743, EPI_ISL_4610118, EPI_ISL_4614003 | | | |
| see above | MVZ Labor Dr. Limbach & Kollegen GbR | Robert Koch Institute | |
| EPI_ISL_4614001 | MVZ Medizinisches Labor Hannover GmbH | Robert Koch Institute | |
| EPI_ISL_4611227, EPI_ISL_4611230, EPI_ISL_4611232 | Medizinisches Zentrallabor Altenburg GmbH & Co.KG | Robert Koch Institute | |
| EPI_ISL_4612749 | SLK-Kliniken Heilbronn GmbH - Institut für Laboratoriumsmedizin | Robert Koch Institute | |
| EPI_ISL_4613371, EPI_ISL_4613372, EPI_ISL_4613376, EPI_ISL_4613379, EPI_ISL_4613382, EPI_ISL_4613384, EPI_ISL_4613386, EPI_ISL_4613387, EPI_ISL_4613389, EPI_ISL_4613390, EPI_ISL_4613392, EPI_ISL_4613397, EPI_ISL_4613399, EPI_ISL_4613404, EPI_ISL_4613405, EPI_ISL_4613407, EPI_ISL_4613412, EPI_ISL_4613414, EPI_ISL_4613418, EPI_ISL_4613419, EPI_ISL_4613420, EPI_ISL_4613425, EPI_ISL_4613429, EPI_ISL_4613430, EPI_ISL_4613431, EPI_ISL_4613435, EPI_ISL_4613436, EPI_ISL_4613438, EPI_ISL_4613439, EPI_ISL_4613444, EPI_ISL_4613446, EPI_ISL_4613447, EPI_ISL_4613448, EPI_ISL_4613452, EPI_ISL_4613459, EPI_ISL_4613460, EPI_ISL_4613465, EPI_ISL_4613470, EPI_ISL_4613472, EPI_ISL_4613478, EPI_ISL_4613483, EPI_ISL_4613487, EPI_ISL_4613494, EPI_ISL_4613495, EPI_ISL_4613496, EPI_ISL_4613501, EPI_ISL_4613505, EPI_ISL_4613511, EPI_ISL_4613514, EPI_ISL_4613516, EPI_ISL_4613520, EPI_ISL_4613521, EPI_ISL_4613523, EPI_ISL_4613526, EPI_ISL_4613527, EPI_ISL_4613529, EPI_ISL_4613531, EPI_ISL_4613532, EPI_ISL_4613533, EPI_ISL_4613534, EPI_ISL_4613536, EPI_ISL_4613538, EPI_ISL_4613543, EPI_ISL_4613544, EPI_ISL_4613550, EPI_ISL_4613551, EPI_ISL_4613555, EPI_ISL_4613557, EPI_ISL_4613559, EPI_ISL_4613561, EPI_ISL_4613562, EPI_ISL_4613565, EPI_ISL_4613566, EPI_ISL_4613568, EPI_ISL_4613569, EPI_ISL_4613570, EPI_ISL_4613575, EPI_ISL_4613576, EPI_ISL_4613577, EPI_ISL_4613578, EPI_ISL_4613579, EPI_ISL_4613580, EPI_ISL_4613581, EPI_ISL_4613582, EPI_ISL_4613583, EPI_ISL_4613585, EPI_ISL_4613586, EPI_ISL_4613587, EPI_ISL_4613591, EPI_ISL_4613592, EPI_ISL_4613594, EPI_ISL_4613597, EPI_ISL_4613602, EPI_ISL_4613606, EPI_ISL_4613608, EPI_ISL_4613610, EPI_ISL_4613612, EPI_ISL_4613613, EPI_ISL_4613615, EPI_ISL_4613617, EPI_ISL_4613618, EPI_ISL_4613619, EPI_ISL_4613620, EPI_ISL_4613622, EPI_ISL_4613623, EPI_ISL_4613628, EPI_ISL_4613631, EPI_ISL_4613632, EPI_ISL_4613635, EPI_ISL_4613637, EPI_ISL_4613638, EPI_ISL_4613640, EPI_ISL_4613642, EPI_ISL_4613644, EPI_ISL_4613645, EPI_ISL_4613647, EPI_ISL_4613649, EPI_ISL_4613651, EPI_ISL_4613655, EPI_ISL_4613657, EPI_ISL_4613658, EPI_ISL_4613661, EPI_ISL_4613664, EPI_ISL_4613667, EPI_ISL_4613672, EPI_ISL_4613675, EPI_ISL_4613680, EPI_ISL_4613683, EPI_ISL_4613692, EPI_ISL_4613699, EPI_ISL_4613712, EPI_ISL_4613719, EPI_ISL_4613722, EPI_ISL_4613729, EPI_ISL_4613732, EPI_ISL_4613734, EPI_ISL_4613735, EPI_ISL_4613736, EPI_ISL_4613737, EPI_ISL_4613739, EPI_ISL_4613740, EPI_ISL_4613742, EPI_ISL_4613746, EPI_ISL_4613750, EPI_ISL_4613754, EPI_ISL_4613755, EPI_ISL_4613761, EPI_ISL_4613762, EPI_ISL_4613764, EPI_ISL_4613765, EPI_ISL_4613766, EPI_ISL_4613769, EPI_ISL_4613770, EPI_ISL_4613771, EPI_ISL_4613772, EPI_ISL_4613776, EPI_ISL_4613780, EPI_ISL_4613781, EPI_ISL_4613785, EPI_ISL_4613789, EPI_ISL_4613790, EPI_ISL_4613791, EPI_ISL_4613792, EPI_ISL_4613793, EPI_ISL_4613796, EPI_ISL_4613798, EPI_ISL_4613799, EPI_ISL_4613801, EPI_ISL_4613802, EPI_ISL_4613804, EPI_ISL_4613810, EPI_ISL_4613814, EPI_ISL_4613816, EPI_ISL_4613825, EPI_ISL_4613827, EPI_ISL_4613829, EPI_ISL_4613830, EPI_ISL_4613835, EPI_ISL_4613836, EPI_ISL_4613838, EPI_ISL_4613842, EPI_ISL_4613844, EPI_ISL_4613847, EPI_ISL_4613851, EPI_ISL_4613854, EPI_ISL_4613855, EPI_ISL_4613857, EPI_ISL_4613859, EPI_ISL_4613867, EPI_ISL_4613868, EPI_ISL_4613870, EPI_ISL_4613871, EPI_ISL_4613874, EPI_ISL_4613875, EPI_ISL_4613876, EPI_ISL_4613877, EPI_ISL_4613878, EPI_ISL_4613879, EPI_ISL_4613882, EPI_ISL_4613885, EPI_ISL_4613890, EPI_ISL_4613892, EPI_ISL_4613893, EPI_ISL_4613897, EPI_ISL_4613899, EPI_ISL_4613901, EPI_ISL_4613904, EPI_ISL_4613905, EPI_ISL_4613909, EPI_ISL_4613911, EPI_ISL_4613913, EPI_ISL_4613914, EPI_ISL_4613916, EPI_ISL_4613921, EPI_ISL_4613934, EPI_ISL_4613942, EPI_ISL_4613943, EPI_ISL_4613944, EPI_ISL_4613946, EPI_ISL_4613947, EPI_ISL_4613948, EPI_ISL_4613951, EPI_ISL_4613953, EPI_ISL_4613954, EPI_ISL_4613955, EPI_ISL_4613959, EPI_ISL_4613964, EPI_ISL_4613965, EPI_ISL_4613966, EPI_ISL_4613969, EPI_ISL_4613973, EPI_ISL_4613976, EPI_ISL_4613977, EPI_ISL_4613979, EPI_ISL_4613983, EPI_ISL_4613984, EPI_ISL_4613985, EPI_ISL_4613988, EPI_ISL_4613996 | | | |
| see above | SYNLAB MVZ Leinfelden-Echterdingen | Robert Koch Institute | |
| EPI_ISL_4613373, EPI_ISL_4613375, EPI_ISL_4613381, EPI_ISL_4613391, EPI_ISL_4613393, EPI_ISL_4613395, EPI_ISL_4613396, EPI_ISL_4613398, EPI_ISL_4613400, EPI_ISL_4613401, EPI_ISL_4613402, EPI_ISL_4613403, EPI_ISL_4613406, EPI_ISL_4613408, EPI_ISL_4613426, EPI_ISL_4613427, EPI_ISL_4613428, EPI_ISL_4613434, EPI_ISL_4613437, EPI_ISL_4613441, EPI_ISL_4613442, EPI_ISL_4613443, EPI_ISL_4613445, EPI_ISL_4613453, EPI_ISL_4613454, EPI_ISL_4613457, EPI_ISL_4613461, EPI_ISL_4613466, EPI_ISL_4613467, EPI_ISL_4613468, EPI_ISL_4613473, EPI_ISL_4613476, EPI_ISL_4613477, EPI_ISL_4613479, EPI_ISL_4613486, EPI_ISL_4613488, EPI_ISL_4613490, EPI_ISL_4613491, EPI_ISL_4613492, EPI_ISL_4613497, EPI_ISL_4613499, EPI_ISL_4613508, EPI_ISL_4613515, EPI_ISL_4613517, EPI_ISL_4613518, EPI_ISL_4613519, EPI_ISL_4613528, EPI_ISL_4613540, EPI_ISL_4613545, EPI_ISL_4613553, EPI_ISL_4613554, EPI_ISL_4613558, EPI_ISL_4613564, EPI_ISL_4613571, EPI_ISL_4613573, EPI_ISL_4613589, EPI_ISL_4613596, EPI_ISL_4613607, EPI_ISL_4613609, EPI_ISL_4613614, EPI_ISL_4613616, EPI_ISL_4613621, EPI_ISL_4613625, EPI_ISL_4613626, EPI_ISL_4613627, EPI_ISL_4613629, EPI_ISL_4613630, EPI_ISL_4613634, EPI_ISL_4613636, EPI_ISL_4613648, EPI_ISL_4613650, EPI_ISL_4613653, EPI_ISL_4613654, EPI_ISL_4613659, EPI_ISL_4613660, EPI_ISL_4613665, EPI_ISL_4613674, EPI_ISL_4613677, EPI_ISL_4613679, EPI_ISL_4613682, EPI_ISL_4613685, EPI_ISL_4613686, EPI_ISL_4613689, EPI_ISL_4613695, EPI_ISL_4613696, EPI_ISL_4613701, EPI_ISL_4613704, EPI_ISL_4613706, EPI_ISL_4613707, EPI_ISL_4613708, EPI_ISL_4613709, EPI_ISL_4613710, EPI_ISL_4613711, EPI_ISL_4613713, EPI_ISL_4613715, EPI_ISL_4613721, EPI_ISL_4613724, EPI_ISL_4613725, EPI_ISL_4613727, EPI_ISL_4613728, EPI_ISL_4613730, EPI_ISL_4613731, EPI_ISL_4613738, EPI_ISL_4613741, EPI_ISL_4613743, EPI_ISL_4613744, EPI_ISL_4613747, EPI_ISL_4613752, EPI_ISL_4613767, EPI_ISL_4613768, EPI_ISL_4613774, EPI_ISL_4613779, EPI_ISL_4613782, EPI_ISL_4613784, EPI_ISL_4613787, EPI_ISL_4613788, EPI_ISL_4613794, EPI_ISL_4613803, EPI_ISL_4613805, EPI_ISL_4613806, EPI_ISL_4613808, EPI_ISL_4613813, EPI_ISL_4613815, EPI_ISL_4613817, EPI_ISL_4613818, EPI_ISL_4613820, EPI_ISL_4613821, EPI_ISL_4613822, EPI_ISL_4613824, EPI_ISL_4613828, EPI_ISL_4613831, EPI_ISL_4613832, EPI_ISL_4613834, EPI_ISL_4613837, EPI_ISL_4613839, EPI_ISL_4613843, EPI_ISL_4613845, EPI_ISL_4613849, EPI_ISL_4613850, EPI_ISL_4613853, EPI_ISL_4613858, EPI_ISL_4613860, EPI_ISL_4613861, EPI_ISL_4613862, EPI_ISL_4613863, EPI_ISL_4613864, EPI_ISL_4613872, EPI_ISL_4613873, EPI_ISL_4613886, EPI_ISL_4613891, EPI_ISL_4613898, EPI_ISL_4613900, EPI_ISL_4613902, EPI_ISL_4613903, EPI_ISL_4613907, EPI_ISL_4613915, EPI_ISL_4613920, EPI_ISL_4613922, EPI_ISL_4613923, EPI_ISL_4613924, EPI_ISL_4613933, EPI_ISL_4613936, EPI_ISL_4613938, EPI_ISL_4613939, EPI_ISL_4613940, EPI_ISL_4613945, EPI_ISL_4613949, EPI_ISL_4613952, EPI_ISL_4613962, EPI_ISL_4613963, EPI_ISL_4613970, EPI_ISL_4613971, EPI_ISL_4613972, EPI_ISL_4613974, EPI_ISL_4613975, EPI_ISL_4613980, EPI_ISL_4613986, EPI_ISL_4613987, EPI_ISL_4613989, EPI_ISL_4613990, EPI_ISL_4613992, EPI_ISL_4613998, EPI_ISL_4613999 | | | |
| see above | SYNLAB MVZ Leverkusen | Robert Koch Institute | |
| EPI_ISL_4614376, EPI_ISL_4614379, EPI_ISL_4614381, EPI_ISL_4614462, EPI_ISL_4614470, EPI_ISL_4614482 | Sonic - MVZ Medizinisches Labor Bremen GmbH | Robert Koch Institute | |
| EPI_ISL_4609692 | Universitätsklinikum Münster Institut für Virologie | Robert Koch Institute | |
| EPI_ISL_4551725, EPI_ISL_4551726, EPI_ISL_4551727, EPI_ISL_4551728, EPI_ISL_4551729, EPI_ISL_4551730, EPI_ISL_4551731, EPI_ISL_4551732, EPI_ISL_4551733, EPI_ISL_4551734, EPI_ISL_4551738, EPI_ISL_4551739, EPI_ISL_4551740, EPI_ISL_4551741, EPI_ISL_4551742, EPI_ISL_4551743, EPI_ISL_4551744, EPI_ISL_4551745, EPI_ISL_4551746, EPI_ISL_4551747, EPI_ISL_4551748, EPI_ISL_4551749, EPI_ISL_4551750, EPI_ISL_4551751, EPI_ISL_4551752, EPI_ISL_4551753, EPI_ISL_4551754, EPI_ISL_4551755, EPI_ISL_4551756, EPI_ISL_4551757, EPI_ISL_4551758, EPI_ISL_4551759, EPI_ISL_4551760, EPI_ISL_4551761, EPI_ISL_4551762, EPI_ISL_4551763, EPI_ISL_4551764, EPI_ISL_4551765, EPI_ISL_4551766, EPI_ISL_4551767, EPI_ISL_4551768, EPI_ISL_4551769, EPI_ISL_4551770, EPI_ISL_4551771, EPI_ISL_4551772, EPI_ISL_4551773, EPI_ISL_4551774, EPI_ISL_4551775, EPI_ISL_4551776, EPI_ISL_4551777, EPI_ISL_4551778, EPI_ISL_4551779, EPI_ISL_4551868, EPI_ISL_4551870, EPI_ISL_4551872, EPI_ISL_4551874, EPI_ISL_4551876, EPI_ISL_4551878, EPI_ISL_4551956, EPI_ISL_4551957, EPI_ISL_4551958, EPI_ISL_4551959, EPI_ISL_4551960, EPI_ISL_4551961, EPI_ISL_4551962, EPI_ISL_4551963, EPI_ISL_4551964, EPI_ISL_4551965, EPI_ISL_4551966, EPI_ISL_4551967, EPI_ISL_4551968, EPI_ISL_4551969, EPI_ISL_4551970, EPI_ISL_4551971, EPI_ISL_4551972, EPI_ISL_4551973, EPI_ISL_4551974, EPI_ISL_4551975, EPI_ISL_4551976, EPI_ISL_4551977, EPI_ISL_4551978, EPI_ISL_4551979, EPI_ISL_4551980, EPI_ISL_4551981, EPI_ISL_4551982, EPI_ISL_4551983, EPI_ISL_4551984, EPI_ISL_4551985, | | | |

| | | | |
|---|---|---|---|
| EPI_ISL_4551987, EPI_ISL_4551988, EPI_ISL_4551989, EPI_ISL_4551997, EPI_ISL_4551998, EPI_ISL_4551999, EPI_ISL_4552000, EPI_ISL_4552001, EPI_ISL_4552002, EPI_ISL_4552003, EPI_ISL_4552004 | | | |
| see above | ZOTZ KLIMAS MVZ Düsseldorf-Centrum GbR ÜBAG für Labormedizin, Genetik, Zytologie, Pathologie | Center of Medical Microbiology, Virology, and Hospital Hygiene, University of Duesseldorf | Alexander Dilthey; Andreas Walker; Daniel Strelow; Jessica Nicolai; Jörg Timm; Katrin Hoffmann; Klaus Pfeffer; Lisanna Hülse; Malte Kohns Vasconcelos; Maximilian Damagnez; Nadine Lübke; Patrick Finzer; Rainer Zotz; Tobias Wienemann; Torsten Houwaart |
| EPI_ISL_4612067 | amedes Medlab Arnold Analytik MVZ GmbH | Robert Koch Institute | |
| EPI_ISL_4611875, EPI_ISL_4611880, EPI_ISL_4611890, EPI_ISL_4611892, EPI_ISL_4611893, EPI_ISL_4611898, EPI_ISL_4611899, EPI_ISL_4611908, EPI_ISL_4611910, EPI_ISL_4611913, EPI_ISL_4611915, EPI_ISL_4611916, EPI_ISL_4611918, EPI_ISL_4611919, EPI_ISL_4611920, EPI_ISL_4611921, EPI_ISL_4611922, EPI_ISL_4611923, EPI_ISL_4611924, EPI_ISL_4611925, EPI_ISL_4611926, EPI_ISL_4611927, EPI_ISL_4611928, EPI_ISL_4611929, EPI_ISL_4611930, EPI_ISL_4611931, EPI_ISL_4611932, EPI_ISL_4611933, EPI_ISL_4611934, EPI_ISL_4611935, EPI_ISL_4611936, EPI_ISL_4611937, EPI_ISL_4611938, EPI_ISL_4611939, EPI_ISL_4611940, EPI_ISL_4611941, EPI_ISL_4611942, EPI_ISL_4611943, EPI_ISL_4611944, EPI_ISL_4611945, EPI_ISL_4611946, EPI_ISL_4611947, EPI_ISL_4611953, EPI_ISL_4611955, EPI_ISL_4611956, EPI_ISL_4611957, EPI_ISL_4611959, EPI_ISL_4611966, EPI_ISL_4611967, EPI_ISL_4611969, EPI_ISL_4611973, EPI_ISL_4611974, EPI_ISL_4611976, EPI_ISL_4611977, EPI_ISL_4611978, EPI_ISL_4611979, EPI_ISL_4611980, EPI_ISL_4611982, EPI_ISL_4611983, EPI_ISL_4611984, EPI_ISL_4611985, EPI_ISL_4611986, EPI_ISL_4611995, EPI_ISL_4612011 | | | |
| see above | nordlab - Partnerschaftspraxis für Laboratoriumsmedizin | Robert Koch Institute | |
