## Supplementary material for "SARS-COV-2 δ variant drives the pandemic in India and Europe via two subvariants": Acknowledgement table on the GISAID genomes used in this study

We gratefully acknowledge the following Authors from the Originating laboratories responsible for obtaining the specimens, as well as the Submitting laboratories where the genome data were generated and shared via GISAID, on which this research is based.

All Submitters of data may be contacted directly via www.gisaid.org

Authors are sorted alphabetically.

| Accession ID | Originating Laboratory | Submitting Laboratory | Authors |
|---|---|---|---|
| EPI_ISL_4436491, EPI_ISL_4436495, EPI_ISL_4436550 | A.S.L. CITTA DI TORINO - OSPEDALE MAURIZIANO | Fondazione del Piemonte per l'Oncologia IRCCS | Antonino Sottile; Giorgio Giardina; Paola Marino; Silvia Brossa |
| EPI_ISL_4572067, EPI_ISL_4572205 | A.S.L. CUNEO 1 | Fondazione del Piemonte per l'Oncologia IRCCS | Antonino Sottile; Giorgio Giardina; Paola Marino; Silvia Brossa |
| EPI_ISL_4606209 | ASTRALAB TASSIGNY | CNR Virus des Infections Respiratoires - France SUD | Antonin Bal; Bruno Lina; Gregory Destras; Gwendolyne Burfin; Hadrien Regue; Laurence Josset; Martine Valette; Quentin Semanas |
| EPI_ISL_4557564 | AULSS 2 Marca Trevigiana | Istituto Zooprofilattico Sperimentale delle Venezie | Adelaide Milani; Alessia Schivo; Alice Fusaro; Ambra Pastori; Annalisa Salviato; Antonia Ricci; Calogero Terregino; Edoardo Giussani; Elisa Palumbo; Erika Giorgia Quaranta; Isabella Monne; Luca Tassoni |
| EPI_ISL_4571221, EPI_ISL_4571230 | AULSS 3 Venezia | UOSD Genetica e Citogenetica - Azienda ULSS 3 Serenissima; Istituto Zooprofilattico Sperimentale delle Venezie | Adelaide Milani; Alessia Schivo; Alice Fusaro; Ambra Pastori; Annalisa Salviato; Antonia Ricci; Calogero Terregino; Claudia Perini; Edoardo Giussani; Elisa Palumbo; Elisa Squarcina; Erika Giorgia Quaranta; Isabella Monne; Laura Bevilacqua; Laura Squarzon; Luca Sorino; Luca Tassoni; Mosé Favarato |
| EPI_ISL_4571129 | AULSS 6 Euganea | Istituto Zooprofilattico Sperimentale delle Venezie | Adelaide Milani; Alessia Schivo; Alice Fusaro; Ambra Pastori; Annalisa Salviato; Antonia Ricci; Calogero Terregino; Edoardo Giussani; Elisa Palumbo; Erika Giorgia Quaranta; Isabella Monne; Luca Tassoni |
| EPI_ISL_4571275 | AULSS 9 Scaligera - Legnago | UOSD Genetica e Citogenetica - Azienda ULSS 3 Serenissima; Istituto Zooprofilattico Sperimentale delle Venezie | Adelaide Milani; Alessia Schivo; Alice Fusaro; Ambra Pastori; Annalisa Salviato; Antonia Ricci; Calogero Terregino; Claudia Perini; Edoardo Giussani; Elisa Palumbo; Elisa Squarcina; Erika Giorgia Quaranta; Isabella Monne; Laura Bevilacqua; Laura Squarzon; Luca Sorino; Luca Tassoni; Mosé Favarato |
| EPI_ISL_4606281 | AX BIO OCEAN | CNR Virus des Infections Respiratoires - France SUD | Antonin Bal; Bruno Lina; Gregory Destras; Gwendolyne Burfin; Hadrien Regue; Laurence Josset; Martine Valette; Quentin Semanas |
| EPI_ISL_3933472, EPI_ISL_4205130, EPI_ISL_4213138, EPI_ISL_4345131, EPI_ISL_4345393, EPI_ISL_4364136, EPI_ISL_4364137, EPI_ISL_4364154, EPI_ISL_4364347, EPI_ISL_4364395, EPI_ISL_4468597, EPI_ISL_4468598, EPI_ISL_4468622, EPI_ISL_4545256, EPI_ISL_4545263, EPI_ISL_4545269, EPI_ISL_4545276, EPI_ISL_4545286, EPI_ISL_4545315, EPI_ISL_4601121, EPI_ISL_4601122, EPI_ISL_4601123, EPI_ISL_4601125, EPI_ISL_4601126, EPI_ISL_4601138 | | | |
| see above | AZDelta | AZ Delta Medical Laboratories in Roeselare, Belgium | Dieter De Smet; Frederik Van Hoecke; Geert Martens; on behalf of AZ Delta COVID-19 Genomics core (member of Genomic surveillance of SARS-CoV-2 in Belgium network) |
| EPI_ISL_4494549 | Aegis Sciences Corporation | Centers for Disease Control and Prevention Division of Viral Diseases, Pathogen Discovery | Alec Vest; Benjamin Rambo-Martin; Christopher Gulvick; Clinton Paden; Cyndi Clark; Dakota Howard; Dhwani Batra; Dillon Nall; Duncan MacCannell; Erisa Sula; Ethan Sanders; Holly Houdeshell; Jason Caravas; Kristine Lacek; Matthew Hardison; Matthew Schmerer; Ola Kvalvaag; Patrick Campbell; Peter Cook; Rob Case; Scott Sammons; Shatavia Morrison; Shaun Westlund; Tymeckia Kendall; Victoria Caban Figueroa; Vikramsinha Ghorpade; Yvette Unoarumhi |
| EPI_ISL_4479431, EPI_ISL_4508183 | Akershus University Hospital, Department for Microbiology and Infectious Disease Control | Norwegian Institute of Public Health, Department of Virology | Atiya R Ali; Debech Nadia; Engebretsen Serina Beate; Garcia Llorente Ignacio; Hilde Elshaug; Hilde Vollan; Jon Bråte; Kamilla Heddeland Instefjord; Karoline Bragstad; Kathrine Stene-Johansen; Line Victoria Moen; Marie Paulsen Madsen; Olav Hungnes; Pedersen Benedikte Nevjen; Rasmus Riis Kopperud |
| EPI_ISL_4275365, EPI_ISL_4275369, EPI_ISL_4275393 | Algemeen Medisch Labo | Labo Klinische Biologie, UZA | Basil Britto Xavier; Christine Lammens; Herman Goossens; Ines Verbesselt; Jasmine Coppens; Kathleen Holemans; Marie Le Mercier; Veerle Matheeussen |
| EPI_ISL_4303874 | Althaia. Xarxa Assistencial Universitària de Manresa | IrsiCaixa | Antonia Flor; Bonaventura Clotet; Bonaventura Clotet Gloria Trujillo; Carolina Gonzalez-Fernandez; Eulalia Grau; Francesc Catala-Moll; Jaume Trapé; Marc Noguera-Julian; Maria Casadellà; Mariona Parera; Miquel Micó; Pilar Armengol; Rafel Perez Vidal; Roger Paredes |
| EPI_ISL_4510581, EPI_ISL_4510582 | Arlon | Plateforme de testing Namuroise | Degosserie Jonathan; Demars Aurore; Denis Olivier; Lesly Nyinkeu Kemamen; Maschietto Céline; Mullier François; Nicolas Gilliard; Nobis Chloé; Otto Gaetan |
| EPI_ISL_4284581 | Azienda Sanitaria dell'Alto Adige - Laboratorio Aziendale di Microbiologia e Virologia | Azienda Sanitaria dell'Alto Adige | Irene Bianconi |
| EPI_ISL_4605315, EPI_ISL_4605391 | BIOLAB 33 | CNR Virus des Infections Respiratoires - France SUD | Antonin Bal; Bruno Lina; Gregory Destras; Gwendolyne Burfin; Hadrien Regue; Laurence Josset; Martine Valette; Quentin Semanas |
| EPI_ISL_4620583, EPI_ISL_4620769 | Bayerisches Landesamt für Gesundheit und Lebensmittelsicherheit (LGL) | Robert Koch Institute | |
| EPI_ISL_4227834, EPI_ISL_4227940, EPI_ISL_4438023, EPI_ISL_4438090, EPI_ISL_4438197, EPI_ISL_4441321, EPI_ISL_4441327, EPI_ISL_4441427, EPI_ISL_4441486, EPI_ISL_4441694, EPI_ISL_4611443, EPI_ISL_4611487, EPI_ISL_4614388, EPI_ISL_4614628, EPI_ISL_4614651, EPI_ISL_4614684, EPI_ISL_4614786, EPI_ISL_4614869 | | | |
| see above | Bioscientia Labor Wermsdorf | Robert Koch Institute | |
| EPI_ISL_4094623, EPI_ISL_4094869, EPI_ISL_4095063, EPI_ISL_4095123, EPI_ISL_4095222, EPI_ISL_4095234, EPI_ISL_4095276, EPI_ISL_4095392, EPI_ISL_4095660, EPI_ISL_4095689, EPI_ISL_4285096, EPI_ISL_4285289, EPI_ISL_4285293, EPI_ISL_4285569, EPI_ISL_4285784, EPI_ISL_4286299, EPI_ISL_4286361, EPI_ISL_4286370, EPI_ISL_4286440, EPI_ISL_4304090, EPI_ISL_4304254, EPI_ISL_4304603, EPI_ISL_4304685, EPI_ISL_4454435, EPI_ISL_4454719, EPI_ISL_4487945, EPI_ISL_4487954, EPI_ISL_4488086, EPI_ISL_4497322, EPI_ISL_4516605, EPI_ISL_4516858, EPI_ISL_4516890, EPI_ISL_4516906, EPI_ISL_4517034, EPI_ISL_4517431, EPI_ISL_4517481, EPI_ISL_4517555, EPI_ISL_4517564, EPI_ISL_4517566, EPI_ISL_4517668, EPI_ISL_4517722 | | | |
| see above | Broad Institute Clinical Research Sequencing Platform | Infectious Disease Program, Broad Institute of Harvard and MIT | Adams, G.; B.L.; B.W.; Bauer, M.; Birren; Blumenstiel, B.; Brown, C.; Carter, A.; Chaluvadi, S.; D.J.; DeFelice, M.; DeRuff, K.; Dodge, S.; Gabriel, S.; Gallagher, G.; Gladden-Young, A.; Granger, B.; J.E.; K.J.; Lagerborg, K.; Larkin, K.; Lee, M.; Lemieux; Lennon, N.; Loreth, C.; Madoff, L.; McGovern, S.; Meldrim, J.; Normandin, E.; P.C.; Park; Pearlman, L.; Reilly, S.; Rudy, M.; Sabeti; Siddle; Smole, S.; Tomkins-Tinch, C.; Vicente, G.; and MacInnis |
| EPI_ISL_4539760 | C.H. LE DAMANY LANNION-TRESTEL | CHU Pontchaillou | DE TAYRAC Marie; DENOUAL Florent; DUFOUR Marie Jose; ETCHEVERRY Amandine; FEBREAU Christine; GALIBERT Marie Dominique; GROLHIER Claire; JAGLINE Steven; PRONIER Charlotte; QUENET Benjamin; SASSI Mohamed; THIBAULT Vincent |
| EPI_ISL_4440511, EPI_ISL_4440560 | CENTOGENE Frankfurt Laboratory: Niederlassung Industriepark Höchst | Robert Koch Institute | |
| EPI_ISL_4605009, EPI_ISL_4605418, EPI_ISL_4605427, EPI_ISL_4605458, EPI_ISL_4605484, EPI_ISL_4606741 | CERBALLIANCE OCCITANIE | CNR Virus des Infections Respiratoires - France SUD | Antonin Bal; Bruno Lina; Gregory Destras; Gwendolyne Burfin; Hadrien Regue; Laurence Josset; Martine Valette; Quentin Semanas |
| EPI_ISL_4127720, EPI_ISL_4254807, EPI_ISL_4254808, EPI_ISL_4254809 | CHM - SITE CHR | Institut de Pathologie et Genetique (IPG) | Jérémie Gras; Pascale Hilbert |
| EPI_ISL_4413130, EPI_ISL_4632210 | CHUV | Laboratory of genomics and metagenomics | Claire Bertelli; Damien Jacot; Gilbert Greub; Sébastien Aeby; Trestan Pillonel |
| EPI_ISL_4201559 | CLILAB | Microbiology Department | Aida Gonzalez-Diaz; Carmen Ardanuy; Jordi Camara; Jordi Niubó; Laura Calatayud; M Angeles Domínguez; Sara Marti; Yolanda Hernandez |
| EPI_ISL_4325390, EPI_ISL_4606010 | CNR Virus des Infections Respiratoires - France SUD | CNR Virus des Infections Respiratoires - France SUD | Antonin Bal; Bruno Lina; Gregory Destras; Gwendolyne Burfin; Hadrien Regue; Laurence Josset; Martine Valette; Quentin Semanas |
| EPI_ISL_4551949, EPI_ISL_4551950, EPI_ISL_4551954, EPI_ISL_4551955 | Center of Medical Microbiology, Virology, and Hospital Hygiene, University of Duesseldorf | Center of Medical Microbiology, Virology, and Hospital Hygiene, University of Duesseldorf | Alexander Dilthey; Andreas Walker; Daniel Strelow; Jessica Nicolai; Jörg Timm; Klaus Pfeffer; Lisanna Hülse; Malte Kohns Vasconcelos; Maximilian Damagnez; Nadine Lübke; Tobias Wienemann; Torsten Houwaart |
| EPI_ISL_4078034 | Centre Hospitalier Universitaire Clermont-Ferrand | CHU Clermont-Ferrand, service de virologie | Bisseux Maxime; Combes Patricia; Henquell Cécile; Mirand Audrey |
| EPI_ISL_4505601 | Centrum voor Medische Analyse | Labo Klinische Biologie, UZA | Basil Britto Xavier; Christine Lammens; Herman Goossens; Ines Verbesselt; Jasmine Coppens; Kathleen Holemans; Marie Le Mercier; Veerle Matheeussen |
| EPI_ISL_4131379, EPI_ISL_4131439, EPI_ISL_4131455, EPI_ISL_4131470, EPI_ISL_4521282, EPI_ISL_4521300, EPI_ISL_4521318, EPI_ISL_4521319, EPI_ISL_4521335, EPI_ISL_4521337, EPI_ISL_4521340, EPI_ISL_4521386 | | | |
| see above | Clinical Microbiology, Infection Prevention and Control | Section for Molecular Diagnostics | Björn Hallström; Jonas Björkman |
| EPI_ISL_4450886 | Clinical Virology | Clinical Bacteriology | Adrian Egli; Alfredo Mari; Fanny Wegner; Hans Hirsch; Helena MB Seth-Smith; Julia Bielicki; Karoline Leuzinger; Manuel Battegay; Tim Roloff |
| EPI_ISL_4492600 | Clinique Notre Dame à Hermalle | GIGA Medical Genomics | Bouchra Boujemla; Claire Gourzonès; Cécile Meex; Keith Durkin; Laurent Gillet; Maria Artesi; Marie-Pierre Hayette; Nadine Cambisano; Nathalie Renotte; Olivier Ek; Sébastien Bontems; Vincent Bours |

| | | | |
|---|---|---|---|
| EPI_ISL_4069549, EPI_ISL_4274807, EPI_ISL_4274829, EPI_ISL_4274845, EPI_ISL_4274857, EPI_ISL_4493370, EPI_ISL_4493374, EPI_ISL_4493383, EPI_ISL_4493385, EPI_ISL_4493411, EPI_ISL_4629170 | | | |
| see above | Cliniques universitaires Saint-Luc | UCLouvain/IREC/MBLG | Benoit Kabamba Mukadi; Bertrand Bearzatto; Jean Ruelle |
| EPI_ISL_3989411, EPI_ISL_3989443, EPI_ISL_3989474, EPI_ISL_3989486, EPI_ISL_3989515, EPI_ISL_3989532, EPI_ISL_3989622, EPI_ISL_3989646, EPI_ISL_3989675, EPI_ISL_3989676, EPI_ISL_3989790, EPI_ISL_3989827, EPI_ISL_3989834, EPI_ISL_3989892, EPI_ISL_3989955, EPI_ISL_3989964, EPI_ISL_3990005, EPI_ISL_3990078, EPI_ISL_3990128, EPI_ISL_3990167, EPI_ISL_3990182, EPI_ISL_3990308, EPI_ISL_4006987, EPI_ISL_4007004, EPI_ISL_4007012, EPI_ISL_4007016, EPI_ISL_4007049, EPI_ISL_4007091, EPI_ISL_4007102, EPI_ISL_4007138, EPI_ISL_4007214, EPI_ISL_4007258, EPI_ISL_4007285, EPI_ISL_4007289, EPI_ISL_4007293, EPI_ISL_4007299, EPI_ISL_4007340, EPI_ISL_4007349, EPI_ISL_4007362, EPI_ISL_4007372, EPI_ISL_4007434, EPI_ISL_4007442, EPI_ISL_4007606, EPI_ISL_4007660, EPI_ISL_4007690, EPI_ISL_4007722, EPI_ISL_4007800, EPI_ISL_4007870, EPI_ISL_4007973, EPI_ISL_4007991, EPI_ISL_4008007, EPI_ISL_4008084, EPI_ISL_4008147, EPI_ISL_4008291, EPI_ISL_4008323, EPI_ISL_4008350, EPI_ISL_4049719, EPI_ISL_4049727, EPI_ISL_4049734, EPI_ISL_4049745, EPI_ISL_4049747, EPI_ISL_4049755, EPI_ISL_4049855, EPI_ISL_4049902, EPI_ISL_4049903, EPI_ISL_4049919, EPI_ISL_4049956, EPI_ISL_4049971, EPI_ISL_4049981, EPI_ISL_4049997, EPI_ISL_4050061, EPI_ISL_4050069, EPI_ISL_4050075, EPI_ISL_4050081, EPI_ISL_4050093, EPI_ISL_4050098, EPI_ISL_4050105, EPI_ISL_4050111, EPI_ISL_4050141, EPI_ISL_4050169, EPI_ISL_4050186, EPI_ISL_4050224, EPI_ISL_4050275, EPI_ISL_4050287, EPI_ISL_4050288, EPI_ISL_4050297, EPI_ISL_4050314, EPI_ISL_4050327, EPI_ISL_4050334, EPI_ISL_4050341, EPI_ISL_4050474, EPI_ISL_4050478, EPI_ISL_4050485, EPI_ISL_4050546, EPI_ISL_4050550, EPI_ISL_4050572, EPI_ISL_4069409, EPI_ISL_4069412, EPI_ISL_4069421, EPI_ISL_4069423, EPI_ISL_4069432, EPI_ISL_4069473, EPI_ISL_4105486, EPI_ISL_4105508, EPI_ISL_4105541, EPI_ISL_4105551, EPI_ISL_4105565, EPI_ISL_4105572, EPI_ISL_4105573, EPI_ISL_4105577, EPI_ISL_4105582, EPI_ISL_4105630, EPI_ISL_4105669, EPI_ISL_4105699, EPI_ISL_4105707, EPI_ISL_4105710, EPI_ISL_4105791, EPI_ISL_4105826, EPI_ISL_4105842, EPI_ISL_4105861, EPI_ISL_4105879, EPI_ISL_4105887, EPI_ISL_4105930, EPI_ISL_4105932, EPI_ISL_4105972, EPI_ISL_4105981, EPI_ISL_4170405, EPI_ISL_4170430, EPI_ISL_4170432, EPI_ISL_4170434, EPI_ISL_4170442, EPI_ISL_4170468, EPI_ISL_4170474, EPI_ISL_4170503, EPI_ISL_4170520, EPI_ISL_4170533, EPI_ISL_4170571, EPI_ISL_4170623, EPI_ISL_4170643, EPI_ISL_4170645, EPI_ISL_4170651, EPI_ISL_4170682, EPI_ISL_4170692, EPI_ISL_4199665, EPI_ISL_4199682, EPI_ISL_4199763, EPI_ISL_4199801, EPI_ISL_4199802, EPI_ISL_4199811, EPI_ISL_4199831, EPI_ISL_4199836, EPI_ISL_4199845, EPI_ISL_4199849, EPI_ISL_4199857, EPI_ISL_4199878, EPI_ISL_4199882, EPI_ISL_4199889, EPI_ISL_4199909, EPI_ISL_4199919, EPI_ISL_4199925, EPI_ISL_4199943, EPI_ISL_4199961, EPI_ISL_4199973, EPI_ISL_4199982, EPI_ISL_4199995, EPI_ISL_4200002, EPI_ISL_4200032, EPI_ISL_4200042, EPI_ISL_4200065, EPI_ISL_4200068, EPI_ISL_4200077, EPI_ISL_4200116, EPI_ISL_4200144, EPI_ISL_4200153, EPI_ISL_4200158, EPI_ISL_4200218, EPI_ISL_4200235, EPI_ISL_4200266, EPI_ISL_4200270, EPI_ISL_4200294, EPI_ISL_4200307, EPI_ISL_4200321, EPI_ISL_4200353, EPI_ISL_4200373, EPI_ISL_4200377, EPI_ISL_4200384, EPI_ISL_4200385, EPI_ISL_4200387, EPI_ISL_4200388, EPI_ISL_4200400, EPI_ISL_4200430, EPI_ISL_4200445, EPI_ISL_4200447, EPI_ISL_4200466, EPI_ISL_4200479, EPI_ISL_4200500, EPI_ISL_4200525, EPI_ISL_4200527, EPI_ISL_4200546, EPI_ISL_4200572, EPI_ISL_4200586, EPI_ISL_4200596, EPI_ISL_4200598, EPI_ISL_4200602, EPI_ISL_4200621, EPI_ISL_4200629, EPI_ISL_4200635, EPI_ISL_4200652, EPI_ISL_4200655, EPI_ISL_4200710, EPI_ISL_4232687, EPI_ISL_4232688, EPI_ISL_4232701, EPI_ISL_4232711, EPI_ISL_4232712, EPI_ISL_4232716, EPI_ISL_4232718, EPI_ISL_4232722, EPI_ISL_4232736, EPI_ISL_4232741, EPI_ISL_4232766, EPI_ISL_4232782, EPI_ISL_4232843, EPI_ISL_4232847, EPI_ISL_4232851, EPI_ISL_4232902, EPI_ISL_4232925, EPI_ISL_4232931, EPI_ISL_4233010, EPI_ISL_4233021, EPI_ISL_4233052, EPI_ISL_4233060, EPI_ISL_4233071, EPI_ISL_4233080, EPI_ISL_4233102, EPI_ISL_4233130, EPI_ISL_4233134, EPI_ISL_4233208, EPI_ISL_4233209, EPI_ISL_4233226, EPI_ISL_4233276, EPI_ISL_4233300, EPI_ISL_4256634, EPI_ISL_4256681, EPI_ISL_4256712, EPI_ISL_4256725, EPI_ISL_4257995, EPI_ISL_4258000, EPI_ISL_4258023, EPI_ISL_4258024, EPI_ISL_4258058, EPI_ISL_4258099, EPI_ISL_4258100, EPI_ISL_4258101, EPI_ISL_4258117, EPI_ISL_4258122, EPI_ISL_4258127, EPI_ISL_4258136, EPI_ISL_4258140, EPI_ISL_4258150, EPI_ISL_4258181, EPI_ISL_4258197, EPI_ISL_4258205, EPI_ISL_4258216, EPI_ISL_4258221, EPI_ISL_4258245, EPI_ISL_4258249, EPI_ISL_4258254, EPI_ISL_4258258, EPI_ISL_4258268, EPI_ISL_4258273, EPI_ISL_4258293, EPI_ISL_4258305, EPI_ISL_4258307, EPI_ISL_4258320, EPI_ISL_4258349, EPI_ISL_4303421, EPI_ISL_4303436, EPI_ISL_4303437, EPI_ISL_4303451, EPI_ISL_4303481, EPI_ISL_4303482, EPI_ISL_4303528, EPI_ISL_4303532, EPI_ISL_4303544, EPI_ISL_4303547, EPI_ISL_4303554, EPI_ISL_4303558, EPI_ISL_4303559, EPI_ISL_4303568, EPI_ISL_4303573, EPI_ISL_4303578, EPI_ISL_4303590, EPI_ISL_4303605, EPI_ISL_4303624, EPI_ISL_4303625, EPI_ISL_4303658, EPI_ISL_4354720, EPI_ISL_4354735, EPI_ISL_4354755, EPI_ISL_4354776, EPI_ISL_4354817, EPI_ISL_4354828, EPI_ISL_4354834, EPI_ISL_4354848, EPI_ISL_4354955, EPI_ISL_4354963, EPI_ISL_4355021, EPI_ISL_4355118, EPI_ISL_4355181, EPI_ISL_4355283, EPI_ISL_4355340, EPI_ISL_4355452, EPI_ISL_4355456, EPI_ISL_4355488, EPI_ISL_4355627, EPI_ISL_4355633, EPI_ISL_4355647, EPI_ISL_4355658, EPI_ISL_4355659, EPI_ISL_4355668, EPI_ISL_4355675, EPI_ISL_4355690, EPI_ISL_4355693, EPI_ISL_4355714, EPI_ISL_4355720, EPI_ISL_4355752, EPI_ISL_4355759, EPI_ISL_4355768, EPI_ISL_4355794, EPI_ISL_4355800, EPI_ISL_4355809, EPI_ISL_4355833, EPI_ISL_4355864, EPI_ISL_4355885, EPI_ISL_4355928, EPI_ISL_4394097, EPI_ISL_4447627, EPI_ISL_4447653, EPI_ISL_4447655, EPI_ISL_4447671, EPI_ISL_4447706, EPI_ISL_4447721, EPI_ISL_4447726, EPI_ISL_4447758, EPI_ISL_4447806, EPI_ISL_4447822, EPI_ISL_4447824, EPI_ISL_4447841, EPI_ISL_4447872, EPI_ISL_4447881, EPI_ISL_4447889, EPI_ISL_4448077, EPI_ISL_4448147, EPI_ISL_4448227, EPI_ISL_4448243, EPI_ISL_4448267, EPI_ISL_4448308, EPI_ISL_4448327, EPI_ISL_4448333, EPI_ISL_4448338, EPI_ISL_4448356, EPI_ISL_4448361, EPI_ISL_4448368, EPI_ISL_4448389, EPI_ISL_4448398, EPI_ISL_4448433, EPI_ISL_4448474, EPI_ISL_4448498, EPI_ISL_4448581, EPI_ISL_4448587, EPI_ISL_4448605, EPI_ISL_4448622, EPI_ISL_4448657, EPI_ISL_4448664, EPI_ISL_4448669, EPI_ISL_4448677, EPI_ISL_4448679, EPI_ISL_4448736, EPI_ISL_4448770, EPI_ISL_4448808, EPI_ISL_4448811, EPI_ISL_4448826, EPI_ISL_4448839, EPI_ISL_4448854, EPI_ISL_4448858, EPI_ISL_4448882, EPI_ISL_4448898, EPI_ISL_4448899, EPI_ISL_4448919, EPI_ISL_4448938, EPI_ISL_4448990, EPI_ISL_4448992, EPI_ISL_4449013, EPI_ISL_4449014, EPI_ISL_4449021, EPI_ISL_4449032, EPI_ISL_4449039, EPI_ISL_4449046, EPI_ISL_4449049, EPI_ISL_4449075, EPI_ISL_4449090, EPI_ISL_4449122, EPI_ISL_4449129, EPI_ISL_4473488, EPI_ISL_4473489, EPI_ISL_4473491, EPI_ISL_4473511, EPI_ISL_4473524, EPI_ISL_4473601, EPI_ISL_4473640, EPI_ISL_4473651, EPI_ISL_4473676, EPI_ISL_4473698, EPI_ISL_4473713, EPI_ISL_4473747, EPI_ISL_4473768, EPI_ISL_4473775, EPI_ISL_4473812, EPI_ISL_4473877, EPI_ISL_4520857, EPI_ISL_4520875, EPI_ISL_4520876, EPI_ISL_4520889, EPI_ISL_4520927, EPI_ISL_4520928, EPI_ISL_4520955, EPI_ISL_4520959, EPI_ISL_4520961, EPI_ISL_4520970, EPI_ISL_4520979, EPI_ISL_4520986, EPI_ISL_4520998, EPI_ISL_4521013, EPI_ISL_4521017, EPI_ISL_4521035, EPI_ISL_4521036, EPI_ISL_4521047, EPI_ISL_4521068, EPI_ISL_4521076, EPI_ISL_4521086, EPI_ISL_4521095, EPI_ISL_4521129, EPI_ISL_4521131, EPI_ISL_4521138, EPI_ISL_4521141, EPI_ISL_4521166, EPI_ISL_4521173, EPI_ISL_4521193, EPI_ISL_4521225, EPI_ISL_4521243, EPI_ISL_4551362, EPI_ISL_4551377, EPI_ISL_4551410, EPI_ISL_4551487, EPI_ISL_4551500, EPI_ISL_4551532 | | | |
| see above | Department of Bacteria, Parasites and Fungi, Statens Serum Institut, Copenhagen, Denmark | Statens Serum Institut Bioinformatics and Microbial Genomics | Danish Covid-19 Genome Consortium |
| EPI_ISL_4489328, EPI_ISL_4492662, EPI_ISL_4492667 | Department of Clinical Microbiology | GIGA Medical Genomics | Bouchra Boujemla; Claire Gourzonès; Cécile Meex; Keith Durkin; Laurent Gillet; Maria Artesi; Marie-Pierre Hayette; Nadine Cambisano; Nathalie Renotte; Olivier Ek; Sébastien Bontems; Vincent Bours |
| EPI_ISL_4303431, EPI_ISL_4303574 | Department of Clinical Microbiology and Center for Genomic Medicine, Rigshospitalet, Copenhagen, Denmark | Statens Serum Institut Bioinformatics and Microbial Genomics | Danish Covid-19 Genome Consortium |
| EPI_ISL_4256716, EPI_ISL_4256717, EPI_ISL_4256724, EPI_ISL_4447666, EPI_ISL_4449065, EPI_ISL_4449082 | Department of Clinical Microbiology, Odense University Hospital, Odense, Denmark | Statens Serum Institut Bioinformatics and Microbial Genomics | Danish Covid-19 Genome Consortium |
| EPI_ISL_4398123, EPI_ISL_4398251 | Department of Medical Microbiology - section Molde, Molde Hospital | Norwegian Institute of Public Health, Department of Virology | Atiya R Ali; Debech Nadia; Engebretsen Serina Beate; Garcia Llorente Ignacio; Hilde Elshaug; Hilde Vollan; Jon Bråte; Kamilla Heddeland Instefjord; Karoline Bragstad; Kathrine Stene-Johansen; Line Victoria Moen; Marie Paulsen Madsen; Olav Hungnes; Pedersen Benedikte Nevjen; Rasmus Riis Kopperud |
| EPI_ISL_4505349 | Dept. of Microbiology and Infection Control, Akershus University Hospital HF | Dept. of Microbiology and Infection Control, Akershus University Hospital HF | Alexander Hesselberg Løvestad; Hege Vangstein Aamot |
| EPI_ISL_4446887 | Diagnosticum - Labor Neukirchen | Robert Koch Institute | |
| EPI_ISL_4413063 | Dianalabs SA | Genesupport | Geraldine Jost; Katia Jaton; Nadia Liassine; Tanguy ARAUD |
| EPI_ISL_4204382, EPI_ISL_4204388, EPI_ISL_4204396, EPI_ISL_4633367, EPI_ISL_4633734 | Division of Emerging Infectious Diseases, Bureau of Infectious Diseases Diagnosis Control, Korea Disease Control and Prevention Agency | Division of Emerging Infectious Diseases, Bureau of Infectious Diseases Diagnosis Control, Korea Disease Control and Prevention Agency | Ae Kyung Park; Chae Young Lee; Eun-Jin Kim; Heui Man Kim; Il-Hwan Kim; Jeong-Ah Kim |
| EPI_ISL_4482941 | Dutch COVID-19 response team | Erasmus Medical Center | Anne van der Linden; Annemiek van der Eijk; Bas Oude Munnink; Corine GeurtsvanKessel; David Nieuwenhuijse; Emmanuelle Munger; Irina Chestakova; Marion Koopmans; Marjan Boter; Reina Sikkema; Richard Molenkamp; on behalf of the Dutch national COVID-19 response team. |
| EPI_ISL_4399875, EPI_ISL_4400273, EPI_ISL_4400762, EPI_ISL_4400765, EPI_ISL_4400807, EPI_ISL_4401112, EPI_ISL_4401200, EPI_ISL_4401891, EPI_ISL_4402178, EPI_ISL_4544165, EPI_ISL_4544177, EPI_ISL_4544182, EPI_ISL_4544183, EPI_ISL_4544195, EPI_ISL_4544213, EPI_ISL_4544381, EPI_ISL_4544392, EPI_ISL_4544413, EPI_ISL_4544541, EPI_ISL_4544590, EPI_ISL_4544615, EPI_ISL_4544653, EPI_ISL_4544791, EPI_ISL_4544792, EPI_ISL_4544815, EPI_ISL_4544818, EPI_ISL_4544840 | | | |
| see above | Dutch COVID-19 response team | National Institute for Public Health and the Environment (RIVM) | Adam Meijer; AnneMarie van den Brandt; Annelies Kroneman; Bas van der Veer; Chantal Reusken; Dennis Schmitz; Dirk Eggink; Euníce Then; Florian Zwagemaker; Harry Vennema; Ivo van Walle; Jeroen Cremer; Karim Hajji; Kim Freriks; Lisa Wijsman; Lynn Aarts; Ryanne Jaarsma; Sanne Bos; Sharon van den Brink; Stijn van Rossum; on behalf of the national COVID-19 response team |
| EPI_ISL_4413146 | EHNV | Laboratory of genomics and metagenomics | Claire Bertelli; Damien Jacot; Gilbert Greub; Sébastien Aeby; Trestan Pillonel |
| EPI_ISL_4607088 | EXALAB LE HAILLAN | CNR Virus des Infections Respiratoires - France SUD | Antonin Bal; Bruno Lina; Gregory Destras; Gwendolyne Burfin; Hadrien Regue; Laurence Josset; Martine Valette; Quentin Semanas |
| EPI_ISL_4615436 | Eurofins LifeCodexx GmbH | Robert Koch Institute | |
| EPI_ISL_4042813, EPI_ISL_4042845, EPI_ISL_4042938, EPI_ISL_4228730, EPI_ISL_4616205 | Eurofins MVZ Labor Gelsenkirchen | Robert Koch Institute | |
| EPI_ISL_4543176 | Fundacio Althaia-Manresa | Banc de Sang i Teixits | Carlos Hobeich; Francisco Vidal; Irene Corrales; Lorena Ramírez; Maria Glòria Soria; Natália Comes; Nina Borràs; Noemí Gonzalez; Silvia Sauleda |
| EPI_ISL_4269603 | Gravity Diagnostics, LLC | Gravity Diagnostics, LLC | Gravity Diagnostics |
| EPI_ISL_4201574 | HOSPITAL UNIVERSITARI DE BELLVITGE | Microbiology Department | Aida Gonzalez-Diaz; Carmen Ardanuy; Jordi Camara; Jordi Niubó; Laura Calatayud; M Angeles Domínguez; Sara Marti; Yolanda Hernandez |
| EPI_ISL_4547936, EPI_ISL_4547942 | HOSPITAL UNIVERSITARIO CENTRAL DE ASTURIAS | Laboratorio de Virología HUCA | ; Alba L; Alvarez-Arguelles ME; Boga JA; Costales I; Coto E; González-Alba JM; Gómez de Oña J; Martín-Rodríguez G; Melón S; Perez-Martínez Z; Rojo S; Sandoval M |
| EPI_ISL_4137153, EPI_ISL_4137167, EPI_ISL_4137216, EPI_ISL_4350099, EPI_ISL_4350100, EPI_ISL_4350137, EPI_ISL_4350170, EPI_ISL_4350213, EPI_ISL_4350254, EPI_ISL_4550889, EPI_ISL_4550891, EPI_ISL_4550893, EPI_ISL_4550897 | | | |
| see above | HOSPITAL UNIVERSITARIO SON ESPASES | HOSPITAL UNIVERSITARIO SON ESPASES | Dr. Antonio Oliver; Dr. Carla López-Causapé; Dr. Gabriel Cabot; Hospital Universitario Son Espases; on behalf of Servicio de Microbiología |
| EPI_ISL_4066928 | Health Services Laboratories | Wellcome Sanger Institute for the COVID-19 Genomics UK (COG-UK) Consortium | Cordelia Langford; David K. Jackson; Dominic Kwiatkowski; Ewan Harrison; Health Services Laboratories and Alex Alderton; Ian Johnston; Jeffrey Barrett; John Sillitoe on behalf of the Wellcome Sanger Institute COVID-19 Surveillance Team; Roberto Amato; Sonia Goncalves |
| EPI_ISL_4204689 | Heilig hart Lier | Imelda Ziekenhuis | Dagmar Obbels; Hanne Valgaeren; Johan Frans |
| EPI_ISL_4461630, EPI_ISL_4461780, EPI_ISL_4503709, EPI_ISL_4503807, EPI_ISL_4545361, EPI_ISL_4545365 | Hospital | National Reference Center for Viruses of Respiratory Infections, Institut Pasteur, Paris | Alexandra Ducancelle; Angela Brisebarre; Camille Capel; Christophe Malabat; Corinne Maufrais; Etienne Simon-Lorière; Frédéric Lemoine; Isabelle Gros; Julien Fumey; JéRome Guinard; L. Courdavault; Louise Lefrançois; Marianne Burgard; Marion Barbet; Maud Vanpeene; Méline Bizard; Slim El Khiari; Slim El-Khiari; Sylvie Behillil; Sylvie Van der Werf; Vincent Enouf |
| EPI_ISL_4395984, EPI_ISL_4395988 | Hospital General Universitario Gregorio Marañón | Hospital General Universitario Gregorio Marañón | Cristina Rodriguez-Grande; Darío García de Viedma; Julia Suárez; Laura Pérez-Lago; Marta Herranz Martin; Patricia Muñoz; Pedro Sola Campoy; Pilar Catalán; Sergio Buenestado Serrano; Victor Manuel de la Cueva |
| EPI_ISL_4578040 | Hospital General de Valdepeñas | Hospital General Universitario de Ciudad Real | Cristina Colmenarejo; José Martínez-Alarcón; Lidia García-Agudo; Marta Torres-Narbona; Soledad Illescas Fernández-Bermejo |
| EPI_ISL_4548976 | Hospital Universitari Arnau de Vilanova | Hospital Universitari Vall d'Hebron - Vall d'Hebron Institut de Recerca | Alejandra González-Sánchez; Andrés Antón; Ariadna Rando; Carla Castillo; Cristina Andrés; Damir Garcia-Cehic; Josep Quer; Juliana Esperalba; Karen García; Maria Carmen Martin; Maria Gema Codina; Maria Piñana; Rodrigo Vásquez; Tomàs Pumarola |
| EPI_ISL_4444201, EPI_ISL_4444215, EPI_ISL_4444236 | Hospital Universitari Joan XXIII | Hospital Universitari Joan XXIII | Carla Martín; Clara Benavent; Cristina Gutiérrez; Ester Picó; Gemma Recio; Margarida Terrón |
| EPI_ISL_4112968 | Hospital Universitari Vall d'Hebron - | Hospital Universitari Vall d'Hebron - | Alejandra González-Sánchez; Andrés Antón; Ariadna Rando; Carla Castillo; Cristina Andrés; Damir Garcia-Cehic; Josep Quer; Juliana Esperalba; Karen García; Maria Carmen Martin; Maria Gema Codina; Maria Piñana; Rodrigo Vásquez; Tomàs Pumarola |

| | | | |
|---|---|---|---|
| | Vall d'Hebron Institut de Recerca | Vall d'Hebron Institut de Recerca | |
| EPI_ISL_4578048 | Hospital Virgen De Altagracia | Hospital General Universitario de Ciudad Real | Cristina Colmenarejo; José Martínez-Alarcón; Lidia García-Agudo; Marta Torres-Narbona; Soledad Illescas Fernández-Bermejo |
| EPI_ISL_4611670, EPI_ISL_4611682 | IMD Labor Frankfurt | Robert Koch Institute | |
| EPI_ISL_4204707 | Imelda Ziekenhuis | Imelda Ziekenhuis | Dagmar Obbels; Hanne Valgaeren; Johan Frans |
| EPI_ISL_4632119 | Institut de Chimie Clinique Sarl | Laboratory of genomics and metagenomics | Claire Bertelli; Damien Jacot; Gilbert Greub; Sébastien Aeby; Trestan Pillonel |
| EPI_ISL_4269656, EPI_ISL_4269700, EPI_ISL_4486270, EPI_ISL_4625133 | Institute for Infectious Diseases | Institute for Infectious Diseases, University of Bern | Alban Ramette; Christian Baumann; Cora Sägesser; Franziska Suter-Riniker; Loïc Borcard; Miguel A Terrazos Miani; Nicole Liechti; Pascal Bittel; Peter Keller; Sonja Gempeler; Stefan Neuenschwander; Stephen L Leib |
| EPI_ISL_4185848, EPI_ISL_4185850, EPI_ISL_4185853, EPI_ISL_4185925, EPI_ISL_4185961, EPI_ISL_4186072, EPI_ISL_4186111, EPI_ISL_4270069, EPI_ISL_4270074, EPI_ISL_4270075, EPI_ISL_4634191 | | | |
| see above | Jessa | Jessa | Marijke Raymaekers et al. on behalf of the Jessa_cmdLab; Severine Berden et al. on behalf of the Jessa_cmdLab |
| EPI_ISL_4365231, EPI_ISL_4365236, EPI_ISL_4365237, EPI_ISL_4365239, EPI_ISL_4365240, EPI_ISL_4365255, EPI_ISL_4365258, EPI_ISL_4365294, EPI_ISL_4365412, EPI_ISL_4572020, EPI_ISL_4572041 | | | |
| see above | KU Leuven, Rega Institute, Clinical and Epidemiological Virology | KU Leuven, Rega Institute, Clinical and Epidemiological Virology | Bert Vanmechelen; Joan Marti-Carerras; Piet Maes; Tony Wawina-Bokalanga |
| EPI_ISL_4175151, EPI_ISL_4175152, EPI_ISL_4330413 | Karolinska University Hospital Huddinge | Karolinska University Hospital | Annelie Bjerkner; Isak Sylvin; Jan Albert; Karolina Ininbergs; Lina Guerra Blomqvist; Lynda Eneh; Martin Ekman; Martina Wahlund; Robert Dyrdak; Sandra Broddesson; Tanja Normark; Tobias Allander; Valtteri Wirta; Zhibing Yun |
| EPI_ISL_4505720, EPI_ISL_4505721, EPI_ISL_4505735 | Klinisch Laboratorium ZNA | Klinisch Laboratorium ZNA | Verstrepen et al. |
| EPI_ISL_4606836, EPI_ISL_4606839, EPI_ISL_4606851, EPI_ISL_4606900 | LABORATOIRE ANABIO BERGSON | CNR Virus des Infections Respiratoires - France SUD | Antonin Bal; Bruno Lina; Gregory Destras; Gwendolyne Burfin; Hadrien Regue; Laurence Josset; Martine Valette; Quentin Semanas |
| EPI_ISL_4605757, EPI_ISL_4605817 | LABORATOIRE BIOESTEREL | CNR Virus des Infections Respiratoires - France SUD | Antonin Bal; Bruno Lina; Gregory Destras; Gwendolyne Burfin; Hadrien Regue; Laurence Josset; Martine Valette; Quentin Semanas |
| EPI_ISL_4605770, EPI_ISL_4605803, EPI_ISL_4605813, EPI_ISL_4605851, EPI_ISL_4605856, EPI_ISL_4605866, EPI_ISL_4606692 | | | |
| see above | LABORATOIRE CERBALLIANCE PLT VILLON | CNR Virus des Infections Respiratoires - France SUD | Antonin Bal; Bruno Lina; Gregory Destras; Gwendolyne Burfin; Hadrien Regue; Laurence Josset; Martine Valette; Quentin Semanas |
| EPI_ISL_4606759 | LABORATOIRE FORTE BIO DAX | CNR Virus des Infections Respiratoires - France SUD | Antonin Bal; Bruno Lina; Gregory Destras; Gwendolyne Burfin; Hadrien Regue; Laurence Josset; Martine Valette; Quentin Semanas |
| EPI_ISL_4606990, EPI_ISL_4606994, EPI_ISL_4607017 | LABORATOIRE LABAZUR | CNR Virus des Infections Respiratoires - France SUD | Antonin Bal; Bruno Lina; Gregory Destras; Gwendolyne Burfin; Hadrien Regue; Laurence Josset; Martine Valette; Quentin Semanas |
| EPI_ISL_4606061, EPI_ISL_4606065, EPI_ISL_4606075, EPI_ISL_4606077 | LABORATOIRE LBA JAYAN AGEN | CNR Virus des Infections Respiratoires - France SUD | Antonin Bal; Bruno Lina; Gregory Destras; Gwendolyne Burfin; Hadrien Regue; Laurence Josset; Martine Valette; Quentin Semanas |
| EPI_ISL_4605665 | LABORATOIRE UNILIANS DECINES | CNR Virus des Infections Respiratoires - France SUD | Antonin Bal; Bruno Lina; Gregory Destras; Gwendolyne Burfin; Hadrien Regue; Laurence Josset; Martine Valette; Quentin Semanas |
| EPI_ISL_4445602, EPI_ISL_4608964, EPI_ISL_4608994, EPI_ISL_4609014, EPI_ISL_4610987, EPI_ISL_4610997, EPI_ISL_4611044, EPI_ISL_4611045, EPI_ISL_4611143, EPI_ISL_4611185, EPI_ISL_4616773, EPI_ISL_4616801, EPI_ISL_4616864, EPI_ISL_4618249, EPI_ISL_4619228 | | | |
| see above | LADR Zentrallabor DR. Kramer & Kollegen Geesthacht | Robert Koch Institute | |
| EPI_ISL_4606523 | LBM DYNABIO CARREAU | CNR Virus des Infections Respiratoires - France SUD | Antonin Bal; Bruno Lina; Gregory Destras; Gwendolyne Burfin; Hadrien Regue; Laurence Josset; Martine Valette; Quentin Semanas |
| EPI_ISL_4606532, EPI_ISL_4606534 | LBM TRONQUIERES | CNR Virus des Infections Respiratoires - France SUD | Antonin Bal; Bruno Lina; Gregory Destras; Gwendolyne Burfin; Hadrien Regue; Laurence Josset; Martine Valette; Quentin Semanas |
| EPI_ISL_4053034 | LKO | Jessa | Marijke Raymaekers et al. on behalf of the Jessa_cmdLab |
| EPI_ISL_4525778 | Lab. Microbiologia e Virologia Cotugno A.O. dei Colli | TIGEM | Antonio Grimaldi Patrizia Annunziata Francesco Panariello Claudia Tiberio Teresa Giuliano Valentina Bouche Chiara Colantuono Lucio Di Filippo Anna Manfredi Marcello Salvi Antonio Limone Luigi Atripaldi Andrea Ballabio Davide Cacchiarelli |
| EPI_ISL_4224792, EPI_ISL_4624787 | LabKom - Labor Augsburg MVZ GmbH | Robert Koch Institute | |
| EPI_ISL_4614785, EPI_ISL_4614830, EPI_ISL_4614931 | LabKom - Labor Mainz MVZ GmbH | Robert Koch Institute | |
| EPI_ISL_4614594 | LabKom - MVZ Labor Bochum MLB GmbH | Robert Koch Institute | |
| EPI_ISL_4461444, EPI_ISL_4461654, EPI_ISL_4461709, EPI_ISL_4503745, EPI_ISL_4503760, EPI_ISL_4503762, EPI_ISL_4503833, EPI_ISL_4503879, EPI_ISL_4503881, EPI_ISL_4503931, EPI_ISL_4600659, EPI_ISL_4600670, EPI_ISL_4600671, EPI_ISL_4600724, EPI_ISL_4600821, EPI_ISL_4600844, EPI_ISL_4600846, EPI_ISL_4601056 | | | |
| see above | Labo Analyses Med | National Reference Center for Viruses of Respiratory Infections, Institut Pasteur, Paris | Angela Brisebarre; Anne Duterrail; Baptiste Moreira; Camille Capel; Christophe Malabat; Corinne Maufrais; Domitille Leman; Etienne Simon-Lorière; Frédéric Lemoine; Gilles Abs; Guillaume Aubin; Jean-FrançOis Comes; Julien Fumey; Leonard; Louise Lefrançois; Marion Barbet; Maud Vanpeene; Méline Bizard; Nabila Belhouachi; OphéLie Said-Delattre; Philippe Girard; Slim El Khiari; Slim El-Khiari; Sophie Chalmin; Stephanie Hainos; Sylvie Behillil; Sylvie Van der Werf; Vincent Enouf |
| EPI_ISL_4310262 | Labor Berlin Charité Vivantes GmbH / Institut für Virologie | Charité Universitätsmedizin Berlin, Institut für Virologie/Labor Berlin | Barbara Mühlemann; Christian Drosten; Christine Stephan; Peter Menzel; Rolf Schwarzer; Terry Jones; Victor M Corman |
| EPI_ISL_4230901, EPI_ISL_4230978, EPI_ISL_4231109, EPI_ISL_4231129, EPI_ISL_4231383, EPI_ISL_4231422, EPI_ISL_4444613, EPI_ISL_4444710, EPI_ISL_4444714, EPI_ISL_4444754, EPI_ISL_4444819, EPI_ISL_4444855, EPI_ISL_4444864, EPI_ISL_4444998, EPI_ISL_4609282, EPI_ISL_4609299, EPI_ISL_4609388, EPI_ISL_4609395, EPI_ISL_4609404, EPI_ISL_4609423, EPI_ISL_4609465, EPI_ISL_4609527, EPI_ISL_4614039, EPI_ISL_4614080, EPI_ISL_4614206, EPI_ISL_4614209 | | | |
| see above | Labor Dr. Wisplinghoff - Köln | Robert Koch Institute | |
| EPI_ISL_4224309, EPI_ISL_4437713, EPI_ISL_4437730, EPI_ISL_4439885, EPI_ISL_4442472, EPI_ISL_4446545, EPI_ISL_4611798, EPI_ISL_4612193, EPI_ISL_4615093, EPI_ISL_4615096, EPI_ISL_4615187, EPI_ISL_4615202, EPI_ISL_4615209, EPI_ISL_4615219 | | | |
| see above | Labor Mönchengladbach MVZ Dr. Stein + Kollegen GbR | Robert Koch Institute | |
| EPI_ISL_4617034 | Laborarztpraxis Osnabrück | Robert Koch Institute | |
| EPI_ISL_4543386, EPI_ISL_4545397, EPI_ISL_4634213 | Laboratori de Referencia de Catalunya | Laboratori de Referencia de Catalunya | Bellosillo B.; Canal M.; Hernandez JJ.; Padilla E.; Ramirez A.; Vilas A. |
| EPI_ISL_4483618, EPI_ISL_4538746 | Laboratory Corporation of America | Centers for Disease Control and Prevention Division of Viral Diseases, Pathogen Discovery | Amanda Douglas; Amanda Suchanek; Andrea Throop; Ayla Burns; Benjamin Rambo-Martin; Bobbi Croy; Brian Krueger; Brian Norvell; Christopher Gulvick; Christos Petropoulos; Clinton Paden; Craig Lukasik; Dakota Howard; Debbie Boles; Dhwani Batra; Duncan MacCannell; Eyad Almasri; Goran Stevovic; Howard Engler; Hrushikesh Deshmukh; Jake Humphrey; Jana Schroth; Jason Caravas; Joe Voshell; John Pruitt; Jonathan Meltzer; Jonathan Williams; Kimberly Wagner; Kristine Lacek; Lax Iyer; Lisa Pfefferle; Lyndon Tilson; Manoj Jain; Marcia Eisenberg; Mary Cristobal; Mary Williamson; Matthew Robinson; Matthew Schmerer; Michael Levandoski; Mike Sapeta; Mindy Nye; Minoo Agarwal; Mohan Kolli; Nuthawin Charoensri; Oren Cohen; Peter Cook; Prashant Gupta; Qian Zeng; Rama Ghatti; Scott Parker; Scott Ryan; Scott Sammons; Shatavia Morrison; Stanley Letovsky; Steven Ragan; Suresh Selvaraju; Susan Countryman; Susan Hicks; Suzanne Dale; Thomas Urban; Tim Kuphal; Tricia Zwiefelhofer; Tymeckia Kendall; Victoria Caban Figueroa; Vincent Drouillon; Yvette Unoarumhi |
| EPI_ISL_4312406, EPI_ISL_4312861, EPI_ISL_4313301, EPI_ISL_4313638, EPI_ISL_4314142, EPI_ISL_4314645 | Laboratory of Microbiology, ASST Ospedale di Circolo, Varese viale borri 57 21100 Varese, Italy | Laboratory of Microbiology, ASST Ospedale di Circolo, Varese viale borri 57 21100 Varese, Italy | Andreina Baj; Angelo Genoni; Daniela Dalla Gasperina; Daniele Focosi; Fabrizio Maggi; Federica Novazzi; Francesca Dragoferrante |
| EPI_ISL_4190649 | Lighthouse Lab in Alderley Park | Wellcome Sanger Institute for the COVID-19 Genomics UK (COG-UK) Consortium | Cordelia Langford; David K. Jackson; Dominic Kwiatkowski; Ewan Harrison; Ian Johnston; Jacquelyn Wynn; Jeffrey Barrett; John Sillitoe on behalf of the Wellcome Sanger Institute COVID-19 Surveillance Team; Mairead Hyland; Roberto Amato; Sonia Goncalves; The Lighthouse Lab in Alderley Park and Alex Alderton |
| EPI_ISL_4067974 | Lighthouse Lab in Glasgow | Wellcome Sanger Institute for the COVID-19 Genomics UK (COG-UK) Consortium | Anna Dominiczak and Alex Alderton; Carol Clugston; Cordelia Langford; David Gray; David K. Jackson; Dominic Kwiatkowski; Ewan Harrison; Harper VanSteenhouse; Ian Johnston; Jeffrey Barrett; John Sillitoe on behalf of the Wellcome Sanger Institute COVID-19 Surveillance Team; Roberto Amato; Sonia Goncalves; Yumi Kasai |

| | | | |
|---|---|---|---|
| EPI_ISL_4143330, EPI_ISL_4143847, EPI_ISL_4265161, EPI_ISL_4320666, EPI_ISL_4404310, EPI_ISL_4478842, EPI_ISL_4522086 | | | |
| see above | Lighthouse Lab in Milton Keynes | Wellcome Sanger Institute for the COVID-19 Genomics UK (COG-UK) Consortium | Cordelia Langford; David K. Jackson; Dominic Kwiatkowski; Ewan Harrison; Ian Johnston; Jeffrey Barrett; John Sillitoe on behalf of the Wellcome Sanger Institute COVID-19 Surveillance Team; Roberto Amato; Sonia Goncalves; The Lighthouse Lab in Milton Keynes and Alex Alderton |
| EPI_ISL_4622959 | Limbach - MVZ Labor Dr. Volkmann & Kollegen | Robert Koch Institute | |
| EPI_ISL_4235115 | MCL Medizinische Laboratorien, Hauptstandort Niederwangen | Institute for Infectious Diseases, University of Bern | Alban Ramette; Christian Baumann; Cora Sägesser; Franziska Suter-Riniker; Loïc Borcard; Miguel A Terrazos Miani; Nicole Liechti; Pascal Bittel; Peter Keller; Sonja Gempeler; Stefan Neuenschwander; Stephen L Leib |
| EPI_ISL_4437698, EPI_ISL_4612161, EPI_ISL_4624864 | MDI Limbach Berlin GmbH; MVZ Labor Berlin | Robert Koch Institute | |
| EPI_ISL_4606392, EPI_ISL_4606983 | MIRIALIS CLUSES BECHET | CNR Virus des Infections Respiratoires - France SUD | Antonin Bal; Bruno Lina; Gregory Destras; Gwendolyne Burfin; Hadrien Regue; Laurence Josset; Martine Valette; Quentin Semanas |
| EPI_ISL_4610160, EPI_ISL_4610208 | MVZ Dr. Eberhard & Partner Dortmund | Robert Koch Institute | |
| EPI_ISL_4609748 | MVZ Labor Dr. Limbach & Kollegen GbR | Robert Koch Institute | |
| EPI_ISL_4616604 | MVZ Labor Krone GbR | Robert Koch Institute | |
| EPI_ISL_4258880, EPI_ISL_4258911, EPI_ISL_4258915 | Medical Laboratories Duesseldorf | Center of Medical Microbiology, Virology, and Hospital Hygiene, University of Duesseldorf | Alexander Dilthey; Andreas Walker; Angelika Helmer; Christian Lange; Daniel Strelow; Jessica Nicolai; Jörg Timm; Klaus Pfeffer; Lisanna Hülse; Malte Kohns Vasconcelos; Maximilian Damagnez; Nadine Lübke; Tobias Wienemann; Torsten Houwaart |
| EPI_ISL_4225558, EPI_ISL_4225587, EPI_ISL_4439827, EPI_ISL_4439838, EPI_ISL_4439861, EPI_ISL_4609576, EPI_ISL_4609595, EPI_ISL_4609610, EPI_ISL_4612079, EPI_ISL_4612089 | | | |
| see above | Medizinische Laboratorien Düsseldorf | Robert Koch Institute | |
| EPI_ISL_4080639, EPI_ISL_4279179, EPI_ISL_4279181 | Microbiology Department - University Hospital Brussel | Microbiology Department - University Hospital Brussel | Florence Crombé; Oriane Soetens; Thomas Demuyser |
| EPI_ISL_4172014, EPI_ISL_4172034, EPI_ISL_4172083, EPI_ISL_4303778, EPI_ISL_4303846, EPI_ISL_4359775, EPI_ISL_4359924, EPI_ISL_4359926, EPI_ISL_4359929, EPI_ISL_4359985, EPI_ISL_4533573, EPI_ISL_4533581, EPI_ISL_4533590, EPI_ISL_4533597, EPI_ISL_4533610, EPI_ISL_4533783, EPI_ISL_4533784, EPI_ISL_4533861 | | | |
| see above | Microbiology Department, Laboratori Clínic Metropolitana Nord. Hospital Universitari Germans Trias i Pujol | Can Ruti SARS-CoV-2 Sequencing Hub (HUGTiP/IrsiCaixa/IGTP) | Alexia París; Anna Not; Antoni E Bordoy; Bonaventura Clotet; Cristina Casañ; David Panisello; Francesc Catala-Moll; Gemma Clara; Ignacio Blanco; Laia Soler; Lauro Sumoy; Marc Noguera-Julian; Maria Casadellà; Mariona Parera; Mercedes Guerrero; Montserrat Giménez; Pere-Joan Cardona; Pilar Armengol; Roger Paredes; Verónica Saludes; and Elisa Martró on behalf of the Can Ruti SARS-CoV-2 Sequencing Hub |
| EPI_ISL_4416774 | Microvida | Microvida | Jaco Verweij; Joep Stohr; Suzan D. Pas |
| EPI_ISL_4429345 | Ministry of Health Turkey | Ministry of Health Turkey | Fatma Bayrakdar; Gülay Korukluoğlu; Süleyman Yalcin; Yasemin Coşgun |
| EPI_ISL_4199096 | NYU Langone Health | Departments of Pathology and Medicine, New York University School of Medicine | Adriana Heguy; Christian Marier; Dacia Dimartino; Emily Guzman; Gael Westby; Guiqing Wang; Paolo Cotzia; Paul Zappile; Peter Meyn; Sitharam Ramaswami; Yutong Zhang |
| EPI_ISL_4195076, EPI_ISL_4195098, EPI_ISL_4195102, EPI_ISL_4195109, EPI_ISL_4195111, EPI_ISL_4195113, EPI_ISL_4195115, EPI_ISL_4515925, EPI_ISL_4515950, EPI_ISL_4516317 | | | |
| see above | National Platform bis UMONS/Jolimont | National Platform bis UMONS/Jolimont | Eric Tarantino; Florian Juszczak; Gautier Detry; Guillaume Bayon-Vicente; Laetitia Gheysen; Ruddy Wattiez |
| EPI_ISL_4604584 | ORIAPOLE | CNR Virus des Infections Respiratoires - France SUD | Antonin Bal; Bruno Lina; Gregory Destras; Gwendolyne Burfin; Hadrien Regue; Laurence Josset; Martine Valette; Quentin Semanas |
| EPI_ISL_4315062 | Originating lab: Wales Specialist Virology Centre Sequencing lab: Pathogen Genomics Unit | Public Health Wales Microbiology Cardiff Wales Specialist Virology Centre | Alec Birchley; Alexander Adams; Amy Gaskin; Angela Marchbank; Bree Gatica-Wilcox; Catherine Moore; Jason Coombes; Joanne Watkins; Joel Southgate; Johnathan Evans; Laura Gifford; Lauren Gilbert; Lee Graham; Malorie Perry; Matthew Bull; Nicole Pacchiarini; Sally Corden; Sara Kumziene-Summerhayes; Sara Rey; Sarah Taylor; Simon Cottrell; Sophie Jones; Tom Connor |
| EPI_ISL_4416995 | Osp. S.Pertini | INMI Lazzaro Spallanzani IRCCS | A Di Caro; B Bartolini; CEM Gruber; E Giombini; F Messina; F Santini; G Bonfiglio; M Rueca; MR Capobianchi; O Butera |
| EPI_ISL_4492287 | PAMM | Laboratory of Medical Microbiology and Pathology, PAMM | Christel van Herk; Inge Briels; Jeroen van de Bovenkamp; Maaike Broeders; Pleunie van Alphen |
| EPI_ISL_4546886 | Pandemic Response Lab - NYC | Pandemic Response Lab, R&D | Alex Carpio; Cybill del Castillo; Dylan Law; Haiping Hao; Henry Lee; Isabel Fernandez Escapa; Jon Laurent; Melissa Hopkins; Michael Hammerling; Pradeep Bugga; Shinyoung Clair Kang; Sol Rey; William Ward |
| EPI_ISL_4050677, EPI_ISL_4050685, EPI_ISL_4050691, EPI_ISL_4071905, EPI_ISL_4071909, EPI_ISL_4186337, EPI_ISL_4186480, EPI_ISL_4219262, EPI_ISL_4261670, EPI_ISL_4261688, EPI_ISL_4261689, EPI_ISL_4263086, EPI_ISL_4419282, EPI_ISL_4419283, EPI_ISL_4633216, EPI_ISL_4633218, EPI_ISL_4633222 | | | |
| see above | Plateforme de testing Namuroise | Plateforme de testing Namuroise | Degosserie Jonathan; Demars Aurore; Denis Olivier; Gilliard Nicolas; Lesly Nyinkeu Kemamen; Maschietto Céline; Mullier François; Nicolas Gilliard; Nobis Chloé; Nyinkeu Kemamen Lesly; Otto Gaetan |
| EPI_ISL_4059368, EPI_ISL_4059376, EPI_ISL_4059389, EPI_ISL_4211035, EPI_ISL_4211050, EPI_ISL_4211069, EPI_ISL_4211125, EPI_ISL_4211127, EPI_ISL_4212599, EPI_ISL_4212606, EPI_ISL_4212613, EPI_ISL_4212618, EPI_ISL_4275299, EPI_ISL_4275346, EPI_ISL_4392335, EPI_ISL_4392346, EPI_ISL_4392376, EPI_ISL_4392384, EPI_ISL_4392385, EPI_ISL_4392392, EPI_ISL_4505549, EPI_ISL_4505634 | | | |
| see above | Platform BIS UZA/UAntwerpen | Labo Klinische Biologie, UZA | Basil Britto Xavier; Christine Lammens; Herman Goossens; Ines Verbesselt; Jasmine Coppens; Kathleen Holemans; Marie Le Mercier; Veerle Matheeussen |
| EPI_ISL_4106050, EPI_ISL_4413089 | Polyanalytic SA | Genesupport | Geraldine Jost; Katia Jaton; Nadia Liassine; Tanguy ARAUD |
| EPI_ISL_4548550 | Pracownia Diagnostyki Molekularnej COVID-19 Wojewódzkiego Szpitala Specjalistycznego we Wrocławiu, Ośrodka Badawczo-Rozwojowego | Wojewodzka Stacja Sanitarno-Epidemiologiczna w Gorzowie Wielkopolskim | Elżbieta Justyńska; Klaudia Kobendza-Wlodarczak; Patrycja Faberska and Marek Magol; Renata Siegel |
| EPI_ISL_4307554, EPI_ISL_4307796, EPI_ISL_4307797, EPI_ISL_4307854 | Quadram Institute Bioscience | COVID-19 Genomics UK (COG-UK) Consortium | Alexander J Trotter; Alison E. Mather; Alp Aydin; Ana P. Tedim; Anastasia Kolyva; Andrew Bell; Andrew J. Page; Christopher Jeanes; Claire Stuart; Dave J. Baker; Ebenezer Foster-Nyarko; Gemma L. Kay; John Wain; Justin O'Grady; Leonardo de Oliveira Martins; Lewis G. Spurgin; Lindsay Coupland; Lizzie Meadows; Luke Bedford; Maria Diaz; Mark Webber; Martin Lott; Muhammed Yasir; Nabil-Fareed Alikhan; Ngozi Elumogo; Nicholas M. Thomson; Rachael Stanley; Rachel Gilroy; Reenesh Prakash; Rose K Davidson; Samir Dervisevic; Samuel Bloomfield; Sophie J. Prosolek; Steven Rudder; Thanh Le-Viet |
| EPI_ISL_4606800, EPI_ISL_4606827, EPI_ISL_4606926 | REUNILAB | CNR Virus des Infections Respiratoires - France SUD | Antonin Bal; Bruno Lina; Gregory Destras; Gwendolyne Burfin; Hadrien Regue; Laurence Josset; Martine Valette; Quentin Semanas |
| EPI_ISL_4117688, EPI_ISL_4117752, EPI_ISL_4117782, EPI_ISL_4117796, EPI_ISL_4117835, EPI_ISL_4118705, EPI_ISL_4118738, EPI_ISL_4118766, EPI_ISL_4118776, EPI_ISL_4118784, EPI_ISL_4118794, EPI_ISL_4121346, EPI_ISL_4121407, EPI_ISL_4121526, EPI_ISL_4121702, EPI_ISL_4122029, EPI_ISL_4122339, EPI_ISL_4122786, EPI_ISL_4122794, EPI_ISL_4123735, EPI_ISL_4308371, EPI_ISL_4308419, EPI_ISL_4308549, EPI_ISL_4308600, EPI_ISL_4309009, EPI_ISL_4309027, EPI_ISL_4309050, EPI_ISL_4309239, EPI_ISL_4311857, EPI_ISL_4311965, EPI_ISL_4529926, EPI_ISL_4530106, EPI_ISL_4530312, EPI_ISL_4530472, EPI_ISL_4530672, EPI_ISL_4530871, EPI_ISL_4531384, EPI_ISL_4531411, EPI_ISL_4531570, EPI_ISL_4531634, EPI_ISL_4531847, EPI_ISL_4531848 | | | |
| see above | Respiratory Virus Unit, Microbiology Services Colindale, Public Health England | COVID-19 Genomics UK (COG-UK) Consortium | PHE Covid Sequencing Team |
| EPI_ISL_4506526 | SARS-CoV-2 Sequencing Castilla y Leon-Spain Consortium | SARS-CoV-2 Sequencing Castilla y Leon-Spain Consortium | Antonio Orduña-Domingo; Carlos Fuster Foz; Carmen Aldea-Mansilla; Carmen Gimeno Crespo; David Abad; Gabriel March Rosello; Gregoria Megías Lobón; Jose María Eiros Bouza; M. Isabel Fernandez-Natal; Marta Dominguez-Gil; Marta Hernandez; María Antonia García Castro; Mª Fe Brezmes-Valdivieso; Noelia Arenal Andrés; Silvia Rojo; Sonsoles Garcinuño Pérez |
| EPI_ISL_4489273, EPI_ISL_4489286, EPI_ISL_4489314, EPI_ISL_4489342, EPI_ISL_4489346, EPI_ISL_4489447, EPI_ISL_4489633, EPI_ISL_4489643, EPI_ISL_4489700, EPI_ISL_4489715, EPI_ISL_4489749, EPI_ISL_4492577, EPI_ISL_4492629, EPI_ISL_4492656, EPI_ISL_4600156, EPI_ISL_4600175 | | | |
| see above | SYNLAB | GIGA Medical Genomics | Bouchra Boujemla; Claire Gourzonès; Cécile Meex; Keith Durkin; Laurent Gillet; Maria Artesi; Marie-Pierre Hayette; Nadine Cambisano; Nathalie Renotte; Olivier Ek; Sébastien Bontems; Vincent Bours |
| EPI_ISL_4438679, EPI_ISL_4613250, EPI_ISL_4613319, EPI_ISL_4613447, EPI_ISL_4613740, EPI_ISL_4613878 | SYNLAB MVZ Leinfelden-Echterdingen | Robert Koch Institute | |
| EPI_ISL_4045353, EPI_ISL_4222245, EPI_ISL_4222774, EPI_ISL_4222979, EPI_ISL_4222989, EPI_ISL_4223602, EPI_ISL_4223696, EPI_ISL_4223717, EPI_ISL_4223982, EPI_ISL_4224007, EPI_ISL_4224179, EPI_ISL_4228831, EPI_ISL_4228880, EPI_ISL_4229153, EPI_ISL_4229239, EPI_ISL_4229294, EPI_ISL_4229320, EPI_ISL_4229322, EPI_ISL_4229584, EPI_ISL_4230184, EPI_ISL_4230357, EPI_ISL_4230754, EPI_ISL_4436802, EPI_ISL_4436836, EPI_ISL_4436854, EPI_ISL_4437147, EPI_ISL_4438664, EPI_ISL_4442646, EPI_ISL_4442730, EPI_ISL_4442876, EPI_ISL_4443215, EPI_ISL_4443327, EPI_ISL_4443335, EPI_ISL_4443369, EPI_ISL_4443719, EPI_ISL_4443757, EPI_ISL_4444007, EPI_ISL_4444011, EPI_ISL_4446099, EPI_ISL_4446102, EPI_ISL_4446158, EPI_ISL_4446201, EPI_ISL_4446202, EPI_ISL_4446251, EPI_ISL_4446375, EPI_ISL_4446387, EPI_ISL_4610223, EPI_ISL_4610276, EPI_ISL_4610361, EPI_ISL_4610607, EPI_ISL_4610749, EPI_ISL_4613019, EPI_ISL_4613046, EPI_ISL_4613106, EPI_ISL_4613476, EPI_ISL_4613589, EPI_ISL_4613609, EPI_ISL_4615551, EPI_ISL_4615632, EPI_ISL_4615719, EPI_ISL_4615887, EPI_ISL_4615895, EPI_ISL_4615912, EPI_ISL_4617676, EPI_ISL_4617936, EPI_ISL_4617945, EPI_ISL_4617973, EPI_ISL_4617999 | | | |
| see above | SYNLAB MVZ Leverkusen | Robert Koch Institute | |
| EPI_ISL_4043757, EPI_ISL_4044227, EPI_ISL_4044751, EPI_ISL_4044832, EPI_ISL_4045014, EPI_ISL_4045042, EPI_ISL_4222291, EPI_ISL_4224095, EPI_ISL_4226258, EPI_ISL_4226262, EPI_ISL_4226424, EPI_ISL_4230010, EPI_ISL_4230013, EPI_ISL_4230174, EPI_ISL_4230463, EPI_ISL_4443036, EPI_ISL_4443258, EPI_ISL_4610401 | | | |
| see above | SYNLAB MVZ Trier | Robert Koch Institute | |
| EPI_ISL_4438496, EPI_ISL_4613687, EPI_ISL_4616062, EPI_ISL_4616067 | SYNLAB MVZ Weiden | Robert Koch Institute | |
| EPI_ISL_4441476, | Sonic - Bioscientia - MVZ Labor Saar | Robert Koch Institute | |

| | | | |
|---|---|---|---|
| EPI_ISL_4441501, EPI_ISL_4611382, EPI_ISL_4614431, EPI_ISL_4614863 | GmbH | | |
| EPI_ISL_4108518, EPI_ISL_4108522, EPI_ISL_4108542, EPI_ISL_4108563, EPI_ISL_4108567, EPI_ISL_4108572, EPI_ISL_4108584, EPI_ISL_4108613, EPI_ISL_4108622, EPI_ISL_4335864, EPI_ISL_4335899, EPI_ISL_4335914, EPI_ISL_4335922, EPI_ISL_4335956, EPI_ISL_4335992, EPI_ISL_4336024, EPI_ISL_4336032, EPI_ISL_4336056, EPI_ISL_4336099, EPI_ISL_4336110, EPI_ISL_4336112, EPI_ISL_4336120, EPI_ISL_4336166, EPI_ISL_4336179, EPI_ISL_4336207, EPI_ISL_4336216, EPI_ISL_4336220, EPI_ISL_4336225, EPI_ISL_4336236, EPI_ISL_4336248, EPI_ISL_4336255, EPI_ISL_4336266, EPI_ISL_4336298, EPI_ISL_4336309, EPI_ISL_4336326, EPI_ISL_4336371, EPI_ISL_4336386, EPI_ISL_4336406, EPI_ISL_4336442, EPI_ISL_4336448, EPI_ISL_4336520, EPI_ISL_4336522, EPI_ISL_4336527 | | | |
| see above | Swedish national genomic surveillance program of SARS-CoV-2 | The Public Health Agency of Sweden | Alma Brolund; Maria Lind Karlberg; Maximilian Riess; Swedish national genomic surveillance program of SARS-CoV-2 |
| EPI_ISL_4229820, EPI_ISL_4230835, EPI_ISL_4230837, EPI_ISL_4230838, EPI_ISL_4444068, EPI_ISL_4608795 | Synlab MVZ Augsburg | Robert Koch Institute | |
| EPI_ISL_3876210, EPI_ISL_3997151, EPI_ISL_4170837, EPI_ISL_4357232 | U.O. Microbiologia Laboratorio Unico Centro Servizi - AUSL della Romagna | U.O. Microbiologia, Laboratorio Unico Centro Servizi - AUSL della Romagna | Giorgio Dirani |
| EPI_ISL_4477279 | UAB Diagnostikos laboratorija | National Public Health Surveillance Laboratory | Ana Steponkiene; Danas Baksa; Jelena Razmuk; Lukas Vasionis; Lukas Zemaitis; Migle Gabrielaite; Svajune Muralyte |
| EPI_ISL_4345204 | UMC Groningen, Clinical Virology, Department of Medical Microbiology and Infection Prevention | UMC Groningen, Clinical Virology, Department of Medical Microbiology and Infection Prevention | Alexander Friedrich; Coretta Van Leer-Buter; Erley Lizarazo-Forero; Hubert Niesters; Lilli Gard; Marjolein Knoester; Monika Fliss; Sigrid Rosema; Xuewei Zhou |
| EPI_ISL_4396619 | UNILABS | Instituto Nacional de Saude (INSA) | Borges et al |
| EPI_ISL_4528342 | University College London, Great Ormond Street Hospital for Children NHS Foundation Trust, Imperial College Healthcare NHS Trust | COVID-19 Genomics UK (COG-UK) Consortium | Charlotte Williams; Helena Tutill; Judith Breuer; Marius Cotic; Mark Kristiansen; Nadua Bayzid; Patricia Dyal; Rachel Williams; Sergi Castellano; Sunando Roy |
| EPI_ISL_4274240, EPI_ISL_4274287, EPI_ISL_4274417, EPI_ISL_4508046, EPI_ISL_4508149, EPI_ISL_4634030, EPI_ISL_4634120 | | | |
| see above | University Hospitals of Geneva, Laboratory of Virology | HUG, Laboratory of Virology and the Health2030 Genome Center | Aline Mamin; Ana Rita Goncalves; Cedric Howald; Deborah Penet; Francisco Perez; Henri Pegeot; Ioannis Xenarios; Keith Harshman; Laurent Kaiser; Lorenzo Cerutti; Melyssa Elies; Samuel Cordey |
| EPI_ISL_4489244, EPI_ISL_4492758, EPI_ISL_4492784, EPI_ISL_4492792, EPI_ISL_4548683, EPI_ISL_4548692, EPI_ISL_4548703, EPI_ISL_4548705, EPI_ISL_4548719, EPI_ISL_4548723, EPI_ISL_4548726, EPI_ISL_4548728, EPI_ISL_4548756, EPI_ISL_4548764 | | | |
| see above | University of Liège COVID-19 testing center | GIGA Medical Genomics | Bouchra Boujemla; Claire Gourzonès; Cécile Meex; Keith Durkin; Laurent Gillet; Maria Artesi; Marie-Pierre Hayette; Nadine Cambisano; Nathalie Renotte; Olivier Ek; Sébastien Bontems; Vincent Bours |
| EPI_ISL_4227593, EPI_ISL_4441301 | Universitätsklinikum Frankfurt - Institut für Medizinische Virologie | Robert Koch Institute | |
| EPI_ISL_4575737, EPI_ISL_4603447 | Utah Public Health Laboratory | Utah Public Health Laboratory | Erin L. Young; John Arnn; Kelly F. Oakeson; Olinto Linares-Perdomo; Pooja Gupta |
| EPI_ISL_4580731, EPI_ISL_4580785, EPI_ISL_4631594 | Viollier AG | Department of Biosystems Science and Engineering, ETH Zürich | Andrea Patrignani; Andreia Cabral de Gouvea; Catharine Aquino; Chaoran Chen; Christian Beisel; Christiane Beckmann; Christoph Noppen; Daniel Ehrsam; Doris Popovic; Elodie Burcklen; Griffin White; Ina Nissen; Isabel Stürmer; Ivan Topolsky; Jay Tracy; Kim Philipp Jablonski; Lara Fuhrmann; Laura Neff; Lennart Opitz; Louis du Plessis; Maria Domenica Moccia; Maurice Redondo; Mirjam Feldkamp; Natascha Santacroce; Niko Beerenwinkel; Olivier Kobel; Ralph Schlapbach; Rebecca Denes; Sarah Nadeau; Simon Grüter; Tanja Stadler; Timothy Sykes |
| EPI_ISL_4576919 | Yale Clinical Virology Lab | Grubaugh Lab - Yale School of Public Health | Anderson Brito; Annie Watkins; Chaney Kalinich; Chantal Vogels; Isabel Ott; Jessica Rothman; Joseph Fauver; Kendall Billig; Mallery Breban; Marie L. Landry; Mary Petrone; Nathan Grubaugh; Tara Alpert; Tobias Koch |
| EPI_ISL_4031083, EPI_ISL_4031087, EPI_ISL_4053013, EPI_ISL_4053016, EPI_ISL_4053018, EPI_ISL_4053026, EPI_ISL_4053030, EPI_ISL_4185858, EPI_ISL_4185871, EPI_ISL_4511716, EPI_ISL_4560589 | | | |
| see above | ZOL | Jessa | Marijke Raymaekers et al. on behalf of the Jessa_cmdLab; Severine Berden et al. on behalf of the Jessa_cmdLab |
| EPI_ISL_4224981, EPI_ISL_4224984, EPI_ISL_4224993, EPI_ISL_4447467, EPI_ISL_4447505, EPI_ISL_4447519, EPI_ISL_4456016, EPI_ISL_4456024, EPI_ISL_4456031, EPI_ISL_4458217, EPI_ISL_4458241, EPI_ISL_4458248, EPI_ISL_4458257, EPI_ISL_4458262, EPI_ISL_4458264, EPI_ISL_4458265, EPI_ISL_4458266, EPI_ISL_4458269, EPI_ISL_4458282, EPI_ISL_4458321, EPI_ISL_4458367, EPI_ISL_4458372, EPI_ISL_4551718 | | | |
| see above | ZOTZ KLIMAS MVZ Düsseldorf-Centrum GbR ÜBAG für Labormedizin, Genetik, Zytologie, Pathologie | Center of Medical Microbiology, Virology, and Hospital Hygiene, University of Duesseldorf | Alexander Dilthey; Andreas Walker; Daniel Strelow; Jessica Nicolai; Jörg Timm; Katrin Hoffmann; Klaus Pfeffer; Lisanna Hülse; Malte Kohns Vasconcelos; Maximilian Damagnez; Nadine Lübke; Patrick Finzer; Rainer Zotz; Tobias Wienemann; Torsten Houwaart |
| EPI_ISL_4440074, EPI_ISL_4611844 | amedes MVZ Hannover | Robert Koch Institute | |
| EPI_ISL_4630672 | labor team w AG | Department of Biosystems Science and Engineering, ETH Zürich | Andrea Patrignani; Andreas Lindauer; Andreia Cabral de Gouvea; Catharine Aquino; Chaoran Chen; Daniel Ehrsam; Doris Popovic; Griffin White; Isabel Stürmer; Ivan Topolsky; Jay Tracy; Kim Philipp Jablonski; Lara Fuhrmann; Laura Neff; Lennart Opitz; Louis du Plessis; Maria Domenica Moccia; Monika Bucher; Niko Beerenwinkel; Ralph Schlapbach; Rebekka Pohl; Sarah Nadeau; Simon Grüter; Tanja Stadler; Timothy Sykes |
