## Supplementary material for "SARS-COV-2 δ variant drives the pandemic in India and Europe via two subvariants": Acknowledgement table on the GISAID genomes used in this study

We gratefully acknowledge the following Authors from the Originating laboratories responsible for obtaining the specimens, as well as the Submitting laboratories where the genome data were generated and shared via GISAID, on which this research is based.

All Submitters of data may be contacted directly via www.gisaid.org

Authors are sorted alphabetically.

| Accession ID | Originating Laboratory | Submitting Laboratory | Authors |
|---|---|---|---|
| EPI_ISL_3935142, EPI_ISL_3935165, EPI_ISL_3935166, EPI_ISL_3935178, EPI_ISL_3935212, EPI_ISL_3935213, EPI_ISL_3935224, EPI_ISL_3935227, EPI_ISL_3935228, EPI_ISL_3935230, EPI_ISL_3935239, EPI_ISL_3935241, EPI_ISL_3935242, EPI_ISL_3935249, EPI_ISL_3935250, EPI_ISL_3935252, EPI_ISL_3935260, EPI_ISL_3935262, EPI_ISL_3935271, EPI_ISL_3935273, EPI_ISL_3935274, EPI_ISL_3935277, EPI_ISL_3935281, EPI_ISL_3935283, EPI_ISL_3935285, EPI_ISL_3935286, EPI_ISL_3935293, EPI_ISL_3935294, EPI_ISL_3935295, EPI_ISL_3935297, EPI_ISL_3935306, EPI_ISL_3935308, EPI_ISL_3935318, EPI_ISL_4545895, EPI_ISL_4545897, EPI_ISL_4545902, EPI_ISL_4545903, EPI_ISL_4545904, EPI_ISL_4545907, EPI_ISL_4545908, EPI_ISL_4545926, EPI_ISL_4545927, EPI_ISL_4545928, EPI_ISL_4545932, EPI_ISL_4545936, EPI_ISL_4545940, EPI_ISL_4545941, EPI_ISL_4545943, EPI_ISL_4546031, EPI_ISL_4546035, EPI_ISL_4546046, EPI_ISL_4546049, EPI_ISL_4546059, EPI_ISL_4546061, EPI_ISL_4546065, EPI_ISL_4546069, EPI_ISL_4546071, EPI_ISL_4546077, EPI_ISL_4546082, EPI_ISL_4546088, EPI_ISL_4546099, EPI_ISL_4546102, EPI_ISL_4546103, EPI_ISL_4546108, EPI_ISL_4546115, EPI_ISL_4546118, EPI_ISL_4546120, EPI_ISL_4546129, EPI_ISL_4546130, EPI_ISL_4546132, EPI_ISL_4546135, EPI_ISL_4546136, EPI_ISL_4546137, EPI_ISL_4546138, EPI_ISL_4546139, EPI_ISL_4546140, EPI_ISL_4546141, EPI_ISL_4546142, EPI_ISL_4546143, EPI_ISL_4546144, EPI_ISL_4546147, EPI_ISL_4546148, EPI_ISL_4546149, EPI_ISL_4546150, EPI_ISL_4546152, EPI_ISL_4546153, EPI_ISL_4546156, EPI_ISL_4546157, EPI_ISL_4546158, EPI_ISL_4546159, EPI_ISL_4546161, EPI_ISL_4546163, EPI_ISL_4546166, EPI_ISL_4546167, EPI_ISL_4546170, EPI_ISL_4546171, EPI_ISL_4546173, EPI_ISL_4546175, EPI_ISL_4546177, EPI_ISL_4546178, EPI_ISL_4546179, EPI_ISL_4546180, EPI_ISL_4546181, EPI_ISL_4546182, EPI_ISL_4546183, EPI_ISL_4546185, EPI_ISL_4546186, EPI_ISL_4546187, EPI_ISL_4546190, EPI_ISL_4546191, EPI_ISL_4546192, EPI_ISL_4546197, EPI_ISL_4546199, EPI_ISL_4546201, EPI_ISL_4546202, EPI_ISL_4546206, EPI_ISL_4546208, EPI_ISL_4546209, EPI_ISL_4546211, EPI_ISL_4546212, EPI_ISL_4546273, EPI_ISL_4546454, EPI_ISL_4546455, EPI_ISL_4546458, EPI_ISL_4546459, EPI_ISL_4546463, EPI_ISL_4546464, EPI_ISL_4546465, EPI_ISL_4546466, EPI_ISL_4546467, EPI_ISL_4546468, EPI_ISL_4546469, EPI_ISL_4546474, EPI_ISL_4546477, EPI_ISL_4546478, EPI_ISL_4546479, EPI_ISL_4546480, EPI_ISL_4546486, EPI_ISL_4546487, EPI_ISL_4546489, EPI_ISL_4546490, EPI_ISL_4546491, EPI_ISL_4546493, EPI_ISL_4546494, EPI_ISL_4546497, EPI_ISL_4546498, EPI_ISL_4546499, EPI_ISL_4546500, EPI_ISL_4546502, EPI_ISL_4546503, EPI_ISL_4546504, EPI_ISL_4546505, EPI_ISL_4546506, EPI_ISL_4546509, EPI_ISL_4546510, EPI_ISL_4546511, EPI_ISL_4546512, EPI_ISL_4546513, EPI_ISL_4546514, EPI_ISL_4546515, EPI_ISL_4546516, EPI_ISL_4546517, EPI_ISL_4546521, EPI_ISL_4546523, EPI_ISL_4546524, EPI_ISL_4546525, EPI_ISL_4546526, EPI_ISL_4546527, EPI_ISL_4546530, EPI_ISL_4546531, EPI_ISL_4546532, EPI_ISL_4546535, EPI_ISL_4546542, EPI_ISL_4546543, EPI_ISL_4546544 | | | |
| see above | AP SSO | CSIR-Centre for Cellular and Molecular Biology - INSACOG | ; Amareshwar Vodapalli; Ara Sreenivas; Archana Bharadwaj Siva; B Himasri; Divya Tej Sowpati; Jandhyala Sai Krishna; Karthik Bharadwaj Tallapaka; Krishna Khairnar; Lamuk Zaveri; Onkar Kulkarni; Payel Mukherjee; Priya Nurkurthy; Rakesh K Mishra; Shreekant Verma; Sofia Banu; Sumedha Avadhanula; Tulasi Nagabandi; Valli Nagalakshmi Undamatla; Vidhyadhari Methuku |
| EPI_ISL_4129618 | AURIGA LABS | INSACOG-KA, NIMHANS | Ananthapadmanabha Kotambail; Anita S Desai; Anson Kunjumon George; Chetan G K; Ellango Ramasamy; Gautham Arunachal Udupi; Mahesh Kumar.C.S; Sony Sharma; V Ravi |
| EPI_ISL_4533541, EPI_ISL_4533932 | Ampligene Diagnostics Private Limited, Ahmedabad | Gujarat Biotechnology Research Centre | Apurvasinh Puvar; Arpit Shukla; Bhadreshsinh Gohil; Chaitanya Joshi; Chirag Pandya; Dinesh Kumar; Janvi Raval; Madhvi Joshi; Nimesh Patel; Nitin Savaliya; Nitin Shukla; Priyank Chavda; Ramesh Pandit; Sonal Sharma; Zarna Patel |
| EPI_ISL_4533550 | Apollo Hospital International Limited ,Bhat Gandhinagar | Gujarat Biotechnology Research Centre | Apurvasinh Puvar; Arpit Shukla; Bhadreshsinh Gohil; Chaitanya Joshi; Dinesh Kumar; Janvi Raval; Madhvi Joshi; Nimesh Patel; Nitin Savaliya; Nitin Shukla; Pankti Shah; Priyank Chavda; Ramesh Pandit; Sonal Sharma; Zarna Patel |
| EPI_ISL_4129936, EPI_ISL_4129942 | BBMP Urban | INSACOG-KA, NIMHANS | Ananthapadmanabha Kotambail; Anita S Desai; Anson Kunjumon George; Chetan G K; Ellango Ramasamy; Gautham Arunachal Udupi; Mahesh Kumar.C.S; Sony Sharma; V Ravi |
| EPI_ISL_4415454, EPI_ISL_4415455, EPI_ISL_4415456, EPI_ISL_4415459, EPI_ISL_4415460, EPI_ISL_4415462, EPI_ISL_4415464, EPI_ISL_4415465, EPI_ISL_4415468, EPI_ISL_4415470, EPI_ISL_4415473, EPI_ISL_4415474, EPI_ISL_4415476, EPI_ISL_4415479, EPI_ISL_4415480, EPI_ISL_4415481, EPI_ISL_4415483, EPI_ISL_4415484, EPI_ISL_4415489, EPI_ISL_4415490, EPI_ISL_4415493, EPI_ISL_4415494, EPI_ISL_4415495, EPI_ISL_4415499, EPI_ISL_4415501, EPI_ISL_4415502, EPI_ISL_4415503, EPI_ISL_4415508, EPI_ISL_4415509, EPI_ISL_4415510, EPI_ISL_4415511, EPI_ISL_4415512, EPI_ISL_4415513, EPI_ISL_4415514, EPI_ISL_4415515, EPI_ISL_4415516, EPI_ISL_4415519, EPI_ISL_4415522, EPI_ISL_4415526, EPI_ISL_4415529, EPI_ISL_4415530, EPI_ISL_4415538, EPI_ISL_4415553, EPI_ISL_4415555, EPI_ISL_4415560, EPI_ISL_4415563, EPI_ISL_4415573, EPI_ISL_4415580, EPI_ISL_4415581, EPI_ISL_4415606, EPI_ISL_4415607, EPI_ISL_4415612, EPI_ISL_4415619, EPI_ISL_4415620, EPI_ISL_4415622, EPI_ISL_4415624, EPI_ISL_4415626, EPI_ISL_4415627, EPI_ISL_4415628, EPI_ISL_4415629, EPI_ISL_4415638, EPI_ISL_4415643, EPI_ISL_4415644, EPI_ISL_4415648, EPI_ISL_4415651, EPI_ISL_4415655, EPI_ISL_4415659, EPI_ISL_4415660, EPI_ISL_4415661, EPI_ISL_4415669, EPI_ISL_4415671, EPI_ISL_4415689, EPI_ISL_4415692, EPI_ISL_4415695, EPI_ISL_4415696, EPI_ISL_4415697, EPI_ISL_4415698, EPI_ISL_4415699, EPI_ISL_4415700, EPI_ISL_4415708, EPI_ISL_4415712, EPI_ISL_4415713, EPI_ISL_4415719, EPI_ISL_4415720, EPI_ISL_4415723, EPI_ISL_4415724, EPI_ISL_4415731, EPI_ISL_4415732, EPI_ISL_4415738, EPI_ISL_4415742, EPI_ISL_4415744, EPI_ISL_4415746, EPI_ISL_4415755, EPI_ISL_4415756, EPI_ISL_4415759, EPI_ISL_4415763 | | | |
| see above | BJMC, Pune | IISER Pune-INSACOG | Aurnab Ghose; Joy Merwin Monteiro; Krishanpal Karmodiya; Rajesh Karyakarte; Suvarna Joshi |
| EPI_ISL_4533554 | Bhavsinhaji General Civil Hospital,Porbandar | Gujarat Biotechnology Research Centre | Apurvasinh Puvar; Arpit Shukla; Bhadreshsinh Gohil; Chaitanya Joshi; Dinesh Kumar; JD Parmar; Janvi Raval; Madhvi Joshi; Nimesh Patel; Nitin Savaliya; Nitin Shukla; Priyank Chavda; Ramesh Pandit; Sonal Sharma; Zarna Patel |
| EPI_ISL_4546375 | CSIR - Central Leather Research Institute | CSIR-Centre for Cellular and Molecular Biology - INSACOG | Amareshwar Vodapalli; Ara Sreenivas; Archana Bharadwaj Siva; B Himasri; Divya Tej Sowpati; Jandhyala Sai Krishna; Karthik Bharadwaj Tallapaka; Lamuk Zaveri; Onkar Kulkarni; Payel Mukherjee; Priya Nurkurthy; Rakesh K Mishra; Shreekant Verma; Sofia Banu; Sumedha Avadhanula; Tulasi Nagabandi; Valli Nagalakshmi Undamatla; Vidhyadhari Methuku |
| EPI_ISL_4545952, EPI_ISL_4545953, EPI_ISL_4545954, EPI_ISL_4545955, EPI_ISL_4545965, EPI_ISL_4545966, EPI_ISL_4545974, EPI_ISL_4545975, EPI_ISL_4545976, EPI_ISL_4545984, EPI_ISL_4545985, EPI_ISL_4545987, EPI_ISL_4545994, EPI_ISL_4545997, EPI_ISL_4546004, EPI_ISL_4546005, EPI_ISL_4546007, EPI_ISL_4546015, EPI_ISL_4546017, EPI_ISL_4546018, EPI_ISL_4546027, EPI_ISL_4546028, EPI_ISL_4546029 | | | |
| see above | CSIR-Central Leather Research Institute | CSIR-Centre for Cellular and Molecular Biology - INSACOG | Amareshwar Vodapalli; Ara Sreenivas; Archana Bharadwaj Siva; B Himasri; Divya Tej Sowpati; Jandhyala Sai Krishna; Karthik Bharadwaj Tallapaka; Lamuk Zaveri; Onkar Kulkarni; Payel Mukherjee; Priya Nurkurthy; Rakesh K Mishra; Shreekant Verma; Sofia Banu; Sumedha Avadhanula; Tulasi Nagabandi; Valli Nagalakshmi Undamatla; Vidhyadhari Methuku |
| EPI_ISL_3935130, EPI_ISL_3935226, EPI_ISL_4546040, EPI_ISL_4546052, EPI_ISL_4546075, EPI_ISL_4546232, EPI_ISL_4546450, EPI_ISL_4546451, EPI_ISL_4546452, EPI_ISL_4546533 | | | |
| see above | CSIR-Centre for Cellular and Molecular Biology - INSACOG | CSIR-Centre for Cellular and Molecular Biology - INSACOG | Amareshwar Vodapalli; Ara Sreenivas; Archana Bharadwaj Siva; B Himasri; Divya Tej Sowpati; Jandhyala Sai Krishna; Karthik Bharadwaj Tallapaka; Krishna Khairnar; Lamuk Zaveri; Onkar Kulkarni; Payel Mukherjee; Priya Nurkurthy; Rakesh K Mishra; Shreekant Verma; Sofia Banu; Sumedha Avadhanula; Tulasi Nagabandi; Valli Nagalakshmi Undamatla; Vidhyadhari Methuku |
| EPI_ISL_4546032, EPI_ISL_4546043, EPI_ISL_4546044, EPI_ISL_4546055, EPI_ISL_4546068, EPI_ISL_4546079, EPI_ISL_4546080, EPI_ISL_4546092, EPI_ISL_4546093, EPI_ISL_4546105, EPI_ISL_4546116, EPI_ISL_4546382, EPI_ISL_4546384, EPI_ISL_4546386, EPI_ISL_4546393, EPI_ISL_4546395, EPI_ISL_4546402, EPI_ISL_4546403, EPI_ISL_4546411, EPI_ISL_4546412, EPI_ISL_4546413, EPI_ISL_4546414, EPI_ISL_4546424, EPI_ISL_4546431, EPI_ISL_4546432, EPI_ISL_4546441, EPI_ISL_4546442 | | | |
| see above | CSIR-National Environmental Engineering Research Institute | CSIR-Centre for Cellular and Molecular Biology - INSACOG | Amareshwar Vodapalli; Ara Sreenivas; Archana Bharadwaj Siva; B Himasri; Divya Tej Sowpati; Jandhyala Sai Krishna; Karthik Bharadwaj Tallapaka; Krishna Khairnar; Lamuk Zaveri; Onkar Kulkarni; Payel Mukherjee; Priya Nurkurthy; Rakesh K Mishra; Shreekant Verma; Sofia Banu; Sumedha Avadhanula; Tulasi Nagabandi; Valli Nagalakshmi Undamatla; Vidhyadhari Methuku |
| EPI_ISL_4415402, EPI_ISL_4415403, EPI_ISL_4415404, EPI_ISL_4415405, EPI_ISL_4415408, EPI_ISL_4415409, EPI_ISL_4415532, EPI_ISL_4415564, EPI_ISL_4415566, EPI_ISL_4415600, EPI_ISL_4415609, EPI_ISL_4415632, EPI_ISL_4415633, EPI_ISL_4415635, EPI_ISL_4415645, EPI_ISL_4415677, EPI_ISL_4415691, EPI_ISL_4415702, EPI_ISL_4415717, EPI_ISL_4415727, EPI_ISL_4415760 | | | |
| see above | Department of Microbiology, Government Medical College, Baramati | IISER Pune-INSACOG | Aurnab Ghose; Joy Merwin Monteiro; Krishanpal Karmodiya |
| EPI_ISL_4533906, EPI_ISL_4533967 | Desai Metropolis Health Service Pvt. Ltd, surat | Gujarat Biotechnology Research Centre | Apurvasinh Puvar; Arpit Shukla; Bhadreshsinh Gohil; Chaitanya Joshi; Dinesh Kumar; Janvi Raval; Madhvi Joshi; Manali Shah; Nimesh Patel; Nitin Savaliya; Nitin Shukla; Priyank Chavda; Ramesh Pandit; Sonal Sharma; Zarna Patel |
| EPI_ISL_4533930 | Dr. Bhatt Lab, Rajkot | Gujarat Biotechnology Research Centre | Apurvasinh Puvar; Arpit Shukla; Bhadreshsinh Gohil; Chaitanya Joshi; Dinesh Kumar; Janvi Raval; Madhvi Joshi; Nimesh Patel; Nitin Savaliya; Nitin Shukla; Priyank Chavda; Ramesh Pandit; Sonal Sharma; Sonam Maniyar; Zarna Patel |
| EPI_ISL_4104772, EPI_ISL_4104788, EPI_ISL_4104795, EPI_ISL_4104825 | GMERS Junagadh Medical College | Gujarat Biotechnology Research Centre | Arpit Shukla; Bhadreshsinh Gohil; Chaitanya Joshi; Dinesh Kumar; Janvi Raval; Madhvi Joshi; Nimesh Patel; Niraj Kumar Biswas; Nitin Savaliya; Nitin Shukla; Ramesh Pandit; Sonal Sharma; Twinkle Soni; Zarna Patel |
| EPI_ISL_4533927 | GMERS Medical College, Gandhinagar | Gujarat Biotechnology Research Centre | Apurvasinh Puvar; Arpit Shukla; Bhadreshsinh Gohil; Chaitanya Joshi; Dinesh Kumar; Gaurishankar Shrimali; Janvi Raval; Madhvi Joshi; Nimesh Patel; Nitin Savaliya; Nitin Shukla; Priyank Chavda; Ramesh Pandit; Sonal Sharma; Zarna Patel |
| EPI_ISL_4104777, EPI_ISL_4104812, EPI_ISL_4104813 | GMERS Medical College, Valsad | Gujarat Biotechnology Research Centre | Arpit Shukla; Bhadreshsinh Gohil; Chaitanya Joshi; Dinesh Kumar; Janvi Raval; Madhvi Joshi; Nimesh Patel; Nitin Savaliya; Nitin Shukla; Ramesh Pandit; Sonal Sharma; Twinkle Soni; Vicky Gandhi; Zarna Patel |
| EPI_ISL_4058069, EPI_ISL_4058071, EPI_ISL_4058073, EPI_ISL_4058074, EPI_ISL_4058076, EPI_ISL_4058078, EPI_ISL_4058079, EPI_ISL_4058081, EPI_ISL_4058083, EPI_ISL_4058086 | | | |
| see above | Gandhi Hospital | CDFD | Ashwin Dalal; Asmita Gupta; Divya Vashisht; Murali Bashyam; Nagamani Kammili; Vinay Donipadi |
| EPI_ISL_4058052, EPI_ISL_4058056, EPI_ISL_4058068, EPI_ISL_4058148, EPI_ISL_4058175, EPI_ISL_4058201, EPI_ISL_4058203, EPI_ISL_4058238, EPI_ISL_4058240 | | | |
| see above | Gandhi Medical College | CDFD | Ashwin Dalal; Asmita Gupta; Divya Vashisht; Murali Bashyam; Nagamani Kammili; Vinay Donipadi |
| EPI_ISL_4104822 | General Hospital, Chotaudaipur | Gujarat Biotechnology Research Centre | Arpit Shukla; Arti Thakur; Bhadreshsinh Gohil; Chaitanya Joshi; Dinesh Kumar; Janvi Raval; Madhvi Joshi; Nimesh Patel; Nitin Savaliya; Nitin Shukla; Ramesh Pandit; Sonal Sharma; Twinkle Soni; Zarna Patel |
| EPI_ISL_4104776, EPI_ISL_4104781, EPI_ISL_4104782, EPI_ISL_4104783, EPI_ISL_4104833 | Goverment Medical College, Bhavnagar | Gujarat Biotechnology Research Centre | Arpit Shukla; Bhadreshsinh Gohil; Chaitanya Joshi; Dinesh Kumar; Janvi Raval; Kairavi Jwalantbhai Desai; Madhvi Joshi; Nimesh Patel; Nitin Savaliya; Nitin Shukla; Ramesh Pandit; Sonal Sharma; Twinkle Soni; Zarna Patel |
| EPI_ISL_4058091, EPI_ISL_4058093, EPI_ISL_4058101, EPI_ISL_4058103, EPI_ISL_4058104, EPI_ISL_4058109, EPI_ISL_4058192, EPI_ISL_4058196, EPI_ISL_4058198, EPI_ISL_4058199 | | | |
| see above | Government Fever Hospital | CDFD | Ashwin Dalal; Asmita Gupta; Divya Vashisht; Murali Bashyam; Nagamani Kammili; Vinay Donipadi |
| EPI_ISL_4058205, EPI_ISL_4058211, EPI_ISL_4058213, EPI_ISL_4058215, EPI_ISL_4058217, EPI_ISL_4058218, EPI_ISL_4058220, EPI_ISL_4058222, EPI_ISL_4058223 | | | |
| see above | Government General Hospital | CDFD | Ashwin Dalal; Asmita Gupta; Divya Vashisht; Murali Bashyam; Nagamani Kammili; Vinay Donipadi |
| EPI_ISL_3507489, EPI_ISL_3507499, EPI_ISL_3507500, EPI_ISL_3507501 | Government General Hospital | Centre for DNA Fingerprinting and Diagnostics | Ashwin Dalal; Asmita Gupta; Divya Vashisht; Murali Bashyam; Nagamani Kammili; Reelina Basu; Vinay Donipadi |
| EPI_ISL_4058061, EPI_ISL_4058062, EPI_ISL_4058064, EPI_ISL_4058066, EPI_ISL_4058174, EPI_ISL_4058245, EPI_ISL_4058250, EPI_ISL_4058252 | | | |
| see above | Government Maternity Hospital | CDFD | Ashwin Dalal; Asmita Gupta; Divya Vashisht; Murali Bashyam; Nagamani Kammili; Vinay Donipadi |
| EPI_ISL_4533959, EPI_ISL_4533960 | Green Cross Pathology & Molecular Laboratory | Gujarat Biotechnology Research Centre | Apurvasinh Puvar; Arpit Shukla; Bhadreshsinh Gohil; Chaitanya Joshi; Dinesh Kumar; Janvi Raval; Madhvi Joshi; Narendra M Patel; Nimesh Patel; Nitin Savaliya; Nitin Shukla; Priyank Chavda; Ramesh Pandit; Sonal Sharma; Zarna Patel |
| EPI_ISL_4104824 | Green Cross Pathology & Molecular Laboratory, Ahmedabad | Gujarat Biotechnology Research Centre | Arpit Shukla; Bhadreshsinh Gohil; Chaitanya Joshi; Dinesh Kumar; Janvi Raval; Madhvi Joshi; Narendra Patel; Nimesh Patel; Nitin Savaliya; Nitin Shukla; Ramesh Pandit; Sonal Sharma; Twinkle Soni; Zarna Patel |
| EPI_ISL_3717583 | Green Cross Pathology and Molecular Laboratory, Ahmedabad | Gujarat Biotechnology Research Centre | Arpit Shukla; Bhadreshsinh Gohil; Chaitanya Joshi; Dinesh Kumar; Janvi Raval; Madhvi Joshi; Narendra Patel; Nimesh Patel; Nitin Savaliya; Nitin Shukla; Ramesh Pandit; Sonal Sharma; Twinkle Soni; Zarna Patel |
| EPI_ISL_4533937, EPI_ISL_4533938, EPI_ISL_4533939, EPI_ISL_4533940, EPI_ISL_4533941, EPI_ISL_4533942 | Green cross Genetic Lab Pvt. Ltd, Ahmedabad | Gujarat Biotechnology Research Centre | Apurvasinh Puvar; Arpit Shukla; Bhadreshsinh Gohil; Chaitanya Joshi; Dinesh Kumar; Janvi Raval; Madhvi Joshi; Nimesh Patel; Nitin Savaliya; Nitin Shukla; Priyank Chavda; Ramesh Pandit; Sonal Sharma; Umang Patel; Zarna Patel |
| EPI_ISL_4533923, EPI_ISL_4533924 | Gujarat Pathology Laboratory, Ahmedabad | Gujarat Biotechnology Research Centre | Apurvasinh Puvar; Arpit Shukla; Bhadreshsinh Gohil; Chaitanya Joshi; Dinesh Kumar; Janvi Raval; Madhvi Joshi; Nimesh Patel; Niraj Kothari; Nitin Savaliya; Nitin Shukla; Priyank Chavda; Ramesh Pandit; Sonal Sharma; Zarna Patel |
| EPI_ISL_4129745, EPI_ISL_4129747 | Hassan Institute of Medical Sciences | INSACOG-KA, NIMHANS | Ananthapadmanabha Kotambail; Anita S Desai; Anson Kunjumon George; Chetan G K; Ellango Ramasamy; Gautham Arunachal Udupi; Mahesh Kumar.C.S; Sony Sharma; V Ravi |
| EPI_ISL_3533110, EPI_ISL_3533111, EPI_ISL_3533112, EPI_ISL_3533115, EPI_ISL_3533116, EPI_ISL_3982433, EPI_ISL_3982437, EPI_ISL_3982438, EPI_ISL_3982439, EPI_ISL_3982440, EPI_ISL_3982441, EPI_ISL_3982442, EPI_ISL_3982443, EPI_ISL_3982444, EPI_ISL_3982445, EPI_ISL_3982446, EPI_ISL_3982447, EPI_ISL_3982448, EPI_ISL_3982449, EPI_ISL_3982450, EPI_ISL_3982451, EPI_ISL_3982452, EPI_ISL_3982453, EPI_ISL_3982454, EPI_ISL_3982455, EPI_ISL_3982456, EPI_ISL_3982457, EPI_ISL_3982458, EPI_ISL_3982459, EPI_ISL_3982460, EPI_ISL_3982461, EPI_ISL_3982462, EPI_ISL_3982463, EPI_ISL_3982465, EPI_ISL_3982466, EPI_ISL_3982467, EPI_ISL_3982468, EPI_ISL_3982469, EPI_ISL_3982470, EPI_ISL_3982475, EPI_ISL_3982478, EPI_ISL_3982479, EPI_ISL_3982480, EPI_ISL_3982481, EPI_ISL_3982482, EPI_ISL_3982483, EPI_ISL_3982485, EPI_ISL_3982486, EPI_ISL_3982487, EPI_ISL_3982489, EPI_ISL_3982490, EPI_ISL_3982495, EPI_ISL_3982497, EPI_ISL_3982498, EPI_ISL_3982499, EPI_ISL_3982500, EPI_ISL_3982502, EPI_ISL_3982503, EPI_ISL_3982504, EPI_ISL_3982506, EPI_ISL_3982507, EPI_ISL_3982508, EPI_ISL_3982509, EPI_ISL_3982510, EPI_ISL_3982511, EPI_ISL_3982512, | | | |

| | | | |
|---|---|---|---|
| EPI_ISL_3982513, EPI_ISL_3982515, EPI_ISL_3982516, EPI_ISL_3982521, EPI_ISL_3982522, EPI_ISL_3982523, EPI_ISL_3982526, EPI_ISL_3982527, EPI_ISL_3982528, EPI_ISL_3982529, EPI_ISL_3982530, EPI_ISL_3982531, EPI_ISL_3982535, EPI_ISL_3982538, EPI_ISL_3982540, EPI_ISL_3982541, EPI_ISL_3982547, EPI_ISL_3982549, EPI_ISL_3982550, EPI_ISL_3982552, EPI_ISL_3982554, EPI_ISL_3982555, EPI_ISL_3982558, EPI_ISL_3982560, EPI_ISL_3982561, EPI_ISL_3982565, EPI_ISL_3982566, EPI_ISL_3982567, EPI_ISL_3982568, EPI_ISL_3982569, EPI_ISL_3982570, EPI_ISL_3982578, EPI_ISL_3982579, EPI_ISL_3982580, EPI_ISL_3982581, EPI_ISL_3982583, EPI_ISL_3982584, EPI_ISL_3984265, EPI_ISL_3984266 | | | |
| see above | ICMR-National Institute of Virology - INSACOG | NIV Influenza | Dr. Varsha Potdar and NIC Team |
| EPI_ISL_3982974, EPI_ISL_3982975, EPI_ISL_3982976, EPI_ISL_3982977, EPI_ISL_3982978, EPI_ISL_3982979, EPI_ISL_3982986, EPI_ISL_3982987, EPI_ISL_3982990, EPI_ISL_3982991, EPI_ISL_3982992, EPI_ISL_3982993, EPI_ISL_3982994, EPI_ISL_3982995, EPI_ISL_3982996, EPI_ISL_3982998, EPI_ISL_3983000, EPI_ISL_3983001, EPI_ISL_3983002 | | | |
| see above | INSACOG-Assam | National Institute of Biomedical Genomics – INSACOG | Ajanta Sharma; Arindam Maitra; Kailash Chamuah; Lahari Saikia; Nidhan Kumar Biswas; Saumitra Das; Sreedhar Chinnaswamy |
| EPI_ISL_4504794, EPI_ISL_4504798, EPI_ISL_4504806, EPI_ISL_4504809 | INSACOG-MANIPUR | National Institute of Biomedical Genomics - INSACOG | Arindam Maitra; Kh. Ranjana Devi; L. Shivadutta Singh; Nidhan Kumar Biswas; R.K.Manojkumar Singh; Saumitra Das; Sreedhar Chinnaswamy |
| EPI_ISL_4312064, EPI_ISL_4312077, EPI_ISL_4312513, EPI_ISL_4312535, EPI_ISL_4312617, EPI_ISL_4312674, EPI_ISL_4312741, EPI_ISL_4312746, EPI_ISL_4312809, EPI_ISL_4312850, EPI_ISL_4312930, EPI_ISL_4313104 | | | |
| see above | INSACOG-Mizoram | National Institute of Biomedical Genomics – INSACOG | Arindam Maitra; Gracy Laldinmawii; N Senthil Kumar; Nidhan Kumar Biswas; Saumitra Das; Sreedhar Chinnaswamy; Swagnik Roy |
| EPI_ISL_4504571, EPI_ISL_4504573, EPI_ISL_4504574, EPI_ISL_4504575, EPI_ISL_4504577, EPI_ISL_4504578, EPI_ISL_4504579, EPI_ISL_4504580, EPI_ISL_4504581, EPI_ISL_4504584, EPI_ISL_4504585, EPI_ISL_4504586, EPI_ISL_4504587, EPI_ISL_4504588, EPI_ISL_4504589, EPI_ISL_4504592, EPI_ISL_4504593, EPI_ISL_4504595, EPI_ISL_4504596, EPI_ISL_4504597, EPI_ISL_4504598, EPI_ISL_4504600, EPI_ISL_4504601, EPI_ISL_4504604, EPI_ISL_4504605, EPI_ISL_4504606, EPI_ISL_4504607, EPI_ISL_4504609, EPI_ISL_4504611, EPI_ISL_4504613, EPI_ISL_4504614, EPI_ISL_4504615, EPI_ISL_4504616, EPI_ISL_4504617, EPI_ISL_4504618, EPI_ISL_4504620, EPI_ISL_4504621, EPI_ISL_4504622, EPI_ISL_4504624, EPI_ISL_4504625, EPI_ISL_4504627, EPI_ISL_4504628, EPI_ISL_4504629, EPI_ISL_4504631, EPI_ISL_4504632, EPI_ISL_4504633, EPI_ISL_4504634, EPI_ISL_4504635, EPI_ISL_4504637, EPI_ISL_4504639, EPI_ISL_4504640, EPI_ISL_4504642 | | | |
| see above | INSACOG-Sikkim | National Institute of Biomedical Genomics - INSACOG | Arindam Maitra; Dr. Shrijana Gurung; Kaden Zangmu Bhutia; Nidhan Kumar Biswas; Saumitra Das; Sreedhar Chinnaswamy Tshering Ongmu Bhutia |
| EPI_ISL_4504193, EPI_ISL_4504194, EPI_ISL_4504195, EPI_ISL_4504196, EPI_ISL_4504197, EPI_ISL_4504199, EPI_ISL_4504200, EPI_ISL_4504201, EPI_ISL_4504202, EPI_ISL_4504203, EPI_ISL_4504204, EPI_ISL_4504205, EPI_ISL_4504206, EPI_ISL_4504207, EPI_ISL_4504208, EPI_ISL_4504210, EPI_ISL_4504212, EPI_ISL_4504213, EPI_ISL_4504214, EPI_ISL_4504216, EPI_ISL_4504217, EPI_ISL_4504218, EPI_ISL_4504219, EPI_ISL_4504220, EPI_ISL_4504221, EPI_ISL_4504222, EPI_ISL_4504226, EPI_ISL_4504227, EPI_ISL_4504228, EPI_ISL_4504229, EPI_ISL_4504231, EPI_ISL_4504234, EPI_ISL_4504236, EPI_ISL_4504238, EPI_ISL_4504239, EPI_ISL_4504240, EPI_ISL_4504241, EPI_ISL_4504243, EPI_ISL_4504244, EPI_ISL_4504246, EPI_ISL_4504247, EPI_ISL_4504248, EPI_ISL_4504249, EPI_ISL_4504250, EPI_ISL_4504251, EPI_ISL_4504252, EPI_ISL_4504253, EPI_ISL_4504254, EPI_ISL_4504255, EPI_ISL_4504256, EPI_ISL_4504257, EPI_ISL_4504258, EPI_ISL_4504259, EPI_ISL_4504260, EPI_ISL_4504261, EPI_ISL_4504262, EPI_ISL_4504263, EPI_ISL_4504264, EPI_ISL_4504265, EPI_ISL_4504266, EPI_ISL_4504268, EPI_ISL_4504269, EPI_ISL_4504270, EPI_ISL_4504272, EPI_ISL_4504273, EPI_ISL_4504275, EPI_ISL_4504277, EPI_ISL_4504278, EPI_ISL_4504279, EPI_ISL_4504280, EPI_ISL_4504282, EPI_ISL_4504283, EPI_ISL_4504284, EPI_ISL_4504285, EPI_ISL_4504286, EPI_ISL_4504298, EPI_ISL_4504299, EPI_ISL_4504300, EPI_ISL_4504301, EPI_ISL_4504302, EPI_ISL_4504303, EPI_ISL_4504304, EPI_ISL_4504305, EPI_ISL_4504307, EPI_ISL_4504311, EPI_ISL_4504312, EPI_ISL_4504314, EPI_ISL_4504316, EPI_ISL_4504318, EPI_ISL_4504319, EPI_ISL_4504320, EPI_ISL_4504321, EPI_ISL_4504322, EPI_ISL_4504323, EPI_ISL_4504324, EPI_ISL_4504325, EPI_ISL_4504326, EPI_ISL_4504337, EPI_ISL_4504338, EPI_ISL_4504342, EPI_ISL_4504343, EPI_ISL_4504344, EPI_ISL_4504387, EPI_ISL_4504389, EPI_ISL_4504390, EPI_ISL_4504391, EPI_ISL_4504393, EPI_ISL_4504394, EPI_ISL_4504395, EPI_ISL_4504396, EPI_ISL_4504397, EPI_ISL_4504398, EPI_ISL_4504399, EPI_ISL_4504400, EPI_ISL_4504402, EPI_ISL_4504403, EPI_ISL_4504406, EPI_ISL_4504408, EPI_ISL_4504410, EPI_ISL_4504411, EPI_ISL_4504414, EPI_ISL_4504415, EPI_ISL_4504417, EPI_ISL_4504418, EPI_ISL_4504419, EPI_ISL_4504420, EPI_ISL_4504421, EPI_ISL_4504422, EPI_ISL_4504423, EPI_ISL_4504426, EPI_ISL_4504427, EPI_ISL_4504428, EPI_ISL_4504429, EPI_ISL_4504430, EPI_ISL_4504433, EPI_ISL_4504434, EPI_ISL_4504435, EPI_ISL_4504436, EPI_ISL_4504437, EPI_ISL_4504439, EPI_ISL_4504440, EPI_ISL_4504442, EPI_ISL_4504443, EPI_ISL_4504444, EPI_ISL_4504445, EPI_ISL_4504446, EPI_ISL_4504448, EPI_ISL_4504450, EPI_ISL_4504455, EPI_ISL_4504456, EPI_ISL_4504457, EPI_ISL_4504458, EPI_ISL_4504459, EPI_ISL_4504460, EPI_ISL_4504461, EPI_ISL_4504462, EPI_ISL_4504463, EPI_ISL_4504464, EPI_ISL_4504465, EPI_ISL_4504466, EPI_ISL_4504467, EPI_ISL_4504468, EPI_ISL_4504471, EPI_ISL_4504472, EPI_ISL_4504473, EPI_ISL_4504474, EPI_ISL_4504475, EPI_ISL_4504476, EPI_ISL_4504477, EPI_ISL_4504478, EPI_ISL_4504479, EPI_ISL_4504480, EPI_ISL_4504481, EPI_ISL_4504482, EPI_ISL_4504483, EPI_ISL_4504484, EPI_ISL_4504486, EPI_ISL_4504489, EPI_ISL_4504491, EPI_ISL_4504492, EPI_ISL_4504493, EPI_ISL_4504495, EPI_ISL_4504496, EPI_ISL_4504497, EPI_ISL_4504498, EPI_ISL_4504499, EPI_ISL_4504500, EPI_ISL_4504501, EPI_ISL_4504502, EPI_ISL_4504503, EPI_ISL_4504504, EPI_ISL_4504505, EPI_ISL_4504506, EPI_ISL_4504507, EPI_ISL_4504508, EPI_ISL_4504509, EPI_ISL_4504510, EPI_ISL_4504511, EPI_ISL_4504512, EPI_ISL_4504513, EPI_ISL_4504514, EPI_ISL_4504515, EPI_ISL_4504516, EPI_ISL_4504517, EPI_ISL_4504518, EPI_ISL_4504519, EPI_ISL_4504520, EPI_ISL_4504521, EPI_ISL_4504522, EPI_ISL_4504523, EPI_ISL_4504524, EPI_ISL_4504525, EPI_ISL_4504528, EPI_ISL_4504529, EPI_ISL_4504530, EPI_ISL_4504534, EPI_ISL_4504535, EPI_ISL_4504536, EPI_ISL_4504538, EPI_ISL_4504540, EPI_ISL_4504541, EPI_ISL_4504542, EPI_ISL_4504544, EPI_ISL_4504545, EPI_ISL_4504546, EPI_ISL_4504547, EPI_ISL_4504548, EPI_ISL_4504549, EPI_ISL_4504550, EPI_ISL_4504551, EPI_ISL_4504554, EPI_ISL_4504555, EPI_ISL_4504556, EPI_ISL_4504557, EPI_ISL_4504558, EPI_ISL_4504559, EPI_ISL_4504560, EPI_ISL_4504561, EPI_ISL_4504563, EPI_ISL_4504565, EPI_ISL_4504566, EPI_ISL_4504567, EPI_ISL_4504568 | | | |
| see above | INSACOG-WB | National Institute of Biomedical Genomics - INSACOG | Ajay Chakraborti; Arindam Maitra; Bhaswati Bandyopadhyay; Nidhan Kumar Biswas; Saumitra Das; Sreedhar Chinnaswamy; Tamal Ghosh |
| EPI_ISL_4192927, EPI_ISL_4192928, EPI_ISL_4193023, EPI_ISL_4193027, EPI_ISL_4193029, EPI_ISL_4193031, EPI_ISL_4193034, EPI_ISL_4193035, EPI_ISL_4193057, EPI_ISL_4193058, EPI_ISL_4193063, EPI_ISL_4193065, EPI_ISL_4193066, EPI_ISL_4193067, EPI_ISL_4193068, EPI_ISL_4193069, EPI_ISL_4193070, EPI_ISL_4193079, EPI_ISL_4193084, EPI_ISL_4193085, EPI_ISL_4193086, EPI_ISL_4193102, EPI_ISL_4193104, EPI_ISL_4193113, EPI_ISL_4193120, EPI_ISL_4193121, EPI_ISL_4193127, EPI_ISL_4193129, EPI_ISL_4193130, EPI_ISL_4193131, EPI_ISL_4193148, EPI_ISL_4193149, EPI_ISL_4193150, EPI_ISL_4193155, EPI_ISL_4193157, EPI_ISL_4193158, EPI_ISL_4193161, EPI_ISL_4193162, EPI_ISL_4193163, EPI_ISL_4193164, EPI_ISL_4193165, EPI_ISL_4193166, EPI_ISL_4193167, EPI_ISL_4193169, EPI_ISL_4193172, EPI_ISL_4193174, EPI_ISL_4193178, EPI_ISL_4193179, EPI_ISL_4193181, EPI_ISL_4193182, EPI_ISL_4193183, EPI_ISL_4193186, EPI_ISL_4193191, EPI_ISL_4193200, EPI_ISL_4193202, EPI_ISL_4193206, EPI_ISL_4193215, EPI_ISL_4193219, EPI_ISL_4193226, EPI_ISL_4193227, EPI_ISL_4193242, EPI_ISL_4193245, EPI_ISL_4193251, EPI_ISL_4193271, EPI_ISL_4193286, EPI_ISL_4193288, EPI_ISL_4193295, EPI_ISL_4193297, EPI_ISL_4193301, EPI_ISL_4193303, EPI_ISL_4193318, EPI_ISL_4193328, EPI_ISL_4193334, EPI_ISL_4193335, EPI_ISL_4193336, EPI_ISL_4193344, EPI_ISL_4193350, EPI_ISL_4193352, EPI_ISL_4193353, EPI_ISL_4193354, EPI_ISL_4193357, EPI_ISL_4193359, EPI_ISL_4193365, EPI_ISL_4193370, EPI_ISL_4193372, EPI_ISL_4193381, EPI_ISL_4193382, EPI_ISL_4193385, EPI_ISL_4193387, EPI_ISL_4193388, EPI_ISL_4193392, EPI_ISL_4193396, EPI_ISL_4193399, EPI_ISL_4193405, EPI_ISL_4193413, EPI_ISL_4193416, EPI_ISL_4193429, EPI_ISL_4193430, EPI_ISL_4193437, EPI_ISL_4193442, EPI_ISL_4193445, EPI_ISL_4193457, EPI_ISL_4193462, EPI_ISL_4193466, EPI_ISL_4193473, EPI_ISL_4193477, EPI_ISL_4193479, EPI_ISL_4193481, EPI_ISL_4193487, EPI_ISL_4193491, EPI_ISL_4193492, EPI_ISL_4193494, EPI_ISL_4193495, EPI_ISL_4193496, EPI_ISL_4193498, EPI_ISL_4193500, EPI_ISL_4193502 | | | |
| see above | INSACOG-WB | National Institute of Biomedical Genomics – INSACOG | Ajay Chakraborti; Arindam Maitra; Bhaswati Bandyopadhyay; Nidhan Kumar Biswas; Saumitra Das; Sreedhar Chinnaswamy; Tamal Ghosh |
| EPI_ISL_4312113, EPI_ISL_4312119, EPI_ISL_4312121, EPI_ISL_4312124, EPI_ISL_4312126, EPI_ISL_4312128, EPI_ISL_4312141, EPI_ISL_4312158, EPI_ISL_4312160, EPI_ISL_4312180, EPI_ISL_4312204, EPI_ISL_4312221, EPI_ISL_4312279, EPI_ISL_4312281, EPI_ISL_4312315, EPI_ISL_4312328, EPI_ISL_4312337, EPI_ISL_4312347, EPI_ISL_4312358, EPI_ISL_4312363, EPI_ISL_4312367, EPI_ISL_4312382, EPI_ISL_4312389, EPI_ISL_4312391, EPI_ISL_4312393, EPI_ISL_4312398, EPI_ISL_4312429, EPI_ISL_4312435, EPI_ISL_4312448, EPI_ISL_4312466, EPI_ISL_4312482, EPI_ISL_4312492, EPI_ISL_4312507, EPI_ISL_4312518, EPI_ISL_4312532, EPI_ISL_4312539, EPI_ISL_4312560, EPI_ISL_4312566, EPI_ISL_4312619, EPI_ISL_4312646, EPI_ISL_4312653, EPI_ISL_4312662, EPI_ISL_4312672, EPI_ISL_4312694, EPI_ISL_4312705, EPI_ISL_4312707, EPI_ISL_4312751, EPI_ISL_4312778, EPI_ISL_4312783, EPI_ISL_4312845, EPI_ISL_4312880, EPI_ISL_4312887, EPI_ISL_4312904, EPI_ISL_4312950, EPI_ISL_4312963, EPI_ISL_4312985, EPI_ISL_4312987, EPI_ISL_4313017, EPI_ISL_4313023, EPI_ISL_4313030, EPI_ISL_4313039, EPI_ISL_4313070, EPI_ISL_4313072, EPI_ISL_4313085, EPI_ISL_4313090 | | | |
| see above | INSACOG-WB | National Institute of Biomedical Genomics – INSACOG | Ajay Chakraborti; Arindam Maitra; Bhaswati Bandyopadhyay; Nidhan Kumar Biswas; Saumitra Das; Sreedhar Chinnaswamy; Tamal Ghosh |
| EPI_ISL_3717582 | Indian Red Cross Society, Vadaj, Ahmedabad | Gujarat Biotechnology Research Centre | Arpit Shukla; Bhadreshsinh Gohil; Chaitanya Joshi; Dinesh Kumar; Janvi Raval; Madhvi Joshi; Nimesh Patel; Nitin Savaliya; Nitin Shukla; No details; Ramesh Pandit; Sonal Sharma; Twinkle Soni; Zarna Patel |
| EPI_ISL_4058164, EPI_ISL_4058165, EPI_ISL_4058167, EPI_ISL_4058227, EPI_ISL_4058229, EPI_ISL_4058233 | Institute of Preventive Medicine | CDFD | Ashwin Dalal; Asmita Gupta; Divya Vashisht; Murali Bashyam; Nagamani Kammili; Vinay Donipadi |
| EPI_ISL_3833606, EPI_ISL_3905544, EPI_ISL_4533929, EPI_ISL_4533933 | Invitro Speciality laboratory, Ahmedabad | Gujarat Biotechnology Research Centre | Apurvasinh Puvar; Arpit Shukla; Bhadreshsinh Gohil; Chaitanya Joshi; Dinesh Kumar; Janvi Raval; Madhvi Joshi; Nimesh Patel; Nitin Savaliya; Nitin Shukla; Priyank Chavda; Ramesh Pandit; Sonal Sharma; Twinkle Soni; Viral N Pathak; Zarna Patel |
| EPI_ISL_4558039, EPI_ISL_4558040, EPI_ISL_4558041, EPI_ISL_4558042, EPI_ISL_4558043, EPI_ISL_4558044, EPI_ISL_4558047, EPI_ISL_4558048, EPI_ISL_4558049, EPI_ISL_4558050, EPI_ISL_4558051, EPI_ISL_4558060, EPI_ISL_4558063, EPI_ISL_4558064, EPI_ISL_4558067, EPI_ISL_4558073 | | | |
| see above | MEDICA Superspeciality Hospital | CSIR-Indian Institute of Chemical Biology | Arpita Ghosh Mitra; Arun Bandyopadhyay; Aviral Roy; Partha Chakrabarti; Poulomi Sarkar; Saikat Chakrabarti; Sarthak Banerjee; Siddik Sarkar; Soumen Saha |
| EPI_ISL_4104818, EPI_ISL_4104831, EPI_ISL_4104832 | Mangalam Invitro Laboratory, Rajkot | Gujarat Biotechnology Research Centre | Arpit Shukla; Bhadreshsinh Gohil; Chaitanya Joshi; Dinesh Kumar; Janvi Raval; Madhvi Joshi; Nimesh Patel; Nitin Savaliya; Nitin Shukla; Rahul P Thumbar; Ramesh Pandit; Sonal Sharma; Twinkle Soni; Zarna Patel |
| EPI_ISL_3935185, EPI_ISL_3935222, EPI_ISL_3935247, EPI_ISL_3935258, EPI_ISL_3935278, EPI_ISL_3935289, EPI_ISL_3935290 | | | |
| see above | Mapmygenome | CSIR-Centre for Cellular and Molecular Biology - INSACOG | Amareshwar Vodapalli; Ara Sreenivas; Archana Bharadwaj Siva; B Himasri; Divya Tej Sowpati; Jandhyala Sai Krishna; Karthik Bharadwaj Tallapaka; Lamuk Zaveri; Onkar Kulkarni; Payel Mukherjee; Priya Nurkurthy; Rakesh K Mishra; Shreekant Verma; Sofia Banu; Sumedha Avadhanula; Tulasi Nagabandi; Valli Nagalakshmi Undamatla; Vidhyadhari Methuku |
| EPI_ISL_4533925, EPI_ISL_4533926 | Medical College, Baroda | Gujarat Biotechnology Research Centre | Apurvasinh Puvar; Arpit Shukla; Bhadreshsinh Gohil; Chaitanya Joshi; Dinesh Kumar; Janvi Raval; Madhvi Joshi; Nimesh Patel; Nitin Savaliya; Nitin Shukla; Priyank Chavda; Ramesh Pandit; Sonal Sharma; Tanuja Javdekar; Zarna Patel |
| EPI_ISL_4129753, EPI_ISL_4129756, EPI_ISL_4129762, EPI_ISL_4129765, EPI_ISL_4129769, EPI_ISL_4129771 | Mysuru Medical College and Research Institute | INSACOG-KA, NIMHANS | Ananthapadmanabha Kotambail; Anita S Desai; Anson Kunjumon George; Chetan G K; Ellango Ramasamy; Gautham Arunachal Udupi; Mahesh Kumar.C.S; Sony Sharma; V Ravi |
| EPI_ISL_4104773, EPI_ISL_4104809, EPI_ISL_4104810, EPI_ISL_4104811, EPI_ISL_4104821, EPI_ISL_4104829, EPI_ISL_4533961, EPI_ISL_4533962, EPI_ISL_4533963, EPI_ISL_4533964 | | | |
| see above | NHL Medical College, Ahmedabad | Gujarat Biotechnology Research Centre | Apurvasinh Puvar; Arpit Shukla; Bhadreshsinh Gohil; Chaitanya Joshi; Dinesh Kumar; Janvi Raval; Jayshri Pethani; Madhvi Joshi; Nimesh Patel; Nitin Savaliya; Nitin Shukla; Priyank Chavda; Ramesh Pandit; Sonal Sharma; Twinkle Soni; Zarna Patel |
| EPI_ISL_4129791, EPI_ISL_4129794, EPI_ISL_4129865, EPI_ISL_4129933, EPI_ISL_4129940, EPI_ISL_4129945 | NIMHANS | INSACOG-KA, NIMHANS | Ananthapadmanabha Kotambail; Anita S Desai; Anson Kunjumon George; Chetan G K; Ellango Ramasamy; Gautham Arunachal Udupi; Mahesh Kumar.C.S; Sony Sharma; V Ravi |
| EPI_ISL_4533551, EPI_ISL_4533552 | Nadiyad Civil Hospital | Gujarat Biotechnology Research Centre | Apurvasinh Puvar; Arpit Shukla; Bhadreshsinh Gohil; Chaitanya Joshi; Dinesh Kumar; Janvi Raval; Madhvi Joshi; Navendu Joshi; Nimesh Patel; Nitin Savaliya; Nitin Shukla; Priyank Chavda; Ramesh Pandit; Sonal Sharma; Zarna Patel |
| EPI_ISL_3717577, EPI_ISL_3717578 | Neuberg Supratech Reference Laboratory, Ahmedabad | Gujarat Biotechnology Research Centre | Arpit Shukla; Bhadreshsinh Gohil; Chaitanya Joshi; Dinesh Kumar; Janvi Raval; Madhvi Joshi; Nimesh Patel; Nitin Savaliya; Nitin Shukla; Ramesh Pandit; Shiva; Sonal Sharma; Twinkle Soni; Zarna Patel |
| EPI_ISL_3833597, EPI_ISL_3833599, EPI_ISL_4104816 | Neuberg Supratech Reference Laboratory, Surat | Gujarat Biotechnology Research Centre | Arpit Shukla; Bhadreshsinh Gohil; Chaitanya Joshi; Dinesh Kumar; Janvi Raval; Madhvi Joshi; Manthan Shah; Nimesh Patel; Nitin Savaliya; Nitin Shukla; Ramesh Pandit; Sonal Sharma; Twinkle Soni; Zarna Patel |
| EPI_ISL_4533968 | Neuberg Supratech Refrence Laboratory, Surat | Gujarat Biotechnology Research Centre | Apurvasinh Puvar; Arpit Shukla; Bhadreshsinh Gohil; Chaitanya Joshi; Dinesh Kumar; Janvi Raval; Madhvi Joshi; Manthan Shah; Nimesh Patel; Nitin Savaliya; Nitin Shukla; Priyank Chavda; Ramesh Pandit; Sonal Sharma; Zarna Patel |
| EPI_ISL_4533918, EPI_ISL_4533919, EPI_ISL_4533920 | Northstar Pathology Lab, Ahmedabad | Gujarat Biotechnology Research Centre | Apurvasinh Puvar; Arpit Shukla; Bhadreshsinh Gohil; Chaitanya Joshi; Dinesh Kumar; Janvi Raval; Jigar Suthar; Madhvi Joshi; Nimesh Patel; Nitin Savaliya; Nitin Shukla; Priyank Chavda; Ramesh Pandit; Sonal Sharma; Zarna Patel |
| EPI_ISL_4058057, EPI_ISL_4058121, EPI_ISL_4058126, EPI_ISL_4058129, EPI_ISL_4058131, EPI_ISL_4058133 | Osmania Medical College | CDFD | Ashwin Dalal; Asmita Gupta; Divya Vashisht; Murali Bashyam; Nagamani Kammili; Vinay Donipadi |
| EPI_ISL_4104774, EPI_ISL_4104796, EPI_ISL_4104797, EPI_ISL_4104798, EPI_ISL_4104799, EPI_ISL_4104801, EPI_ISL_4104803, EPI_ISL_4104826, EPI_ISL_4104827 | | | |
| see above | Pangenomics, Ahmedabad | Gujarat Biotechnology Research Centre | Arpit Shukla; Bhadreshsinh Gohil; Chaitanya Joshi; Dinesh Kumar; Janvi Raval; KM Singh; Madhvi Joshi; Nimesh Patel; Nitin Savaliya; Nitin Shukla; Ramesh Pandit; Sonal Sharma; Twinkle Soni; Zarna Patel |
| EPI_ISL_3948558, EPI_ISL_3948559, EPI_ISL_3948564, EPI_ISL_3948565, EPI_ISL_3948566, EPI_ISL_3948567, EPI_ISL_3948568, EPI_ISL_3948569 | | | |
| see above | Pathocare Pathology Laboratory, Vadodara | Gujarat Biotechnology Research Centre | Arpit Shukla; Bhadreshsinh Gohil; Chaitanya Joshi; Dinesh Kumar; Janvi Raval; Madhvi Joshi; Mital Vakani; Nimesh Patel; Nitin Savaliya; Nitin Shukla; Ramesh Pandit; Sonal Sharma; Twinkle Soni; Zarna Patel |
| EPI_ISL_4533965, EPI_ISL_4533966 | Rivaara Lab, Ahmedabad | Gujarat Biotechnology Research Centre | Apurvasinh Puvar; Arpit Shukla; B Sharan Sharma; Bhadreshsinh Gohil; Chaitanya Joshi; Dinesh Kumar; Janvi Raval; Madhvi Joshi; Nimesh Patel; Nitin Savaliya; Nitin Shukla; Priyank Chavda; Ramesh Pandit; Sonal Sharma; Zarna Patel |
| EPI_ISL_4533553 | S.S.G Hospital ,Vadodara | Gujarat Biotechnology Research Centre | Apurvasinh Puvar; Arpit Shukla; Bhadreshsinh Gohil; Chaitanya Joshi; Dinesh Kumar; Janvi Raval; Jigna Karia; Madhvi Joshi; Nimesh Patel; Nitin Savaliya; Nitin Shukla; Priyank Chavda; Ramesh Pandit; Sonal Sharma; Zarna Patel |
| EPI_ISL_4533555, EPI_ISL_4533557, EPI_ISL_4533558, EPI_ISL_4533559, EPI_ISL_4533560, EPI_ISL_4533935, EPI_ISL_4533978, EPI_ISL_4533980, EPI_ISL_4533981 | | | |
| see above | SN Genelab Pvt. Ltd, Surat | Gujarat Biotechnology Research Centre | Apurvasinh Puvar; Arpit Shukla; Bhadreshsinh Gohil; Chaitanya Joshi; Dinesh Kumar; Janvi Raval; Madhvi Joshi; Nimesh Patel; Nirmal A Vaniwala; Nitin Savaliya; Nitin Shukla; Priyank Chavda; Ramesh Pandit; Sonal Sharma; Zarna Patel |

| | | | |
|---|---|---|---|
| EPI_ISL_4533956, EPI_ISL_4533957 | Scientific Diagnostic Centre Pvt. Ltd., Ahmedabad | Gujarat Biotechnology Research Centre | Apurvasinh Puvar; Arpit Shukla; Bhadreshsinh Gohil; Chaitanya Joshi; Dinesh Kumar; Janvi Raval; Madhvi Joshi; Moksha B Narechania; Nimesh Patel; Nitin Savaliya; Nitin Shukla; Priyank Chavda; Ramesh Pandit; Sonal Sharma; Zarna Patel |
| EPI_ISL_3948556, EPI_ISL_4533921 | Shraddhadeep Green Cross Pathology Laboratory, Gandhinagar | Gujarat Biotechnology Research Centre | Apurvasinh Puvar; Arpit Shukla; Bhadreshsinh Gohil; Chaitanya Joshi; Dinesh Kumar; Dinesh Rathod; Janvi Raval; Krupa Shah; Madhvi Joshi; Nimesh Patel; Nitin Savaliya; Nitin Shukla; Priyank Chavda; Ramesh Pandit; Sonal Sharma; Twinkle Soni; Zarna Patel |
| EPI_ISL_4533911, EPI_ISL_4533912, EPI_ISL_4533913, EPI_ISL_4533914, EPI_ISL_4533915, EPI_ISL_4533916, EPI_ISL_4533917 | | | |
| see above | Smimer Medical College ,Surat | Gujarat Biotechnology Research Centre | Apurvasinh Puvar; Arpit Shukla; Bhadreshsinh Gohil; Chaitanya Joshi; Dinesh Kumar; Janvi Raval; Madhvi Joshi; Manish Patel; Nimesh Patel; Nitin Savaliya; Nitin Shukla; Priyank Chavda; Ramesh Pandit; Sonal Sharma; Zarna Patel |
| EPI_ISL_4533543, EPI_ISL_4533951, EPI_ISL_4533952, EPI_ISL_4533953, EPI_ISL_4533954 | Sterling Accuris, Bhavnagar | Gujarat Biotechnology Research Centre | Apurvasinh Puvar; Arpit Shukla; Bhadreshsinh Gohil; Chaitanya Joshi; Dinesh Kumar; Hemal Salot; Janvi Raval; Madhvi Joshi; Nimesh Patel; Nitin Savaliya; Nitin Shukla; Priyank Chavda; Ramesh Pandit; Sonal Sharma; Zarna Patel |
| EPI_ISL_4533955 | Sunflower Laboratory, Ahmedabad | Gujarat Biotechnology Research Centre | Apurvasinh Puvar; Arpit Shukla; Bhadreshsinh Gohil; Chaitanya Joshi; Dinesh Kumar; Janvi Raval; Madhvi Joshi; Mantrit Kaur; Nimesh Patel; Nitin Savaliya; Nitin Shukla; Priyank Chavda; Ramesh Pandit; Sonal Sharma; Zarna Patel |
| EPI_ISL_3948549, EPI_ISL_3948550, EPI_ISL_3948551, EPI_ISL_3948552, EPI_ISL_3948555, EPI_ISL_4104775, EPI_ISL_4104778, EPI_ISL_4104779, EPI_ISL_4104780, EPI_ISL_4104814, EPI_ISL_4533545, EPI_ISL_4533548, EPI_ISL_4533974, EPI_ISL_4533975, EPI_ISL_4533977 | | | |
| see above | Supratech Micropath Laboratory Research Institute Pvt Ltd, Ahmedabad | Gujarat Biotechnology Research Centre | Apurvasinh Puvar; Arpit Shukla; Bhadreshsinh Gohil; Chaitanya Joshi; Dinesh Kumar; Janvi Raval; Madhvi Joshi; Mahendra Parikh; Nimesh Patel; Nitin Savaliya; Nitin Shukla; Priyank Chavda; Ramesh Pandit; Shiva; Sonal Sharma; Twinkle Soni; Zarna Patel |
| EPI_ISL_4104786, EPI_ISL_4104787, EPI_ISL_4533542, EPI_ISL_4533943, EPI_ISL_4533947, EPI_ISL_4533948, EPI_ISL_4533950 | | | |
| see above | Toprani Advance Lab system,Vadodara | Gujarat Biotechnology Research Centre | Apurvasinh Puvar; Arpit Shukla; Bhadreshsinh Gohil; Chaitanya Joshi; Dinesh Kumar; Janvi Raval; Madhvi Joshi; Nimesh Patel; Nitin Savaliya; Nitin Shukla; Priyank Chavda; Ramesh Pandit; Sonal Sharma; Tushar Toprani; Twinkle Soni; Zarna Patel |
| EPI_ISL_4415442, EPI_ISL_4415443, EPI_ISL_4415444, EPI_ISL_4415445, EPI_ISL_4415446, EPI_ISL_4415449, EPI_ISL_4415451, EPI_ISL_4415452, EPI_ISL_4415544, EPI_ISL_4415552, EPI_ISL_4415554, EPI_ISL_4415558, EPI_ISL_4415559, EPI_ISL_4415562, EPI_ISL_4415565, EPI_ISL_4415568, EPI_ISL_4415569, EPI_ISL_4415570, EPI_ISL_4415571, EPI_ISL_4415576, EPI_ISL_4415577, EPI_ISL_4415582, EPI_ISL_4415583, EPI_ISL_4415584, EPI_ISL_4415586, EPI_ISL_4415588, EPI_ISL_4415590, EPI_ISL_4415595, EPI_ISL_4415596, EPI_ISL_4415603, EPI_ISL_4415611, EPI_ISL_4415615, EPI_ISL_4415617, EPI_ISL_4415625, EPI_ISL_4415630, EPI_ISL_4415637, EPI_ISL_4415654, EPI_ISL_4415656, EPI_ISL_4415662, EPI_ISL_4415663, EPI_ISL_4415665, EPI_ISL_4415668, EPI_ISL_4415670, EPI_ISL_4415682, EPI_ISL_4415687, EPI_ISL_4415688, EPI_ISL_4415690, EPI_ISL_4415693, EPI_ISL_4415694, EPI_ISL_4415701, EPI_ISL_4415703, EPI_ISL_4415705, EPI_ISL_4415709, EPI_ISL_4415710, EPI_ISL_4415711, EPI_ISL_4415714, EPI_ISL_4415715, EPI_ISL_4415716, EPI_ISL_4415718, EPI_ISL_4415722, EPI_ISL_4415725, EPI_ISL_4415726, EPI_ISL_4415728, EPI_ISL_4415729, EPI_ISL_4415740, EPI_ISL_4415743, EPI_ISL_4415745, EPI_ISL_4415750, EPI_ISL_4415752, EPI_ISL_4415754, EPI_ISL_4415758, EPI_ISL_4415761, EPI_ISL_4415762, EPI_ISL_4415764 | | | |
| see above | Virus Research & Diagnostic Laboratory | IISER Pune-INSACOG | Aurnab Ghose; Joy Merwin Monteiro; Krishanpal Karmodiya |
| EPI_ISL_4129648 | bangalore Medical college and Research Institute | INSACOG-KA, NIMHANS | Ananthapadmanabha Kotambail; Anita S Desai; Anson Kunjumon George; Chetan G K; Ellango Ramasamy; Gautham Arunachal Udupi; Mahesh Kumar.C.S; Sony Sharma; V Ravi |
