## Supplementary material for "SARS-COV-2 δ variant drives the pandemic in India and Europe via two subvariants": Acknowledgement table on the GISAID genomes used in this study

We gratefully acknowledge the following Authors from the Originating laboratories responsible for obtaining the specimens, as well as the Submitting laboratories where the genome data were generated and shared via GISAID, on which this research is based.

All Submitters of data may be contacted directly via www.gisaid.org

Authors are sorted alphabetically.

| Accession ID | Originating Laboratory | Submitting Laboratory | Authors |
|---|---|---|---|
| EPI_ISL_3935229, EPI_ISL_3935253, EPI_ISL_3935264, EPI_ISL_4026639, EPI_ISL_4545905, EPI_ISL_4545942, EPI_ISL_4546165, EPI_ISL_4546189, EPI_ISL_4546195, EPI_ISL_4546481, EPI_ISL_4546482, EPI_ISL_4546492 | | | |
| see above | AP SSO | CSIR-Centre for Cellular and Molecular Biology - INSACOG | Amareshwar Vodapalli; Ara Sreenivas; Archana Bharadwaj Siva; B Himasri; Divya Tej Sowpati; Jandhyala Sai Krishna; Karthik Bharadwaj Tallapaka; Lamuk Zaveri; Onkar Kulkarni; Payel Mukherjee; Priya Nurkurthy; Rakesh K Mishra; Shreekant Verma; Sofia Banu; Sumedha Avadhanula; Tulasi Nagabandi; Valli Nagalakshmi Undamatla; Vidhyadhari Methuku |
| EPI_ISL_4415458, EPI_ISL_4415461, EPI_ISL_4415463, EPI_ISL_4415466, EPI_ISL_4415467, EPI_ISL_4415469, EPI_ISL_4415471, EPI_ISL_4415472, EPI_ISL_4415475, EPI_ISL_4415485, EPI_ISL_4415486, EPI_ISL_4415498, EPI_ISL_4415517, EPI_ISL_4415518, EPI_ISL_4415521, EPI_ISL_4415525, EPI_ISL_4415594, EPI_ISL_4415602, EPI_ISL_4415618, EPI_ISL_4415626 | | | |
| see above | BJMC, Pune | IISER Pune-INSACOG | Aurnab Ghose; Joy Merwin Monteiro; Krishanpal Karmodiya; Rajesh Karyakarte; Suvarna Joshi |
| EPI_ISL_4058169 | CDFD | CDFD | Ashwin Dalal; Asmita Gupta; Divya Vashisht; Murali Bashyam; Nagamani Kammili; Vinay Donipadi |
| EPI_ISL_4545995 | CSIR-Central Leather Research Institute | CSIR-Centre for Cellular and Molecular Biology - INSACOG | Amareshwar Vodapalli; Ara Sreenivas; Archana Bharadwaj Siva; B Himasri; Divya Tej Sowpati; Jandhyala Sai Krishna; Karthik Bharadwaj Tallapaka; Lamuk Zaveri; Onkar Kulkarni; Payel Mukherjee; Priya Nurkurthy; Rakesh K Mishra; Shreekant Verma; Sofia Banu; Sumedha Avadhanula; Tulasi Nagabandi; Valli Nagalakshmi Undamatla; Vidhyadhari Methuku |
| EPI_ISL_4545899 | CSIR-Centre for Cellular and Molecular Biology - INSACOG | CSIR-Centre for Cellular and Molecular Biology - INSACOG | Amareshwar Vodapalli; Ara Sreenivas; Archana Bharadwaj Siva; B Himasri; Divya Tej Sowpati; Jandhyala Sai Krishna; Karthik Bharadwaj Tallapaka; Lamuk Zaveri; Onkar Kulkarni; Payel Mukherjee; Priya Nurkurthy; Rakesh K Mishra; Shreekant Verma; Sofia Banu; Sumedha Avadhanula; Tulasi Nagabandi; Valli Nagalakshmi Undamatla; Vidhyadhari Methuku |
| EPI_ISL_4546056, EPI_ISL_4546080, EPI_ISL_4546093, EPI_ISL_4546105, EPI_ISL_4546381, EPI_ISL_4546422 | CSIR-National Environmental Engineering Research Institute | CSIR-Centre for Cellular and Molecular Biology - INSACOG | Amareshwar Vodapalli; Ara Sreenivas; Archana Bharadwaj Siva; B Himasri; Divya Tej Sowpati; Jandhyala Sai Krishna; Karthik Bharadwaj Tallapaka; Krishna Khairnar; Lamuk Zaveri; Onkar Kulkarni; Payel Mukherjee; Priya Nurkurthy; Rakesh K Mishra; Shreekant Verma; Sofia Banu; Sumedha Avadhanula; Tulasi Nagabandi; Valli Nagalakshmi Undamatla; Vidhyadhari Methuku |
| EPI_ISL_4415406, EPI_ISL_4415407, EPI_ISL_4415601, EPI_ISL_4415657, EPI_ISL_4415734 | Department of Microbiology, Government Medical College, Baramati | IISER Pune-INSACOG | Aurnab Ghose; Joy Merwin Monteiro; Krishanpal Karmodiya |
| EPI_ISL_4533958 | Desai Metropolis Health Service Pvt. Ltd, surat | Gujarat Biotechnology Research Centre | Apurvasinh Puvar; Arpit Shukla; Bhadreshsinh Gohil; Chaitanya Joshi; Dinesh Kumar; Janvi Raval; Madhvi Joshi; Manali Shah; Nimesh Patel; Nitin Savaliya; Nitin Shukla; Priyank Chavda; Ramesh Pandit; Sonal Sharma; Zarna Patel |
| EPI_ISL_4533928 | GMERS Medical College, Gandhinagar | Gujarat Biotechnology Research Centre | Apurvasinh Puvar; Arpit Shukla; Bhadreshsinh Gohil; Chaitanya Joshi; Dinesh Kumar; Gaurishankar Shrimali; Janvi Raval; Madhvi Joshi; Nimesh Patel; Nitin Savaliya; Nitin Shukla; Priyank Chavda; Ramesh Pandit; Sonal Sharma; Zarna Patel |
| EPI_ISL_4058084 | Gandhi Hospital | CDFD | Ashwin Dalal; Asmita Gupta; Divya Vashisht; Murali Bashyam; Nagamani Kammili; Vinay Donipadi |
| EPI_ISL_4058054, EPI_ISL_4058153, EPI_ISL_4058170, EPI_ISL_4058241 | Gandhi Medical College | CDFD | Ashwin Dalal; Asmita Gupta; Divya Vashisht; Murali Bashyam; Nagamani Kammili; Vinay Donipadi |
| EPI_ISL_4104784, EPI_ISL_4104785 | General Hospital, Chotaudaipur | Gujarat Biotechnology Research Centre | Arpit Shukla; Arti Thakur; Bhadreshsinh Gohil; Chaitanya Joshi; Dinesh Kumar; Janvi Raval; Madhvi Joshi; Nimesh Patel; Nitin Savaliya; Nitin Shukla; Ramesh Pandit; Sonal Sharma; Twinkle Soni; Zarna Patel |
| EPI_ISL_4533931 | General Hospital, Nadiad | Gujarat Biotechnology Research Centre | Apurvasinh Puvar; Arpit Shukla; Bhadreshsinh Gohil; Chaitanya Joshi; Dinesh Kumar; Janvi Raval; Madhvi Joshi; Navendu Joshi; Nimesh Patel; Nitin Savaliya; Nitin Shukla; Priyank Chavda; Ramesh Pandit; Sonal Sharma; Zarna Patel |
| EPI_ISL_4058098, EPI_ISL_4058106, EPI_ISL_4058110, EPI_ISL_4058112, EPI_ISL_4058114, EPI_ISL_4058194 | Government Fever Hospital | CDFD | Ashwin Dalal; Asmita Gupta; Divya Vashisht; Murali Bashyam; Nagamani Kammili; Vinay Donipadi |
| EPI_ISL_4058206, EPI_ISL_4058210 | Government General Hospital | CDFD | Ashwin Dalal; Asmita Gupta; Divya Vashisht; Murali Bashyam; Nagamani Kammili; Vinay Donipadi |
| EPI_ISL_4058059, EPI_ISL_4058243, EPI_ISL_4058247, EPI_ISL_4058248, EPI_ISL_4058254 | Government Maternity Hospital | CDFD | Ashwin Dalal; Asmita Gupta; Divya Vashisht; Murali Bashyam; Nagamani Kammili; Vinay Donipadi |
| EPI_ISL_3982476, EPI_ISL_3982482, EPI_ISL_3982484, EPI_ISL_3982491, EPI_ISL_3982494, EPI_ISL_3982496, EPI_ISL_3982501, EPI_ISL_3982505, EPI_ISL_3982514, EPI_ISL_3982519, EPI_ISL_3982520, EPI_ISL_3982525, EPI_ISL_3982532, EPI_ISL_3982533, EPI_ISL_3982534, EPI_ISL_3982536, EPI_ISL_3982537, EPI_ISL_3982539, EPI_ISL_3982542, EPI_ISL_3982543, EPI_ISL_3982544, EPI_ISL_3982551, EPI_ISL_3982553, EPI_ISL_3982556, EPI_ISL_3982559, EPI_ISL_3982572, EPI_ISL_3982573, EPI_ISL_3982574, EPI_ISL_3982575, EPI_ISL_3982576, EPI_ISL_3982577, EPI_ISL_3982582 | | | |
| see above | ICMR-National Institute of Virology - INSACOG | NIV Influenza | Dr. Varsha Potdar and NIC Team |
| EPI_ISL_3982988, EPI_ISL_3982989, EPI_ISL_3982997 | INSACOG-Assam | National Institute of Biomedical Genomics – INSACOG | Ajanta Sharma; Arindam Maitra; Kailash Chamuah; Lahari Saikia; Nidhan Kumar Biswas; Saumitra Das; Sreedhar Chinnaswamy |
| EPI_ISL_4504800 | INSACOG-MANIPUR | National Institute of Biomedical Genomics – INSACOG | Arindam Maitra; Kh. Ranjana Devi; L. Shivadutta Singh; Nidhan Kumar Biswas; R.K.Manojkumar Singh; Saumitra Das; Sreedhar Chinnaswamy |
| EPI_ISL_4504569, EPI_ISL_4504572, EPI_ISL_4504602, EPI_ISL_4504603, EPI_ISL_4504608, EPI_ISL_4504610, EPI_ISL_4504612, EPI_ISL_4504619, EPI_ISL_4504623, EPI_ISL_4504626, EPI_ISL_4504630, EPI_ISL_4504643, EPI_ISL_4504644, EPI_ISL_4504646 | | | |
| see above | INSACOG-Sikkim | National Institute of Biomedical Genomics – INSACOG | Arindam Maitra; Dr. Shrijana Gurung; Kaden Zangmu Bhutia; Nidhan Kumar Biswas; Saumitra Das; Sreedhar Chinnaswamy Tshering Ongmu Bhutia |
| EPI_ISL_4504198, EPI_ISL_4504209, EPI_ISL_4504211, EPI_ISL_4504223, EPI_ISL_4504233, EPI_ISL_4504235, EPI_ISL_4504274, EPI_ISL_4504306, EPI_ISL_4504308, EPI_ISL_4504313, EPI_ISL_4504345, EPI_ISL_4504407, EPI_ISL_4504413, EPI_ISL_4504416, EPI_ISL_4504449, EPI_ISL_4504453, EPI_ISL_4504487, EPI_ISL_4504490, EPI_ISL_4504494, EPI_ISL_4504526, EPI_ISL_4504527, EPI_ISL_4504531, EPI_ISL_4504532, EPI_ISL_4504533, EPI_ISL_4504543, EPI_ISL_4504553, EPI_ISL_4504562 | | | |
| see above | INSACOG-WB | National Institute of Biomedical Genomics – INSACOG | Ajay Chakraborti; Arindam Maitra; Bhaswati Bandyopadhyay; Nidhan Kumar Biswas; Saumitra Das; Sreedhar Chinnaswamy; Tamal Ghosh |
| EPI_ISL_4193024, EPI_ISL_4193025, EPI_ISL_4193026, EPI_ISL_4193028, EPI_ISL_4193030, EPI_ISL_4193033, EPI_ISL_4193062, EPI_ISL_4193071, EPI_ISL_4193072, EPI_ISL_4193212, EPI_ISL_4193260, EPI_ISL_4193263, EPI_ISL_4193367, EPI_ISL_4193455, EPI_ISL_4193459, EPI_ISL_4193468, EPI_ISL_4193483, EPI_ISL_4193485, EPI_ISL_4193490 | | | |
| see above | INSACOG-WB | National Institute of Biomedical Genomics – INSACOG | Ajay Chakraborti; Arindam Maitra; Bhaswati Bandyopadhyay; Nidhan Kumar Biswas; Saumitra Das; Sreedhar Chinnaswamy; Tamal Ghosh |
| EPI_ISL_4312134, EPI_ISL_4312540, EPI_ISL_4312977, EPI_ISL_4312989 | INSACOG-WB | National Institute of Biomedical Genomics – INSACOG | Ajay Chakraborti; Arindam Maitra; Bhaswati Bandyopadhyay; Nidhan Kumar Biswas; Saumitra Das; Sreedhar Chinnaswamy; Tamal Ghosh |
| EPI_ISL_4058156, EPI_ISL_4058160, EPI_ISL_4058225, EPI_ISL_4058231, EPI_ISL_4058235, EPI_ISL_4058236 | Institute of Preventive Medicine | CDFD | Ashwin Dalal; Asmita Gupta; Divya Vashisht; Murali Bashyam; Nagamani Kammili; Vinay Donipadi |
| EPI_ISL_3833605, EPI_ISL_3905543 | Invitro Speciality laboratory, Ahmedabad | Gujarat Biotechnology Research Centre | Arpit Shukla; Bhadreshsinh Gohil; Chaitanya Joshi; Dinesh Kumar; Janvi Raval; Madhvi Joshi; Nimesh Patel; Nitin Savaliya; Nitin Shukla; Ramesh Pandit; Sonal Sharma; Twinkle Soni; Viral N Pathak; Zarna Patel |
| EPI_ISL_4558045, EPI_ISL_4558046, EPI_ISL_4558052 | MEDICA Superspeciality Hospital | CSIR-Indian Institute of Chemical Biology | Arpita Ghosh Mitra; Arun Bandyopadhyay; Aviral Roy; Partha Chakrabarti; Poulomi Sarkar; Saikat Chakrabarti; Sarthak Banerjee; Siddik Sarkar; Soumen Saha |
| EPI_ISL_3935172, EPI_ISL_3935196, EPI_ISL_3935233, EPI_ISL_3935313 | Mapmygenome | CSIR-Centre for Cellular and Molecular Biology - INSACOG | Amareshwar Vodapalli; Ara Sreenivas; Archana Bharadwaj Siva; B Himasri; Divya Tej Sowpati; Jandhyala Sai Krishna; Karthik Bharadwaj Tallapaka; Lamuk Zaveri; Onkar Kulkarni; Payel Mukherjee; Priya Nurkurthy; Rakesh K Mishra; Shreekant Verma; Sofia Banu; Sumedha Avadhanula; Tulasi Nagabandi; Valli Nagalakshmi Undamatla; Vidhyadhari Methuku |
| EPI_ISL_4546241 | Mapmygenome India LTD | CSIR-Centre for Cellular and Molecular Biology - INSACOG | Amareshwar Vodapalli; Ara Sreenivas; Archana Bharadwaj Siva; B Himasri; Divya Tej Sowpati; Jandhyala Sai Krishna; Karthik Bharadwaj Tallapaka; Lamuk Zaveri; Onkar Kulkarni; Payel Mukherjee; Priya Nurkurthy; Rakesh K Mishra; Shreekant Verma; Sofia Banu; Sumedha Avadhanula; Tulasi Nagabandi; Valli Nagalakshmi Undamatla; Vidhyadhari Methuku |
| EPI_ISL_4129760, EPI_ISL_4129822, EPI_ISL_4129867 | Mysuru Medical College and Research Institute | INSACOG-KA, NIMHANS | Ananthapadmanabha Kotambail; Anita S Desai; Anson Kunjumon George; Chetan G K; Ellango Ramasamy; Gautham Arunachal Udupi; Mahesh Kumar.C.S; Sony Sharma; V Ravi |
| EPI_ISL_4104808 | NHL Medical College, Ahmedabad | Gujarat Biotechnology Research Centre | Arpit Shukla; Bhadreshsinh Gohil; Chaitanya Joshi; Dinesh Kumar; Janvi Raval; Jayshri Pethani; Madhvi Joshi; Nimesh Patel; Nitin Savaliya; Nitin Shukla; Ramesh Pandit; Sonal Sharma; Twinkle Soni; Zarna Patel |
| EPI_ISL_4129796 | NIMHANS | INSACOG-KA, NIMHANS | Ananthapadmanabha Kotambail; Anita S Desai; Anson Kunjumon George; Chetan G K; Ellango Ramasamy; Gautham Arunachal Udupi; Mahesh Kumar.C.S; Sony Sharma; V Ravi |
| EPI_ISL_3717584 | Neuberg Supratech Reference Laboratory, Ahmedabad | Gujarat Biotechnology Research Centre | Arpit Shukla; Bhadreshsinh Gohil; Chaitanya Joshi; Dinesh Kumar; Janvi Raval; Madhvi Joshi; Nimesh Patel; Nitin Savaliya; Nitin Shukla; Ramesh Pandit; Shiva; Sonal Sharma; Twinkle Soni; Zarna Patel |
| EPI_ISL_3833598, EPI_ISL_3833607, EPI_ISL_4104817, EPI_ISL_4104820, EPI_ISL_4104830 | Neuberg Supratech Reference Laboratory, Surat | Gujarat Biotechnology Research Centre | Arpit Shukla; Bhadreshsinh Gohil; Chaitanya Joshi; Dinesh Kumar; Janvi Raval; Madhvi Joshi; Manthan Shah; Nimesh Patel; Nitin Savaliya; Nitin Shukla; Ramesh Pandit; Sonal Sharma; Twinkle Soni; Zarna Patel |
| EPI_ISL_4058124, EPI_ISL_4058127, EPI_ISL_4058134, EPI_ISL_4058136, EPI_ISL_4058150, EPI_ISL_4058151, EPI_ISL_4058189 | | | |
| see above | Osmania Medical College | CDFD | Ashwin Dalal; Asmita Gupta; Divya Vashisht; Murali Bashyam; Nagamani Kammili; Vinay Donipadi |
| EPI_ISL_4104774, EPI_ISL_4104802, EPI_ISL_4104828 | Pangenomics, Ahmedabad | Gujarat Biotechnology Research Centre | Arpit Shukla; Bhadreshsinh Gohil; Chaitanya Joshi; Dinesh Kumar; Janvi Raval; KM Singh; Madhvi Joshi; Nimesh Patel; Nitin Savaliya; Nitin Shukla; Ramesh Pandit; Sonal Sharma; Twinkle Soni; Zarna Patel |
| EPI_ISL_3948560, EPI_ISL_3948561, EPI_ISL_3948562, EPI_ISL_3948563 | Pathocare Pathology Laboratory, Vadodara | Gujarat Biotechnology Research Centre | Arpit Shukla; Bhadreshsinh Gohil; Chaitanya Joshi; Dinesh Kumar; Janvi Raval; Madhvi Joshi; Mital Vakani; Nimesh Patel; Nitin Savaliya; Nitin Shukla; Ramesh Pandit; Sonal Sharma; Twinkle Soni; Zarna Patel |
| EPI_ISL_4533556, EPI_ISL_4533934, EPI_ISL_4533936, EPI_ISL_4533980 | SN Genelab Pvt. Ltd, Surat | Gujarat Biotechnology Research Centre | Apurvasinh Puvar; Arpit Shukla; Bhadreshsinh Gohil; Chaitanya Joshi; Dinesh Kumar; Janvi Raval; Madhvi Joshi; Nimesh Patel; Nirmal A Vaniwala; Nitin Savaliya; Nitin Shukla; Priyank Chavda; Ramesh Pandit; Sonal Sharma; Zarna Patel |
| EPI_ISL_3717579, EPI_ISL_3717580 | SSG Hospital Vadodara | Gujarat Biotechnology Research Centre | Arpit Shukla; Bhadreshsinh Gohil; Chaitanya Joshi; Dinesh Kumar; Janvi Raval; Jigna Karia; Madhvi Joshi; Nimesh Patel; Nitin Savaliya; Nitin Shukla; Ramesh Pandit; Sonal Sharma; Twinkle Soni; Zarna Patel |
| EPI_ISL_3948557 | Shraddhadeep Green Cross Pathology Laboratory, Gandhinagar | Gujarat Biotechnology Research Centre | Arpit Shukla; Bhadreshsinh Gohil; Chaitanya Joshi; Dinesh Kumar; Janvi Raval; Krupa Shah; Madhvi Joshi; Nimesh Patel; Nitin Savaliya; Nitin Shukla; Ramesh Pandit; Sonal Sharma; Twinkle Soni; Zarna Patel |

| | | | |
|---|---|---|---|
| EPI_ISL_4533907, EPI_ISL_4533908, EPI_ISL_4533909, EPI_ISL_4533910 | Smimer Medical College ,Surat | Gujarat Biotechnology Research Centre | Apurvasinh Puvar; Arpit Shukla; Bhadreshsinh Gohil; Chaitanya Joshi; Dinesh Kumar; Janvi Raval; Madhvi Joshi; Manish Patel; Nimesh Patel; Nitin Savaliya; Nitin Shukla; Priyank Chavda; Ramesh Pandit; Sonal Sharma; Zarna Patel |
| EPI_ISL_3717581 | Sterling Accuris Vadodara | Gujarat Biotechnology Research Centre | Arpit Shukla; Bhadreshsinh Gohil; Chaitanya Joshi; Dinesh Kumar; Janvi Raval; Jyoti Patankar; Madhvi Joshi; Nimesh Patel; Nitin Savaliya; Nitin Shukla; Ramesh Pandit; Sonal Sharma; Twinkle Soni; Zarna Patel |
| EPI_ISL_4533544 | Sterling Accuris, Surat | Gujarat Biotechnology Research Centre | Apurvasinh Puvar; Arpit Shukla; Bhadreshsinh Gohil; Chaitanya Joshi; Dinesh Kumar; Gaurav Shashtri; Janvi Raval; Madhvi Joshi; Nimesh Patel; Nitin Savaliya; Nitin Shukla; Priyank Chavda; Ramesh Pandit; Sonal Sharma; Zarna Patel |
| EPI_ISL_3948554, EPI_ISL_4104815, EPI_ISL_4533546, EPI_ISL_4533549, EPI_ISL_4533976 | Supratech Micropath Laboratory Research Institute Pvt Ltd, Ahmedabad | Gujarat Biotechnology Research Centre | Apurvasinh Puvar; Arpit Shukla; Bhadreshsinh Gohil; Chaitanya Joshi; Dinesh Kumar; Janvi Raval; Madhvi Joshi; Mahendra Parikh; Nimesh Patel; Nitin Savaliya; Nitin Shukla; Priyank Chavda; Ramesh Pandit; Shiva; Sonal Sharma; Twinkle Soni; Zarna Patel |
| EPI_ISL_4104823, EPI_ISL_4533944, EPI_ISL_4533945, EPI_ISL_4533946 | Toprani Advance Lab system,Vadodara | Gujarat Biotechnology Research Centre | Apurvasinh Puvar; Arpit Shukla; Bhadreshsinh Gohil; Chaitanya Joshi; Dinesh Kumar; Janvi Raval; Madhvi Joshi; Nimesh Patel; Nitin Savaliya; Nitin Shukla; Priyank Chavda; Ramesh Pandit; Sonal Sharma; Tushar Toprani; Twinkle Soni; Zarna Patel |
| EPI_ISL_4415447, EPI_ISL_4415450, EPI_ISL_4415453, EPI_ISL_4415537, EPI_ISL_4415575, EPI_ISL_4415674, EPI_ISL_4415675, EPI_ISL_4415741 | | | |
| see above | Virus Research & Diagnostic Laboratory | IISER Pune-INSACOG | Aurnab Ghose; Joy Merwin Monteiro; Krishanpal Karmodiya |
| EPI_ISL_4129629, EPI_ISL_4129638, EPI_ISL_4129658 | bangalore Medical college and Research Institute | INSACOG-KA, NIMHANS | Ananthapadmanabha Kotambail; Anita S Desai; Anson Kunjumon George; Chetan G K; Ellango Ramasamy; Gautham Arunachal Udupi; Mahesh Kumar.C.S; Sony Sharma; V Ravi |
