## Supplementary material for "SARS-COV-2 δ variant drives the pandemic in India and Europe via two subvariants": Acknowledgement table on the GISAID genomes used in this study

We gratefully acknowledge the following Authors from the Originating laboratories responsible for obtaining the specimens, as well as the Submitting laboratories where the genome data were generated and shared via GISAID, on which this research is based.

All Submitters of data may be contacted directly via www.gisaid.org

Authors are sorted alphabetically.

| Accession ID | Originating Laboratory | Submitting Laboratory | Authors |
|---|---|---|---|
| EPI_ISL_4584508, EPI_ISL_4584588, EPI_ISL_4584693, EPI_ISL_4584774, EPI_ISL_4584806, EPI_ISL_4584807, EPI_ISL_4584880, EPI_ISL_4585033, EPI_ISL_4586436, EPI_ISL_4586468, EPI_ISL_4586653, EPI_ISL_4586697, EPI_ISL_4586699, EPI_ISL_4586736, EPI_ISL_4586779, EPI_ISL_4586886, EPI_ISL_4586942, EPI_ISL_4587054, EPI_ISL_4587157, EPI_ISL_4589756, EPI_ISL_4589837, EPI_ISL_4589944, EPI_ISL_4589951, EPI_ISL_4590166, EPI_ISL_4590422, EPI_ISL_4590516, EPI_ISL_4590543, EPI_ISL_4591135, EPI_ISL_4591670, EPI_ISL_4591674, EPI_ISL_4591688, EPI_ISL_4591739, EPI_ISL_4591779, EPI_ISL_4592410, EPI_ISL_4592458, EPI_ISL_4592586, EPI_ISL_4592608, EPI_ISL_4592677, EPI_ISL_4592742, EPI_ISL_4592828, EPI_ISL_4592914, EPI_ISL_4592959, EPI_ISL_4592960, EPI_ISL_4593004, EPI_ISL_4593031, EPI_ISL_4593088, EPI_ISL_4593159, EPI_ISL_4593895, EPI_ISL_4596560, EPI_ISL_4618837, EPI_ISL_4618854, EPI_ISL_4619081, EPI_ISL_4619352, EPI_ISL_4619392, EPI_ISL_4619566, EPI_ISL_4619584, EPI_ISL_4621276, EPI_ISL_4623293 | | | |
| see above | Berkshire and Surrey Pathology Services Lighthouse Laboratory | Wellcome Sanger Institute for the COVID-19 Genomics UK (COG-UK) Consortium | Berkshire and Surrey Pathology Services Lighthouse Laboratory and Alex Alderton; Cordelia Langford; David K. Jackson; Dominic Kwiatkowski; Ewan Harrison; Ian Johnston; Jeffrey Barrett; John Sillitoe on behalf of the Wellcome Sanger Institute COVID-19 Surveillance Team; Roberto Amato; Sonia Goncalves |
| EPI_ISL_4674921, EPI_ISL_4674974, EPI_ISL_4675533, EPI_ISL_4675566, EPI_ISL_4675581, EPI_ISL_4675713, EPI_ISL_4676227, EPI_ISL_4676237, EPI_ISL_4676238, EPI_ISL_4676364, EPI_ISL_4676524 | | | |
| see above | Health Services Laboratories | Wellcome Sanger Institute for the COVID-19 Genomics UK (COG-UK) Consortium | Cordelia Langford; David K. Jackson; Dominic Kwiatkowski; Ewan Harrison; Health Services Laboratories and Alex Alderton; Ian Johnston; Jeffrey Barrett; John Sillitoe on behalf of the Wellcome Sanger Institute COVID-19 Surveillance Team; Roberto Amato; Sonia Goncalves |
| EPI_ISL_4585211, EPI_ISL_4585325, EPI_ISL_4585476, EPI_ISL_4585596, EPI_ISL_4587236, EPI_ISL_4587237, EPI_ISL_4587241, EPI_ISL_4587353, EPI_ISL_4587357, EPI_ISL_4587364, EPI_ISL_4587545, EPI_ISL_4587575, EPI_ISL_4587590, EPI_ISL_4587612, EPI_ISL_4587660, EPI_ISL_4587687, EPI_ISL_4587711, EPI_ISL_4587795, EPI_ISL_4587859, EPI_ISL_4587872, EPI_ISL_4587894, EPI_ISL_4587934, EPI_ISL_4587935, EPI_ISL_4588827, EPI_ISL_4588841, EPI_ISL_4588866, EPI_ISL_4588877, EPI_ISL_4588889, EPI_ISL_4588932, EPI_ISL_4588957, EPI_ISL_4588994, EPI_ISL_4589037, EPI_ISL_4589075, EPI_ISL_4589248, EPI_ISL_4596187, EPI_ISL_4596202, EPI_ISL_4596469, EPI_ISL_4597332, EPI_ISL_4597336, EPI_ISL_4597388, EPI_ISL_4597399, EPI_ISL_4597401, EPI_ISL_4597402, EPI_ISL_4597422, EPI_ISL_4597423, EPI_ISL_4597498, EPI_ISL_4597499, EPI_ISL_4597561, EPI_ISL_4597615, EPI_ISL_4597772, EPI_ISL_4597927, EPI_ISL_4597989, EPI_ISL_4598009, EPI_ISL_4598052, EPI_ISL_4598062, EPI_ISL_4598095, EPI_ISL_4598102, EPI_ISL_4619955, EPI_ISL_4620009, EPI_ISL_4620045, EPI_ISL_4620225, EPI_ISL_4620450, EPI_ISL_4620479, EPI_ISL_4620541, EPI_ISL_4620626, EPI_ISL_4620670, EPI_ISL_4620816, EPI_ISL_4620834, EPI_ISL_4621040, EPI_ISL_4621048, EPI_ISL_4621076, EPI_ISL_4621102, EPI_ISL_4621221, EPI_ISL_4621820, EPI_ISL_4621851, EPI_ISL_4621886, EPI_ISL_4622078, EPI_ISL_4622204, EPI_ISL_4670362, EPI_ISL_4670564, EPI_ISL_4670580, EPI_ISL_4670668, EPI_ISL_4673331, EPI_ISL_4673337, EPI_ISL_4673407, EPI_ISL_4673416, EPI_ISL_4673490, EPI_ISL_4673508, EPI_ISL_4673745, EPI_ISL_4674142, EPI_ISL_4674266, EPI_ISL_4674336, EPI_ISL_4674376, EPI_ISL_4678607, EPI_ISL_4678665, EPI_ISL_4678693, EPI_ISL_4678751, EPI_ISL_4678805, EPI_ISL_4678815, EPI_ISL_4678885, EPI_ISL_4678892, EPI_ISL_4678918, EPI_ISL_4679085, EPI_ISL_4679176, EPI_ISL_4679186, EPI_ISL_4679321, EPI_ISL_4679364, EPI_ISL_4679377, EPI_ISL_4679388, EPI_ISL_4679497, EPI_ISL_4679528, EPI_ISL_4679556, EPI_ISL_4679627, EPI_ISL_4679662, EPI_ISL_4679727, EPI_ISL_4685500, EPI_ISL_4685542, EPI_ISL_4685731 | | | |
| see above | Integrated Covid Hub North East | Wellcome Sanger Institute for the COVID-19 Genomics UK (COG-UK) Consortium | Cordelia Langford; David K. Jackson; Dominic Kwiatkowski; Ewan Harrison; Ian Johnston; Integrated Covid Hub North East and Alex Alderton; Jeffrey Barrett; John Sillitoe on behalf of the Wellcome Sanger Institute COVID-19 Surveillance Team; Roberto Amato; Sonia Goncalves |
| EPI_ISL_4579122, EPI_ISL_4579176, EPI_ISL_4579201, EPI_ISL_4579209, EPI_ISL_4579230, EPI_ISL_4579248, EPI_ISL_4579290, EPI_ISL_4579322, EPI_ISL_4579354, EPI_ISL_4579500, EPI_ISL_4579505, EPI_ISL_4579519, EPI_ISL_4579531, EPI_ISL_4579539, EPI_ISL_4579546, EPI_ISL_4579551, EPI_ISL_4579559, EPI_ISL_4579577, EPI_ISL_4579596, EPI_ISL_4579608, EPI_ISL_4579630, EPI_ISL_4579698, EPI_ISL_4579775, EPI_ISL_4579812, EPI_ISL_4579898, EPI_ISL_4579964, EPI_ISL_4579987, EPI_ISL_4579993, EPI_ISL_4580008, EPI_ISL_4580104, EPI_ISL_4580124, EPI_ISL_4580144, EPI_ISL_4583385, EPI_ISL_4583474, EPI_ISL_4583530, EPI_ISL_4583629, EPI_ISL_4583631, EPI_ISL_4583687, EPI_ISL_4583853, EPI_ISL_4583866, EPI_ISL_4583944, EPI_ISL_4583988, EPI_ISL_4584178, EPI_ISL_4584280, EPI_ISL_4584313, EPI_ISL_4584336, EPI_ISL_4584349, EPI_ISL_4584357, EPI_ISL_4584360, EPI_ISL_4584422, EPI_ISL_4584438, EPI_ISL_4584455, EPI_ISL_4584492, EPI_ISL_4584511, EPI_ISL_4584610, EPI_ISL_4584725, EPI_ISL_4584790, EPI_ISL_4584853, EPI_ISL_4586638, EPI_ISL_4586667, EPI_ISL_4586739, EPI_ISL_4588081, EPI_ISL_4588119, EPI_ISL_4588133, EPI_ISL_4588523, EPI_ISL_4588611, EPI_ISL_4588703, EPI_ISL_4589361, EPI_ISL_4589447, EPI_ISL_4589527, EPI_ISL_4589560, EPI_ISL_4589563, EPI_ISL_4589578, EPI_ISL_4589713, EPI_ISL_4589730, EPI_ISL_4589734, EPI_ISL_4589809, EPI_ISL_4589888, EPI_ISL_4590173, EPI_ISL_4590336, EPI_ISL_4590402, EPI_ISL_4590508, EPI_ISL_4590557, EPI_ISL_4590631, EPI_ISL_4590804, EPI_ISL_4590893, EPI_ISL_4590936, EPI_ISL_4591014, EPI_ISL_4591060, EPI_ISL_4591113, EPI_ISL_4591292, EPI_ISL_4591351, EPI_ISL_4591517, EPI_ISL_4591546, EPI_ISL_4592479, EPI_ISL_4592610, EPI_ISL_4592984, EPI_ISL_4593242, EPI_ISL_4593316, EPI_ISL_4593512, EPI_ISL_4593689, EPI_ISL_4593809, EPI_ISL_4593918, EPI_ISL_4594102, EPI_ISL_4594113, EPI_ISL_4594135, EPI_ISL_4594139, EPI_ISL_4594141, EPI_ISL_4594151, EPI_ISL_4594158, EPI_ISL_4594171, EPI_ISL_4594174, EPI_ISL_4594214, EPI_ISL_4594219, EPI_ISL_4594257, EPI_ISL_4594278, EPI_ISL_4594290, EPI_ISL_4594294, EPI_ISL_4594299, EPI_ISL_4594305, EPI_ISL_4594309, EPI_ISL_4594324, EPI_ISL_4618326, EPI_ISL_4618625, EPI_ISL_4618627, EPI_ISL_4618670, EPI_ISL_4618693, EPI_ISL_4620086, EPI_ISL_4621583, EPI_ISL_4621716, EPI_ISL_4623961, EPI_ISL_4624065, EPI_ISL_4624335, EPI_ISL_4669424, EPI_ISL_4669578, EPI_ISL_4669680, EPI_ISL_4669731, EPI_ISL_4669759, EPI_ISL_4669868, EPI_ISL_4670037, EPI_ISL_4670099, EPI_ISL_4670134, EPI_ISL_4671181, EPI_ISL_4671254, EPI_ISL_4671653, EPI_ISL_4671853, EPI_ISL_4671955, EPI_ISL_4672213, EPI_ISL_4672230, EPI_ISL_4672329, EPI_ISL_4672340, EPI_ISL_4672373, EPI_ISL_4672392, EPI_ISL_4672411, EPI_ISL_4672430, EPI_ISL_4672497, EPI_ISL_4672542, EPI_ISL_4672566, EPI_ISL_4672572, EPI_ISL_4672616, EPI_ISL_4672701, EPI_ISL_4672715, EPI_ISL_4672949, EPI_ISL_4672976, EPI_ISL_4673173, EPI_ISL_4673251, EPI_ISL_4674594, EPI_ISL_4674600, EPI_ISL_4674622, EPI_ISL_4674750, EPI_ISL_4674934, EPI_ISL_4675052, EPI_ISL_4675121, EPI_ISL_4675131, EPI_ISL_4675328, EPI_ISL_4675582, EPI_ISL_4675586, EPI_ISL_4675679, EPI_ISL_4675695, EPI_ISL_4675706, EPI_ISL_4675760, EPI_ISL_4675775, EPI_ISL_4676110, EPI_ISL_4679833, EPI_ISL_4679904, EPI_ISL_4679947, EPI_ISL_4679953, EPI_ISL_4679977, EPI_ISL_4679989, EPI_ISL_4680009, EPI_ISL_4680050, EPI_ISL_4680104, EPI_ISL_4680132, EPI_ISL_4680363, EPI_ISL_4680381, EPI_ISL_4680382, EPI_ISL_4680466, EPI_ISL_4680595, EPI_ISL_4680658, EPI_ISL_4680664, EPI_ISL_4680710, EPI_ISL_4680790, EPI_ISL_4680801, EPI_ISL_4680910, EPI_ISL_4680973, EPI_ISL_4681005, EPI_ISL_4681094, EPI_ISL_4681133, EPI_ISL_4681394, EPI_ISL_4681403, EPI_ISL_4681447, EPI_ISL_4681508, EPI_ISL_4681513, EPI_ISL_4681518, EPI_ISL_4681531, EPI_ISL_4681547, EPI_ISL_4681548, EPI_ISL_4681601, EPI_ISL_4681609 | | | |
| see above | Lighthouse Lab in Alderley Park | Wellcome Sanger Institute for the COVID-19 Genomics UK (COG-UK) Consortium | Cordelia Langford; David K. Jackson; Dominic Kwiatkowski; Ewan Harrison; Ian Johnston; Jacquelyn Wynn; Jeffrey Barrett; John Sillitoe on behalf of the Wellcome Sanger Institute COVID-19 Surveillance Team; Mairead Hyland; Roberto Amato; Sonia Goncalves; The Lighthouse Lab in Alderley Park and Alex Alderton |
| EPI_ISL_4579016, EPI_ISL_4579108, EPI_ISL_4579359, EPI_ISL_4579412, EPI_ISL_4579605, EPI_ISL_4579679, EPI_ISL_4579715, EPI_ISL_4579804, EPI_ISL_4579920, EPI_ISL_4583730, EPI_ISL_4584512, EPI_ISL_4584564, EPI_ISL_4584570, EPI_ISL_4584776, EPI_ISL_4584845, EPI_ISL_4584944, EPI_ISL_4585656, EPI_ISL_4585673, EPI_ISL_4585728, EPI_ISL_4585736, EPI_ISL_4585761, EPI_ISL_4585789, EPI_ISL_4585856, EPI_ISL_4585872, EPI_ISL_4585883, EPI_ISL_4585888, EPI_ISL_4585899, EPI_ISL_4585944, EPI_ISL_4586091, EPI_ISL_4586128, EPI_ISL_4586168, EPI_ISL_4586233, EPI_ISL_4586259, EPI_ISL_4586271, EPI_ISL_4586272, EPI_ISL_4586529, EPI_ISL_4586580, EPI_ISL_4586660, EPI_ISL_4586693, EPI_ISL_4589781, EPI_ISL_4590024, EPI_ISL_4590065, EPI_ISL_4591859, EPI_ISL_4591958, EPI_ISL_4591980, EPI_ISL_4592135, EPI_ISL_4592208, EPI_ISL_4592226, EPI_ISL_4592234, EPI_ISL_4592255, EPI_ISL_4592316, EPI_ISL_4592359, EPI_ISL_4592453, EPI_ISL_4592488, EPI_ISL_4593100, EPI_ISL_4593277, EPI_ISL_4593367, EPI_ISL_4593436, EPI_ISL_4593476, EPI_ISL_4593508, EPI_ISL_4593525, EPI_ISL_4593700, EPI_ISL_4593868, EPI_ISL_4593882, EPI_ISL_4596361, EPI_ISL_4596411, EPI_ISL_4596527, EPI_ISL_4596697, EPI_ISL_4596702, EPI_ISL_4596781, EPI_ISL_4596849, EPI_ISL_4596868, EPI_ISL_4596884, EPI_ISL_4596947, EPI_ISL_4596959, EPI_ISL_4596962, EPI_ISL_4596963, EPI_ISL_4597012, EPI_ISL_4597028, EPI_ISL_4597031, EPI_ISL_4597064, EPI_ISL_4597084, EPI_ISL_4597093, EPI_ISL_4597179, EPI_ISL_4597216, EPI_ISL_4597261, EPI_ISL_4597265, EPI_ISL_4597275, EPI_ISL_4597277, EPI_ISL_4598206, EPI_ISL_4598228, EPI_ISL_4598248, EPI_ISL_4598327, EPI_ISL_4598446, EPI_ISL_4598455, EPI_ISL_4598482, EPI_ISL_4598512, EPI_ISL_4598536, EPI_ISL_4598574, EPI_ISL_4598612, EPI_ISL_4598641, EPI_ISL_4598662, EPI_ISL_4598668, EPI_ISL_4598684, EPI_ISL_4598723, EPI_ISL_4598802, EPI_ISL_4598830, EPI_ISL_4600146, EPI_ISL_4618584, EPI_ISL_4618692, EPI_ISL_4621357, EPI_ISL_4621369, EPI_ISL_4621438, EPI_ISL_4621463, EPI_ISL_4621564, EPI_ISL_4669332, EPI_ISL_4669356, EPI_ISL_4669372, EPI_ISL_4669481, EPI_ISL_4669510, EPI_ISL_4669734, EPI_ISL_4669735, EPI_ISL_4669742, EPI_ISL_4669751, EPI_ISL_4669827, EPI_ISL_4669907, EPI_ISL_4670002, EPI_ISL_4670005, EPI_ISL_4670052, EPI_ISL_4670058, EPI_ISL_4670111, EPI_ISL_4670216, EPI_ISL_4671173, EPI_ISL_4671242, EPI_ISL_4671287, EPI_ISL_4671329, EPI_ISL_4671356, EPI_ISL_4671403, EPI_ISL_4671427, EPI_ISL_4671634, EPI_ISL_4671661, EPI_ISL_4671741, EPI_ISL_4671742, EPI_ISL_4671865, EPI_ISL_4671912, EPI_ISL_4671927, EPI_ISL_4671928, EPI_ISL_4671957, EPI_ISL_4671965, EPI_ISL_4672008, EPI_ISL_4672060, EPI_ISL_4672074, EPI_ISL_4676711, EPI_ISL_4676745, EPI_ISL_4676964, EPI_ISL_4676987, EPI_ISL_4677424, EPI_ISL_4677438, EPI_ISL_4677619, EPI_ISL_4677869, EPI_ISL_4677928, EPI_ISL_4678059, EPI_ISL_4678385, EPI_ISL_4678405, EPI_ISL_4678429, EPI_ISL_4678455, EPI_ISL_4681626, EPI_ISL_4681634, EPI_ISL_4681674, EPI_ISL_4681680, EPI_ISL_4681696, EPI_ISL_4681711, EPI_ISL_4681727, EPI_ISL_4681766, EPI_ISL_4681777, EPI_ISL_4681786, EPI_ISL_4681845, EPI_ISL_4681860, EPI_ISL_4681867, EPI_ISL_4681963, EPI_ISL_4682133, EPI_ISL_4682146, EPI_ISL_4682175, EPI_ISL_4682336, EPI_ISL_4682409, EPI_ISL_4682484, EPI_ISL_4682485, EPI_ISL_4682533, EPI_ISL_4682548, EPI_ISL_4682593, EPI_ISL_4682644, EPI_ISL_4682766, EPI_ISL_4682800, EPI_ISL_4682806, EPI_ISL_4682807, EPI_ISL_4682832, EPI_ISL_4682852, EPI_ISL_4682863, EPI_ISL_4682893, EPI_ISL_4682931, EPI_ISL_4682955, EPI_ISL_4683000, EPI_ISL_4683086, EPI_ISL_4683107, EPI_ISL_4683115, EPI_ISL_4683179, EPI_ISL_4683190, EPI_ISL_4683221, EPI_ISL_4683224, EPI_ISL_4683264, EPI_ISL_4683265, EPI_ISL_4683304, EPI_ISL_4683335, EPI_ISL_4683343, EPI_ISL_4683449, EPI_ISL_4683468, EPI_ISL_4683504, EPI_ISL_4683506, EPI_ISL_4683557, EPI_ISL_4683558, EPI_ISL_4683614, EPI_ISL_4683678, EPI_ISL_4683746, EPI_ISL_4683754, EPI_ISL_4683835, EPI_ISL_4683842, EPI_ISL_4683845, EPI_ISL_4683854, EPI_ISL_4683861, EPI_ISL_4683862, EPI_ISL_4683872, EPI_ISL_4683882, EPI_ISL_4683898, EPI_ISL_4683901, EPI_ISL_4683907, EPI_ISL_4683945, EPI_ISL_4683953, EPI_ISL_4683976, EPI_ISL_4683985, EPI_ISL_4684277, EPI_ISL_4684300, EPI_ISL_4684306, EPI_ISL_4684335, EPI_ISL_4684388, EPI_ISL_4684461, EPI_ISL_4684478, EPI_ISL_4684505, EPI_ISL_4684532, EPI_ISL_4684551, EPI_ISL_4684579, EPI_ISL_4684594, EPI_ISL_4684606, EPI_ISL_4684613, EPI_ISL_4684614, EPI_ISL_4684632, EPI_ISL_4684637, EPI_ISL_4684652, EPI_ISL_4684683, EPI_ISL_4684708, EPI_ISL_4684720, EPI_ISL_4684813, EPI_ISL_4684816, EPI_ISL_4684838, EPI_ISL_4684848, EPI_ISL_4684870, EPI_ISL_4684894, EPI_ISL_4684895, EPI_ISL_4684947, EPI_ISL_4685010, EPI_ISL_4685016, EPI_ISL_4685048, EPI_ISL_4685115, EPI_ISL_4685116, EPI_ISL_4685127, EPI_ISL_4685142, EPI_ISL_4685159, EPI_ISL_4685192, EPI_ISL_4685224, EPI_ISL_4685287 | | | |
| see above | Lighthouse Lab in Glasgow | Wellcome Sanger Institute for the COVID-19 Genomics UK (COG-UK) Consortium | Anna Dominiczak and Alex Alderton; Carol Clugston; Cordelia Langford; David Gray; David K. Jackson; Dominic Kwiatkowski; Ewan Harrison; Harper VanSteenhouse; Ian Johnston; Jeffrey Barrett; John Sillitoe on behalf of the Wellcome Sanger Institute COVID-19 Surveillance Team; Roberto Amato; Sonia Goncalves; Yumi Kasai |
| EPI_ISL_4578908, EPI_ISL_4578972, EPI_ISL_4578988, EPI_ISL_4579086, EPI_ISL_4579118, EPI_ISL_4579173, EPI_ISL_4579213, EPI_ISL_4579226, EPI_ISL_4579288, EPI_ISL_4579313, EPI_ISL_4579318, EPI_ISL_4579326, EPI_ISL_4579365, EPI_ISL_4579371, EPI_ISL_4579651, EPI_ISL_4579654, EPI_ISL_4579772, EPI_ISL_4579848, EPI_ISL_4580461, EPI_ISL_4580491, EPI_ISL_4580533, EPI_ISL_4580645, EPI_ISL_4580818, EPI_ISL_4580819, EPI_ISL_4580880, EPI_ISL_4580890, EPI_ISL_4580966, EPI_ISL_4581042, EPI_ISL_4581147, EPI_ISL_4581148, EPI_ISL_4581160, EPI_ISL_4581185, EPI_ISL_4581256, EPI_ISL_4581362, EPI_ISL_4581426, EPI_ISL_4581528, EPI_ISL_4581748, EPI_ISL_4581835, EPI_ISL_4581874, EPI_ISL_4581905, EPI_ISL_4581995, EPI_ISL_4582190, EPI_ISL_4582392, EPI_ISL_4582534, EPI_ISL_4582616, EPI_ISL_4582665, EPI_ISL_4582692, EPI_ISL_4582698, EPI_ISL_4582728, EPI_ISL_4582813, EPI_ISL_4582986, EPI_ISL_4583100, EPI_ISL_4583203, EPI_ISL_4583380, EPI_ISL_4583406, EPI_ISL_4583418, EPI_ISL_4583508, EPI_ISL_4583704, EPI_ISL_4583708, EPI_ISL_4584131, EPI_ISL_4584182, EPI_ISL_4584245, EPI_ISL_4584343, EPI_ISL_4584379, EPI_ISL_4585151, EPI_ISL_4585207, EPI_ISL_4585222, EPI_ISL_4585228, EPI_ISL_4585265, EPI_ISL_4585342, EPI_ISL_4585348, EPI_ISL_4585365, EPI_ISL_4585386, EPI_ISL_4585393, EPI_ISL_4585410, EPI_ISL_4585417, EPI_ISL_4585420, EPI_ISL_4585426, EPI_ISL_4585517, EPI_ISL_4585540, EPI_ISL_4585546, EPI_ISL_4585571, EPI_ISL_4585613, EPI_ISL_4585632, EPI_ISL_4585636, EPI_ISL_4585889, EPI_ISL_4585970, EPI_ISL_4585993, EPI_ISL_4586013, EPI_ISL_4586025, EPI_ISL_4586030, EPI_ISL_4586105, EPI_ISL_4586146, EPI_ISL_4586204, EPI_ISL_4586245, EPI_ISL_4586247, EPI_ISL_4586353, EPI_ISL_4586433, EPI_ISL_4586454, EPI_ISL_4586996, EPI_ISL_4588011, EPI_ISL_4588037, EPI_ISL_4588093, EPI_ISL_4588105, EPI_ISL_4588135, EPI_ISL_4588385, EPI_ISL_4588628, EPI_ISL_4588685, EPI_ISL_4589324, EPI_ISL_4589414, EPI_ISL_4589675, EPI_ISL_4589715, EPI_ISL_4589724, EPI_ISL_4589725, EPI_ISL_4589747, EPI_ISL_4589754, EPI_ISL_4592471, EPI_ISL_4592533, EPI_ISL_4592684, EPI_ISL_4593029, EPI_ISL_4593043, EPI_ISL_4593049, EPI_ISL_4593079, EPI_ISL_4594402, EPI_ISL_4594430, EPI_ISL_4594436, EPI_ISL_4594457, EPI_ISL_4594517, EPI_ISL_4594540, EPI_ISL_4594542, EPI_ISL_4594562, EPI_ISL_4594587, EPI_ISL_4594635, EPI_ISL_4594640, EPI_ISL_4594651, EPI_ISL_4597659, EPI_ISL_4597905, EPI_ISL_4597979, EPI_ISL_4598060, EPI_ISL_4598077, EPI_ISL_4621006, EPI_ISL_4623285, EPI_ISL_4623307, EPI_ISL_4623363, EPI_ISL_4623585, EPI_ISL_4623693, EPI_ISL_4623741, EPI_ISL_4623763, EPI_ISL_4623781, EPI_ISL_4669425, EPI_ISL_4669497, EPI_ISL_4669536, EPI_ISL_4669715, EPI_ISL_4669769, EPI_ISL_4669813, EPI_ISL_4670028, EPI_ISL_4670240, EPI_ISL_4670297, EPI_ISL_4670300, EPI_ISL_4670420, EPI_ISL_4670450, EPI_ISL_4670464, EPI_ISL_4670483, EPI_ISL_4670519, EPI_ISL_4670526, EPI_ISL_4670591, EPI_ISL_4670600, EPI_ISL_4670728, EPI_ISL_4670754, EPI_ISL_4670845, EPI_ISL_4670955, EPI_ISL_4670957, EPI_ISL_4670981, EPI_ISL_4670988, EPI_ISL_4671001, EPI_ISL_4671015, EPI_ISL_4671104, EPI_ISL_4671107, EPI_ISL_4671125, EPI_ISL_4671331, EPI_ISL_4671720, EPI_ISL_4671902, EPI_ISL_4672243, EPI_ISL_4672282, EPI_ISL_4672298, EPI_ISL_4672371, EPI_ISL_4672519, EPI_ISL_4672558, EPI_ISL_4673111, EPI_ISL_4673136, EPI_ISL_4673172, EPI_ISL_4675938, EPI_ISL_4675940, EPI_ISL_4676046, EPI_ISL_4676055, EPI_ISL_4676218, EPI_ISL_4676309, EPI_ISL_4676342, EPI_ISL_4676451, EPI_ISL_4676500, EPI_ISL_4676577, EPI_ISL_4676686, EPI_ISL_4676687, EPI_ISL_4676824, EPI_ISL_4677248, EPI_ISL_4677328, EPI_ISL_4678297, EPI_ISL_4678347, EPI_ISL_4684215 | | | |
| see above | Lighthouse Lab in Milton Keynes | Wellcome Sanger Institute for the COVID-19 Genomics UK (COG-UK) Consortium | Cordelia Langford; David K. Jackson; Dominic Kwiatkowski; Ewan Harrison; Ian Johnston; Jeffrey Barrett; John Sillitoe on behalf of the Wellcome Sanger Institute COVID-19 Surveillance Team; Roberto Amato; Sonia Goncalves; The Lighthouse Lab in Milton Keynes and Alex Alderton |
| EPI_ISL_4578934, EPI_ISL_4578941, EPI_ISL_4578945, EPI_ISL_4578950, EPI_ISL_4578959, EPI_ISL_4578984, EPI_ISL_4579006, EPI_ISL_4579026, EPI_ISL_4579050, EPI_ISL_4579055, EPI_ISL_4579480, EPI_ISL_4579595, EPI_ISL_4579699, EPI_ISL_4579839, EPI_ISL_4579904, EPI_ISL_4579919, EPI_ISL_4581351, EPI_ISL_4581611, EPI_ISL_4581664, EPI_ISL_4581741, EPI_ISL_4581811, EPI_ISL_4581941, EPI_ISL_4583289, EPI_ISL_4583328, EPI_ISL_4583348, EPI_ISL_4583361, EPI_ISL_4583362, EPI_ISL_4583484, EPI_ISL_4583491, EPI_ISL_4583506, EPI_ISL_4583515, EPI_ISL_4583561, EPI_ISL_4583598, EPI_ISL_4583609, EPI_ISL_4583617, EPI_ISL_4583640, EPI_ISL_4583655, EPI_ISL_4583661, EPI_ISL_4583706, EPI_ISL_4584751, EPI_ISL_4589876, EPI_ISL_4589887, EPI_ISL_4589905, EPI_ISL_4590072, EPI_ISL_4590200, EPI_ISL_4590311, EPI_ISL_4590489, EPI_ISL_4590636, EPI_ISL_4591044, EPI_ISL_4591341, EPI_ISL_4591411, EPI_ISL_4592298, EPI_ISL_4594662, EPI_ISL_4594670, EPI_ISL_4594675, EPI_ISL_4594689, EPI_ISL_4594698, EPI_ISL_4594721, EPI_ISL_4594770, EPI_ISL_4594877, EPI_ISL_4594905, EPI_ISL_4594981, EPI_ISL_4595010, EPI_ISL_4595095, EPI_ISL_4595124, EPI_ISL_4595186, EPI_ISL_4595225, EPI_ISL_4595226, EPI_ISL_4595310, EPI_ISL_4595318, EPI_ISL_4595364, EPI_ISL_4595365, EPI_ISL_4595374, EPI_ISL_4595391, EPI_ISL_4596145, EPI_ISL_4596161, EPI_ISL_4596189, EPI_ISL_4596199, EPI_ISL_4596299, EPI_ISL_4596345, EPI_ISL_4596386, EPI_ISL_4596416, EPI_ISL_4596465, EPI_ISL_4596478, EPI_ISL_4596484, EPI_ISL_4596538, EPI_ISL_4597310, EPI_ISL_4597328, EPI_ISL_4597420, EPI_ISL_4597447, EPI_ISL_4597460, EPI_ISL_4597622, EPI_ISL_4597635, EPI_ISL_4597645, EPI_ISL_4597711, EPI_ISL_4598016, EPI_ISL_4598021, EPI_ISL_4618779, EPI_ISL_4618879, EPI_ISL_4618993, EPI_ISL_4618995, EPI_ISL_4619149, EPI_ISL_4619211, EPI_ISL_4619227, EPI_ISL_4619365, EPI_ISL_4619400, EPI_ISL_4619421, EPI_ISL_4619424, EPI_ISL_4619500, EPI_ISL_4619533, EPI_ISL_4619618, EPI_ISL_4619623, EPI_ISL_4619713, EPI_ISL_4679485 | | | |
| see above | Lighthouse Laboratory Plymouth | Wellcome Sanger Institute for the COVID-19 Genomics UK (COG-UK) Consortium | Cordelia Langford; David K. Jackson; Dominic Kwiatkowski; Ewan Harrison; Ian Johnston; Jeffrey Barrett; John Sillitoe on behalf of the Wellcome Sanger Institute COVID-19 Surveillance Team; Lighthouse Laboratory Plymouth and Alex Alderton; Roberto Amato; Sonia Goncalves |
| EPI_ISL_4529787, EPI_ISL_4529788, EPI_ISL_4529800, EPI_ISL_4529801, EPI_ISL_4529865, EPI_ISL_4530189, EPI_ISL_4530315, EPI_ISL_4530340, EPI_ISL_4530347, EPI_ISL_4530416, EPI_ISL_4530451, EPI_ISL_4530513, EPI_ISL_4530516, EPI_ISL_4530519, EPI_ISL_4530534, EPI_ISL_4530668, EPI_ISL_4530793, EPI_ISL_4532013 | | | |
| see above | Respiratory Virus Unit, Microbiology Services Colindale, Public Health England | COVID-19 Genomics UK (COG-UK) Consortium | PHE Covid Sequencing Team |
| EPI_ISL_4581209, EPI_ISL_4581511, EPI_ISL_4581769, EPI_ISL_4581873, EPI_ISL_4593709, EPI_ISL_4674490, EPI_ISL_4674932, EPI_ISL_4675112, EPI_ISL_4675144, EPI_ISL_4675400, EPI_ISL_4675601 | | | |
| see above | Rosalind Franklin Laboratory | Wellcome Sanger Institute for the COVID-19 Genomics UK (COG-UK) Consortium | Cordelia Langford; David K. Jackson; Dominic Kwiatkowski; Donald Fraser; Ewan Harrison; Ian Johnston; Jeffrey Barrett; John Sillitoe on behalf of the Wellcome Sanger Institute COVID-19 Surveillance Team; Rob Howes; Roberto Amato; Sonia Goncalves; Suki Lee; The Rosalind Franklin Laboratory and Alex Alderton |
