## Supplementary material for "SARS-COV-2 δ variant drives the pandemic in India and Europe via two subvariants": Acknowledgement table on the GISAID genomes used in this study

We gratefully acknowledge the following Authors from the Originating laboratories responsible for obtaining the specimens, as well as the Submitting laboratories where the genome data were generated and shared via GISAID, on which this research is based.

All Submitters of data may be contacted directly via www.gisaid.org

Authors are sorted alphabetically.

| Accession ID | Originating Laboratory | Submitting Laboratory | Authors |
|---|---|---|---|
| EPI_ISL_3771684, EPI_ISL_4305875 | Barts Health NHS Trust | COVID-19 Genomics UK (COG-UK) Consortium | BROAD; Beatrix; CUTINO-MOGUEL; Claire; David; Dola; HARRINGTON; KELE; KULASEGARAN-SHYLINI; Maria-Teresa; OWOYEMI; Raghavendran |
| EPI_ISL_4620243 | Bayerisches Landesamt für Gesundheit und Lebensmittelsicherheit (LGL) | Robert Koch Institute | |
| EPI_ISL_3169110, EPI_ISL_3284230, EPI_ISL_3314841, EPI_ISL_3363726, EPI_ISL_3379525, EPI_ISL_3435706, EPI_ISL_3435847, EPI_ISL_3435963, EPI_ISL_3436574, EPI_ISL_3436607, EPI_ISL_3436999, EPI_ISL_3437195, EPI_ISL_3437327, EPI_ISL_3437384, EPI_ISL_3438529, EPI_ISL_3441540, EPI_ISL_3441590, EPI_ISL_3441640, EPI_ISL_3441809, EPI_ISL_3441826, EPI_ISL_3441877, EPI_ISL_3442803, EPI_ISL_3442882, EPI_ISL_3442965, EPI_ISL_3467214, EPI_ISL_3467314, EPI_ISL_3467329, EPI_ISL_3467461, EPI_ISL_3528010, EPI_ISL_3528677, EPI_ISL_3529262, EPI_ISL_3560387, EPI_ISL_3560930, EPI_ISL_3561136, EPI_ISL_3561648, EPI_ISL_3562414, EPI_ISL_3562476, EPI_ISL_3562726, EPI_ISL_3563543, EPI_ISL_3563705, EPI_ISL_3641428, EPI_ISL_3642027, EPI_ISL_3702689, EPI_ISL_3729777, EPI_ISL_3765594, EPI_ISL_3767282, EPI_ISL_3952039, EPI_ISL_3954036, EPI_ISL_3964077, EPI_ISL_4013442, EPI_ISL_4068038, EPI_ISL_4085738, EPI_ISL_4087763, EPI_ISL_4088162, EPI_ISL_4088226, EPI_ISL_4217171, EPI_ISL_4247636, EPI_ISL_4264702, EPI_ISL_4264714, EPI_ISL_4288614, EPI_ISL_4288707, EPI_ISL_4288749, EPI_ISL_4288823, EPI_ISL_4290308, EPI_ISL_4320418, EPI_ISL_4323080, EPI_ISL_4324189, EPI_ISL_4326842, EPI_ISL_4326886, EPI_ISL_4326980, EPI_ISL_4327035, EPI_ISL_4327057, EPI_ISL_4349745, EPI_ISL_4349774, EPI_ISL_4350781, EPI_ISL_4350915, EPI_ISL_4350923, EPI_ISL_4351022, EPI_ISL_4351363, EPI_ISL_4352147, EPI_ISL_4402997, EPI_ISL_4451846, EPI_ISL_4453987, EPI_ISL_4477247, EPI_ISL_4477768, EPI_ISL_4500022, EPI_ISL_4500131, EPI_ISL_4500805, EPI_ISL_4502196, EPI_ISL_4502240, EPI_ISL_4502415, EPI_ISL_4502874, EPI_ISL_4521405, EPI_ISL_4524559, EPI_ISL_4524583, EPI_ISL_4524626, EPI_ISL_4524817, EPI_ISL_4525422, EPI_ISL_4526476, EPI_ISL_4526543, EPI_ISL_4526837, EPI_ISL_4527568, EPI_ISL_4527688, EPI_ISL_4584693, EPI_ISL_4586435, EPI_ISL_4586697, EPI_ISL_4589756, EPI_ISL_4591642, EPI_ISL_4591760, EPI_ISL_4592458, EPI_ISL_4592608, EPI_ISL_4592677, EPI_ISL_4592742, EPI_ISL_4592959 | | | |
| see above | Berkshire and Surrey Pathology Services Lighthouse Laboratory | Wellcome Sanger Institute for the COVID-19 Genomics UK (COG-UK) Consortium | Berkshire and Surrey Pathology Services Lighthouse Laboratory and Alex Alderton; Cordelia Langford; David K. Jackson; Dominic Kwiatkowski; Ewan Harrison; Ian Johnston; Jeffrey Barrett; John Sillitoe on behalf of the Wellcome Sanger Institute COVID-19 Surveillance Team; Roberto Amato; Sonia Goncalves |
| EPI_ISL_4609995 | CENTOGENE Frankfurt Laboratory: Niederlassung Industriepark Höchst | Robert Koch Institute | |
| EPI_ISL_4064785 | Center for Laboratory Medicine | Center for Laboratory Medicine | Yannick Gerth |
| EPI_ISL_3357245 | Department of Bacteria, Parasites and Fungi, Statens Serum Institut, Copenhagen, Denmark | Statens Serum Institut Bioinformatics and Microbial Genomics | Danish Covid-19 Genome Consortium |
| EPI_ISL_4558691 | Diamisanto GmbH | Institute of Medical Virology | Alexandra Trkola; Annette Audigé; Cyril Shah; Gabriela Ziltener; Guido Bloemberg; Jon Huder; Jürg Böni; Kevin Steiner; Maria Grünberg; Maryam Zaheri; Michael Huber; Riccarda Capaul; Stefan Schmutz; Verena Kufner |
| EPI_ISL_4228651, EPI_ISL_4442586 | Eurofins LifeCodexx GmbH | Robert Koch Institute | |
| EPI_ISL_4660614, EPI_ISL_4660623 | HOSPITAL DE BELLVITGE | Microbiology Department | Aida Gonzalez-Diaz; Carmen Ardanuy; Jordi Camara; Jordi Niubó; Laura Calatayud; M Angeles Domínguez; Sara Marti; Yolanda Hernandez |
| EPI_ISL_3309956, EPI_ISL_3311570, EPI_ISL_3366548, EPI_ISL_3437063, EPI_ISL_3441198, EPI_ISL_3442890, EPI_ISL_3455233, EPI_ISL_3455307, EPI_ISL_3469857, EPI_ISL_3499512, EPI_ISL_3560646, EPI_ISL_3580925, EPI_ISL_3581679, EPI_ISL_3641093, EPI_ISL_3641549, EPI_ISL_3641675, EPI_ISL_3642223, EPI_ISL_3702935, EPI_ISL_3703101, EPI_ISL_3764694, EPI_ISL_3767802, EPI_ISL_3884756, EPI_ISL_3964111, EPI_ISL_3992565, EPI_ISL_3992621, EPI_ISL_3992733, EPI_ISL_4012480, EPI_ISL_4015585, EPI_ISL_4046996, EPI_ISL_4047043, EPI_ISL_4048319, EPI_ISL_4086585, EPI_ISL_4100926, EPI_ISL_4100967, EPI_ISL_4323546, EPI_ISL_4325560, EPI_ISL_4325587, EPI_ISL_4325596, EPI_ISL_4325855, EPI_ISL_4326144, EPI_ISL_4326147, EPI_ISL_4326239, EPI_ISL_4326573, EPI_ISL_4500726, EPI_ISL_4676364 | | | |
| see above | Health Services Laboratories | Wellcome Sanger Institute for the COVID-19 Genomics UK (COG-UK) Consortium | Cordelia Langford; David K. Jackson; Dominic Kwiatkowski; Ewan Harrison; Health Services Laboratories and Alex Alderton; Ian Johnston; Jeffrey Barrett; John Sillitoe on behalf of the Wellcome Sanger Institute COVID-19 Surveillance Team; Roberto Amato; Sonia Goncalves |
| EPI_ISL_3969139, EPI_ISL_3969140, EPI_ISL_3969869, EPI_ISL_3970538, EPI_ISL_3975252, EPI_ISL_4015939, EPI_ISL_4101170, EPI_ISL_4101296, EPI_ISL_4144878, EPI_ISL_4192007, EPI_ISL_4192046, EPI_ISL_4192216, EPI_ISL_4192226, EPI_ISL_4290021, EPI_ISL_4290625, EPI_ISL_4290827, EPI_ISL_4327336, EPI_ISL_4329045, EPI_ISL_4353317, EPI_ISL_4499222, EPI_ISL_4522422, EPI_ISL_4522431, EPI_ISL_4522669, EPI_ISL_4522828, EPI_ISL_4522992, EPI_ISL_4523099, EPI_ISL_4523194, EPI_ISL_4585476, EPI_ISL_4585596, EPI_ISL_4587237, EPI_ISL_4587357, EPI_ISL_4587364, EPI_ISL_4587934, EPI_ISL_4588737, EPI_ISL_4589037, EPI_ISL_4589075, EPI_ISL_4596469, EPI_ISL_4597332, EPI_ISL_4597467, EPI_ISL_4597498, EPI_ISL_4620225, EPI_ISL_4621886, EPI_ISL_4622204, EPI_ISL_4670668, EPI_ISL_4673337, EPI_ISL_4673407, EPI_ISL_4673416, EPI_ISL_4674336, EPI_ISL_4678665, EPI_ISL_4679497, EPI_ISL_4733560, EPI_ISL_4733579, EPI_ISL_4733594, EPI_ISL_4735828, EPI_ISL_4736278 | | | |
| see above | Integrated Covid Hub North East | Wellcome Sanger Institute for the COVID-19 Genomics UK (COG-UK) Consortium | Cordelia Langford; David K. Jackson; Dominic Kwiatkowski; Ewan Harrison; Ian Johnston; Integrated Covid Hub North East and Alex Alderton; Jeffrey Barrett; John Sillitoe on behalf of the Wellcome Sanger Institute COVID-19 Surveillance Team; Roberto Amato; Sonia Goncalves |
| EPI_ISL_4609124, EPI_ISL_4616900 | LADR Zentrallabor DR. Kramer & Kollegen Geesthacht | Robert Koch Institute | |
| EPI_ISL_4624706 | LabKom - Labor Augsburg MVZ GmbH | Robert Koch Institute | |
| EPI_ISL_4615379 | Labor Becker & Kollegen (Standort München) | Robert Koch Institute | |
| EPI_ISL_4612196 | Labor Mönchengladbach MVZ Dr. Stein + Kollegen GbR | Robert Koch Institute | |
| EPI_ISL_3915579 | Laboratorium Badawczo-Rozwojowe Vitagenum | WSSE_Gorzow | Elżbieta Justyńska; Klaudia Kobendza-Włodarczak; Patrycja Faberska and Marek Magol; Renata Siegel |
| EPI_ISL_3824128 | Laboratorium Badawczo-Rozwojowe Vitagenum | Wojewódzka Stacja Sanitarno-Epidemiologiczna w Katowicach | Beata Rozwadowska |
| EPI_ISL_3225463, EPI_ISL_3580742, EPI_ISL_3883580, EPI_ISL_3951111, EPI_ISL_3953611, EPI_ISL_3955588, EPI_ISL_3957006, EPI_ISL_3990763, EPI_ISL_3993590, EPI_ISL_3994202, EPI_ISL_3995491, EPI_ISL_3995906, EPI_ISL_4011439, EPI_ISL_4047935, EPI_ISL_4049038, EPI_ISL_4065962, EPI_ISL_4098362, EPI_ISL_4098472, EPI_ISL_4143842, EPI_ISL_4189599, EPI_ISL_4189629, EPI_ISL_4189902, EPI_ISL_4190637, EPI_ISL_4190990, EPI_ISL_4215522, EPI_ISL_4215745, EPI_ISL_4215814, EPI_ISL_4215971, EPI_ISL_4250617, EPI_ISL_4250969, EPI_ISL_4250987, EPI_ISL_4250994, EPI_ISL_4263575, EPI_ISL_4287267, EPI_ISL_4287416, EPI_ISL_4287619, EPI_ISL_4288447, EPI_ISL_4289725, EPI_ISL_4319530, EPI_ISL_4319868, EPI_ISL_4326055, EPI_ISL_4327334, EPI_ISL_4327836, EPI_ISL_4349256, EPI_ISL_4350348, EPI_ISL_4352276, EPI_ISL_4355315, EPI_ISL_4355449, EPI_ISL_4402469, EPI_ISL_4402619, EPI_ISL_4405484, EPI_ISL_4406304, EPI_ISL_4452052, EPI_ISL_4452593, EPI_ISL_4452912, EPI_ISL_4453573, EPI_ISL_4453596, EPI_ISL_4453880, EPI_ISL_4453881, EPI_ISL_4454008, EPI_ISL_4477360, EPI_ISL_4478785, EPI_ISL_4498996, EPI_ISL_4499005, EPI_ISL_4500092, EPI_ISL_4500236, EPI_ISL_4500669, EPI_ISL_4501593, EPI_ISL_4501711, EPI_ISL_4501887, EPI_ISL_4501931, EPI_ISL_4502034, EPI_ISL_4502277, EPI_ISL_4502395, EPI_ISL_4521954, EPI_ISL_4521966, EPI_ISL_4522085, EPI_ISL_4522152, EPI_ISL_4522212, EPI_ISL_4522233, EPI_ISL_4522242, EPI_ISL_4522304, EPI_ISL_4522678, EPI_ISL_4523383, EPI_ISL_4523419, EPI_ISL_4523568, EPI_ISL_4525179, EPI_ISL_4525334, EPI_ISL_4525415, EPI_ISL_4525494, EPI_ISL_4525561, EPI_ISL_4525621, EPI_ISL_4525941, EPI_ISL_4526211, EPI_ISL_4526226, EPI_ISL_4527048, EPI_ISL_4527138, EPI_ISL_4527206, EPI_ISL_4527213, EPI_ISL_4527310, EPI_ISL_4527339, EPI_ISL_4527381, EPI_ISL_4579176, EPI_ISL_4579531, EPI_ISL_4579775, EPI_ISL_4580008, EPI_ISL_4583530, EPI_ISL_4584158, EPI_ISL_4584178, EPI_ISL_4584357, EPI_ISL_4584492, EPI_ISL_4584511, EPI_ISL_4584725, EPI_ISL_4584790, EPI_ISL_4586471, EPI_ISL_4586793, EPI_ISL_4588043, EPI_ISL_4590402, EPI_ISL_4590557, EPI_ISL_4590961, EPI_ISL_4591005, EPI_ISL_4591228, EPI_ISL_4593689, EPI_ISL_4594073, EPI_ISL_4594098, EPI_ISL_4594323, EPI_ISL_4618326, EPI_ISL_4622985, EPI_ISL_4623681, EPI_ISL_4623845, EPI_ISL_4623871, EPI_ISL_4624335, EPI_ISL_4671853, EPI_ISL_4674594, EPI_ISL_4674622, EPI_ISL_4674934, EPI_ISL_4675679, EPI_ISL_4681394, EPI_ISL_4732035, EPI_ISL_4732076, EPI_ISL_4732112, EPI_ISL_4732271, EPI_ISL_4732767, EPI_ISL_4732837, EPI_ISL_4733475, EPI_ISL_4733885, EPI_ISL_4734332, EPI_ISL_4734397, EPI_ISL_4734419, EPI_ISL_4734486, EPI_ISL_4735330, EPI_ISL_4735469, EPI_ISL_4735889, EPI_ISL_4735968, EPI_ISL_4750842, EPI_ISL_4750949, EPI_ISL_4751071, EPI_ISL_4751412, EPI_ISL_4751558, EPI_ISL_4751913, EPI_ISL_4751983, EPI_ISL_4752025, EPI_ISL_4752753, EPI_ISL_4752951, EPI_ISL_4753046, EPI_ISL_4753104, EPI_ISL_4753170, EPI_ISL_4753368, EPI_ISL_4753523, EPI_ISL_4753542, EPI_ISL_4753628, EPI_ISL_4753653, EPI_ISL_4753659, EPI_ISL_4755966, EPI_ISL_4762732, EPI_ISL_4763045, EPI_ISL_4763061 | | | |
| see above | Lighthouse Lab in Alderley Park | Wellcome Sanger Institute for the COVID-19 Genomics UK (COG-UK) Consortium | Cordelia Langford; David K. Jackson; Dominic Kwiatkowski; Ewan Harrison; Ian Johnston; Jacquelyn Wynn; Jeffrey Barrett; John Sillitoe on behalf of the Wellcome Sanger Institute COVID-19 Surveillance Team; Mairead Hyland; Roberto Amato; Sonia Goncalves; The Lighthouse Lab in Alderley Park and Alex Alderton |
| EPI_ISL_3582651, EPI_ISL_3697606, EPI_ISL_3700396, EPI_ISL_3704032, EPI_ISL_3768445, EPI_ISL_3807954, EPI_ISL_3812463, EPI_ISL_4086193, EPI_ISL_4324962, EPI_ISL_4354419, EPI_ISL_4354454, EPI_ISL_4402371, EPI_ISL_4403234, EPI_ISL_4480672, EPI_ISL_4480854, EPI_ISL_4481064, EPI_ISL_4502940, EPI_ISL_4579715, EPI_ISL_4579804, EPI_ISL_4579920, EPI_ISL_4599251, EPI_ISL_4599946, EPI_ISL_4621564, EPI_ISL_4735281 | | | |
| see above | Lighthouse Lab in Glasgow | Wellcome Sanger Institute for the COVID-19 Genomics UK (COG-UK) Consortium | Anna Dominiczak and Alex Alderton; Carol Clugston; Cordelia Langford; David Gray; David K. Jackson; Dominic Kwiatkowski; Ewan Harrison; Harper VanSteenhouse; Ian Johnston; Jeffrey Barrett; John Sillitoe on behalf of the Wellcome Sanger Institute COVID-19 Surveillance Team; Roberto Amato; Sonia Goncalves; Yumi Kasai |
| EPI_ISL_3167358, EPI_ISL_3223752, EPI_ISL_3249514, EPI_ISL_3284876, EPI_ISL_3285828, EPI_ISL_3312706, EPI_ISL_3314018, EPI_ISL_3335245, EPI_ISL_3335803, EPI_ISL_3335890, EPI_ISL_3335898, EPI_ISL_3336285, EPI_ISL_3337108, EPI_ISL_3337431, EPI_ISL_3365031, EPI_ISL_3379775, EPI_ISL_3435529, EPI_ISL_3437507, EPI_ISL_3437587, EPI_ISL_3437698, EPI_ISL_3446328, EPI_ISL_3453973, EPI_ISL_3454469, EPI_ISL_3466051, EPI_ISL_3466113, EPI_ISL_3466664, EPI_ISL_3466675, EPI_ISL_3467845, EPI_ISL_3468727, EPI_ISL_3469128, EPI_ISL_3469407, EPI_ISL_3469924, EPI_ISL_3470402, EPI_ISL_3470554, EPI_ISL_3498298, EPI_ISL_3499306, EPI_ISL_3499333, EPI_ISL_3499382, EPI_ISL_3528718, EPI_ISL_3528990, EPI_ISL_3528992, EPI_ISL_3529017, EPI_ISL_3560377, EPI_ISL_3563515, EPI_ISL_3563553, EPI_ISL_3563590, EPI_ISL_3564202, EPI_ISL_3564369, EPI_ISL_3637809, EPI_ISL_3640478, EPI_ISL_3641748, EPI_ISL_3641890, EPI_ISL_3641899, EPI_ISL_3642100, EPI_ISL_3642275, EPI_ISL_3700867, EPI_ISL_3700963, EPI_ISL_3701523, EPI_ISL_3701899, EPI_ISL_3702637, EPI_ISL_3703993, EPI_ISL_3704108, EPI_ISL_3704469, EPI_ISL_3704899, EPI_ISL_3706088, EPI_ISL_3706207, EPI_ISL_3706411, EPI_ISL_3726981, EPI_ISL_3727811, EPI_ISL_3728760, EPI_ISL_3763089, EPI_ISL_3763607, EPI_ISL_3763739, EPI_ISL_3764307, EPI_ISL_3765464, EPI_ISL_3766019, EPI_ISL_3767054, EPI_ISL_3767417, EPI_ISL_3767656, EPI_ISL_3768238, EPI_ISL_3768316, EPI_ISL_3768535, EPI_ISL_3807387, EPI_ISL_3807408, EPI_ISL_3807434, EPI_ISL_3807618, EPI_ISL_3808652, EPI_ISL_3808869, EPI_ISL_3808917, EPI_ISL_3810167, EPI_ISL_3810269, EPI_ISL_3836522, EPI_ISL_3836589, EPI_ISL_3882582, EPI_ISL_3882675, EPI_ISL_3884148, EPI_ISL_3884531, EPI_ISL_3950230, EPI_ISL_3950341, EPI_ISL_3950657, EPI_ISL_3952205, EPI_ISL_3952508, EPI_ISL_3952550, EPI_ISL_3952575, EPI_ISL_3956035, EPI_ISL_3964109, EPI_ISL_3964441, EPI_ISL_3964496, EPI_ISL_3964691, EPI_ISL_3965018, EPI_ISL_3965759, EPI_ISL_3966830, EPI_ISL_3967063, EPI_ISL_3967113, EPI_ISL_3967121, EPI_ISL_3967334, EPI_ISL_3967441, EPI_ISL_3967641, EPI_ISL_3990710, EPI_ISL_3991393, EPI_ISL_3991821, EPI_ISL_3992474, EPI_ISL_3994110, EPI_ISL_4009984, EPI_ISL_4011086, EPI_ISL_4011108, EPI_ISL_4011773, EPI_ISL_4012048, EPI_ISL_4012374, EPI_ISL_4012569, EPI_ISL_4012634, EPI_ISL_4013590, EPI_ISL_4013788, EPI_ISL_4013938, EPI_ISL_4014070, EPI_ISL_4016208, EPI_ISL_4017218, EPI_ISL_4017734, EPI_ISL_4017779, EPI_ISL_4017824, EPI_ISL_4017953, EPI_ISL_4018169, EPI_ISL_4018193, EPI_ISL_4018258, EPI_ISL_4018277, EPI_ISL_4018411, EPI_ISL_4020545, EPI_ISL_4046737, EPI_ISL_4047860, EPI_ISL_4048171, EPI_ISL_4065230, EPI_ISL_4065334, EPI_ISL_4065647, EPI_ISL_4065663, EPI_ISL_4065833, EPI_ISL_4067090, EPI_ISL_4067692, EPI_ISL_4086430, EPI_ISL_4089338, EPI_ISL_4096890, EPI_ISL_4097571, EPI_ISL_4097650, EPI_ISL_4099220, EPI_ISL_4099240, EPI_ISL_4099241, EPI_ISL_4099672, EPI_ISL_4142937, EPI_ISL_4142943, EPI_ISL_4142990, EPI_ISL_4143029, EPI_ISL_4143139, EPI_ISL_4143175, EPI_ISL_4143182, EPI_ISL_4143369, EPI_ISL_4143975, EPI_ISL_4144094, EPI_ISL_4188824, EPI_ISL_4188959, EPI_ISL_4188998, EPI_ISL_4189666, EPI_ISL_4191377, EPI_ISL_4191409, EPI_ISL_4191450, EPI_ISL_4192437, EPI_ISL_4192567, EPI_ISL_4213479, EPI_ISL_4213895, EPI_ISL_4213996, EPI_ISL_4215110, EPI_ISL_4215115, EPI_ISL_4215549, EPI_ISL_4216235, EPI_ISL_4216789, EPI_ISL_4217298, EPI_ISL_4217307, EPI_ISL_4218420, EPI_ISL_4248001, EPI_ISL_4248381, EPI_ISL_4248417, EPI_ISL_4248458, EPI_ISL_4248481, EPI_ISL_4248832, EPI_ISL_4249128, EPI_ISL_4250110, EPI_ISL_4263984, EPI_ISL_4266113, EPI_ISL_4266282, EPI_ISL_4266546, EPI_ISL_4266708, EPI_ISL_4266744, EPI_ISL_4266815, EPI_ISL_4266827, EPI_ISL_4286991, EPI_ISL_4288022, EPI_ISL_4320588, EPI_ISL_4320792, EPI_ISL_4320880, EPI_ISL_4321004, EPI_ISL_4321037, EPI_ISL_4321287, EPI_ISL_4321432, EPI_ISL_4321460, EPI_ISL_4321718, EPI_ISL_4321819, EPI_ISL_4322222, EPI_ISL_4323855, EPI_ISL_4324626, EPI_ISL_4325176, EPI_ISL_4349595, EPI_ISL_4349727, EPI_ISL_4349804, EPI_ISL_4349830, EPI_ISL_4349868, EPI_ISL_4349927, EPI_ISL_4350010, EPI_ISL_4350236, EPI_ISL_4350717, EPI_ISL_4350739, EPI_ISL_4352270, EPI_ISL_4352404, EPI_ISL_4402343, EPI_ISL_4402586, EPI_ISL_4403020, EPI_ISL_4403156, EPI_ISL_4404287, EPI_ISL_4404369, EPI_ISL_4404412, EPI_ISL_4404414, EPI_ISL_4404534, EPI_ISL_4404608, EPI_ISL_4404639, EPI_ISL_4404904, EPI_ISL_4405552, EPI_ISL_4405836, EPI_ISL_4405939, EPI_ISL_4405961, EPI_ISL_4451493, EPI_ISL_4451611, EPI_ISL_4451633, EPI_ISL_4451748, EPI_ISL_4451769, EPI_ISL_4452764, EPI_ISL_4452994, EPI_ISL_4453496, EPI_ISL_4453536, EPI_ISL_4453815, EPI_ISL_4478145, EPI_ISL_4478318, EPI_ISL_4478694, EPI_ISL_4479599, EPI_ISL_4498946, EPI_ISL_4498975, EPI_ISL_4499709, EPI_ISL_4500325, EPI_ISL_4500369, EPI_ISL_4500370, EPI_ISL_4500380, EPI_ISL_4500389, EPI_ISL_4500416, EPI_ISL_4500420, EPI_ISL_4500489, EPI_ISL_4500595, EPI_ISL_4502741, EPI_ISL_4521565, EPI_ISL_4521636, EPI_ISL_4521751, EPI_ISL_4521992, EPI_ISL_4522024, EPI_ISL_4522075, EPI_ISL_4522255, EPI_ISL_4522318, EPI_ISL_4526090, EPI_ISL_4526195, EPI_ISL_4526804, EPI_ISL_4579365, EPI_ISL_4580966, EPI_ISL_4585222, EPI_ISL_4585265, EPI_ISL_4585348, EPI_ISL_4585889, EPI_ISL_4586247, EPI_ISL_4586353, EPI_ISL_4586454, EPI_ISL_4588105, EPI_ISL_4588685, EPI_ISL_4594562, EPI_ISL_4594564, EPI_ISL_4595579, EPI_ISL_4595755, EPI_ISL_4595879, EPI_ISL_4595881, EPI_ISL_4596042, EPI_ISL_4596044, EPI_ISL_4596100, EPI_ISL_4597659, EPI_ISL_4598060, EPI_ISL_4669425, EPI_ISL_4670420, EPI_ISL_4670845, EPI_ISL_4672371, EPI_ISL_4676451, EPI_ISL_4676577, EPI_ISL_4676686, EPI_ISL_4676687, EPI_ISL_4734646, EPI_ISL_4735016, EPI_ISL_4735068, EPI_ISL_4735173, EPI_ISL_4735174, EPI_ISL_4735529, EPI_ISL_4735552, EPI_ISL_4735555, EPI_ISL_4735625, EPI_ISL_4735642, EPI_ISL_4735813, EPI_ISL_4735978, EPI_ISL_4736003, EPI_ISL_4750622, EPI_ISL_4750857, EPI_ISL_4753026, EPI_ISL_4754542, EPI_ISL_4754594, EPI_ISL_4754596, EPI_ISL_4754602, EPI_ISL_4754611, EPI_ISL_4754995, EPI_ISL_4755058, EPI_ISL_4755180, EPI_ISL_4755187, EPI_ISL_4755297, EPI_ISL_4755299, EPI_ISL_4755439, EPI_ISL_4755596, EPI_ISL_4762741, EPI_ISL_4763082, EPI_ISL_4763091, EPI_ISL_4763277, EPI_ISL_4763295, EPI_ISL_4763309, EPI_ISL_4763418, EPI_ISL_4767392 | | | |
| see above | Lighthouse Lab in Milton Keynes | Wellcome Sanger Institute for the COVID-19 Genomics UK (COG-UK) Consortium | Cordelia Langford; David K. Jackson; Dominic Kwiatkowski; Ewan Harrison; Ian Johnston; Jeffrey Barrett; John Sillitoe on behalf of the Wellcome Sanger Institute COVID-19 Surveillance Team; Roberto Amato; Sonia Goncalves; The Lighthouse Lab in Milton Keynes and Alex Alderton |
| EPI_ISL_3169553, EPI_ISL_3467997, EPI_ISL_3498864, EPI_ISL_3640955, EPI_ISL_3701006, EPI_ISL_3705013, EPI_ISL_3809785, EPI_ISL_3809833, EPI_ISL_3882023, EPI_ISL_3882372, EPI_ISL_3882491, EPI_ISL_3951994, EPI_ISL_3955293, EPI_ISL_3964422, EPI_ISL_4011821, EPI_ISL_4046320, EPI_ISL_4046367, EPI_ISL_4047262, EPI_ISL_4064908, EPI_ISL_4065004, EPI_ISL_4068928, EPI_ISL_4087482, EPI_ISL_4087851, EPI_ISL_4089517, EPI_ISL_4098160, EPI_ISL_4098403, EPI_ISL_4218091, EPI_ISL_4250044, EPI_ISL_4320434, EPI_ISL_4351887, EPI_ISL_4499796, EPI_ISL_4499907, EPI_ISL_4499930, EPI_ISL_4500039, EPI_ISL_4579904, EPI_ISL_4581941, EPI_ISL_4583484, EPI_ISL_4589961, EPI_ISL_4595095, EPI_ISL_4595225, EPI_ISL_4596189, EPI_ISL_4596199, EPI_ISL_4596386, EPI_ISL_4597328, EPI_ISL_4597447, EPI_ISL_4619424, EPI_ISL_4733338, EPI_ISL_4733375, EPI_ISL_4733854, EPI_ISL_4751348, EPI_ISL_4752238, EPI_ISL_4754918 | | | |
| see above | Lighthouse Laboratory Plymouth | Wellcome Sanger Institute for the COVID-19 Genomics UK (COG-UK) Consortium | Cordelia Langford; David K. Jackson; Dominic Kwiatkowski; Ewan Harrison; Ian Johnston; Jeffrey Barrett; John Sillitoe on behalf of the Wellcome Sanger Institute COVID-19 Surveillance Team; Lighthouse Laboratory Plymouth and Alex Alderton; Roberto Amato; Sonia Goncalves |
| EPI_ISL_4439649 | Limbach - MVZ Humangenetik Ulm | Robert Koch Institute | |

| | | | |
|---|---|---|---|
| EPI_ISL_4731435 | National Center of Infectious and Parasitic Diseases | National Center of Infectious and Parasitic Diseases | Alexiev et al |
| EPI_ISL_3421451, EPI_ISL_3571246, EPI_ISL_4114147, EPI_ISL_4115217, EPI_ISL_4115307, EPI_ISL_4306504, EPI_ISL_4306733 | | | |
| see above | Northumbria University / South Tees Hospitals NHS Foundation Trust / North Cumbria Integrated Care NHS Foundation Trust / North Tees and Hartlepool NHS Foundation Trust / Newcastle Hospitals NHS Foundation Trust | COVID-19 Genomics UK (COG-UK) Consortium | Andrew Nelson; Brendan Payne; Clive Graham; Darren L Smith; Debra Padgett; Edward Barton; Emma Swindells; Garren Scott; Gary Black; Gary Eltringham; Giles S Holt; Greg R Young; Jane Greenaway; Jennifer Collins; John Allan; Joshua Loh; Lynn Dover; Matthew Bashton; Mohammad A Tariq; Paul Baker; Sarah Essex; Steve Liggett; Wen C Yew; Yusri Taha |
| EPI_ISL_3422885, EPI_ISL_3423017, EPI_ISL_3423345, EPI_ISL_3423921, EPI_ISL_3575552, EPI_ISL_3575553, EPI_ISL_3575770, EPI_ISL_3576064, EPI_ISL_3576184, EPI_ISL_3576189, EPI_ISL_3788684, EPI_ISL_3788931, EPI_ISL_3791264, EPI_ISL_3791268, EPI_ISL_3962928, EPI_ISL_3963281, EPI_ISL_3964575, EPI_ISL_3964592, EPI_ISL_3964593, EPI_ISL_3965442, EPI_ISL_4133414, EPI_ISL_4133892, EPI_ISL_4134000, EPI_ISL_4134004, EPI_ISL_4134065, EPI_ISL_4134247, EPI_ISL_4134370, EPI_ISL_4134969, EPI_ISL_4135820, EPI_ISL_4135923, EPI_ISL_4135951, EPI_ISL_4135975, EPI_ISL_4135976, EPI_ISL_4136207, EPI_ISL_4136691, EPI_ISL_4137069, EPI_ISL_4313309, EPI_ISL_4314149, EPI_ISL_4315129, EPI_ISL_4315131, EPI_ISL_4315140, EPI_ISL_4315185, EPI_ISL_4315428, EPI_ISL_4316825, EPI_ISL_4317441, EPI_ISL_4318293, EPI_ISL_4318298, EPI_ISL_4318307, EPI_ISL_4318338, EPI_ISL_4319188, EPI_ISL_4319351, EPI_ISL_4319625, EPI_ISL_4532280, EPI_ISL_4532306, EPI_ISL_4532713, EPI_ISL_4532976, EPI_ISL_4532982, EPI_ISL_4533028 | | | |
| see above | Originating lab: Wales Specialist Virology Centre Sequencing lab: Pathogen Genomics Unit | Public Health Wales Microbiology Cardiff Wales Specialist Virology Centre | Alec Birchley; Alexander Adams; Amy Gaskin; Angela Marchbank; Bree Gatica-Wilcox; Catherine Moore; Jason Coombes; Joanne Watkins; Joel Southgate; Johnathan Evans; Laura Gifford; Lauren Gilbert; Lee Graham; Malorie Perry; Matthew Bull; Nicole Pacchiarini; Sally Corden; Sara Kumziene-Summerhayes; Sara Rey; Sarah Taylor; Simon Cottrell; Sophie Jones; Tom Connor |
| EPI_ISL_3773256, EPI_ISL_4307497, EPI_ISL_4529224, EPI_ISL_4529256, EPI_ISL_4529413, EPI_ISL_4529521, EPI_ISL_4529545 | | | |
| see above | Quadram Institute Bioscience | COVID-19 Genomics UK (COG-UK) Consortium | Alexander J Trotter; Alison E. Mather; Alp Aydin; Ana P. Tedim; Anastasia Kolyva; Andrew Bell; Andrew J. Page; Christopher Jeanes; Claire Stuart; Dave J. Baker; Ebenezer Foster-Nyarko; Gemma L. Kay; John Wain; Justin O'Grady; Leonardo de Oliveira Martins; Lewis G. Spurgin; Lindsay Coupland; Lizzie Meadows; Luke Bedford; Maria Diaz; Mark Webber; Martin Lott; Muhammed Yasir; Nabil-Fareed Alikhan; Ngozi Elumogo; Nicholas M. Thomson; Rachael Stanley; Rachel Gilroy; Reenesh Prakash; Rose K Davidson; Samir Dervisevic; Samuel Bloomfield; Sophie J. Prosolek; Steven Rudder; Thanh Le-Viet |
| EPI_ISL_3573145, EPI_ISL_3774470, EPI_ISL_3774715, EPI_ISL_3775677, EPI_ISL_3781427, EPI_ISL_3960523, EPI_ISL_3960645, EPI_ISL_3960722, EPI_ISL_3960736, EPI_ISL_3961952, EPI_ISL_3962002, EPI_ISL_3962504, EPI_ISL_4116905, EPI_ISL_4122892, EPI_ISL_4123613, EPI_ISL_4123680, EPI_ISL_4123774, EPI_ISL_4127888, EPI_ISL_4130339, EPI_ISL_4130471, EPI_ISL_4130480, EPI_ISL_4131134, EPI_ISL_4309330, EPI_ISL_4309547, EPI_ISL_4309960, EPI_ISL_4309968, EPI_ISL_4310982, EPI_ISL_4311237, EPI_ISL_4311297, EPI_ISL_4312137, EPI_ISL_4312386, EPI_ISL_4530233, EPI_ISL_4530340, EPI_ISL_4530516, EPI_ISL_4530873, EPI_ISL_4531731, EPI_ISL_4531832, EPI_ISL_4532003, EPI_ISL_4532069 | | | |
| see above | Respiratory Virus Unit, Microbiology Services Colindale, Public Health England | COVID-19 Genomics UK (COG-UK) Consortium | PHE Covid Sequencing Team |
| EPI_ISL_4674490, EPI_ISL_4675144, EPI_ISL_4675601 | Rosalind Franklin Laboratory | Wellcome Sanger Institute for the COVID-19 Genomics UK (COG-UK) Consortium | Cordelia Langford; David K. Jackson; Dominic Kwiatkowski; Donald Fraser; Ewan Harrison; Ian Johnston; Jeffrey Barrett; John Sillitoe on behalf of the Wellcome Sanger Institute COVID-19 Surveillance Team; Rob Howes; Roberto Amato; Sonia Goncalves; Suki Lee; The Rosalind Franklin Laboratory and Alex Alderton |
| EPI_ISL_4037229, EPI_ISL_4492583 | SYNLAB | GIGA Medical Genomics | Bouchra Boujemla; Claire Gourzonès; Cécile Meex; Keith Durkin; Laurent Gillet; Maria Artesi; Marie-Pierre Hayette; Nadine Cambisano; Nathalie Renotte; Olivier Ek; Sébastien Bontems; Vincent Bours |
| EPI_ISL_4446215, EPI_ISL_4613592, EPI_ISL_4617156, EPI_ISL_4617159 | SYNLAB MVZ Leinfelden-Echterdingen | Robert Koch Institute | |
| EPI_ISL_4607176 | SYNLAB MVZ Trier | Robert Koch Institute | |
| EPI_ISL_4222723, EPI_ISL_4226441, EPI_ISL_4226449, EPI_ISL_4230205, EPI_ISL_4438848, EPI_ISL_4446062, EPI_ISL_4608670, EPI_ISL_4610512, EPI_ISL_4610527, EPI_ISL_4610598, EPI_ISL_4615589 | | | |
| see above | SYNLAB MVZ Weiden | Robert Koch Institute | |
| EPI_ISL_4624148 | Sonic - Labor Staber Heilbronn | Robert Koch Institute | |
| EPI_ISL_4446445 | Synlab MVZ Augsburg | Robert Koch Institute | |
| EPI_ISL_3420566, EPI_ISL_4528231, EPI_ISL_4528325, EPI_ISL_4528332 | University College London, Great Ormond Street Hospital for Children NHS Foundation Trust, Imperial College Healthcare NHS Trust | COVID-19 Genomics UK (COG-UK) Consortium | Charlotte Williams; Helena Tutill; Judith Breuer; Marius Cotic; Mark Kristiansen; Nadua Bayzid; Patricia Dyal; Rachel Williams; Sergi Castellano; Sunando Roy |
| EPI_ISL_3424893, EPI_ISL_3424895, EPI_ISL_3424933 | Wales Specialist Virology Centre Sequencing lab: Pathogen Genomics Unit | Public Health Wales Microbiology Cardiff Wales Specialist Virology Centre | Alec Birchley; Alexander Adams; Amy Gaskin; Angela Marchbank; Bree Gatica-Wilcox; Catherine Moore; Jason Coombes; Joanne Watkins; Joel Southgate; Johnathan Evans; Laura Gifford; Lauren Gilbert; Lee Graham; Malorie Perry; Matthew Bull; Nicole Pacchiarini; Sally Corden; Sara Kumziene-Summerhayes; Sara Rey; Sarah Taylor; Simon Cottrell; Sophie Jones; Tom Connor |
| EPI_ISL_3983406 | Zakład Diagnostyki Laboratoryjnej | Wojewodzka Stacja Sanitarno-Epidemiologiczna w Gorzowie Wielkopolskim | Elzbieta Justynska; Klaudia Kobendza-Wlodarczak; Patrycja Faberska and Marek Magol; Renata Siegel |
| EPI_ISL_4587499 | labor team w AG | Department of Biosystems Science and Engineering, ETH Zürich | Andrea Patrignani; Andreas Lindauer; Andreia Cabral de Gouvea; Catharine Aquino; Chaoran Chen; Daniel Ehrsam; Doris Popovic; Griffin White; Isabel Stürmer; Ivan Topolsky; Jay Tracy; Kim Philipp Jablonski; Lara Fuhrmann; Laura Neff; Lennart Opitz; Louis du Plessis; Maria Domenica Moccia; Monika Bucher; Niko Beerenwinkel; Ralph Schlapbach; Rebekka Pohl; Sarah Nadeau; Simon Grüter; Tanja Stadler; Timothy Sykes |
