## Supplementary material for "SARS-COV-2 δ variant drives the pandemic in India and Europe via two subvariants": Acknowledgement table on the GISAID genomes used in this study

| | | | |
|---|---|---|---|
| EPI_ISL_4492600 | Clinique Notre Dame à Hermalle | GIGA Medical Genomics | Bouchra Boujemla; Claire Gourzonès; Cécile Meex; Keith Durkin; Laurent Gillet; Maria Artesi; Marie-Pierre Hayette; Nadine Cambisano; Nathalie Renotte; Olivier Ek; Sébastien Bontems; Vincent Bours |
| EPI_ISL_4069549, EPI_ISL_4274807, EPI_ISL_4274829, EPI_ISL_4274845, EPI_ISL_4274857, EPI_ISL_4493370, EPI_ISL_4493374, EPI_ISL_4493383, EPI_ISL_4493385, EPI_ISL_4493411, EPI_ISL_4629170 | | | |
| see above | Cliniques universitaires Saint-Luc | UCLouvain/IREC/MBLG | Benoit Kabamba Mukadi; Bertrand Bearzatto; Jean Ruelle |
| EPI_ISL_3989411, EPI_ISL_3989443, EPI_ISL_3989474, EPI_ISL_3989486, EPI_ISL_3989515, EPI_ISL_3989532, EPI_ISL_3989622, EPI_ISL_3989646, EPI_ISL_3989675, EPI_ISL_3989676, EPI_ISL_3989790, EPI_ISL_3989827, EPI_ISL_3989834, EPI_ISL_3989892, EPI_ISL_3989955, EPI_ISL_3989964, EPI_ISL_3990005, EPI_ISL_3990078, EPI_ISL_3990128, EPI_ISL_3990167, EPI_ISL_3990182, EPI_ISL_3990308, EPI_ISL_4006987, EPI_ISL_4007004, EPI_ISL_4007012, EPI_ISL_4007016, EPI_ISL_4007049, EPI_ISL_4007091, EPI_ISL_4007102, EPI_ISL_4007138, EPI_ISL_4007214, EPI_ISL_4007258, EPI_ISL_4007285, EPI_ISL_4007289, EPI_ISL_4007293, EPI_ISL_4007299, EPI_ISL_4007340, EPI_ISL_4007349, EPI_ISL_4007362, EPI_ISL_4007372, EPI_ISL_4007434, EPI_ISL_4007442, EPI_ISL_4007606, EPI_ISL_4007660, EPI_ISL_4007690, EPI_ISL_4007722, EPI_ISL_4007800, EPI_ISL_4007870, EPI_ISL_4007973, EPI_ISL_4007991, EPI_ISL_4008007, EPI_ISL_4008084, EPI_ISL_4008147, EPI_ISL_4008291, EPI_ISL_4008323, EPI_ISL_4008350, EPI_ISL_4049719, EPI_ISL_4049727, EPI_ISL_4049734, EPI_ISL_4049745, EPI_ISL_4049747, EPI_ISL_4049755, EPI_ISL_4049855, EPI_ISL_4049902, EPI_ISL_4049903, EPI_ISL_4049919, EPI_ISL_4049956, EPI_ISL_4049971, EPI_ISL_4049981, EPI_ISL_4049997, EPI_ISL_4050061, EPI_ISL_4050069, EPI_ISL_4050075, EPI_ISL_4050081, EPI_ISL_4050093, EPI_ISL_4050098, EPI_ISL_4050105, EPI_ISL_4050111, EPI_ISL_4050141, EPI_ISL_4050169, EPI_ISL_4050186, EPI_ISL_4050224, EPI_ISL_4050275, EPI_ISL_4050287, EPI_ISL_4050288, EPI_ISL_4050297, EPI_ISL_4050314, EPI_ISL_4050327, EPI_ISL_4050334, EPI_ISL_4050341, EPI_ISL_4050474, EPI_ISL_4050478, EPI_ISL_4050485, EPI_ISL_4050546, EPI_ISL_4050550, EPI_ISL_4050572, EPI_ISL_4069409, EPI_ISL_4069412, EPI_ISL_4069421, EPI_ISL_4069423, EPI_ISL_4069432, EPI_ISL_4069473, EPI_ISL_4105486, EPI_ISL_4105508, EPI_ISL_4105541, EPI_ISL_4105551, EPI_ISL_4105565, EPI_ISL_4105572, EPI_ISL_4105573, EPI_ISL_4105577, EPI_ISL_4105582, EPI_ISL_4105630, EPI_ISL_4105669, EPI_ISL_4105699, EPI_ISL_4105707, EPI_ISL_4105710, EPI_ISL_4105791, EPI_ISL_4105826, EPI_ISL_4105842, EPI_ISL_4105861, EPI_ISL_4105879, EPI_ISL_4105887, EPI_ISL_4105930, EPI_ISL_4105932, EPI_ISL_4105972, EPI_ISL_4105981, EPI_ISL_4170405, EPI_ISL_4170430, EPI_ISL_4170432, EPI_ISL_4170434, EPI_ISL_4170442, EPI_ISL_4170468, EPI_ISL_4170474, EPI_ISL_4170503, EPI_ISL_4170520, EPI_ISL_4170533, EPI_ISL_4170571, EPI_ISL_4170623, EPI_ISL_4170643, EPI_ISL_4170645, EPI_ISL_4170651, EPI_ISL_4170682, EPI_ISL_4170692, EPI_ISL_4199665, EPI_ISL_4199682, EPI_ISL_4199763, EPI_ISL_4199801, EPI_ISL_4199802, EPI_ISL_4199811, EPI_ISL_4199831, EPI_ISL_4199836, EPI_ISL_4199845, EPI_ISL_4199849, EPI_ISL_4199857, EPI_ISL_4199878, EPI_ISL_4199882, EPI_ISL_4199889, EPI_ISL_4199909, EPI_ISL_4199919, EPI_ISL_4199925, EPI_ISL_4199943, EPI_ISL_4199961, EPI_ISL_4199973, EPI_ISL_4199982, EPI_ISL_4199995, EPI_ISL_4200002, EPI_ISL_4200032, EPI_ISL_4200042, EPI_ISL_4200065, EPI_ISL_4200068, EPI_ISL_4200077, EPI_ISL_4200116, EPI_ISL_4200144, EPI_ISL_4200153, EPI_ISL_4200158, EPI_ISL_4200218, EPI_ISL_4200235, EPI_ISL_4200266, EPI_ISL_4200270, EPI_ISL_4200294, EPI_ISL_4200307, EPI_ISL_4200321, EPI_ISL_4200353, EPI_ISL_4200373, EPI_ISL_4200377, EPI_ISL_4200384, EPI_ISL_4200385, EPI_ISL_4200387, EPI_ISL_4200388, EPI_ISL_4200400, EPI_ISL_4200430, EPI_ISL_4200445, EPI_ISL_4200447, EPI_ISL_4200466, EPI_ISL_4200479, EPI_ISL_4200500, EPI_ISL_4200525, EPI_ISL_4200527, EPI_ISL_4200546, EPI_ISL_4200572, EPI_ISL_4200586, EPI_ISL_4200596, EPI_ISL_4200598, EPI_ISL_4200602, EPI_ISL_4200621, EPI_ISL_4200629, EPI_ISL_4200635, EPI_ISL_4200652, EPI_ISL_4200655, EPI_ISL_4200710, EPI_ISL_4232687, EPI_ISL_4232688, EPI_ISL_4232701, EPI_ISL_4232711, EPI_ISL_4232712, EPI_ISL_4232716, EPI_ISL_4232718, EPI_ISL_4232722, EPI_ISL_4232736, EPI_ISL_4232741, EPI_ISL_4232766, EPI_ISL_4232782, EPI_ISL_4232843, EPI_ISL_4232847, EPI_ISL_4232851, EPI_ISL_4232902, EPI_ISL_4232925, EPI_ISL_4232931, EPI_ISL_4233010, EPI_ISL_4233021, EPI_ISL_4233052, EPI_ISL_4233060, EPI_ISL_4233071, EPI_ISL_4233080, EPI_ISL_4233102, EPI_ISL_4233130, EPI_ISL_4233134, EPI_ISL_4233208, EPI_ISL_4233209, EPI_ISL_4233226, EPI_ISL_4233276, EPI_ISL_4233300, EPI_ISL_4256634, EPI_ISL_4256681, EPI_ISL_4256712, EPI_ISL_4256725, EPI_ISL_4257995, EPI_ISL_4258000, EPI_ISL_4258023, EPI_ISL_4258024, EPI_ISL_4258058, EPI_ISL_4258099, EPI_ISL_4258100, EPI_ISL_4258101, EPI_ISL_4258117, EPI_ISL_4258122, EPI_ISL_4258127, EPI_ISL_4258136, EPI_ISL_4258140, EPI_ISL_4258150, EPI_ISL_4258181, EPI_ISL_4258197, EPI_ISL_4258205, EPI_ISL_4258216, EPI_ISL_4258221, EPI_ISL_4258245, EPI_ISL_4258249, EPI_ISL_4258254, EPI_ISL_4258258, EPI_ISL_4258268, EPI_ISL_4258273, EPI_ISL_4258293, EPI_ISL_4258305, EPI_ISL_4258307, EPI_ISL_4258320, EPI_ISL_4258349, EPI_ISL_4303421, EPI_ISL_4303436, EPI_ISL_4303437, EPI_ISL_4303451, EPI_ISL_4303481, EPI_ISL_4303482, EPI_ISL_4303528, EPI_ISL_4303532, EPI_ISL_4303544, EPI_ISL_4303547, EPI_ISL_4303554, EPI_ISL_4303558, EPI_ISL_4303559, EPI_ISL_4303568, EPI_ISL_4303573, EPI_ISL_4303578, EPI_ISL_4303590, EPI_ISL_4303605, EPI_ISL_4303624, EPI_ISL_4303625, EPI_ISL_4303658, EPI_ISL_4354720, EPI_ISL_4354735, EPI_ISL_4354755, EPI_ISL_4354776, EPI_ISL_4354817, EPI_ISL_4354828, EPI_ISL_4354834, EPI_ISL_4354848, EPI_ISL_4354955, EPI_ISL_4354963, EPI_ISL_4355021, EPI_ISL_4355118, EPI_ISL_4355181, EPI_ISL_4355283, EPI_ISL_4355340, EPI_ISL_4355452, EPI_ISL_4355456, EPI_ISL_4355488, EPI_ISL_4355627, EPI_ISL_4355633, EPI_ISL_4355647, EPI_ISL_4355658, EPI_ISL_4355659, EPI_ISL_4355668, EPI_ISL_4355675, EPI_ISL_4355690, EPI_ISL_4355693, EPI_ISL_4355714, EPI_ISL_4355720, EPI_ISL_4355752, EPI_ISL_4355759, EPI_ISL_4355768, EPI_ISL_4355794, EPI_ISL_4355800, EPI_ISL_4355809, EPI_ISL_4355833, EPI_ISL_4355864, EPI_ISL_4355885, EPI_ISL_4355928, EPI_ISL_4394097, EPI_ISL_4447627, EPI_ISL_4447653, EPI_ISL_4447655, EPI_ISL_4447671, EPI_ISL_4447706, EPI_ISL_4447721, EPI_ISL_4447726, EPI_ISL_4447758, EPI_ISL_4447806, EPI_ISL_4447822, EPI_ISL_4447824, EPI_ISL_4447841, EPI_ISL_4447872, EPI_ISL_4447881, EPI_ISL_4447889, EPI_ISL_4448077, EPI_ISL_4448147, EPI_ISL_4448227, EPI_ISL_4448243, EPI_ISL_4448267, EPI_ISL_4448308, EPI_ISL_4448327, EPI_ISL_4448333, EPI_ISL_4448338, EPI_ISL_4448356, EPI_ISL_4448361, EPI_ISL_4448368, EPI_ISL_4448389, EPI_ISL_4448398, EPI_ISL_4448433, EPI_ISL_4448474, EPI_ISL_4448498, EPI_ISL_4448581, EPI_ISL_4448587, EPI_ISL_4448605, EPI_ISL_4448622, EPI_ISL_4448657, EPI_ISL_4448664, EPI_ISL_4448669, EPI_ISL_4448677, EPI_ISL_4448679, EPI_ISL_4448736, EPI_ISL_4448770, EPI_ISL_4448808, EPI_ISL_4448811, EPI_ISL_4448826, EPI_ISL_4448839, EPI_ISL_4448854, EPI_ISL_4448858, EPI_ISL_4448882, EPI_ISL_4448898, EPI_ISL_4448899, EPI_ISL_4448919, EPI_ISL_4448938, EPI_ISL_4448990, EPI_ISL_4448992, EPI_ISL_4449013, EPI_ISL_4449014, EPI_ISL_4449021, EPI_ISL_4449032, EPI_ISL_4449039, EPI_ISL_4449046, EPI_ISL_4449049, EPI_ISL_4449075, EPI_ISL_4449090, EPI_ISL_4449122, EPI_ISL_4449129, EPI_ISL_4473488, EPI_ISL_4473489, EPI_ISL_4473491, EPI_ISL_4473511, EPI_ISL_4473524, EPI_ISL_4473601, EPI_ISL_4473640, EPI_ISL_4473651, EPI_ISL_4473676, EPI_ISL_4473698, EPI_ISL_4473713, EPI_ISL_4473747, EPI_ISL_4473768, EPI_ISL_4473775, EPI_ISL_4473812, EPI_ISL_4473877, EPI_ISL_4520857, EPI_ISL_4520875, EPI_ISL_4520876, EPI_ISL_4520889, EPI_ISL_4520927, EPI_ISL_4520928, EPI_ISL_4520955, EPI_ISL_4520959, EPI_ISL_4520961, EPI_ISL_4520970, EPI_ISL_4520979, EPI_ISL_4520986, EPI_ISL_4520998, EPI_ISL_4521013, EPI_ISL_4521017, EPI_ISL_4521035, EPI_ISL_4521036, EPI_ISL_4521047, EPI_ISL_4521068, EPI_ISL_4521076, EPI_ISL_4521086, EPI_ISL_4521095, EPI_ISL_4521129, EPI_ISL_4521131, EPI_ISL_4521138, EPI_ISL_4521141, EPI_ISL_4521166, EPI_ISL_4521173, EPI_ISL_4521193, EPI_ISL_4521225, EPI_ISL_4521243, EPI_ISL_4551362, EPI_ISL_4551377, EPI_ISL_4551410, EPI_ISL_4551487, EPI_ISL_4551500, EPI_ISL_4551532, EPI_ISL_4625474, EPI_ISL_4625720, EPI_ISL_4625736, EPI_ISL_4625767, EPI_ISL_4625812, EPI_ISL_4625822, EPI_ISL_4625857, EPI_ISL_4625875, EPI_ISL_4625879, EPI_ISL_4625896, EPI_ISL_4625898, EPI_ISL_4625917, EPI_ISL_4625932, EPI_ISL_4625942, EPI_ISL_4625986, EPI_ISL_4625990, EPI_ISL_4626082, EPI_ISL_4626084, EPI_ISL_4626089, EPI_ISL_4626101, EPI_ISL_4626133, EPI_ISL_4626165, EPI_ISL_4626201, EPI_ISL_4626222, EPI_ISL_4626230, EPI_ISL_4626237, EPI_ISL_4626238, EPI_ISL_4626289, EPI_ISL_4626296, EPI_ISL_4626319, EPI_ISL_4626320, EPI_ISL_4626329, EPI_ISL_4626330, EPI_ISL_4626366, EPI_ISL_4626394, EPI_ISL_4626406, EPI_ISL_4626412, EPI_ISL_4626454, EPI_ISL_4626470, EPI_ISL_4626486, EPI_ISL_4626525, EPI_ISL_4626541, EPI_ISL_4626573, EPI_ISL_4626607, EPI_ISL_4626622, EPI_ISL_4626625, EPI_ISL_4626664, EPI_ISL_4626672, EPI_ISL_4626676, EPI_ISL_4626683, EPI_ISL_4626699, EPI_ISL_4626715, EPI_ISL_4626721, EPI_ISL_4626728, EPI_ISL_4626758, EPI_ISL_4626769, EPI_ISL_4626776, EPI_ISL_4626791, EPI_ISL_4626800, EPI_ISL_4626868, EPI_ISL_4643363, EPI_ISL_4643625, EPI_ISL_4643629, EPI_ISL_4643925, EPI_ISL_4644006, EPI_ISL_4644148, EPI_ISL_4644416, EPI_ISL_4644575, EPI_ISL_4644723, EPI_ISL_4644792, EPI_ISL_4644936, EPI_ISL_4645013, EPI_ISL_4645207, EPI_ISL_4645274, EPI_ISL_4645400, EPI_ISL_4645518, EPI_ISL_4645529, EPI_ISL_4645667, EPI_ISL_4645715 | | | |
| see above | Department of Bacteria, Parasites and Fungi, Statens Serum Institut, Copenhagen, Denmark | Statens Serum Institut Bioinformatics and Microbial Genomics | Danish Covid-19 Genome Consortium |
| EPI_ISL_4489328, EPI_ISL_4492662, EPI_ISL_4492667 | Department of Clinical Microbiology | GIGA Medical Genomics | Bouchra Boujemla; Claire Gourzonès; Cécile Meex; Keith Durkin; Laurent Gillet; Maria Artesi; Marie-Pierre Hayette; Nadine Cambisano; Nathalie Renotte; Olivier Ek; Sébastien Bontems; Vincent Bours |
| EPI_ISL_4303431, EPI_ISL_4303574 | Department of Clinical Microbiology and Center for Genomic Medicine, Rigshospitalet, Copenhagen, Denmark | Statens Serum Institut Bioinformatics and Microbial Genomics | Danish Covid-19 Genome Consortium |
| EPI_ISL_4256716, EPI_ISL_4256717, EPI_ISL_4256724, EPI_ISL_4447666, EPI_ISL_4449065, EPI_ISL_4449082 | Department of Clinical Microbiology, Odense University Hospital, Odense, Denmark | Statens Serum Institut Bioinformatics and Microbial Genomics | Danish Covid-19 Genome Consortium |
| EPI_ISL_4398123, EPI_ISL_4398251, EPI_ISL_4634746 | Department of Medical Microbiology - section Molde, Molde Hospital | Norwegian Institute of Public Health, Department of Virology | Atiya R Ali; Debech Nadia; Engebretsen Serina Beate; Garcia Llorente Ignacio; Hilde Elshaug; Hilde Vollan; Jon Bråte; Kamilla Heddeland Instefjord; Karoline Bragstad; Kathrine Stene-Johansen; Line Victoria Moen; Marie Paulsen Madsen; Olav Hungnes; Pedersen Benedikte Nevjen; Rasmus Riis Kopperud |
| EPI_ISL_4505349 | Dept. of Microbiology and Infection Control, Akershus University Hospital HF | Dept. of Microbiology and Infection Control, Akershus University Hospital HF | Alexander Hesselberg Løvestad; Hege Vangstein Aamot |
| EPI_ISL_4446887 | Diagnosticum - Labor Neukirchen | Robert Koch Institute | |
| EPI_ISL_4413063 | Dianalabs SA | Genesupport | Geraldine Jost; Katia Jaton; Nadia Liassine; Tanguy ARAUD |
| EPI_ISL_4204382, EPI_ISL_4204388, EPI_ISL_4204396, EPI_ISL_4633367, EPI_ISL_4633734 | Division of Emerging Infectious Diseases, Bureau of Infectious Diseases Diagnosis Control, Korea Disease Control and Prevention Agency | Division of Emerging Infectious Diseases, Bureau of Infectious Diseases Diagnosis Control, Korea Disease Control and Prevention Agency | Ae Kyung Park; Chae Young Lee; Eun-Jin Kim; Heui Man Kim; Il-Hwan Kim; Jeong-Ah Kim |
| EPI_ISL_4482941 | Dutch COVID-19 response team | Erasmus Medical Center | Anne van der Linden; Annemiek van der Eijk; Bas Oude Munnink; Corine GeurtsvanKessel; David Nieuwenhuijse; Emmanuelle Munger; Irina Chestakova; Marion Koopmans; Marjan Boter; Reina Sikkema; Richard Molenkamp; on behalf of the Dutch national COVID-19 response team. |
| EPI_ISL_4399875, EPI_ISL_4400273, EPI_ISL_4400762, EPI_ISL_4400765, EPI_ISL_4400807, EPI_ISL_4401112, EPI_ISL_4401200, EPI_ISL_4401891, EPI_ISL_4402178, EPI_ISL_4544165, EPI_ISL_4544177, EPI_ISL_4544182, EPI_ISL_4544183, EPI_ISL_4544195, EPI_ISL_4544213, EPI_ISL_4544381, EPI_ISL_4544392, EPI_ISL_4544413, EPI_ISL_4544541, EPI_ISL_4544590, EPI_ISL_4544615, EPI_ISL_4544653, EPI_ISL_4544791, EPI_ISL_4544792, EPI_ISL_4544815, EPI_ISL_4544818, EPI_ISL_4544840 | | | |
| see above | Dutch COVID-19 response team | National Institute for Public Health and the Environment (RIVM) | Adam Meijer; AnneMarie van den Brandt; Annelies Kroneman; Bas van der Veer; Chantal Reusken; Dennis Schmitz; Dirk Eggink; Euníce Then; Florian Zwagemaker; Harry Vennema; Ivo van Walle; Jeroen Cremer; Karim Hajji; Kim Freriks; Lisa Wijsman; Lynn Aarts; Ryanne Jaarsma; Sanne Bos; Sharon van den Brink; Stijn van Rossum; on behalf of the national COVID-19 response team |
| EPI_ISL_4413146 | EHNV | Laboratory of genomics and metagenomics | Claire Bertelli; Damien Jacot; Gilbert Greub; Sébastien Aeby; Trestan Pillonel |
| EPI_ISL_4607088 | EXALAB LE HAILLAN | CNR Virus des Infections Respiratoires - France SUD | Antonin Bal; Bruno Lina; Gregory Destras; Gwendolyne Burfin; Hadrien Regue; Laurence Josset; Martine Valette; Quentin Semanas |
| EPI_ISL_4615436 | Eurofins LifeCodexx GmbH | Robert Koch Institute | |
| EPI_ISL_4042813, EPI_ISL_4042845, EPI_ISL_4042938, EPI_ISL_4228730, EPI_ISL_4616205 | Eurofins MVZ Labor Gelsenkirchen | Robert Koch Institute | |
| EPI_ISL_4543176, EPI_ISL_4666485 | Fundacio Althaia-Manresa | Banc de Sang i Teixits | Carlos Hobeich; Francisco Vidal; Irene Corrales; Lorena Ramírez; Maria Glòria Soria; Natàlia Comes; Nina Borràs; Noemí Gonzalez; Silvia Sauleda |
| EPI_ISL_4269603 | Gravity Diagnostics, LLC | Gravity Diagnostics, LLC | Gravity Diagnostics |
| EPI_ISL_4660695, EPI_ISL_4660811 | HOSPITAL DE BELLVITGE | Microbiology Department | Aida Gonzalez-Diaz; Carmen Ardanuy; Jordi Camara; Jordi Niubó; Laura Calatayud; M Angeles Domínguez; Sara Marti; Yolanda Hernandez |
| EPI_ISL_4201574, EPI_ISL_4662921, EPI_ISL_4663000 | HOSPITAL UNIVERSITARI DE BELLVITGE | Microbiology Department | Aida Gonzalez-Diaz; Carmen Ardanuy; Jordi Camara; Jordi Niubó; Laura Calatayud; M Angeles Domínguez; Sara Marti; Yolanda Hernandez |
| EPI_ISL_4547936, EPI_ISL_4547942 | HOSPITAL UNIVERSITARIO CENTRAL DE ASTURIAS | Laboratorio de Virología HUCA | ; Alba L; Alvarez-Arguelles ME; Boga JA; Costales I; Coto E; González-Alba JM; Gómez de Oña J; Martín-Rodríguez G; Melón S; Perez-Martínez Z; Rojo S; Sandoval M |
| EPI_ISL_4137153, EPI_ISL_4137167, EPI_ISL_4137216, EPI_ISL_4350099, EPI_ISL_4350100, EPI_ISL_4350137, EPI_ISL_4350170, EPI_ISL_4350213, EPI_ISL_4350254, EPI_ISL_4550889, EPI_ISL_4550891, EPI_ISL_4550893, EPI_ISL_4550897 | | | |
| see above | HOSPITAL UNIVERSITARIO SON ESPASES | HOSPITAL UNIVERSITARIO SON ESPASES | Dr. Antonio Oliver; Dr. Carla López-Causapé; Dr. Gabriel Cabot; Hospital Universitario Son Espases; on behalf of Servicio de Microbiología |
| EPI_ISL_4066928 | Health Services Laboratories | Wellcome Sanger Institute for the COVID-19 Genomics UK (COG-UK) Consortium | Cordelia Langford; David K. Jackson; Dominic Kwiatkowski; Ewan Harrison; Health Services Laboratories and Alex Alderton; Ian Johnston; Jeffrey Barrett; John Sillitoe on behalf of the Wellcome Sanger Institute COVID-19 Surveillance Team; Roberto Amato; Sonia Goncalves |
| EPI_ISL_4204689 | Heilig hart Lier | Imelda Ziekenhuis | Dagmar Obbels; Hanne Valgaeren; Johan Frans |
| EPI_ISL_4461630, EPI_ISL_4461780, EPI_ISL_4503709, EPI_ISL_4503807, EPI_ISL_4545361, EPI_ISL_4545365 | Hospital | National Reference Center for Viruses of Respiratory Infections, Institut Pasteur, Paris | Alexandra Ducancelle; Angela Brisebarre; Camille Capel; Christophe Malabat; Corinne Maufrais; Etienne Simon-Lorière; Frédéric Lemoine; Isabelle Gros; Julien Fumey; JéRome Guinard; L. Courdavault; Louise Lefrançois; Marianne Burgard; Marion Barbet; Maud Vanpeene; Méline Bizard; Slim El Khiari; Slim El-Khiari; Sylvie Behillil; Sylvie Van der Werf; Vincent Enouf |
| EPI_ISL_4395984, | Hospital General Universitario | Hospital General Universitario | Cristina Rodriguez-Grande; Darío García de Viedma; Julia Suárez; Laura Pérez-Lago; Marta Herranz Martin; Patricia Muñoz; Pedro Sola Campoy; Pilar Catalán; Sergio Buenestado Serrano; Victor Manuel de la Cueva |
