## Supplementary material for "SARS-COV-2 δ variant drives the pandemic in India and Europe via two subvariants": Acknowledgement table on the GISAID genomes used in this study

We gratefully acknowledge the following Authors from the Originating laboratories responsible for obtaining the specimens, as well as the Submitting laboratories where the genome data were generated and shared via GISAID, on which this research is based.

All Submitters of data may be contacted directly via www.gisaid.org

Authors are sorted alphabetically.

| Accession ID | Originating Laboratory | Submitting Laboratory | Authors |
| --- | --- | --- | --- |
| EPI_ISL_3163403, EPI_ISL_3228396, EPI_ISL_3228839, EPI_ISL_3228964, EPI_ISL_3240191, EPI_ISL_3240569, EPI_ISL_3240656, EPI_ISL_3240851, EPI_ISL_3240981, EPI_ISL_3252138, EPI_ISL_3252640, EPI_ISL_3340332, EPI_ISL_3340346, EPI_ISL_3340608, EPI_ISL_3357055, EPI_ISL_3357061, EPI_ISL_3357440, EPI_ISL_3357693, EPI_ISL_3357751, EPI_ISL_3357880, EPI_ISL_3383970, EPI_ISL_3417869, EPI_ISL_3417881, EPI_ISL_3418153, EPI_ISL_3418160, EPI_ISL_3418331, EPI_ISL_3449485, EPI_ISL_3449625, EPI_ISL_3450219, EPI_ISL_3450247, EPI_ISL_3450675, EPI_ISL_3462933, EPI_ISL_3463959, EPI_ISL_3464290, EPI_ISL_3530384, EPI_ISL_3530884, EPI_ISL_3530888, EPI_ISL_3531497, EPI_ISL_3531656, EPI_ISL_3531701, EPI_ISL_3531850, EPI_ISL_3531963, EPI_ISL_3531966, EPI_ISL_3532172, EPI_ISL_3532309, EPI_ISL_3566775, EPI_ISL_3566850, EPI_ISL_3567028, EPI_ISL_3567209, EPI_ISL_3567309, EPI_ISL_3567553, EPI_ISL_3567585, EPI_ISL_3567779, EPI_ISL_3567827, EPI_ISL_3567979, EPI_ISL_3631752, EPI_ISL_3631795, EPI_ISL_3632017, EPI_ISL_3632031, EPI_ISL_3632362, EPI_ISL_3632399, EPI_ISL_3632476, EPI_ISL_3632554, EPI_ISL_3632762, EPI_ISL_3632851, EPI_ISL_3632858, EPI_ISL_3633346, EPI_ISL_3633810, EPI_ISL_3633819, EPI_ISL_3664531, EPI_ISL_3664537, EPI_ISL_3664592, EPI_ISL_3664638, EPI_ISL_3664735, EPI_ISL_3664854, EPI_ISL_3664857, EPI_ISL_3665217, EPI_ISL_3665219, EPI_ISL_3665262, EPI_ISL_3665272, EPI_ISL_3665494, EPI_ISL_3707913, EPI_ISL_3708094, EPI_ISL_3708185, EPI_ISL_3708194, EPI_ISL_3708201, EPI_ISL_3708328, EPI_ISL_3708333, EPI_ISL_3708335, EPI_ISL_3708378, EPI_ISL_3708402, EPI_ISL_3708415, EPI_ISL_3708534, EPI_ISL_3708596, EPI_ISL_3708673, EPI_ISL_3708744, EPI_ISL_3708756, EPI_ISL_3724621, EPI_ISL_3724630, EPI_ISL_3724668, EPI_ISL_3724798, EPI_ISL_3724941, EPI_ISL_3725092, EPI_ISL_3725094, EPI_ISL_3725168, EPI_ISL_3725171, EPI_ISL_3725273, EPI_ISL_3725422, EPI_ISL_3725478, EPI_ISL_3725578, EPI_ISL_3725617, EPI_ISL_3725632, EPI_ISL_3725736, EPI_ISL_3725762, EPI_ISL_3769721, EPI_ISL_3769748, EPI_ISL_3769851, EPI_ISL_3769909, EPI_ISL_3769935, EPI_ISL_3769948, EPI_ISL_3769992, EPI_ISL_3770001, EPI_ISL_3770073, EPI_ISL_3770083, EPI_ISL_3770144, EPI_ISL_3770156, EPI_ISL_3770161, EPI_ISL_3770263, EPI_ISL_3770271, EPI_ISL_3770363, EPI_ISL_3770401, EPI_ISL_3770411, EPI_ISL_3806574, EPI_ISL_3839338, EPI_ISL_3839374, EPI_ISL_3839690, EPI_ISL_3839737, EPI_ISL_3839809, EPI_ISL_3839894, EPI_ISL_3839896, EPI_ISL_3839995, EPI_ISL_3840009, EPI_ISL_3840123, EPI_ISL_3840192, EPI_ISL_3840240, EPI_ISL_3840281, EPI_ISL_3840426, EPI_ISL_3840428, EPI_ISL_3903743, EPI_ISL_3903811, EPI_ISL_3903820, EPI_ISL_3903934, EPI_ISL_3904113, EPI_ISL_3904127, EPI_ISL_3904149, EPI_ISL_3904170, EPI_ISL_3904183, EPI_ISL_3904226, EPI_ISL_3904231, EPI_ISL_3904306, EPI_ISL_3904316, EPI_ISL_3904330, EPI_ISL_3904358, EPI_ISL_3904368, EPI_ISL_3904381, EPI_ISL_3916816, EPI_ISL_3916830, EPI_ISL_3916845, EPI_ISL_3916898, EPI_ISL_3916980, EPI_ISL_3917058, EPI_ISL_3917084, EPI_ISL_3917086, EPI_ISL_3917123, EPI_ISL_3917157, EPI_ISL_3917312, EPI_ISL_3917349, EPI_ISL_3917358, EPI_ISL_3917472, EPI_ISL_3917568, EPI_ISL_3917604, EPI_ISL_3917613, EPI_ISL_3917660, EPI_ISL_3917771, EPI_ISL_3917792, EPI_ISL_3917934, EPI_ISL_3948944, EPI_ISL_3949012, EPI_ISL_3949117, EPI_ISL_3949154, EPI_ISL_3949164, EPI_ISL_3949241, EPI_ISL_3949322, EPI_ISL_3949477, EPI_ISL_3989411, EPI_ISL_3989474, EPI_ISL_3989488, EPI_ISL_3989515, EPI_ISL_3989532, EPI_ISL_3989596, EPI_ISL_3989601, EPI_ISL_3989622, EPI_ISL_3989646, EPI_ISL_3989672, EPI_ISL_3989684, EPI_ISL_3989780, EPI_ISL_3989826, EPI_ISL_3989843, EPI_ISL_3989886, EPI_ISL_3990005, EPI_ISL_3990078, EPI_ISL_3990134, EPI_ISL_3990167, EPI_ISL_3990308, EPI_ISL_4006987, EPI_ISL_4007004, EPI_ISL_4007016, EPI_ISL_4007091, EPI_ISL_4007102, EPI_ISL_4007138, EPI_ISL_4007214, EPI_ISL_4007299, EPI_ISL_4007362, EPI_ISL_4007372, EPI_ISL_4007434, EPI_ISL_4007442, EPI_ISL_4007606, EPI_ISL_4007690, EPI_ISL_4007722, EPI_ISL_4007800, EPI_ISL_4007870, EPI_ISL_4007964, EPI_ISL_4008007, EPI_ISL_4008147, EPI_ISL_4008291, EPI_ISL_4008323, EPI_ISL_4008350, EPI_ISL_4049727, EPI_ISL_4049734, EPI_ISL_4049745, EPI_ISL_4049747, EPI_ISL_4049755, EPI_ISL_4049797, EPI_ISL_4049835, EPI_ISL_4049855, EPI_ISL_4049902, EPI_ISL_4049919, EPI_ISL_4049971, EPI_ISL_4049981, EPI_ISL_4049997, EPI_ISL_4050061, EPI_ISL_4050069, EPI_ISL_4050075, EPI_ISL_4050081, EPI_ISL_4050098, EPI_ISL_4050105, EPI_ISL_4050141, EPI_ISL_4050169, EPI_ISL_4050224, EPI_ISL_4050314, EPI_ISL_4050327, EPI_ISL_4050341, EPI_ISL_4050474, EPI_ISL_4050478, EPI_ISL_4050485, EPI_ISL_4050546, EPI_ISL_4050550, EPI_ISL_4069412, EPI_ISL_4069421, EPI_ISL_4069423, EPI_ISL_4069432, EPI_ISL_4069473, EPI_ISL_4105541, EPI_ISL_4105551, EPI_ISL_4105565, EPI_ISL_4105572, EPI_ISL_4105582, EPI_ISL_4105707, EPI_ISL_4105710, EPI_ISL_4105826, EPI_ISL_4105861, EPI_ISL_4105887, EPI_ISL_4105932, EPI_ISL_4105972, EPI_ISL_4105981, EPI_ISL_4170430, EPI_ISL_4170432, EPI_ISL_4170503, EPI_ISL_4170643, EPI_ISL_4170682, EPI_ISL_4199665, EPI_ISL_4199682, EPI_ISL_4199763, EPI_ISL_4199801, EPI_ISL_4199811, EPI_ISL_4199845, EPI_ISL_4199849, EPI_ISL_4199857, EPI_ISL_4199878, EPI_ISL_4199882, EPI_ISL_4199909, EPI_ISL_4199919, EPI_ISL_4199925, EPI_ISL_4199943, EPI_ISL_4199961, EPI_ISL_4199982, EPI_ISL_4199995, EPI_ISL_4200002, EPI_ISL_4200042, EPI_ISL_4200068, EPI_ISL_4200158, EPI_ISL_4200218, EPI_ISL_4200235, EPI_ISL_4200307, EPI_ISL_4200321, EPI_ISL_4200353, EPI_ISL_4200373, EPI_ISL_4200377, EPI_ISL_4200385, EPI_ISL_4200387, EPI_ISL_4200388, EPI_ISL_4200400, EPI_ISL_4200430, EPI_ISL_4200488, EPI_ISL_4200500, EPI_ISL_4200525, EPI_ISL_4200527, EPI_ISL_4200546, EPI_ISL_4200553, EPI_ISL_4200598, EPI_ISL_4200629, EPI_ISL_4200635, EPI_ISL_4200652, EPI_ISL_4232687, EPI_ISL_4232688, EPI_ISL_4232718, EPI_ISL_4232722, EPI_ISL_4232736, EPI_ISL_4232741, EPI_ISL_4232766, EPI_ISL_4232782, EPI_ISL_4232902, EPI_ISL_4232925, EPI_ISL_4233010, EPI_ISL_4233060, EPI_ISL_4233071, EPI_ISL_4233102, EPI_ISL_4233130, EPI_ISL_4233134, EPI_ISL_4233208, EPI_ISL_4233209, EPI_ISL_4233226, EPI_ISL_4233276, EPI_ISL_4256634, EPI_ISL_4256681, EPI_ISL_4256712, EPI_ISL_4256725, EPI_ISL_4257190, EPI_ISL_4257413, EPI_ISL_4257523, EPI_ISL_4257686, EPI_ISL_4257694, EPI_ISL_4257918, EPI_ISL_4258000, EPI_ISL_4258023, EPI_ISL_4258058, EPI_ISL_4258099, EPI_ISL_4258100, EPI_ISL_4258117, EPI_ISL_4258122, EPI_ISL_4258127, EPI_ISL_4258136, EPI_ISL_4258140, EPI_ISL_4258181, EPI_ISL_4258197, EPI_ISL_4258205, EPI_ISL_4258245, EPI_ISL_4258254, EPI_ISL_4258268, EPI_ISL_4258273, EPI_ISL_4258293, EPI_ISL_4258305, EPI_ISL_4258307, EPI_ISL_4258349, EPI_ISL_4303421, EPI_ISL_4303436, EPI_ISL_4303481, EPI_ISL_4303544, EPI_ISL_4303547, EPI_ISL_4303554, EPI_ISL_4303558, EPI_ISL_4303568, EPI_ISL_4303573, EPI_ISL_4303578, EPI_ISL_4303590, EPI_ISL_4303605, EPI_ISL_4303624, EPI_ISL_4303625, EPI_ISL_4303658, EPI_ISL_4354720, EPI_ISL_4354755, EPI_ISL_4354776, EPI_ISL_4354817, EPI_ISL_4354828, EPI_ISL_4354834, EPI_ISL_4354848, EPI_ISL_4354955, EPI_ISL_4355021, EPI_ISL_4355118, EPI_ISL_4355181, EPI_ISL_4355283, EPI_ISL_4355456, EPI_ISL_4355627, EPI_ISL_4355633, EPI_ISL_4355658, EPI_ISL_4355659, EPI_ISL_4355668, EPI_ISL_4355714, EPI_ISL_4355794, EPI_ISL_4355800, EPI_ISL_4355809, EPI_ISL_4355833, EPI_ISL_4355885, EPI_ISL_4355928, EPI_ISL_4394057, EPI_ISL_4394097, EPI_ISL_4447726, EPI_ISL_4447824, EPI_ISL_4447881, EPI_ISL_4448077, EPI_ISL_4448147, EPI_ISL_4448227, EPI_ISL_4448243, EPI_ISL_4448267, EPI_ISL_4448327, EPI_ISL_4448361, EPI_ISL_4448398, EPI_ISL_4448474, EPI_ISL_4448498, EPI_ISL_4448581, EPI_ISL_4448657, EPI_ISL_4448677, EPI_ISL_4448808, EPI_ISL_4448826, EPI_ISL_4448858, EPI_ISL_4448919, EPI_ISL_4449013, EPI_ISL_4449014, EPI_ISL_4449021, EPI_ISL_4449032, EPI_ISL_4449049, EPI_ISL_4449090, EPI_ISL_4473511, EPI_ISL_4473524, EPI_ISL_4473676, EPI_ISL_4473698, EPI_ISL_4473747, EPI_ISL_4473775, EPI_ISL_4473812, EPI_ISL_4520857, EPI_ISL_4520876, EPI_ISL_4520889, EPI_ISL_4520927, EPI_ISL_4520928, EPI_ISL_4520955, EPI_ISL_4520959, EPI_ISL_4520961, EPI_ISL_4520986, EPI_ISL_4521017, EPI_ISL_4521068, EPI_ISL_4521076, EPI_ISL_4521086, EPI_ISL_4521131, EPI_ISL_4521138, EPI_ISL_4521141, EPI_ISL_4521173, EPI_ISL_4521193, EPI_ISL_4521243, EPI_ISL_4551362, EPI_ISL_4551377, EPI_ISL_4625720, EPI_ISL_4625736, EPI_ISL_4625748, EPI_ISL_4625767, EPI_ISL_4625875, EPI_ISL_4625896, EPI_ISL_4625898, EPI_ISL_4625917, EPI_ISL_4625942, EPI_ISL_4626084, EPI_ISL_4626089, EPI_ISL_4626101, EPI_ISL_4626133, EPI_ISL_4626165, EPI_ISL_4626201, EPI_ISL_4626222, EPI_ISL_4626296, EPI_ISL_4626319, EPI_ISL_4626329, EPI_ISL_4626366, EPI_ISL_4626412, EPI_ISL_4626454, EPI_ISL_4626525, EPI_ISL_4626541, EPI_ISL_4626607, EPI_ISL_4626664, EPI_ISL_4626672, EPI_ISL_4626683, EPI_ISL_4626699, EPI_ISL_4626715, EPI_ISL_4626721, EPI_ISL_4626728, EPI_ISL_4626758 | | | |
| see above | Department of Bacteria, Parasites and Fungi, Statens Serum Institut, Copenhagen, Denmark | Statens Serum Institut Bioinformatics and Microbial Genomics | Danish Covid-19 Genome Consortium |
| EPI_ISL_3948940, EPI_ISL_3949518, EPI_ISL_4303431, EPI_ISL_4303574 | Department of Clinical Microbiology and Center for Genomic Medicine, Rigshospitalet, Copenhagen, Denmark | Statens Serum Institut Bioinformatics and Microbial Genomics | Danish Covid-19 Genome Consortium |
| EPI_ISL_3339841, EPI_ISL_3989828 | Department of Clinical Microbiology, Odense University Hospital, Odense, Denmark | Statens Serum Institut Bioinformatics and Microbial Genomics | Danish Covid-19 Genome Consortium |
